# Clinical effectiveness of SGLT2 inhibitors in non-diabetic kidney transplanted patients- a real world data analysis

**DOI:** 10.64898/2026.05.22.26353858

**Authors:** Jamila-Cate Tran, Zhejia Tian, Jonas Willerding, Janis Casper, Kai Schmidt-Ott, Anette Melk, Bernhard MW Schmidt

## Abstract

**Background and hypothesis:** Sodium-glucose cotransporter-2 inhibitors (SGLT2-inhibitors) slow chronic kidney disease progression, but evidence in non-diabetic kidney transplant recipients is limited. We evaluated associations between SGLT2-inhibitor use and major adverse kidney events (MAKE), major adverse cardiovascular events (MACE), and all-cause mortality.

**Methods:** In this retrospective cohort study using the TriNetX federated research network, adult non-diabetic kidney transplant recipients transplanted between January 2015 and January 2022 were identified. SGLT2-inhibitor users initiating therapy ≥1000 days post-transplant were compared with non-users after 1:1 propensity score matching. The primary outcome was MAKE, defined as dialysis initiation or death. Secondary outcomes included all-cause mortality and MACE.

**Results:** Propensity score matching yielded 867 pairs of SGLT2-inhibitor users and non-users. SGLT2-inhibitor use was associated with lower risks of MAKE (adjusted hazard ratio [aHR] 0.64, 95% CI 0.45-0.91) and all-cause mortality (aHR 0.55, 95% CI 0.36-0.85). No significant association was observed for MACE (aHR 0.86, 95% CI 0.64-1.17). No increased risk of urinary tract infections was observed among SGLT2-inhibitor users.

**Conclusion:** SGLT2-inhibitor use was associated with lower risks of MAKE and all-cause mortality in non-diabetic kidney transplant recipients.

## Introduction

Sodium–glucose cotransporter-2 (SGLT2) inhibitors have emerged as a key disease-modifying therapy in chronic kidney disease (CKD)^1^ and heart failure^2^, with benefits extending far beyond glycaemic control. Large randomized trials and meta-analyses^3^ have demonstrated that SGLT2 inhibitors slow kidney function decline, reduce major adverse cardiovascular events, and lower all-cause mortality in patients with CKD, including those without diabetes (DAPA-CKD, EMPA-KIDNEY). Guidelines therefore recommend SGLT2 inhibitors across a broad spectrum of CKD stages and etiologies, irrespective of type 2 diabetes status, particularly in individuals at moderate to very high risk of CKD progression and complications (KDIGO CKD guideline 2024).

In kidney transplant recipients with diabetes, the use of SGLT2 inhibitors is increasingly reported and appears clinically promising. Observational cohorts, registry analyses, systematic reviews^4,5^, and large real-world datasets suggest that SGLT2 inhibitor therapy in diabetic kidney transplant recipients is associated with improved glycaemic control, favourable effects on body weight and blood pressure, and reductions in all-cause mortality, major adverse cardiovascular events (MACE), and major adverse kidney events (MAKE). At the same time, available data indicate an acceptable safety profile without major signals for increased acute rejection, severe volume depletion, or excess genitourinary infections, although follow-up durations remain limited and sample sizes modest. Nonetheless, most published studies have focused on post-transplant patients with pre-existing or post-transplant diabetes, and evidence for non-diabetic kidney transplant recipients is scarce.

Given the expanding indication of SGLT2 inhibitors in non-diabetic CKD and the growing but still incomplete evidence base in diabetic kidney transplant recipients, the potential role of SGLT2 inhibitors in kidney transplant recipients without diabetes remains uncertain. In this context, we investigated the use of SGLT2 inhibitors in non-diabetic kidney transplant recipients and evaluated their association with major adverse kidney events (MAKE), major adverse cardiovascular events (MACE), and all-cause mortality. By focusing on this in studies underrepresented population, our study aims to address a relevant knowledge gap and to inform future strategies for optimizing long-term cardiorenal risk reduction after kidney transplantation.

## Methods

### Data source

We performed a retrospective cohort study using patient data obtained from TriNetX, a federated health research platform that provides access to de-identified standardized electronic medical records from a large number of participating health care organizations (HCOs) worldwide^6^. TriNetX is used for studies on real-world evidence and includes demographic characteristics, diagnoses, procedures, medication records, and laboratory data. The data quality across the network is maintained through routine checks for conformance, completeness, and plausibility^7^.

For this study, all queries and analyses were conducted within the Global Collaborative Network, one of several sub-networks from the TriNetX database, and last updated on December 22, 2025. At the time of the data collection in August 2025, this network comprised 166 HCOs and data from over 160 million patients. The study involved a secondary analysis of pre-existing health records and did not include any direct contact, intervention or interaction with patients. All data in TriNetX are de-identified per the de-identification requirements specified in §164.514(a) of the HIPAA Privacy rule. The de-identification procedures were verified through formal determination by a qualified expert as defined in Section §164.514(b)(1). Given that only de-identified statistical summaries are generated and no protected health information is accessible, the Institutional Review Board (IRB) classified this work as exempt from informed consent. This Study followed the Strengthening the Reporting of Observational Studies in Epidemiology (STROBE) reporting guideline.

### Study Population

We included adult patients (≥18 years) who underwent their first kidney transplantation (Index Event) between January 1, 2015, and January 1, 2022. Only patients without any diagnosis of diabetes mellitus were eligible. To ensure comparable follow-up between cohorts, all patients were required to have at least one recorded health care visit at ≥1,181 days post-transplant reflecting a clinically stable cohort without early post-transplant issues.

Patients were excluded if they had any record of SGLT-2i use pre-transplant, or if death, dialysis or any SGLT-2i use occurred within the first 999 days post-transplant. Additional exclusions were applied to patients in whom any outcome occurred prior to the outcome time window. The diagnosis codes (ICD-10), laboratory codes (LOINC), and medication codes (RXNorm) used to define the cohort selection criteria are listed in the supplementary methods appendix.

To establish a landmark cohort of stable transplant recipients, patients were required to survive without any initiation of dialysis or SGLT2 inhibitor exposure during the first 999 days post-transplant. Cohort assignment occurred on day 1000 onward were classified as SGLT2-inhibitor users, whereas patients without SGLT2 inhibitor exposure served as controls.

To ensure follow-up availability, all patients were additionally required to have at least one recorded healthcare encounter ≥1,181 days after transplantation.

Patients with occurrence of the respective outcome before start of follow-up were excluded. The diagnosis codes (ICD-10), laboratory codes (LOINC), and medication codes (RXNorm) used to define the cohort selection criteria are listed in the Supplementary Methods Appendix.

### Exposure and Comparator Groups

The exposed cohort consisted of kidney transplant recipients who received their first SGLT2 inhibitor prescription from day 1,000 onward post-transplant, while the unexposed cohort comprised recipients without any SGLT2 inhibitor prescription between day 1,000 and day 3,006 post-transplant. Day 1,181 post-transplant marked the beginning of the outcome time-window for both groups, which continued until day 3,006 (end of follow-up).

### Outcomes

The primary outcome was the first occurrence of major adverse kidney events (MAKE), defined as a composite of initiation of dialysis or death from any cause. Secondary outcomes included all-cause mortality and major adverse cardiovascular events (MACE), defined as a composite of myocardial infarction, stroke, or death (supplemental table S5). Outcome definitions were based on standardized clinical coding, with the exact terms detailed in the supplementary Methods.

### Covariates and Matching

Baseline variables were measured 365 days prior to the start of the Index Event and included demographic characteristics, comorbidities, laboratory parameters and use of medications. All baseline covariates were included in the propensity score model (supplemental table S6 and S7) for 1:1 nearest-neighbor matching with a caliper of 0.1. The score was generated from 46 predefined covariates. Post-matching balance was evaluated using standardized mean differences (SMDs), with SMD < 0.1 considered indicative of adequate balance.

### Statistical analysis

For each outcome, both cohorts were analyzed using Kaplan-Meier curves and log-rank tests. Adjusted hazard ratios (aHRs) with 95% confidence intervals (CIs) were estimated using Cox proportional hazards regression. The proportional hazards assumption was assessed using Schoenfeld residuals and the Grambsch–Therneau test and was not violated. E-values were calculated to assess the robustness of the observed effects against potential unmeasured confounding^8^. Competing risk analysis for MAKE, MACE and all-cause mortality component endpoints were performed using the Aalen-Johansen method, treating death as a competing event. Specificity analyses assessed individual components of MAKE and MACE. Safety analyses examined adverse events potentially related to SGLT-2i therapy. To evaluate robustness, we conducted positive and negative outcome controls and positive and negative exposure controls. All hypothesis tests were two-sided, with statistical significance defined as p < 0.05.

## Results

From 80,306 non-diabetic kidney transplant recipients between January 1, 2015, and January 1, 2022, we excluded 1,876 patients aged <18 years and 31 patients with documented SGLT-2i use prior to the transplantation (Figure 1). Furthermore, 9,071 patients were excluded due to death, initiation of dialysis, or SGLT-2i treatment within the first 999 days post-transplant. Among the remaining, 878 patients received their first SGLT-2i prescription from day 1,000 post-transplant onwards, while 68,818 patients had no SGLT-2i exposure between day 1,000 and day 3,006 post-transplant. To ensure adequate follow-up, we excluded additional 9 SGLT-2i users and 19,864 non-users without a recorded healthcare visit ≥1,181 days post-transplant. This resulted in 869 exposed and 48,954 unexposed patients eligible for analysis.

**Figure 1.**
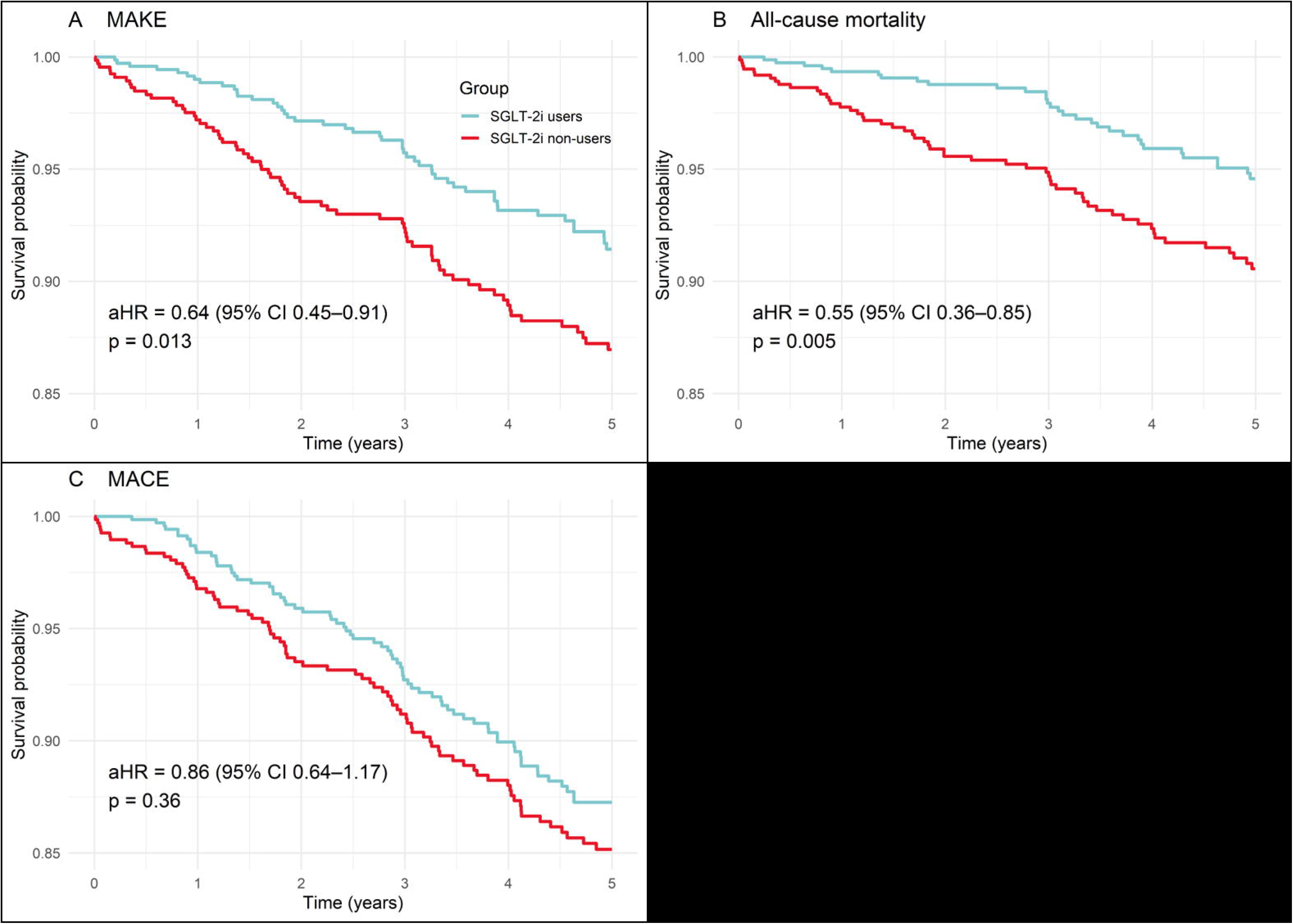
Study cohort selection and propensity score matching worklow. Flow chart of patient selection from the TriNetX Global Collaborative Network and construction of the propensity score-matched cohorts of SGLT2-inhibitor users and non-users.

Propensity score matching yielded 867 matched pairs of SGLT-2i users and non-users with standardized mean differences <0.1 for all baseline covariates, confirming good balance between cohorts (Table 1).

**Table 1.** Baseline characteristics before and after propensity score matching. Baseline demographic characteristics, comorbidities, medication use and laboratory parameters among SGLT2-inhibitor users and non-users before and after propensity score matching (PSM). Covariate balance was assessed using standardized mean differences (SMDs), with an SMD <0.1 indicating adequate balance.

|  | Before PSM |  |  | After PSM |  |  |
| --- | --- | --- | --- | --- | --- | --- |
|  | SGLT2i users | SGLT2i non-users | SMD | SGLT2i users | SGLT2i non-users | SMD |
| N | 869 | 48,954 |  | 867 | 867 |  |
| Demographics |  |  |  |  |  |  |
| Age at index date, mean ± SD | 51.4 ± 14.5 | 48.3 ± 16.4 | 0.19 | 51.3 ± 14.5 | 51.5 ± 15.1 | 0.01 |
| Male, n (%) | 525 (60.41%) | 26,985 (55.12%) | 0.11 | 523 (60.32%) | 517 (59.63%) | 0.01 |
| White, n (%) | 461 (53.04%) | 26,538 (54.21%) | 0.02 | 460 (53.05%) | 481 (55.48%) | 0.04 |
| Not Hispanic or Latino, n (%) | 596 (68.58%) | 29,426 (60.1%) | 0.17 | 594 (68.51%) | 601 (69.31%) | 0.01 |
| Comorbidities, n (%) |  |  |  |  |  |  |
| Ischemic heart diseases | 76 (8.74%) | 2,172 (4.43%) | 0.17 | 74 (8.53%) | 66 (7.61%) | 0.03 |
| Acute myocardial infarction | 14 (1.61%) | 254 (0.51%) | 0.1 | 13 (1.49%) | 10 (1.15%) | 0.03 |
| Diseases of arteries, arterioles and capillaries | 61 (7.02%) | 2,484 (5.07%) | 0.08 | 59 (6.8%) | 58 (6.69%) | <0.01 |
| Essential (primary) hypertension | 320 (36.82%) | 13,427 (27.42%) | 0.2 | 319 (36.79%) | 349 (40.25%) | 0.07 |
| Hypertensive diseases | 397 (45.68%) | 17,047 (34.82%) | 0.22 | 395 (45.55%) | 419 (48.32%) | 0.05 |
| Systolic (congestive) heart failure | 15 (1.72%) | 350 (0.71%) | 0.09 | 15 (1.73%) | 12 (1.38%) | 0.02 |
| Diastolic (congestive) heart failure | 18 (2.07%) | 300 (0.61%) | 0.12 | 16 (1.84%) | 13 (1.49%) | 0.02 |
| Heart failure, unspecified | 36 (4.14%) | 865 (1.76%) | 0.14 | 34 (3.92%) | 29 (3.34%) | 0.03 |
| Pulmonary heart disease and diseases of pulmonary circulation | 37 (4.25%) | 853 (1.74%) | 0.14 | 36 (4.15%) | 35 (4.03%) | <0.01 |
| Cerebrovascular diseases | 22 (2.53%) | 895 (1.82%) | 0.04 | 22 (2.53%) | 16 (1.84%) | 0.04 |
| Acute kidney failure | 63 (7.25%) | 2,623 (5.35%) | 0.07 | 61 (7.03%) | 52 (5.99%) | 0.04 |
| Chronic kidney disease | 388 (44.64%) | 19,079 (38.97%) | 0.11 | 386 (44.52%) | 378 (43.59%) | 0.01 |
| Cystic kidney disease | 44 (5.06%) | 2,614 (5.34%) | 0.01 | 44 (5.07%) | 41 (4.72%) | 0.01 |
| Unspecified kidney failure | 25 (2.87%) | 1,170 (2.39%) | 0.03 | 24 (2.76%) | 25 (2.88%) | <0.01 |
| Proteinuria | 52 (5.98%) | 1,954 (3.99%) | 0.09 | 50 (5.76%) | 51 (5.88%) | <0.01 |
| Urinary tract infection, site not specified | 42 (4.83%) | 2,531 (5.17%) | 0.01 | 42 (4.84%) | 53 (6.11%) | 0.05 |
| Overweight and obesity | 38 (4.37%) | 1,718 (3.5%) | 0.04 | 38 (4.38%) | 30 (3.46%) | 0.04 |
| Disorders of lipoprotein metabolism and other lipidemias | 176 (20.25%) | 6,668 (13.62%) | 0.17 | 176 (20.3%) | 178 (20.53%) | <0.01 |
| Neoplasms | 121 (13.92%) | 4,725 (9.65%) | 0.13 | 119 (13.72%) | 126 (14.53%) | 0.02 |
| Diseases of liver | 45 (5.17%) | 2,202 (4.49%) | 0.03 | 44 (5.07%) | 38 (4.38%) | 0.03 |
| Systemic connective tissue disorders | 48 (5.52%) | 1,536 (3.13%) | 0.11 | 48 (5.53%) | 43 (4.96%) | 0.02 |
| Chronic lower respiratory diseases | 55 (6.32%) | 1,788 (3.65%) | 0.12 | 53 (6.11%) | 50 (5.76%) | 0.01 |
| Medications, n (%) |  |  |  |  |  |  |
| Beta blockers | 295 (33.94%) | 10,520 (21.49%) | 0.28 | 293 (33.79%) | 292 (33.67%) | <0.01 |
| Calcium channel blockers | 226 (26%) | 9,329 (19.05%) | 0.16 | 224 (25.83%) | 223 (25.72%) | <0.01 |
| Antiarrhythmics | 161 (18.52%) | 6,726 (13.73%) | 0.13 | 159 (18.33%) | 146 (16.84%) | 0.03 |
| Diuretics | 201 (23.13%) | 6,996 (14.29%) | 0.22 | 199 (22.95%) | 184 (21.22%) | 0.04 |
| ACE inhibitors | 105 (12.08%) | 3,880 (7.92%) | 0.13 | 104 (11.99%) | 99 (11.41%) | 0.01 |
| Angiotensin II inhibitor | 124 (14.26%) | 3,293 (6.72%) | 0.24 | 122 (11.99%) | 127 (14.64%) | 0.01 |
| Antianginals | 48 (5.52%) | 1,355 (2.76%) | 0.13 | 46 (5.3%) | 46 (5.3%) | <0.01 |
| Antilipemic agents | 216 (24.85%) | 6,510 (13.29%) | 0.29 | 215 (24.79%) | 248 (28.6%) | 0.08 |
| Oral hypoglycemic agents | 10 (1.15%) | 164 (0.33%) | 0.09 | 10 (1.15%) | 11 (1.26%) | 0.01 |
| HMG-CoA reductase inhibitors | 203 (23.36%) | 5,999 (12.25%) | 0.29 | 202 (23.29%) | 233 (26.87%) | 0.08 |
| Laboratory |  |  |  |  |  |  |
| HDL, mg/dL | 47.7 ± 19 | 50.7 ± 19.3 | 0.15 | 47.9 ± 19 | 49.7 ± 18.5 | 0.09 |
| LDL, mg/dL | 91 ± 35.8 | 95.5 ± 38.6 | 0.12 | 90.9 ± 35.9 | 93.1 ± 34.8 | 0.06 |
| Triglyceride, mg/dL | 147 ± 103 | 145 ± 96.9 | 0.01 | 147 ± 103 | 150 ± 121 | 0.02 |
| Hemoglobin, mg/dL | 12 ± 2.09 | 12 ± 2.14 | 0.02 | 12 ± 2.07 | 12.2 ± 2.01 | 0.09 |
| BMI, kg/m<sup>2</sup> | 27.6 ± 5.35 | 26.9 ± 5.69 | 0.12 | 27.6 ± 5.37 | 27.07 ± 5.4 | 0.09 |
| DBP, mmHg | 78.4 ± 13 | 77.4 ± 12.8 | 0.07 | 78.4 ± 13 | 78.4 ± 12.1 | <0.01 |
| SBP, mmHg | 133 ± 20.5 | 131 ± 20.4 | 0.1 | 133 ± 20.3 | 134 ± 18.1 | <0.01 |
| eGFR, mL/min/1.73 m<sup>2</sup> | 36.1 ± 27.6 | 38 ± 30.2 | 0.08 | 36.1 ± 27.5 | 38.7 ± 26.1 | 0.09 |
| Sodium, mmol/L | 139 ± 3.34 | 139 ± 3.3 | <0.01 | 139 ± 3.34 | 139 ± 3.23 | 0.08 |
| Potassium, mmol/L | 4.41 ± 0.63 | 4.36 ± 0.58 | 0.09 | 4.42 ± 0.63 | 4.43 ± 0.56 | 0.01 |

### Outcomes

During the matched follow-up period which had a median duration 3,006 days in both groups, 54 out of 790 patients (6.83%) in the SGLT-2i group experienced a MAKE outcome compared with 76 out of 762 patients (9.97%) in the matched non-user group (Supplemental Table S11). The aHR for MAKE was 0.64 (95% CI: 0.45-0.91), indicating a substantially lower hazard of MAKE among those receiving SGLT-2i therapy. The E-value for the observed aHR was 2.5, with 1.43 for the upper bound of the 95% CI, suggesting the observed association is moderately robust to unmeasured confounding (Figure 2A).

**Figure 2.**
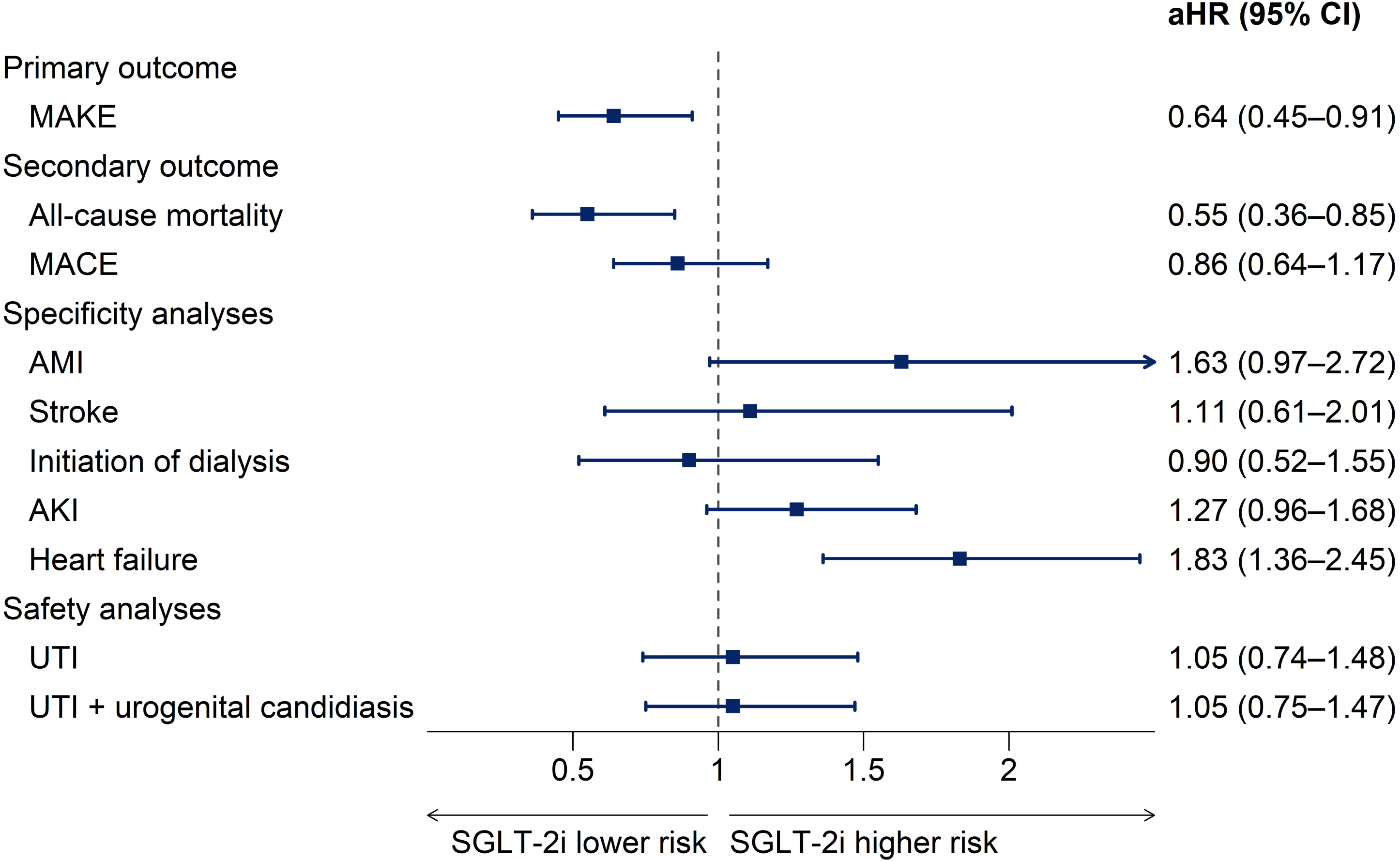
Kaplan-Meier analyses of clinical outcomes. Kaplan-Meier curves comparing SGLT2-inhibitor users and non-users for (A) major adverse kidney events (MAKE), (B) all-cause mortality, and (C) major adverse cardiovascular events (MACE). Adjusted hazard ratios (aHRs) with 95% confidence intervals (CIs) are presented.

Regarding secondary outcomes, all-cause mortality occurred in 35 of 867 patients (4%) in the SGLT-2i group compared with 59 of 867 (6.8%) in the matched control group, corresponding to an aHR of 0.55 (95% CI: 0.36-0.85, log-rank p = 0.0057) and an E-value of 2.62. This demonstrated a persistent reduction in mortality risk with SGLT-2i therapy (Figure 2B).

In contrast, MACE occurred with similar frequency between the groups, with an aHR of 0.86 (95% CI 0.64-1.17) and an E-value of 1.43, showing no statistically significant association with SGLT-2i treatment for MACE in this population (Figure 2C).

Specificity analyses of individual outcome components revealed a more complex picture (Figure 3). Rates of acute myocardial infarction were numerically higher in the SLT-2i group (aHR 1.63, 95% CI: 0.97-2.72), stroke incidence was comparable between groups (aHR 1.11, 95% CI: 0.61-2.01), and incident dialysis was less frequent among treated patients (aHR 0.90, 95% CI 0.52-1.55). There was no statistically significant difference in the risk of acute kidney injury between groups (aHR 1.27, 95% CI: 0.96-1.68), and heart failure was observed more frequently among recipients of SGLT-2i (aHR 1.83, 95% CI 1.36-2.45).

**Figure 3.**
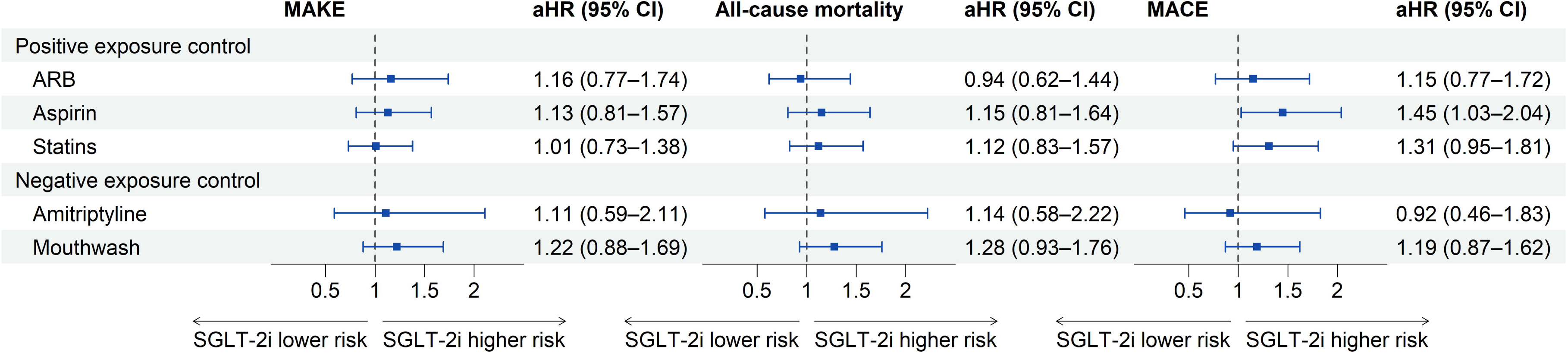
Primary, secondary, specificity and safety outcomes. Forest plot showing aHRs with 95% CIs for primary, secondary, specificity and safety outcomes after PSM. The dashed vertical line represents an aHR of 1.0.

Although the composite endpoint of MAKE was reduced in the SGLT-2i cohort, the individual subcomponents in the sensitivity analyses appeared numerically worse for MAKE and MACE. Competing risk analyses using the Aalen-Johansen method, with death treated as a competing event, demonstrated that these apparent differences were largely attributable to survivorship bias. Patients receiving SGLT-2i lived longer and therefore had a longer time frame during which non-fatal events could occur.

Cumulative incidence at the end of the follow-up window was 4.29% for mortality and 4.25% for MAKE in the SGLT-2i group, compared with 9.79% for mortality and 5.26% for MAKE in non-users. For MACE, cumulative incidence was 12% in the SGLT-2i group versus 5.67% in non-users, again reflecting an increased opportunity for events in the context of longer survival rather than an absolute increase in hazard (Figure 4).

**Figure 4.**
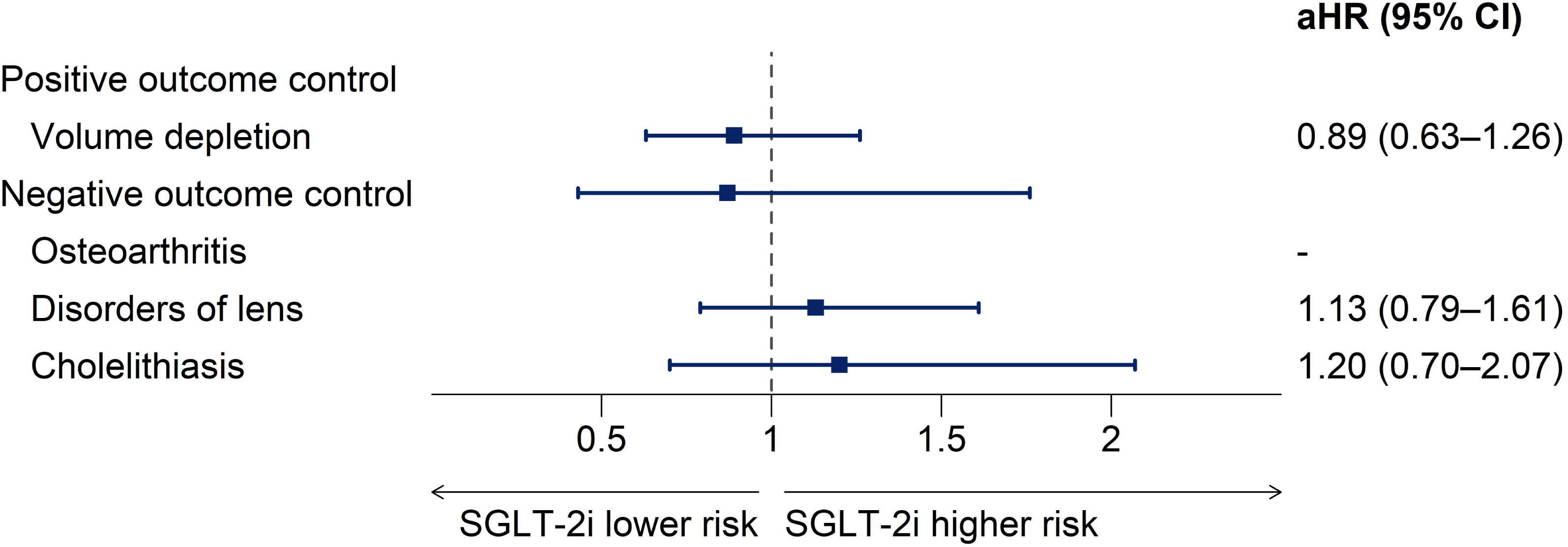
Competing risk analyses. Aalen-Johansen cumulative incidence curves for MAKE, all-cause mortality and MACE among SGLT2-inhibitors users and non-users after PSM. Corresponding cumulative incidence estimates at the end of follow-up are shown.

Safety analyses showed no increase in urinary tract infection (UTI) risk in the SGLT-2i group (aHR 1.05, 95% CI: 0.74-1.48), and combined analyses of UTI with urogenital candidiasis likewise indicated no significant differences between the groups (aHR 1.05, 95% CI: 0.75-1.47) (Figure 3).

### Positive and negative controls

In the positive exposure control analysis, we examined the association of angiotensin receptor blocker (ARB), aspirin, and statin use with the study endpoints (Figure 5).

**Figure 5.**
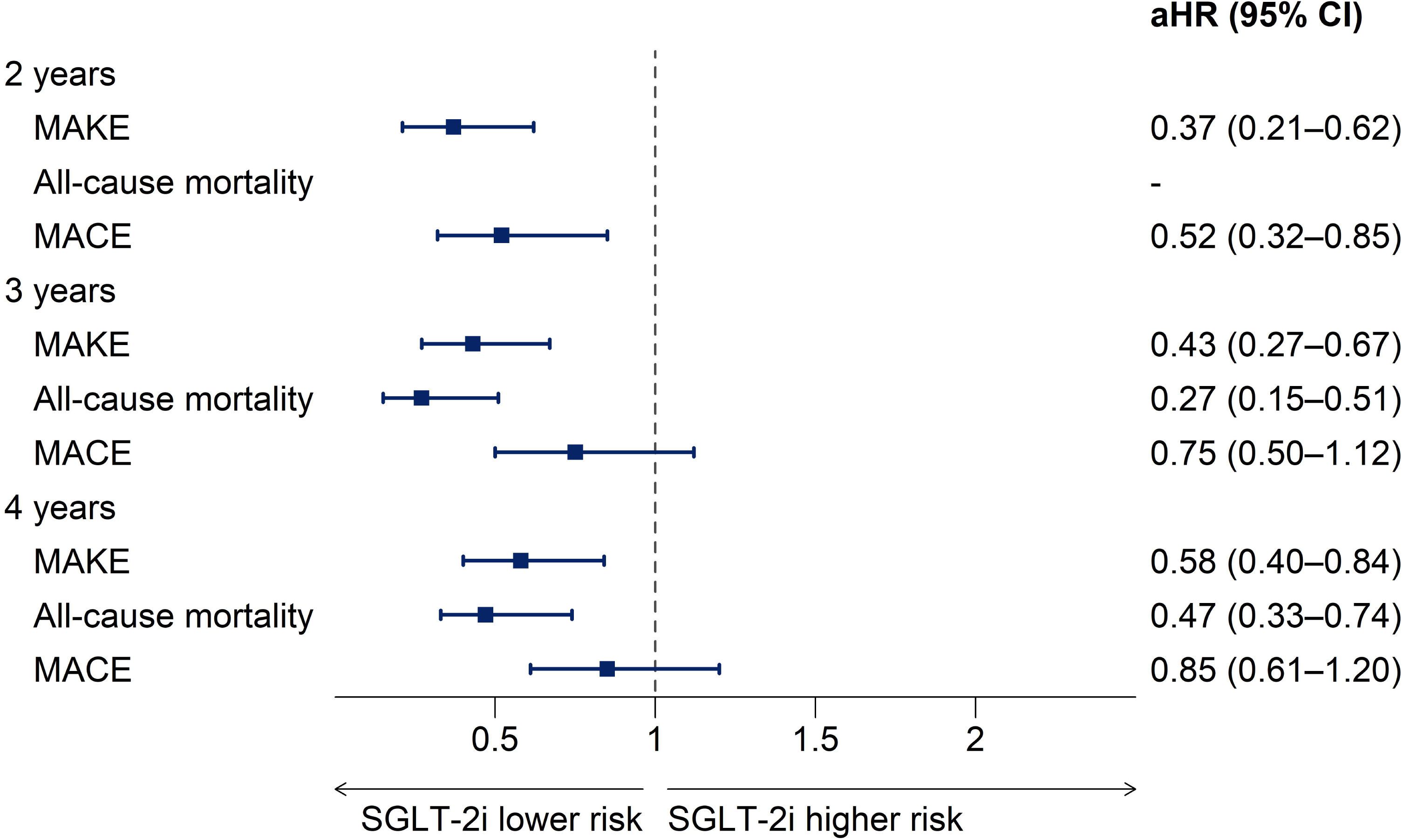
Positive and negative exposure controls. Forest plots of aHRs with 95% CIs for predefined positive and negative exposure controls across MAKE, all-cause mortality and MACE after PSM.

ARB use showed no significant association with MAKE (aHR 1.16, 95% CI: 0.77-1.74), all-cause mortality (aHR 1.15, 95% CI: 0.62-1.44), or MACE (aHR 1.15, 95% CI: 0.77-1.72). Similarly, aspirin use showed no association with MAKE (aHR 1.13, 95% CI: 0.81-1.57) or mortality (aHR 1.15, 95% CI: 0.81-1.64) and a slightly higher risk of MACE in the ARB group, while statin use also yielded no significant results across all endpoints.

In the negative exposure control analyses, amitriptyline and mouthwash consumption were assessed, neither of which demonstrated significant associations with MAKE, mortality, or MACE.

For the positive outcome control, we evaluated volume depletion, which had been linked with SGLT-2i use (Figure 6). This analysis showed no significant association (aHR 0.89, 95% CI: 0.63-1.26). Negative outcome controls, including cholelithiasis, osteoarthritis, and lens disorders, also demonstrated no significant hazard differences between treatment groups, reinforcing the robustness of the analytical framework.

**Figure 6.**
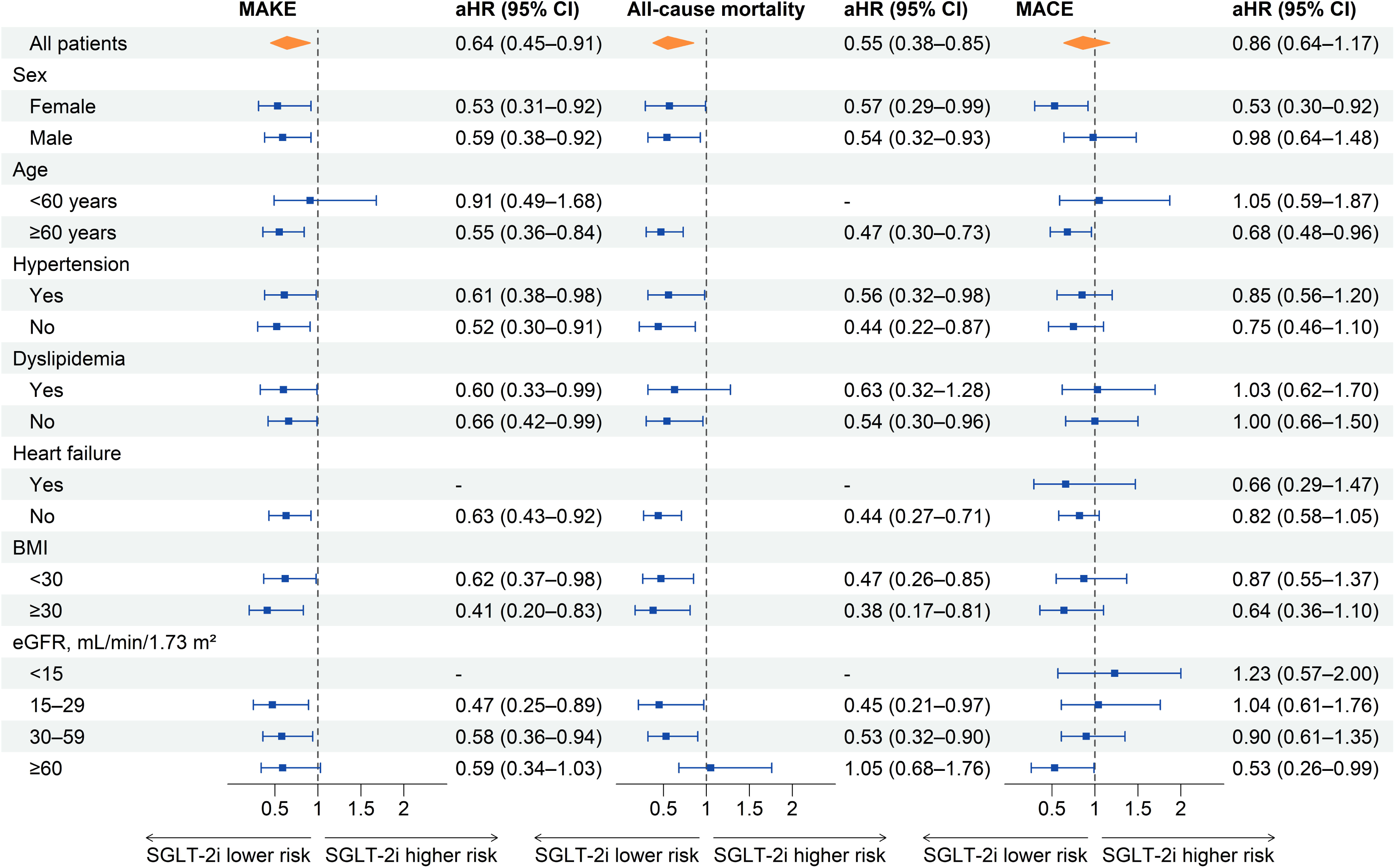
Positive and negative outcome controls. Forest plot of aHRs with 95% CIs for predefined positive and negative outcome controls following PSM.

### Restricted follow-up analyses

In analyses restricted to follow-up periods of 2, 3 and 4 years, the associations with lower risks of MAKE and all-cause mortality remained stable and statistically significant. For MAKE, the aHR at 2 years was 0.37 (95% CI, 0.21-0.62). For all-cause mortality, the aHR at 3 years was 0.27 (95% CI, 0.15-0.51). The association with MACE remained statistically significant at 2 years (aHR 0.52, 95% CI, 0.32-0.85) but was no longer statistically significant with longer follow-up (Figure 7).

**Figure 7.**
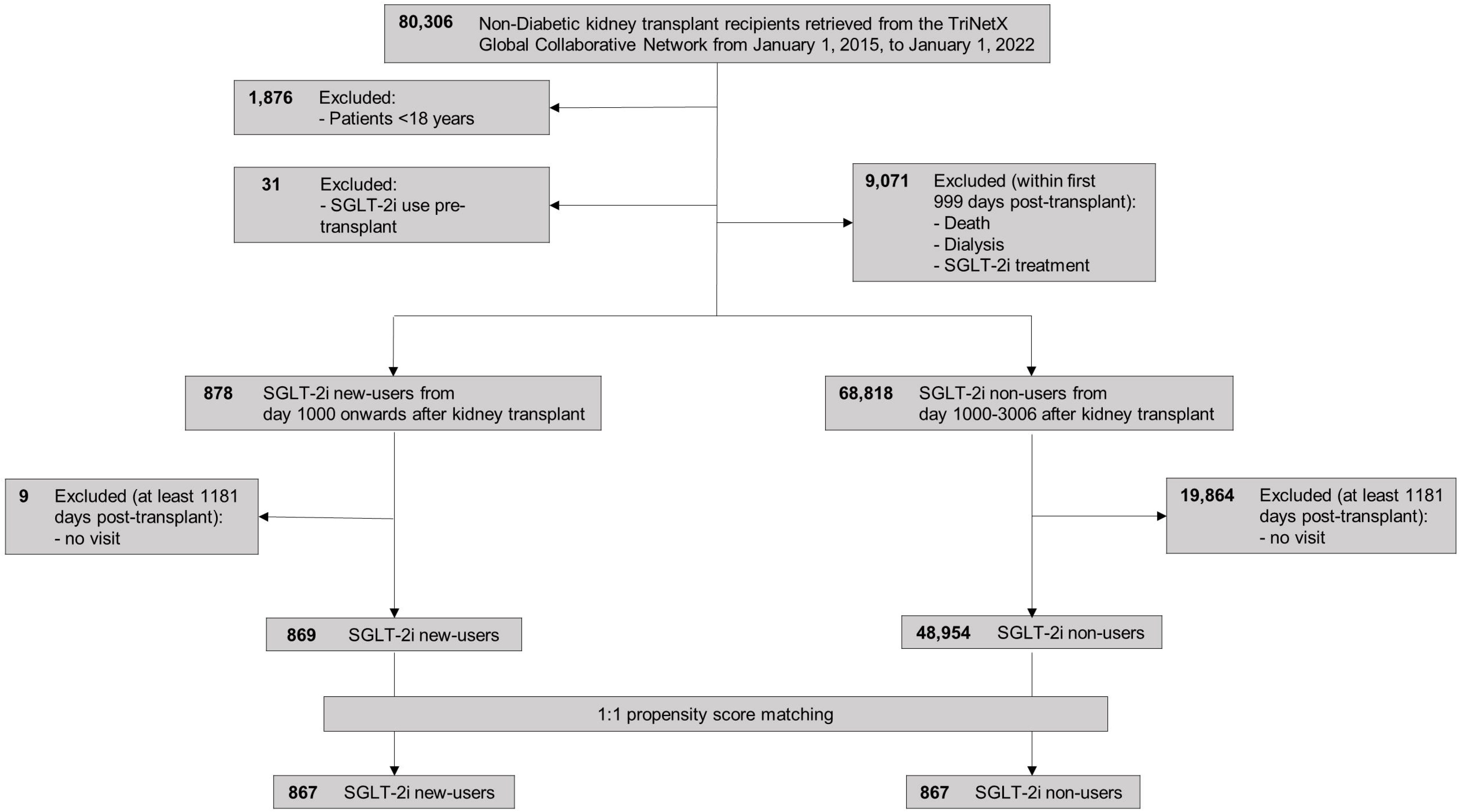
Restricted follow-up analyses. Forest plot of aHRs with 95% CIs for MAKE, all-cause mortality and MACE across restricted follow-up periods of 2, 3, and 4 years after PSM.

### Subgroup analysis

We performed prespecified subgroup analyses to explore potential effect modification across baseline demographic and clinical variables (Figure 8). The renal and survival benefits of SGLT-2i use was broadly consistent across most demographic and clinical variables, but appeared more pronounced in certain patient groups. Stronger relative risk reductions were seen in individuals aged ≥ 60 years, those with a higher BMI of ≥ 30, and recipients with an eGFR of 15-29 mL/min/1.73 m^2^. In contrast, younger patients (<60 years) and in those with a higher eGFR ≥ 60, no statistically significant benefit for MAKE was observed.

**Figure 8.**
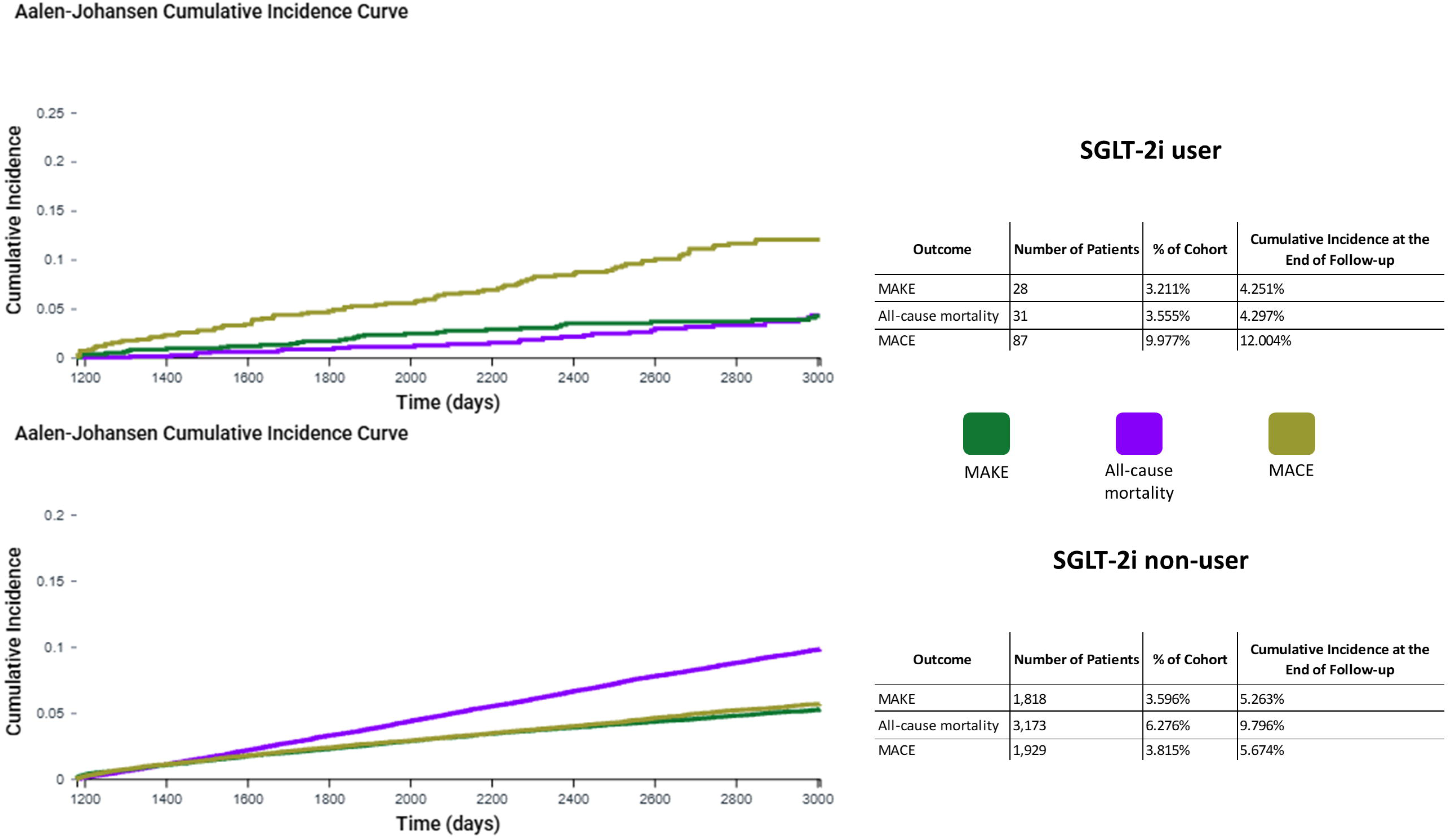
Subgroup analyses. Forest plots of aHRs with 95% CIs for MAKE, all-cause mortality and MACE across predefined demographic and clinical subgroups after PSM.

For all-cause mortality, the hazard reduction was consistent across almost all subgroups, with significant effects in both sexes, patients with and without hypertension and across BMI and eGFR categories <60. Only patients without dyslipidemia had a significant lower risk of mortality outcomes.

In contrast to both MAKE and mortality outcomes, hazard reductions for MACE were less consistent. Benefit was mainly evident in females, older patients, and those with a renal function ≥ 60. Across other subgroups, point estimates did not reach statistical significance.

## Discussion

The present reallZIworld analysis based on propensity scorelZImatched cohorts demonstrates that SGLT2 inhibitor use in nonlZIdiabetic long term kidney transplant recipients is associated with a significant reduction in MAKE and alllZIcause mortality, while no consistent benefit on MACE could be shown. The reduction in MAKE was substantial, with an adjusted hazard ratio (aHR) of 0.64 and robust ElZIvalues, and alllZIcause mortality was markedly lower with an aHR of 0.55. At the same time, we did not observe an increased risk of genitourinary infections or other relevant safety signals, supporting a favourable benefit–risk profile of SGLT2 inhibitors in this specific population.

These findings are broadly consistent with large randomized controlled trials of SGLT2 inhibitors in patients with CKD, in which meaningful reductions in kidney disease progression and, in part, mortality were observed in both diabetic and nonlZIdiabetic populations^9,10^. In DAPAlZICKD and EMPAlZIKIDNEY, SGLT2 inhibition reduced the risk of CKD progression and cardiovascular death across heterogeneous CKD cohorts, independent of diabetes status. Our study extends this evidence by showing comparable renal and survival benefits in a highly selected group of nonlZIdiabetic kidney transplant recipients, who were systematically excluded from the landmark CKD trials.

Our results also align with and complement the growing body of evidence on SGLT2 inhibitor use in diabetic kidney transplant recipients but go beyond this existing literature^11,12^. Previous reallZIworld and registrylZIbased analyses in diabetic transplant recipients have reported lower risks of mortality, MACE and MAKE with SGLT2 inhibitor therapy and have triggered discussions about their routine use in this population. Our data indicate that similar, and potentially even more pronounced, relative benefits on renal endpoints and mortality may be achievable in nonlZIdiabetic transplant recipients, suggesting that the protective effects of SGLT2 inhibitors extend beyond glucoselZIlowering and are clinically relevant in purely nonlZIdiabetic transplant populations.

The observed reduction in alllZIcause mortality is of particular clinical importance, given that cardiovascular events and infections remain leading causes of death in kidney transplant recipients^13,14^ and are only partly mitigated by standard preventive strategies. In line with data from CKD trials and studies in diabetic transplant recipients, in which SGLT2 inhibitors were likewise associated with lower mortality, the additional survival benefit seen in our cohort could contribute meaningfully to improving longlZIterm outcomes after transplantation. The fact that mortality in the control group was approximately twice as high as in SGLT2 inhibitor users underscores the potential clinical impact of implementing this therapy more broadly in eligible nonlZIdiabetic recipients.

A central methodological message of our work is the importance of competing risk analysis for correctly interpreting renal and cardiovascular endpoints in this context. While sensitivity analyses of individual nonlZIfatal outcomes showed numerically higher event rates in the SGLT2 inhibitor group, Aalen–Johansen competing risk analyses^15^ indicated that these patterns were largely driven by survivorship bias, as patients with longer survival inherently have more time to experience nonlZIfatal events. Only by treating death as a competing event did it become evident that the cumulative incidence of MAKE and alllZIcause mortality was actually lower among SGLT2 inhibitor users, and that the apparent increase in some component endpoints mainly reflects differential survival rather than an increased intrinsic hazard.

Several limitations must be acknowledged. First, this is a retrospective, reallZIworld analysis based on electronic health records and, despite extensive propensity score matching, remains susceptible to residual confounding, including confounding by indication. Second, the number of exposed patients is relatively modest, even within a very large source population, which limits the precision of subgroup and safety analyses. Third, competing risk analyses, although essential in this setting, are methodologically complex, and interpretation of individual component outcomes—particularly in the presence of different survival times between groups—must remain cautious and hypothesislZIgenerating.

Despite these limitations, our findings strongly suggest that SGLT2 inhibitors have considerable potential to improve longlZIterm renal outcomes and survival in nonlZIdiabetic kidney transplant recipients and may become an important component of adjunctive therapy alongside maintenance immunosuppression. If confirmed by prospective trials, these results could help drive a paradigm shift in postlZItransplant longlZIterm care, expanding current practice—focused primarily on blood pressure control, lipid management and RAAS blockade—to systematically include SGLT2 inhibition as a standard therapeutic option in appropriate nonlZIdiabetic kidney transplant recipients.

## Data availability statement

The data that support the findings of this study are available from TriNetX, LLC, but restrictions apply to the availability of these data, which were used under license for the current study and are therefore not publicly available.

## Use of artificial intelligence (AI) tools statement

During the preparation of this work, ChatGPT (OpenAI) was used to assist with programming code for data visualization and figure generation, and Gemini (Google) to support language editing and improve readability. All generated content was reviewed, edited, and verified by the authors, who take full responsibility for the final manuscript.

## Abbreviations

ACEi: angiotensin-converting-enzyme inhibitors
ARB: angiotensin receptor blockers
CKD: chronic kidney disease
eGFR: estimated glomerular filtration rate
ESRD: end-stage renal disease
HCOs: healthcare organizations
RAAS: renin-angiotensin-aldosterone system
RAASi: renin-angiotensin-aldosterone system inhibitors
SGLT-2i: sodium glucose cotransporter-2 inhibitors
T2DM: type 2 diabetes mellitus

