## Supplementary Appendix for "Clinical effectiveness of SGLT2 inhibitors in non-diabetic kidney transplanted patients- a real world data analysis"

[**Table S45. Code definitions and selection criteria for SGLT2i users – <30 BMI.**	116](#_Toc230180108)

[**Table S46. Code definitions and selection criteria for SGLT2i non-users – <30 BMI.**	119](#_Toc230180109)

[**Table S49. Code definitions and selection criteria for SGLT2i users – <15 eGFR.**	128](#_Toc230180112)

[**Table S50. Code definitions and selection criteria for SGLT2i non-users – <15 eGFR.**	131](#_Toc230180113)

### 1. **Introduction**

TriNetX is the global federated health research network providing access to electronic medical records (diagnoses, procedures, medications, laboratory values, genomic information) across large healthcare organizations (HCOs). This report was run on the set of HCOs grouped into a network called Global Collaborative Network. This network included 170 HCO(s).

The analysis process includes two main steps:

1) Defining the cohorts through query criteria;

2) Setting up and running the analysis.

Setting up the analysis requires definitions for the index event, outcomes criteria, and the time frame. Compare outcomes supports four analyses: Measures of Association, Survival, Number of Instances and Lab result distribution. These analyses have additional options that are listed in the Outcomes Definitions and Analyses Specifications section below. Furthermore, characteristics of the cohorts that are balanced using propensity score matching are also included in the Propensity Score Matching section.

##### **Table S1. Code definitions and selection criteria for identification for SGLT2i users.**

| Ungrouped terms | | | | | |
| --- | --- | --- | --- | --- | --- |
|  | must have |  | demographics | Age | Age (at least 18 years (most recent occurrence)) |
|  | cannot have |  | diagnosis | UMLS:ICD10CM:E08-E13 | Diabetes mellitus |
| Group 1 | | | | | |
|  | **Kidney transplant** | | | | |
|  | must have | any of | diagnosis | UMLS:ICD10CM:Z94.0 | Kidney transplant status |
|  |  |  | procedure | UMLS:ICD10PCS:0TY00Z0 | Transplantation of Right Kidney, Allogeneic, Open Approach |
|  |  |  | procedure | UMLS:ICD10PCS:0TY10Z0 | Transplantation of Left Kidney, Allogeneic, Open Approach |
|  |  |  | procedure | UMLS:ICD10PCS:0TY10Z1 | Transplantation of Left Kidney, Syngeneic, Open Approach |
|  |  |  | procedure | UMLS:ICD10PCS:0TY10Z2 | Transplantation of Left Kidney, Zooplastic, Open Approach |
|  |  |  | procedure | UMLS:ICD10PCS:0TY00Z1 | Transplantation of Right Kidney, Syngeneic, Open Approach |
|  |  |  | procedure | UMLS:ICD10PCS:0TY00Z2 | Transplantation of Right Kidney, Zooplastic, Open Approach |
|  |  |  | procedure | UMLS:ICD10PCS:0TY0 | Urinary System / Transplantation / Kidney, Right |
|  |  |  | procedure | UMLS:ICD10PCS:0TY1 | Urinary System / Transplantation / Kidney, Left |
|  | date constraint | | The terms in this group occurred between Jan 1, 2015 and Jan 1, 2022 | | |
| Group 2 | | | | | |
|  | **Group 2A Kidney transplant** | | | | |
|  | must have | any of | diagnosis | UMLS:ICD10CM:Z94.0 | Kidney transplant status |
|  |  |  | procedure | UMLS:ICD10PCS:0TY00Z0 | Transplantation of Right Kidney, Allogeneic, Open Approach |
|  |  |  | procedure | UMLS:ICD10PCS:0TY10Z0 | Transplantation of Left Kidney, Allogeneic, Open Approach |
|  |  |  | procedure | UMLS:ICD10PCS:0TY10Z1 | Transplantation of Left Kidney, Syngeneic, Open Approach |
|  |  |  | procedure | UMLS:ICD10PCS:0TY10Z2 | Transplantation of Left Kidney, Zooplastic, Open Approach |
|  |  |  | procedure | UMLS:ICD10PCS:0TY00Z1 | Transplantation of Right Kidney, Syngeneic, Open Approach |
|  |  |  | procedure | UMLS:ICD10PCS:0TY00Z2 | Transplantation of Right Kidney, Zooplastic, Open Approach |
|  |  |  | procedure | UMLS:ICD10PCS:0TY0 | Urinary System / Transplantation / Kidney, Right |
|  |  |  | procedure | UMLS:ICD10PCS:0TY1 | Urinary System / Transplantation / Kidney, Left |
|  | date constraint | | The terms in this group occurred at any time | | |
|  | event relationship | | Any instance of noDialysis, Death, SGLT2i occurred within 1 day and 999 days after the first instance of Kidney transplant | | |
|  | **Group 2B noDialysis, Death, SGLT2i** | | | | |
|  | cannot have |  | procedure | UMLS:CPT:1012740 | Dialysis Services and Procedures |
|  |  | or | procedure | UMLS:CPT:90945 | Dialysis procedure other than hemodialysis (eg, peritoneal dialysis, hemofiltration, or other continuous renal replacement therapies), with single evaluation by a physician or other qualified health care professional |
|  |  | or | procedure | UMLS:CPT:90947 | Dialysis procedure other than hemodialysis (eg, peritoneal dialysis, hemofiltration, or other continuous renal replacement therapies) requiring repeated evaluations by a physician or other qualified health care professional, with or without substantial revision of dialysis prescription |
|  |  | or | procedure | UMLS:CPT:1012752 | Hemodialysis Procedures |
|  |  | or | procedure | UMLS:SNOMED:302497006 | Hemodialysis |
|  |  | or | procedure | UMLS:CPT:90937 | Hemodialysis procedure requiring repeated evaluation(s) with or without substantial revision of dialysis prescription |
|  |  | or | procedure | UMLS:CPT:90935 | Hemodialysis procedure with single evaluation by a physician or other qualified health care professional |
|  |  | or | demographics | Deceased | Deceased |
|  |  | or | diagnosis | UMLS:ICD10CM:R99 | Ill-defined and unknown cause of mortality |
|  |  | or | medication | NLM:ATC:A10BK | Sodium-glucose co-transporter 2 (SGLT2) inhibitors |
| Group 3 | | | | | |
|  | **Group 3A Kidney transplant** | | | | |
|  | must have | any of | diagnosis | UMLS:ICD10CM:Z94.0 | Kidney transplant status |
|  |  |  | procedure | UMLS:ICD10PCS:0TY00Z0 | Transplantation of Right Kidney, Allogeneic, Open Approach |
|  |  |  | procedure | UMLS:ICD10PCS:0TY10Z0 | Transplantation of Left Kidney, Allogeneic, Open Approach |
|  |  |  | procedure | UMLS:ICD10PCS:0TY10Z1 | Transplantation of Left Kidney, Syngeneic, Open Approach |
|  |  |  | procedure | UMLS:ICD10PCS:0TY10Z2 | Transplantation of Left Kidney, Zooplastic, Open Approach |
|  |  |  | procedure | UMLS:ICD10PCS:0TY00Z1 | Transplantation of Right Kidney, Syngeneic, Open Approach |
|  |  |  | procedure | UMLS:ICD10PCS:0TY00Z2 | Transplantation of Right Kidney, Zooplastic, Open Approach |
|  |  |  | procedure | UMLS:ICD10PCS:0TY0 | Urinary System / Transplantation / Kidney, Right |
|  |  |  | procedure | UMLS:ICD10PCS:0TY1 | Urinary System / Transplantation / Kidney, Left |
|  | date constraint | | The terms in this group occurred at any time | | |
|  | event relationship | | Any instance of SGLT2i occurred at least 1000 days after the first instance of Kidney transplant | | |
|  | **Group 3B SGLT2i** | | | | |
|  | must have |  | medication | NLM:ATC:A10BK | Sodium-glucose co-transporter 2 (SGLT2) inhibitors |
| Group 4 | | | | | |
|  | **Group 4A Kidney transplant** | | | | |
|  | must have | any of | diagnosis | UMLS:ICD10CM:Z94.0 | Kidney transplant status |
|  |  |  | procedure | UMLS:ICD10PCS:0TY00Z0 | Transplantation of Right Kidney, Allogeneic, Open Approach |
|  |  |  | procedure | UMLS:ICD10PCS:0TY10Z0 | Transplantation of Left Kidney, Allogeneic, Open Approach |
|  |  |  | procedure | UMLS:ICD10PCS:0TY10Z1 | Transplantation of Left Kidney, Syngeneic, Open Approach |
|  |  |  | procedure | UMLS:ICD10PCS:0TY10Z2 | Transplantation of Left Kidney, Zooplastic, Open Approach |
|  |  |  | procedure | UMLS:ICD10PCS:0TY00Z1 | Transplantation of Right Kidney, Syngeneic, Open Approach |
|  |  |  | procedure | UMLS:ICD10PCS:0TY00Z2 | Transplantation of Right Kidney, Zooplastic, Open Approach |
|  |  |  | procedure | UMLS:ICD10PCS:0TY0 | Urinary System / Transplantation / Kidney, Right |
|  |  |  | procedure | UMLS:ICD10PCS:0TY1 | Urinary System / Transplantation / Kidney, Left |
|  | date constraint | | The terms in this group occurred at any time | | |
|  | event relationship | | Any instance of noSGLT2i occurred at least 1 day before the first instance of Kidney transplant | | |
|  | **Group 4B noSGLT2i** | | | | |
|  | cannot have |  | medication | NLM:ATC:A10BK | Sodium-glucose co-transporter 2 (SGLT2) inhibitors |
| Group 5 | | | | | |
|  | **Group 5A Kidney transplant** | | | | |
|  | must have | any of | diagnosis | UMLS:ICD10CM:Z94.0 | Kidney transplant status |
|  |  |  | procedure | UMLS:ICD10PCS:0TY00Z0 | Transplantation of Right Kidney, Allogeneic, Open Approach |
|  |  |  | procedure | UMLS:ICD10PCS:0TY10Z0 | Transplantation of Left Kidney, Allogeneic, Open Approach |
|  |  |  | procedure | UMLS:ICD10PCS:0TY10Z1 | Transplantation of Left Kidney, Syngeneic, Open Approach |
|  |  |  | procedure | UMLS:ICD10PCS:0TY10Z2 | Transplantation of Left Kidney, Zooplastic, Open Approach |
|  |  |  | procedure | UMLS:ICD10PCS:0TY00Z1 | Transplantation of Right Kidney, Syngeneic, Open Approach |
|  |  |  | procedure | UMLS:ICD10PCS:0TY00Z2 | Transplantation of Right Kidney, Zooplastic, Open Approach |
|  |  |  | procedure | UMLS:ICD10PCS:0TY0 | Urinary System / Transplantation / Kidney, Right |
|  |  |  | procedure | UMLS:ICD10PCS:0TY1 | Urinary System / Transplantation / Kidney, Left |
|  | date constraint | | The terms in this group occurred at any time | | |
|  | event relationship | | Any instance of Visit occurred at least 1181 days after the first instance of Kidney transplant | | |
|  | **Group 5B Visit** | | | | |
|  | must have |  | visit | TNX:Visit | Visit |

##### **Table S2. Code definitions and selection criteria for identification for SGLT2i non-users.**

| Ungrouped terms | | | | | |
| --- | --- | --- | --- | --- | --- |
|  | must have |  | demographics | Age | Age (at least 18 years (most recent occurrence)) |
|  | cannot have |  | diagnosis | UMLS:ICD10CM:E08-E13 | Diabetes mellitus |
| Group 1 | | | | | |
|  | **Kidney transplant** | | | | |
|  | must have | any of | diagnosis | UMLS:ICD10CM:Z94.0 | Kidney transplant status |
|  |  |  | procedure | UMLS:ICD10PCS:0TY00Z0 | Transplantation of Right Kidney, Allogeneic, Open Approach |
|  |  |  | procedure | UMLS:ICD10PCS:0TY10Z0 | Transplantation of Left Kidney, Allogeneic, Open Approach |
|  |  |  | procedure | UMLS:ICD10PCS:0TY10Z1 | Transplantation of Left Kidney, Syngeneic, Open Approach |
|  |  |  | procedure | UMLS:ICD10PCS:0TY10Z2 | Transplantation of Left Kidney, Zooplastic, Open Approach |
|  |  |  | procedure | UMLS:ICD10PCS:0TY00Z1 | Transplantation of Right Kidney, Syngeneic, Open Approach |
|  |  |  | procedure | UMLS:ICD10PCS:0TY00Z2 | Transplantation of Right Kidney, Zooplastic, Open Approach |
|  |  |  | procedure | UMLS:ICD10PCS:0TY0 | Urinary System / Transplantation / Kidney, Right |
|  |  |  | procedure | UMLS:ICD10PCS:0TY1 | Urinary System / Transplantation / Kidney, Left |
|  | date constraint | | The terms in this group occurred between Jan 1, 2015 and Jan 1, 2022 | | |
| Group 2 | | | | | |
|  | **Group 2A Kidney transplant** | | | | |
|  | must have | any of | diagnosis | UMLS:ICD10CM:Z94.0 | Kidney transplant status |
|  |  |  | procedure | UMLS:ICD10PCS:0TY00Z0 | Transplantation of Right Kidney, Allogeneic, Open Approach |
|  |  |  | procedure | UMLS:ICD10PCS:0TY10Z0 | Transplantation of Left Kidney, Allogeneic, Open Approach |
|  |  |  | procedure | UMLS:ICD10PCS:0TY10Z1 | Transplantation of Left Kidney, Syngeneic, Open Approach |
|  |  |  | procedure | UMLS:ICD10PCS:0TY10Z2 | Transplantation of Left Kidney, Zooplastic, Open Approach |
|  |  |  | procedure | UMLS:ICD10PCS:0TY00Z1 | Transplantation of Right Kidney, Syngeneic, Open Approach |
|  |  |  | procedure | UMLS:ICD10PCS:0TY00Z2 | Transplantation of Right Kidney, Zooplastic, Open Approach |
|  |  |  | procedure | UMLS:ICD10PCS:0TY0 | Urinary System / Transplantation / Kidney, Right |
|  |  |  | procedure | UMLS:ICD10PCS:0TY1 | Urinary System / Transplantation / Kidney, Left |
|  | date constraint | | The terms in this group occurred at any time | | |
|  | event relationship | | Any instance of noDialysis, Death, SGLT2i occurred within 1 day and 999 days after the first instance of Kidney transplant | | |
|  | **Group 2B noDialysis, Death, SGLT2i** | | | | |
|  | cannot have |  | procedure | UMLS:CPT:1012740 | Dialysis Services and Procedures |
|  |  | or | procedure | UMLS:CPT:90945 | Dialysis procedure other than hemodialysis (eg, peritoneal dialysis, hemofiltration, or other continuous renal replacement therapies), with single evaluation by a physician or other qualified health care professional |
|  |  | or | procedure | UMLS:CPT:90947 | Dialysis procedure other than hemodialysis (eg, peritoneal dialysis, hemofiltration, or other continuous renal replacement therapies) requiring repeated evaluations by a physician or other qualified health care professional, with or without substantial revision of dialysis prescription |
|  |  | or | procedure | UMLS:CPT:1012752 | Hemodialysis Procedures |
|  |  | or | procedure | UMLS:SNOMED:302497006 | Hemodialysis |
|  |  | or | procedure | UMLS:CPT:90937 | Hemodialysis procedure requiring repeated evaluation(s) with or without substantial revision of dialysis prescription |
|  |  | or | procedure | UMLS:CPT:90935 | Hemodialysis procedure with single evaluation by a physician or other qualified health care professional |
|  |  | or | demographics | Deceased | Deceased |
|  |  | or | diagnosis | UMLS:ICD10CM:R99 | Ill-defined and unknown cause of mortality |
|  |  | or | medication | NLM:ATC:A10BK | Sodium-glucose co-transporter 2 (SGLT2) inhibitors |
| Group 3 | | | | | |
|  | **Group 3A Kidney transplant** | | | | |
|  | must have | any of | diagnosis | UMLS:ICD10CM:Z94.0 | Kidney transplant status |
|  |  |  | procedure | UMLS:ICD10PCS:0TY00Z0 | Transplantation of Right Kidney, Allogeneic, Open Approach |
|  |  |  | procedure | UMLS:ICD10PCS:0TY10Z0 | Transplantation of Left Kidney, Allogeneic, Open Approach |
|  |  |  | procedure | UMLS:ICD10PCS:0TY10Z1 | Transplantation of Left Kidney, Syngeneic, Open Approach |
|  |  |  | procedure | UMLS:ICD10PCS:0TY10Z2 | Transplantation of Left Kidney, Zooplastic, Open Approach |
|  |  |  | procedure | UMLS:ICD10PCS:0TY00Z1 | Transplantation of Right Kidney, Syngeneic, Open Approach |
|  |  |  | procedure | UMLS:ICD10PCS:0TY00Z2 | Transplantation of Right Kidney, Zooplastic, Open Approach |
|  |  |  | procedure | UMLS:ICD10PCS:0TY0 | Urinary System / Transplantation / Kidney, Right |
|  |  |  | procedure | UMLS:ICD10PCS:0TY1 | Urinary System / Transplantation / Kidney, Left |
|  | date constraint | | The terms in this group occurred at any time | | |
|  | event relationship | | Any instance of noSGLT2i occurred within 1000 days and 3006 days after the first instance of Kidney transplant | | |
|  | **Group 3B noSGLT2i** | | | | |
|  | cannot have |  | medication | NLM:ATC:A10BK | Sodium-glucose co-transporter 2 (SGLT2) inhibitors |
| Group 4 | | | | | |
|  | **Group 4A Kidney transplant** | | | | |
|  | must have | any of | diagnosis | UMLS:ICD10CM:Z94.0 | Kidney transplant status |
|  |  |  | procedure | UMLS:ICD10PCS:0TY00Z0 | Transplantation of Right Kidney, Allogeneic, Open Approach |
|  |  |  | procedure | UMLS:ICD10PCS:0TY10Z0 | Transplantation of Left Kidney, Allogeneic, Open Approach |
|  |  |  | procedure | UMLS:ICD10PCS:0TY10Z1 | Transplantation of Left Kidney, Syngeneic, Open Approach |
|  |  |  | procedure | UMLS:ICD10PCS:0TY10Z2 | Transplantation of Left Kidney, Zooplastic, Open Approach |
|  |  |  | procedure | UMLS:ICD10PCS:0TY00Z1 | Transplantation of Right Kidney, Syngeneic, Open Approach |
|  |  |  | procedure | UMLS:ICD10PCS:0TY00Z2 | Transplantation of Right Kidney, Zooplastic, Open Approach |
|  |  |  | procedure | UMLS:ICD10PCS:0TY0 | Urinary System / Transplantation / Kidney, Right |
|  |  |  | procedure | UMLS:ICD10PCS:0TY1 | Urinary System / Transplantation / Kidney, Left |
|  | date constraint | | The terms in this group occurred at any time | | |
|  | event relationship | | Any instance of noSGLT2i occurred at least 1 day before the first instance of Kidney transplant | | |
|  | **Group 4B noSGLT2i** | | | | |
|  | cannot have |  | medication | NLM:ATC:A10BK | Sodium-glucose co-transporter 2 (SGLT2) inhibitors |
| Group 5 | | | | | |
|  | **Group 5A Kidney transplant** | | | | |
|  | must have | any of | diagnosis | UMLS:ICD10CM:Z94.0 | Kidney transplant status |
|  |  |  | procedure | UMLS:ICD10PCS:0TY00Z0 | Transplantation of Right Kidney, Allogeneic, Open Approach |
|  |  |  | procedure | UMLS:ICD10PCS:0TY10Z0 | Transplantation of Left Kidney, Allogeneic, Open Approach |
|  |  |  | procedure | UMLS:ICD10PCS:0TY10Z1 | Transplantation of Left Kidney, Syngeneic, Open Approach |
|  |  |  | procedure | UMLS:ICD10PCS:0TY10Z2 | Transplantation of Left Kidney, Zooplastic, Open Approach |
|  |  |  | procedure | UMLS:ICD10PCS:0TY00Z1 | Transplantation of Right Kidney, Syngeneic, Open Approach |
|  |  |  | procedure | UMLS:ICD10PCS:0TY00Z2 | Transplantation of Right Kidney, Zooplastic, Open Approach |
|  |  |  | procedure | UMLS:ICD10PCS:0TY0 | Urinary System / Transplantation / Kidney, Right |
|  |  |  | procedure | UMLS:ICD10PCS:0TY1 | Urinary System / Transplantation / Kidney, Left |
|  | date constraint | | The terms in this group occurred at any time | | |
|  | event relationship | | Any instance of Visit occurred at least 1181 days after the first instance of Kidney transplant | | |
|  | **Group 5B Visit** | | | | |
|  | must have |  | visit | TNX:Visit | Visit |

### 2. **Analysis Setup**

Dieser This section contains the Index Event and Time Window definitions and a list of selected outcomes and the analyses.

#### Index Event & Time Window Definitions

The index event defines the point in time when each patient in the cohort enters the analysis. To define an index event for the cohort, one or more criteria for the cohort must be selected. The index date for each patient within a cohort is the day on which the patient first met the selected criteria for the cohort (listed in the table below).

As the index event defines the earliest time point after which outcomes are analyzed, the time window defines the duration during which outcomes are analyzed. The time window can start on the same day as the index event or at any specified time interval after the index event. The time window can end any time after the start date. Outcomes are defined as diagnoses, medications, procedures, or laboratory values that happened in the time window starting after the first occurrence of the index event.

#### Time Window Used in this Analysis

This analysis included outcomes that occurred in the time window that started 1 day after the first occurrence of the index event and ended 1825 days after the first occurrence of the index event.

The index event only includes events that occurred up to 20 years ago. Patients whose index event occurred 20 years or more ago are excluded. In this analysis, 0 patients in Cohort 1 and 0 patients in Cohort 2 were excluded because they met the index event more than 20 years ago.

#### Index Events Used in this Analysis

Index events for the Compare Outcomes analysis were derived from the cohort definitions. Index events were defined separately for each cohort and were based on the criteria used in the original cohort definition. Please see Appendix B for the text representation of the index event definition.

##### **Table S3. Definition of index event in the SGLT2i user cohort.**

| Group 1 | | | | | |
| --- | --- | --- | --- | --- | --- |
|  | **Kidney transplant** | | | | |
|  | must have | any of | diagnosis | UMLS:ICD10CM:Z94.0 | Kidney transplant status |
|  |  |  | procedure | UMLS:ICD10PCS:0TY00Z0 | Transplantation of Right Kidney, Allogeneic, Open Approach |
|  |  |  | procedure | UMLS:ICD10PCS:0TY10Z0 | Transplantation of Left Kidney, Allogeneic, Open Approach |
|  |  |  | procedure | UMLS:ICD10PCS:0TY10Z1 | Transplantation of Left Kidney, Syngeneic, Open Approach |
|  |  |  | procedure | UMLS:ICD10PCS:0TY10Z2 | Transplantation of Left Kidney, Zooplastic, Open Approach |
|  |  |  | procedure | UMLS:ICD10PCS:0TY00Z1 | Transplantation of Right Kidney, Syngeneic, Open Approach |
|  |  |  | procedure | UMLS:ICD10PCS:0TY00Z2 | Transplantation of Right Kidney, Zooplastic, Open Approach |
|  |  |  | procedure | UMLS:ICD10PCS:0TY0 | Urinary System / Transplantation / Kidney, Right |
|  |  |  | procedure | UMLS:ICD10PCS:0TY1 | Urinary System / Transplantation / Kidney, Left |
|  | date constraint | | The terms in this group occurred between Jan 1, 2015 and Jan 1, 2022 | | |

##### **Table S4. Definition of index event in the SGLT2i non-user cohort.**

| Group 1 | | | | | |
| --- | --- | --- | --- | --- | --- |
|  | **Kidney transplant** | | | | |
|  | must have | any of | diagnosis | UMLS:ICD10CM:Z94.0 | Kidney transplant status |
|  |  |  | procedure | UMLS:ICD10PCS:0TY00Z0 | Transplantation of Right Kidney, Allogeneic, Open Approach |
|  |  |  | procedure | UMLS:ICD10PCS:0TY10Z0 | Transplantation of Left Kidney, Allogeneic, Open Approach |
|  |  |  | procedure | UMLS:ICD10PCS:0TY10Z1 | Transplantation of Left Kidney, Syngeneic, Open Approach |
|  |  |  | procedure | UMLS:ICD10PCS:0TY10Z2 | Transplantation of Left Kidney, Zooplastic, Open Approach |
|  |  |  | procedure | UMLS:ICD10PCS:0TY00Z1 | Transplantation of Right Kidney, Syngeneic, Open Approach |
|  |  |  | procedure | UMLS:ICD10PCS:0TY00Z2 | Transplantation of Right Kidney, Zooplastic, Open Approach |
|  |  |  | procedure | UMLS:ICD10PCS:0TY0 | Urinary System / Transplantation / Kidney, Right |
|  |  |  | procedure | UMLS:ICD10PCS:0TY1 | Urinary System / Transplantation / Kidney, Left |
|  | date constraint | | The terms in this group occurred between Jan 1, 2015 and Jan 1, 2022 | | |

### 3. Analyses Specifications

The Compare Outcomes Analytic supports four types of analyses: Measure of Association, Survival, Number of Instances, and Lab result distribution. The first three analyses support the “exclude patients with outcomes prior to the window” setting. This option can exclude patients from the analysis if they are not at risk for an outcome (e.g., if the outcome is a chronic disease). When "exclude patients with the outcome prior to the time window" is not checked, all patients in the cohort are included in the analysis, regardless of whether they had the outcome prior to the time window. When "exclude patients with the outcome prior to the time window" is checked, patients are excluded from the analysis if their record includes the outcome prior to the beginning of the time window. This selection will exclude all patients with the outcome prior to the index event. If the start of the time window for the analysis falls some days after the index event, patients will also be excluded if they have the outcome between the index event and the start of the time window.

#### Measure of Association Analysis

The Measure of Association Analysis calculates and compares the fraction of patients with the selected outcome. The output summary includes: Patients in each Cohort (count of patients meeting query criteria); Patients with Outcome in each Cohort (of the patients in the cohort, count of patients that had the outcome in the time window); and Risk (the fraction of patients in the cohort that have the outcome in the time window, i.e. Patients with Outcome / Patients in Cohort). In addition, Risk Difference (the difference in the risks in Cohort 1 and Cohort 2), Risk Ratio (the ratio of the risks in Cohort 1 and Cohort 2), and Odds Ratio (the ratio of the odds in Cohort 1 and Cohort 2). The bar chart shows the risk of the outcome for the both cohorts.

#### Survival Analysis

The Kaplan-Meier Analysis estimates probability of the outcome at a respective time interval (daily time interval is used in this analysis). In order to account for the patients who exited the cohort during the analysis period, and therefore should not be included in the analysis, censoring is applied. In this analysis, patients are removed from the analysis (censored) after the last fact in their record.

The output summary includes: Patients in each Cohort (count of patients meeting query criteria); Patients with Outcome (of the patients in the cohort, count of patients that had the outcome in the time window); Median Survival (the number of days when the survival drops below 50%; the “-” indicates that survival does not drop below 50% during the time window); and Survival Probability at End of Time Window (the % survival at the end of the time window). In addition, Log-Rank test, Hazard Ratio and test for Proportionality.

#### Number of Instances Analysis

The Number of Instances Analysis calculates how many times the outcome occurred in the time window. This analysis includes two additional settings: include patients with zero instances; the definition of an instance.

Selecting to exclude patients with zero instances will remove these patients from the calculations for mean number of instances, standard deviation, or median. The histogram showing the distribution of patients by number of instances will not contain a bar for zero. Alternatively, by selecting to include patients with zero instances, the mean, standard deviation, and median for number of instances will reflect these patients. The histogram will contain a bar for zero patients.

The definition of an instance affects how counts are analyzed. By selecting Date, each calendar date on which any of the terms selected in the outcome are recorded will represent one instance. For example, if the outcome is “Med A or Med B,” and a patient has “Med A” on January 3, then both medications on January 4, then “Med B” on January 6, then that patient is considered to have three instances– January 3, January 4, and January 6. Note that if an outcome occurs across several dates (e.g. Visit: inpatient encounter), then only the start date is tracked for the purpose of counting instances. A patient who begins at stay on January 1, ends that stay on January 3, begins another stay on January 10, and ends that stay on January 15, is considered to have two instances of the outcome.

Selecting Visit as an Instance will count any visit that includes the outcome as one instance, regardless of how many times it occurred. For instance, consider a patient administered an analgesic on each of the three days that make up an inpatient stay following some index event. If analgesic is an outcome, these three administrations will represent only one instance, because all three are associated with the same visit.

The output summary includes: Patients in Cohort (count of patients meeting query criteria); Patients with Outcome (of the patients in the cohort, count of patients that had the outcome in the time window); Mean (mean of the counts); Standard Deviation (standard deviation of the counts); Median (median of the counts); and Median (1+ instances) when patients with zero instances included in the analysis. In addition, T-Test statistics testing for the difference between the cohorts is included.

#### Laboratory Results Analysis

Lab Results can be included in the analysis only for the outcomes that are labs. Only the most recent lab values in the time window are included. For the lab results that are numeric, the outcome summary includes: Patients in Cohort (count of patients meeting query criteria); Patients with Outcome (of the patients in the cohort, count of patients that had the outcome in the time window); Mean (mean of the counts); and Standard Deviation (the standard deviation for lab values across patients in the cohort). In addition, T-Test statistics testing for the difference between the cohorts is included.

For the non-numeric lab results, three values are reported: counts of Negative; Positives; and Unknowns.

The counts are represented in the bar chart as percentages of the total counts.

#### Outcome Definitions

Table below outlines the definitions for each outcome and the analysis specifications. For outcome definitions consisting of more than one term, at least one term must match. Please see Appendix C for the text representation of the outcome definitions.

##### **Table S5. Outcome Definitions**

| Mortality | | | | |
| --- | --- | --- | --- | --- |
|  | **Outcome definition** | | | |
|  | | Demographics | Deceased | Deceased |
|  | | Diagnosis | UMLS:ICD10CM:R99 | Ill-defined and unknown cause of mortality |
|  | **Settings for the performed analyses** | | | |
|  | | Kaplan - Meier survival analysis | | including patients with outcome prior to the time window |
|  | | Risk analysis | | including patients with outcome prior to the time window |
| MACE | | | | |
|  | **Outcome definition** | | | |
|  | | Diagnosis | UMLS:ICD10CM:I21 | Acute myocardial infarction |
|  | | Diagnosis | UMLS:ICD10CM:I46 | Cardiac arrest |
|  | | Diagnosis | UMLS:ICD10CM:I63 | Cerebral infarction |
|  | | Demographics | Deceased | Deceased |
|  | | Diagnosis | UMLS:ICD10CM:R99 | Ill-defined and unknown cause of mortality |
|  | | Diagnosis | UMLS:ICD10CM:I22 | Subsequent ST elevation (STEMI) and non-ST elevation (NSTEMI) myocardial infarction |
|  | **Settings for the performed analyses** | | | |
|  | | Kaplan - Meier survival analysis | | excluding patients with outcome prior to the time window |
|  | | Risk analysis | | excluding patients with outcome prior to the time window |
| MAKE | | | | |
|  | **Outcome definition** | | | |
|  | | Procedure | UMLS:CPT:1012740 | Dialysis Services and Procedures |
|  | | Procedure | UMLS:CPT:90945 | Dialysis procedure other than hemodialysis (eg, peritoneal dialysis, hemofiltration, or other continuous renal replacement therapies), with single evaluation by a physician or other qualified health care professional |
|  | | Procedure | UMLS:CPT:90947 | Dialysis procedure other than hemodialysis (eg, peritoneal dialysis, hemofiltration, or other continuous renal replacement therapies) requiring repeated evaluations by a physician or other qualified health care professional, with or without substantial revision of dialysis prescription |
|  | | Procedure | UMLS:SNOMED:302497006 | Hemodialysis |
|  | | Procedure | UMLS:CPT:90935 | Hemodialysis procedure with single evaluation by a physician or other qualified health care professional |
|  | | Procedure | UMLS:CPT:1012752 | Hemodialysis Procedures |
|  | | Procedure | UMLS:CPT:90937 | Hemodialysis procedure requiring repeated evaluation(s) with or without substantial revision of dialysis prescription |
|  | | Demographics | Deceased | Deceased |
|  | | Diagnosis | UMLS:ICD10CM:R99 | Ill-defined and unknown cause of mortality |
|  | **Settings for the performed analyses** | | | |
|  | | Risk analysis | | excluding patients with outcome prior to the time window |
|  | | Kaplan - Meier survival analysis | | excluding patients with outcome prior to the time window |

### 4. **Propensity score matching**

##### **Table S6. Baseline characteristics before propensity score matching.**

| **Cohort 1 and cohort 2 patient count before and after propensity score matching** | | | | | | | | | | | | |
| --- | --- | --- | --- | --- | --- | --- | --- | --- | --- | --- | --- | --- |
|  | | | Cohort | | | Patient count before matching | | | | Patient count after matching | | |
|  | | | 1 - SGLT-2i-User | | | 869 | | | | 867 | | |
|  | | | 2 - noSGLT-2i-User | | | 48,954 | | | | 867 | | |
| **Propensity score density function - Before and after matching (cohort 1 - purple, cohort 2 - green)** | | | | | | | | | | | | |
|  |  | | 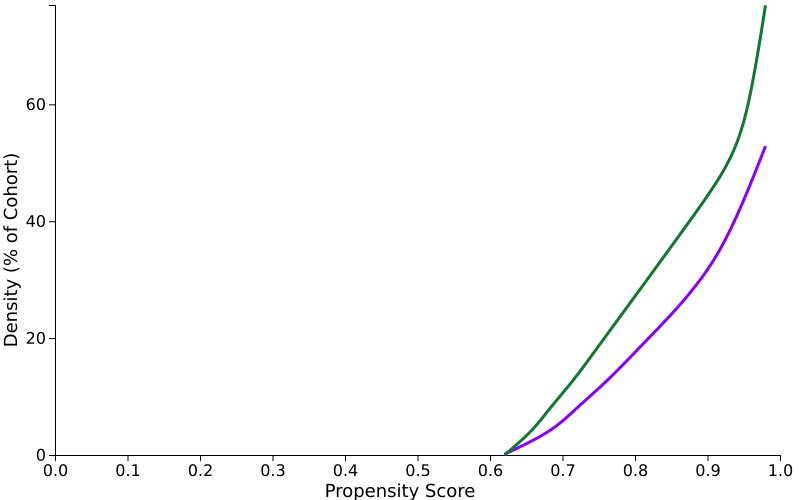 | | | | 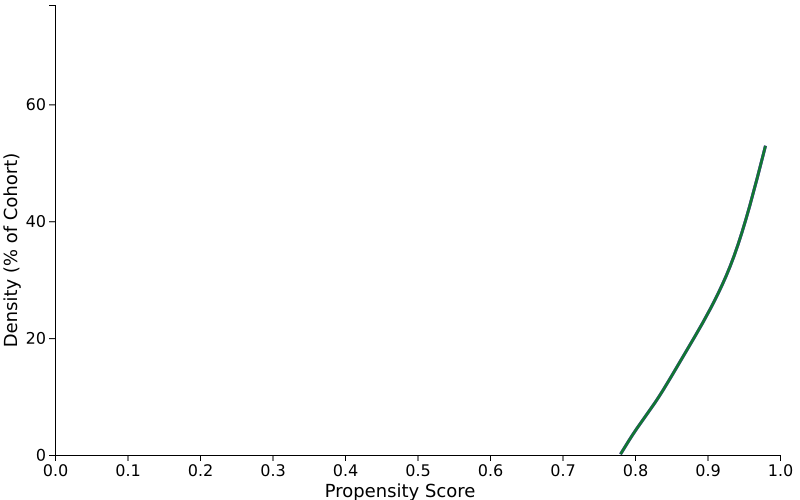 | | | | | |
| **Cohort 1 (N = 869) and cohort 2 (N = 48,954) characteristics before propensity score matching** | | | | | | | | | | | | |
|  | **Demographics** | | | | | | | | | | | |
|  |  | Cohort | | |  | Mean ± SD | | Patients | % of Cohort | | P-Value | Std diff. |
|  |  | 1 2 | | AI | Age at Index | 51.4 +/- 14.5 48.3 +/- 16.3 | | 869 48,954 | 100% 100% | | <0.001 | 0.185 |
|  |  | 1 2 | | 2106-3 | White |  | | 461 26,538 | 53.0% 54.2% | | 0.284 | 0.024 |
|  |  | 1 2 | | 2186-5 | Not Hispanic or Latino |  | | 596 29,426 | 68.6% 60.1% | | <0.001 | 0.172 |
|  |  | 1 2 | | M | Male |  | | 525 26,985 | 60.4% 55.1% | | 0.001 | 0.111 |
|  | **Diagnosis** | | | | | | | | | | | |
|  |  | Cohort | | |  | Mean ± SD | | Patients | % of Cohort | | P-Value | Std diff. |
|  |  | 1 2 | | I10-I1A | Hypertensive diseases |  | | 397 17,047 | 45.7% 34.8% | | <0.001 | 0.218 |
|  |  | 1 2 | | I70-I79 | Diseases of arteries, arterioles and capillaries |  | | 61 2,484 | 7.0% 5.1% | | 0.025 | 0.076 |
|  |  | 1 2 | | I60-I69 | Cerebrovascular diseases |  | | 22 895 | 2.5% 1.8% | | 0.084 | 0.044 |
|  |  | 1 2 | | I26-I28 | Pulmonary heart disease and diseases of pulmonary circulation |  | | 37 853 | 4.3% 1.7% | | <0.001 | 0.144 |
|  |  | 1 2 | | E66 | Overweight and obesity |  | | 38 1,718 | 4.4% 3.5% | | 0.264 | 0.039 |
|  |  | 1 2 | | C00-D49 | Neoplasms |  | | 121 4,725 | 13.9% 9.7% | | <0.001 | 0.131 |
|  |  | 1 2 | | K70-K77 | Diseases of liver |  | | 45 2,202 | 5.2% 4.5% | | 0.167 | 0.034 |
|  |  | 1 2 | | R80 | Proteinuria |  | | 52 1,954 | 6.0% 4.0% | | 0.020 | 0.088 |
|  |  | 1 2 | | M30-M36 | Systemic connective tissue disorders |  | | 48 1,536 | 5.5% 3.1% | | <0.001 | 0.111 |
|  |  | 1 2 | | I10 | Essential (primary) hypertension |  | | 320 13,427 | 36.8% 27.4% | | <0.001 | 0.217 |
|  |  | 1 2 | | I50.2 | Systolic (congestive) heart failure |  | | 15 350 | 1.7% 0.7% | | 0.020 | 0.090 |
|  |  | 1 2 | | I50.3 | Diastolic (congestive) heart failure |  | | 18 300 | 2.1% 0.6% | | <0.001 | 0.120 |
|  |  | 1 2 | | N39.0 | Urinary tract infection, site not specified |  | | 42 2,531 | 4.8% 5.2% | | 0.669 | 0.014 |
|  |  | 1 2 | | I20-I25 | Ischemic heart diseases |  | | 76 2,172 | 8.7% 4.4% | | <0.001 | 0.171 |
|  |  | 1 2 | | J40-J4A | Chronic lower respiratory diseases |  | | 55 1,788 | 6.3% 3.7% | | <0.001 | 0.122 |
|  |  | 1 2 | | E78 | Disorders of lipoprotein metabolism and other lipidemias |  | | 176 6,668 | 20.3% 13.6% | | <0.001 | 0.168 |
|  |  | 1 2 | | I21 | Acute myocardial infarction |  | | 14 254 | 1.6% 0.5% | | <0.001 | 0.114 |
|  |  | 1 2 | | I50.9 | Heart failure, unspecified |  | | 36 865 | 4.1% 1.8% | | <0.001 | 0.141 |
|  |  | 1 2 | | N17 | Acute kidney failure |  | | 63 2,623 | 7.3% 5.4% | | 0.140 | 0.071 |
|  |  | 1 2 | | N18 | Chronic kidney disease (CKD) |  | | 388 19,079 | 44.6% 39.0% | | <0.001 | 0.107 |
|  |  | 1 2 | | N19 | Unspecified kidney failure |  | | 25 1,170 | 2.9% 2.4% | | 0.073 | 0.030 |
|  |  | 1 2 | | Q61 | Cystic kidney disease |  | | 44 2,614 | 5.1% 5.3% | | 0.653 | 0.007 |
|  | **Medication** | | | | | | | | | | | |
|  |  | Cohort | | |  | Mean ± SD | | Patients | % of Cohort | | P-Value | Std diff. |
|  |  | 1 2 | | CV100 | BETA BLOCKERS/RELATED |  | | 295 10,520 | 33.9% 21.5% | | <0.001 | 0.281 |
|  |  | 1 2 | | CV200 | CALCIUM CHANNEL BLOCKERS |  | | 226 9,329 | 26.0% 19.1% | | <0.001 | 0.156 |
|  |  | 1 2 | | CV300 | ANTIARRHYTHMICS |  | | 161 6,726 | 18.5% 13.7% | | 0.001 | 0.132 |
|  |  | 1 2 | | CV700 | DIURETICS |  | | 201 6,996 | 23.1% 14.3% | | <0.001 | 0.220 |
|  |  | 1 2 | | CV350 | ANTILIPEMIC AGENTS |  | | 216 6,510 | 24.9% 13.3% | | <0.001 | 0.289 |
|  |  | 1 2 | | CV800 | ACE INHIBITORS |  | | 105 3,880 | 12.1% 7.9% | | <0.001 | 0.129 |
|  |  | 1 2 | | CV805 | ANGIOTENSIN II INHIBITOR |  | | 124 3,293 | 14.3% 6.7% | | <0.001 | 0.241 |
|  |  | 1 2 | | CV250 | ANTIANGINALS |  | | 48 1,355 | 5.5% 2.8% | | <0.001 | 0.129 |
|  |  | 1 2 | | C10AA | HMG CoA reductase inhibitors |  | | 203 5,999 | 23.4% 12.3% | | <0.001 | 0.290 |
|  |  | 1 2 | | HS502 | ORAL HYPOGLYCEMIC AGENTS,ORAL |  | | 10 164 | 1.2% 0.3% | | <0.001 | 0.089 |
|  | **Laboratory** | | | | | | | | | | | |
|  |  | Cohort | | |  | Mean ± SD | | Patients | % of Cohort | | P-Value | Std diff. |
|  |  | 1 2 | | 9002 | Cholesterol in LDL [Mass/volume] in Serum or Plasma | 91.0 +/- 35.8 95.5 +/- 38.6 | | 229 8,936 | 26.4% 18.4% | | 0.085 | 0.119 |
|  |  | 1 2 | |  | 0 - 55 mg/dL |  | | 44 1,361 | 4.5% 2.5% | | <0.001 | 0.110 |
|  |  | 1 2 | |  | 55 - 100 mg/dL |  | | 146 5,394 | 15.0% 9.9% | | <0.001 | 0.153 |
|  |  | 1 2 | |  | 100 - 130 mg/dL |  | | 80 3,110 | 8.2% 5.7% | | 0.002 | 0.096 |
|  |  | 1 2 | |  | 130 - 160 mg/dL |  | | 35 1,458 | 3.6% 2.7% | | 0.098 | 0.052 |
|  |  | 1 2 | |  | 160 - 190 mg/dL |  | | 11 486 | 1.2% 0.9% | | 0.485 | 0.022 |
|  |  | 1 2 | |  | 190 - 0 mg/dL |  | | 11 194 | 1.2% 0.4% | | 0.001 | 0.085 |
|  |  | 1 2 | | 9001 | Cholesterol in HDL [Mass/volume] in Serum or Plasma | 47.7 +/- 19.0 50.7 +/- 19.3 | | 222 8,972 | 25.5% 18.5% | | 0.017 | 0.155 |
|  |  | 1 2 | |  | 0 - 45 mg/dL |  | | 122 4,519 | 14.1% 9.3% | | <0.001 | 0.140 |
|  |  | 1 2 | |  | 45 - 60 mg/dL |  | | 99 4,025 | 11.4% 8.3% | | 0.002 | 0.098 |
|  |  | 1 2 | |  | 60 - 0 mg/dL |  | | 54 2,867 | 6.2% 5.9% | | 0.674 | 0.013 |
|  |  | 1 2 | | 9083 | BMI | 27.6 +/- 5.3 26.9 +/- 5.7 | | 409 16,809 | 53.0% 38.8% | | 0.016 | 0.124 |
|  |  | 1 2 | |  | 0 - 21 kg/m2 |  | | 50 2,950 | 6.5% 6.8% | | 0.722 | 0.013 |
|  |  | 1 2 | |  | 21 - 26 kg/m2 |  | | 184 7,290 | 23.8% 16.8% | | <0.001 | 0.175 |
|  |  | 1 2 | |  | 26 - 31 kg/m2 |  | | 166 6,474 | 21.5% 14.9% | | <0.001 | 0.171 |
|  |  | 1 2 | |  | 31 - 36 kg/m2 |  | | 102 3,352 | 13.2% 7.7% | | <0.001 | 0.180 |
|  |  | 1 2 | |  | 36 - 0 kg/m2 |  | | 43 1,562 | 5.6% 3.6% | | 0.004 | 0.094 |
|  |  | 1 2 | | 9085 | Blood Pressure, Systolic | 133.0 +/- 20.5 131.1 +/- 20.4 | | 420 17,720 | 54.4% 40.9% | | 0.018 | 0.116 |
|  |  | 1 2 | |  | 0 - 110 mm[Hg] |  | | 120 5,643 | 15.5% 13.0% | | 0.038 | 0.072 |
|  |  | 1 2 | |  | 110 - 130 mm[Hg] |  | | 271 11,519 | 35.1% 26.6% | | <0.001 | 0.186 |
|  |  | 1 2 | |  | 130 - 140 mm[Hg] |  | | 238 9,175 | 30.8% 21.2% | | <0.001 | 0.222 |
|  |  | 1 2 | |  | 140 - 160 mm[Hg] |  | | 246 9,357 | 31.9% 21.6% | | <0.001 | 0.234 |
|  |  | 1 2 | |  | 160 - 0 mm[Hg] |  | | 137 4,833 | 17.7% 11.1% | | <0.001 | 0.189 |
|  |  | 1 2 | | 9086 | Blood Pressure, Diastolic | 78.4 +/- 12.9 77.4 +/- 12.8 | | 421 17,734 | 48.5% 36.5% | | 0.121 | 0.074 |
|  |  | 1 2 | |  | 0 - 70 mm[Hg] |  | | 231 9,677 | 26.6% 19.9% | | <0.001 | 0.157 |
|  |  | 1 2 | |  | 70 - 80 mm[Hg] |  | | 325 12,686 | 37.4% 26.1% | | <0.001 | 0.246 |
|  |  | 1 2 | |  | 80 - 90 mm[Hg] |  | | 290 12,186 | 33.4% 25.1% | | <0.001 | 0.185 |
|  |  | 1 2 | |  | 90 - 0 mm[Hg] |  | | 206 8,110 | 23.7% 16.7% | | <0.001 | 0.176 |
|  |  | 1 2 | | 9004 | Triglyceride [Mass/volume] in Serum, Plasma or Blood | 147.4 +/- 103.3 145.0 +/- 96.9 | | 224 10,013 | 25.8% 20.6% | | 0.801 | 0.014 |
|  |  | 1 2 | |  | 0 - 150 mg/dL |  | | 172 7,868 | 19.8% 16.2% | | 0.007 | 0.095 |
|  |  | 1 2 | |  | 150 - 200 mg/dL |  | | 62 2,673 | 7.1% 5.5% | | 0.057 | 0.065 |
|  |  | 1 2 | |  | 200 - 500 mg/dL |  | | 61 2,429 | 7.0% 5.0% | | 0.014 | 0.082 |
|  |  | 1 2 | |  | 500 - 0 mg/dL |  | | 11 146 | 1.3% 0.3% | | <0.001 | 0.105 |
|  |  | 1 2 | | 9014 | Hemoglobin [Mass/volume] in Blood | 11.9 +/- 2.1 12.0 +/- 2.1 | | 448 21,563 | 51.5% 44.4% | | 0.532 | 0.020 |
|  |  | 1 2 | |  | 0 - 10 g/dL |  | | 161 7,428 | 18.5% 15.3% | | 0.015 | 0.085 |
|  |  | 1 2 | |  | 10 - 12 g/dL |  | | 278 12,831 | 32.0% 26.4% | | <0.001 | 0.124 |
|  |  | 1 2 | |  | 12 - 14 g/dL |  | | 267 12,688 | 30.7% 26.1% | | 0.004 | 0.103 |
|  |  | 1 2 | |  | 14 - 16 g/dL |  | | 123 6,171 | 14.2% 12.7% | | 0.201 | 0.045 |
|  |  | 1 2 | |  | 16 - 0 g/dL |  | | 23 1,363 | 2.7% 2.8% | | 0.922 | 0.004 |
|  |  | 1 2 | | 8001 | Glomerular filtration rate/1.73 sq M.predicted [Volume Rate/Area] in Serum, Plasma or Blood by Creatinine-based formula (MDRD) | 36.1 +/- 27.6 37.9 +/- 30.2 | | 449 19,276 | 51.6% 39.7% | | 0.066 | 0.080 |
|  |  | 1 2 | |  | 0 - 15 mL/min/{1.73_m2} |  | | 240 9,334 | 27.6% 19.2% | | <0.001 | 0.199 |
|  |  | 1 2 | |  | 15 - 30 mL/min/{1.73_m2} |  | | 97 5,540 | 11.1% 11.4% | | 0.833 | 0.008 |
|  |  | 1 2 | |  | 30 - 45 mL/min/{1.73_m2} |  | | 130 5,781 | 15.0% 11.9% | | 0.008 | 0.092 |
|  |  | 1 2 | |  | 45 - 60 mL/min/{1.73_m2} |  | | 182 6,995 | 21.0% 14.4% | | <0.001 | 0.174 |
|  |  | 1 2 | |  | 60 - 1000 mL/min/{1.73_m2} |  | | 164 7,590 | 18.9% 15.6% | | 0.012 | 0.087 |
|  |  | 1 2 | | 9029 | Sodium [Moles/volume] in Serum, Plasma or Blood | 139.0 +/- 3.3 139.0 +/- 3.3 | | 452 21,174 | 58.5% 48.8% | | 0.684 | 0.001 |
|  |  | 1 2 | |  | 0 - 135 mmol/L |  | | 138 7,393 | 15.9% 15.2% | | 0.552 | 0.021 |
|  |  | 1 2 | |  | 135 - 145 mmol/L |  | | 487 22,620 | 56.1% 46.5% | | <0.001 | 0.193 |
|  |  | 1 2 | |  | 145 - 200 mmol/L |  | | 31 1,505 | 3.6% 3.1% | | 0.359 | 0.032 |
|  |  | 1 2 | |  | 200 - 0 mmol/L |  | | 0 10 | 0% 0.0% | | 0.673 | 0.021 |
|  |  | 1 2 | | 9028 | Potassium [Moles/volume] in Serum, Plasma or Blood | 4.4 +/- 0.6 4.4 +/- 0.6 | | 457 21,913 | 59.2% 50.5% | | 0.057 | 0.086 |
|  |  | 1 2 | |  | 0 - 3.50 mmol/L |  | | 89 4,444 | 11.5% 10.2% | | 0.245 | 0.041 |
|  |  | 1 2 | |  | 3.60 - 5 mmol/L |  | | 419 20,386 | 54.3% 47.0% | | <0.001 | 0.146 |
|  |  | 1 2 | |  | 5.10 - 5.50 mmol/L |  | | 125 5,705 | 16.2% 13.2% | | 0.013 | 0.086 |
|  |  | 1 2 | |  | 5.50 - 0 mmol/L |  | | 84 3,729 | 10.9% 8.6% | | 0.025 | 0.077 |

##### **Table S7. Baseline characteristics after propensity score matching.**

| **Cohort 1 (N = 867) and cohort 2 (N = 867) characteristics after propensity score matching** | | | | | | | | | |
| --- | --- | --- | --- | --- | --- | --- | --- | --- | --- |
|  | **Demographics** | | | | | | | | |
|  |  | Cohort | |  | Mean ± SD | Patients | % of Cohort | P-Value | Std diff. |
|  |  | 1 2 | AI | Age at Index | 51.3 +/- 14.5 51.5 +/- 15.1 | 867 867 | 100% 100% | 0.925 | 0.005 |
|  |  | 1 2 | 2106-3 | White |  | 460 481 | 53.1% 55.5% | 0.308 | 0.042 |
|  |  | 1 2 | 2186-5 | Not Hispanic or Latino |  | 594 601 | 68.5% 69.3% | 0.296 | 0.013 |
|  |  | 1 2 | M | Male |  | 523 517 | 60.3% 59.6% | 0.675 | 0.011 |
|  | **Diagnosis** | | | | | | | | |
|  |  | Cohort | |  | Mean ± SD | Patients | % of Cohort | P-Value | Std diff. |
|  |  | 1 2 | I10-I1A | Hypertensive diseases |  | 395 419 | 45.5% 48.3% | 0.202 | 0.054 |
|  |  | 1 2 | I70-I79 | Diseases of arteries, arterioles and capillaries |  | 59 58 | 6.8% 6.7% | 0.429 | 0.001 |
|  |  | 1 2 | I60-I69 | Cerebrovascular diseases |  | 22 16 | 2.5% 1.8% | 0.500 | 0.044 |
|  |  | 1 2 | I26-I28 | Pulmonary heart disease and diseases of pulmonary circulation |  | 36 35 | 4.2% 4.0% | 0.565 | 0.001 |
|  |  | 1 2 | E66 | Overweight and obesity |  | 38 30 | 4.4% 3.5% | 0.620 | 0.035 |
|  |  | 1 2 | C00-D49 | Neoplasms |  | 119 126 | 13.7% 14.5% | 0.651 | 0.023 |
|  |  | 1 2 | K70-K77 | Diseases of liver |  | 44 38 | 5.1% 4.4% | 0.657 | 0.033 |
|  |  | 1 2 | R80 | Proteinuria |  | 50 51 | 5.8% 5.9% | 0.083 | 0.001 |
|  |  | 1 2 | M30-M36 | Systemic connective tissue disorders |  | 48 53 | 5.5% 5.0% | 0.824 | 0.021 |
|  |  | 1 2 | I10 | Essential (primary) hypertension |  | 319 349 | 36.8% 40.3% | 0.248 | 0.069 |
|  |  | 1 2 | I50.2 | Systolic (congestive) heart failure |  | 15 12 | 1.7% 1.4% | 0.826 | 0.021 |
|  |  | 1 2 | I50.3 | Diastolic (congestive) heart failure |  | 16 13 | 1.8% 1.5% | 0.411 | 0.022 |
|  |  | 1 2 | N39.0 | Urinary tract infection, site not specified |  | 42 53 | 4.8% 6.1% | 0.638 | 0.054 |
|  |  | 1 2 | I20-I25 | Ischemic heart diseases |  | 74 66 | 8.5% 7.6% | 0.718 | 0.028 |
|  |  | 1 2 | J40-J4A | Chronic lower respiratory diseases |  | 53 50 | 6.1% 5.8% | 0.915 | 0.010 |
|  |  | 1 2 | E78 | Disorders of lipoprotein metabolism and other lipidemias |  | 176 178 | 20.3% 20.5% | 0.382 | 0.001 |
|  |  | 1 2 | I21 | Acute myocardial infarction |  | 13 10 | 1.5% 1.2% | 0.834 | 0.031 |
|  |  | 1 2 | I50.9 | Heart failure, unspecified |  | 34 29 | 3.9% 3.3% | 0.565 | 0.029 |
|  |  | 1 2 | N17 | Acute kidney failure |  | 61 52 | 7.0% 6.0% | 0.385 | 0.044 |
|  |  | 1 2 | N18 | Chronic kidney disease (CKD) |  | 386 378 | 44.5% 43.6% | 0.539 | 0.011 |
|  |  | 1 2 | N19 | Unspecified kidney failure |  | 24 25 | 2.8% 2.9% | 0.221 | 0.001 |
|  |  | 1 2 | Q61 | Cystic kidney disease |  | 44 41 | 5.1% 4.8% | 0.629 | 0.014 |
|  | **Medication** | | | | | | | | |
|  |  | Cohort | |  | Mean ± SD | Patients | % of Cohort | P-Value | Std diff. |
|  |  | 1 2 | CV100 | BETA BLOCKERS/RELATED |  | 293 292 | 33.8% 33.7% | 0.592 | 0.001 |
|  |  | 1 2 | CV200 | CALCIUM CHANNEL BLOCKERS |  | 224 223 | 25.8% 25.7% | 0.640 | 0.001 |
|  |  | 1 2 | CV300 | ANTIARRHYTHMICS |  | 159 146 | 18.3% 16.8% | 0.589 | 0.028 |
|  |  | 1 2 | CV700 | DIURETICS |  | 199 184 | 23.0% 21.2% | 0.320 | 0.041 |
|  |  | 1 2 | CV350 | ANTILIPEMIC AGENTS |  | 215 248 | 24.8% 28.6% | 0.183 | 0.078 |
|  |  | 1 2 | CV800 | ACE INHIBITORS |  | 104 99 | 12.0% 11.4% | 0.544 | 0.011 |
|  |  | 1 2 | CV805 | ANGIOTENSIN II INHIBITOR |  | 122 127 | 12.0% 14.6% | 0.883 | 0.013 |
|  |  | 1 2 | CV250 | ANTIANGINALS |  | 46 46 | 5.3% 5.3% | 0.119 | 0.001 |
|  |  | 1 2 | C10AA | HMG CoA reductase inhibitors |  | 202 233 | 23.3% 26.9% | 0.285 | 0.084 |
|  |  | 1 2 | HS502 | ORAL HYPOGLYCEMIC AGENTS,ORAL |  | 10 11 | 1.2% 1.3% | 0.826 | 0.011 |
|  | **Laboratory** | | | | | | | | |
|  |  | Cohort | |  | Mean ± SD | Patients | % of Cohort | P-Value | Std diff. |
|  |  | 1 2 | 9002 | Cholesterol in LDL [Mass/volume] in Serum or Plasma | 90.9 +/- 35.9 49.7 +/- 18.5 | 228 233 | 26.2% 26.8% | 0.510 | 0.061 |
|  |  | 1 2 |  | 0 - 55 mg/dL |  | 39 41 | 4.5% 4.7% | 0.818 | 0.012 |
|  |  | 1 2 |  | 55 - 100 mg/dL |  | 129 131 | 14.8% 15.1% | 0.892 | 0.007 |
|  |  | 1 2 |  | 100 - 130 mg/dL |  | 70 76 | 8.1% 8.7% | 0.602 | 0.027 |
|  |  | 1 2 |  | 130 - 160 mg/dL |  | 31 33 | 3.6% 3.8% | 0.798 | 0.013 |
|  |  | 1 2 |  | 160 - 190 mg/dL |  | 10 10 | 1.2% 1.2% | 1 | <0.001 |
|  |  | 1 2 |  | 190 - 0 mg/dL |  | 10 10 | 1.2% 1.2% | 1 | <0.001 |
|  |  | 1 2 | 9001 | Cholesterol in HDL [Mass/volume] in Serum or Plasma | 47.9 +/- 19.0 49.7 +/- 18.5 | 221 232 | 25.4% 26.7% | 0.138 | 0.090 |
|  |  | 1 2 |  | 0 - 45 mg/dL |  | 108 110 | 12.4% 12.7% | 0.884 | 0.007 |
|  |  | 1 2 |  | 45 - 60 mg/dL |  | 87 82 | 10.0% 9.4% | 0.684 | 0.021 |
|  |  | 1 2 |  | 60 - 0 mg/dL |  | 48 60 | 5.5% 6.9% | 0.231 | 0.061 |
|  |  | 1 2 | 9083 | BMI | 27.6 +/- 5.4 27.1 +/- 5.4 | 408 407 | 47.0%  46.8% | 0.565 | 0.090 |
|  |  | 1 2 |  | 0 - 21 kg/m2 |  | 50 44 | 5.8% 5.1% | 0.523 | 0.033 |
|  |  | 1 2 |  | 21 - 26 kg/m2 |  | 183 187 | 21.1% 21.5% | 0.811 | 0.012 |
|  |  | 1 2 |  | 26 - 31 kg/m2 |  | 165 152 | 19.0% 17.5% | 0.413 | 0.042 |
|  |  | 1 2 |  | 31 - 36 kg/m2 |  | 102 93 | 11.7% 10.7% | 0.490 | 0.035 |
|  |  | 1 2 |  | 36 - 0 kg/m2 |  | 43 52 | 5.0% 6.0% | 0.340 | 0.049 |
|  |  | 1 2 | 9085 | Blood Pressure, Systolic | 133.0 +/- 20.3 134.0 +/- 18.1 | 419 423 | 48.2% 48.7% | 0.488 | 0.001 |
|  |  | 1 2 |  | 0 - 110 mm[Hg] |  | 120 109 | 13.8% 12.5% | 0.431 | 0.040 |
|  |  | 1 2 |  | 110 - 130 mm[Hg] |  | 271 262 | 31.2% 30.2% | 0.630 | 0.025 |
|  |  | 1 2 |  | 130 - 140 mm[Hg] |  | 237 246 | 27.3% 28.3% | 0.621 | 0.025 |
|  |  | 1 2 |  | 140 - 160 mm[Hg] |  | 245 236 | 28.2% 27.2% | 0.621 | 0.025 |
|  |  | 1 2 |  | 160 - 0 mm[Hg] |  | 136 123 | 15.6% 14.2% | 0.376 | 0.045 |
|  |  | 1 2 | 9086 | Blood Pressure, Diastolic | 78.4 +/- 13.0 78.4 +/- 12.1 | 420 423 | 48.3% 48.7% | 0.711 | 0.001 |
|  |  | 1 2 |  | 0 - 70 mm[Hg] |  | 204 194 | 23.5% 22.3% | 0.561 | 0.030 |
|  |  | 1 2 |  | 70 - 80 mm[Hg] |  | 288 279 | 33.2% 32.2% | 0.635 | 0.024 |
|  |  | 1 2 |  | 80 - 90 mm[Hg] |  | 257 267 | 29.6% 30.8% | 0.591 | 0.027 |
|  |  | 1 2 |  | 90 - 0 mm[Hg] |  | 182 176 | 20.9% 20.3% | 0.717 | 0.018 |
|  |  | 1 2 | 9004 | Triglyceride [Mass/volume] in Serum, Plasma or Blood | 147.0 +/- 121.1 150.0 +/- 103.2 | 223 233 | 25.8% 26.8% | 0.988 | 0.021 |
|  |  | 1 2 |  | 0 - 150 mg/dL |  | 152 158 | 17.5% 18.2% | 0.703 | 0.019 |
|  |  | 1 2 |  | 150 - 200 mg/dL |  | 55 65 | 6.3% 7.5% | 0.342 | 0.048 |
|  |  | 1 2 |  | 200 - 500 mg/dL |  | 54 51 | 6.2% 5.9% | 0.762 | 0.015 |
|  |  | 1 2 |  | 500 - 0 mg/dL |  | 10 10 | 1.2% 1.2% | 1 | <0.001 |
|  |  | 1 2 | 9014 | Hemoglobin [Mass/volume] in Blood | 12.0 +/- 2.1 12.2 +/- 2.0 | 447 456 | 51.4% 52.5% | 0.021 | 0.094 |
|  |  | 1 2 |  | 0 - 10 g/dL |  | 142 118 | 16.3% 13.6% | 0.103 | 0.083 |
|  |  | 1 2 |  | 10 - 12 g/dL |  | 246 228 | 28.3% 26.2% | 0.320 | 0.051 |
|  |  | 1 2 |  | 12 - 14 g/dL |  | 236 243 | 27.2% 28.0% | 0.700 | 0.020 |
|  |  | 1 2 |  | 14 - 16 g/dL |  | 110 138 | 12.7% 15.9% | 0.052 | 0.099 |
|  |  | 1 2 |  | 16 - 0 g/dL |  | 21 30 | 2.4% 3.5% | 0.200 | 0.065 |
|  |  | 1 2 | 8001 | Glomerular filtration rate/1.73 sq M.predicted [Volume Rate/Area] in Serum, Plasma or Blood by Creatinine-based formula (MDRD) | 36.1 +/- 27.5 38.7 +/- 26.1 | 448 452 | 51.6% 52.0% | 0.005 | 0.092 |
|  |  | 1 2 |  | 0 - 15 mL/min/{1.73_m2} |  | 212 180 | 24.4% 20.7% | 0.061 | 0.095 |
|  |  | 1 2 |  | 15 - 30 mL/min/{1.73_m2} |  | 85 78 | 9.8% 9.0% | 0.562 | 0.030 |
|  |  | 1 2 |  | 30 - 45 mL/min/{1.73_m2} |  | 115 136 | 13.2% 15.6% | 0.147 | 0.074 |
|  |  | 1 2 |  | 45 - 60 mL/min/{1.73_m2} |  | 161 188 | 18.5% 21.6% | 0.100 | 0.084 |
|  |  | 1 2 |  | 60 - 1000 mL/min/{1.73_m2} |  | 145 167 | 16.7% 19.2% | 0.163 | 0.071 |
|  |  | 1 2 | 9029 | Sodium [Moles/volume] in Serum, Plasma or Blood | 139.0 +/- 3.3 139.0 +/- 3.2 | 451 449 | 51.9% 51.7% | 0.305 | 0.078 |
|  |  | 1 2 |  | 0 - 135 mmol/L |  | 122 96 | 14.0% 11.1% | 0.057 | 0.097 |
|  |  | 1 2 |  | 135 - 145 mmol/L |  | 432 437 | 49.7% 50.3% | 0.797 | 0.013 |
|  |  | 1 2 |  | 145 - 200 mmol/L |  | 28 27 | 3.2% 3.1% | 0.891 | 0.007 |
|  |  | 1 2 |  | 200 - 0 mmol/L |  | 0 0 | 0% 0% | -- | -- |
|  |  | 1 2 | 9028 | Potassium [Moles/volume] in Serum, Plasma or Blood | 4.4 +/- 0.6 4.4 +/- 0.6 | 456 458 | 52.5% 52.7% | 0.666 | 0.009 |
|  |  | 1 2 |  | 0 - 3.50 mmol/L |  | 88 73 | 10.1% 8.4% | 0.212 | 0.064 |
|  |  | 1 2 |  | 3.60 - 5 mmol/L |  | 418 423 | 48.1% 48.7% | 0.798 | 0.013 |
|  |  | 1 2 |  | 5.10 - 5.50 mmol/L |  | 125 108 | 14.4% 12.4% | 0.227 | 0.062 |
|  |  | 1 2 |  | 5.50 - 0 mmol/L |  | 84 61 | 9.7% 7.0% | 0.045 | 0.102 |

### 5. **Results**

##### **Table S8. Follow-up duration in both cohorts before and after propensity score matching.**

| **Follow-up Time (Before Matching)** | | | | | | |
| --- | --- | --- | --- | --- | --- | --- |
|  |  | Cohort | Mean Follow-up (Days) | Standard Deviation | Median Follow-up (Days) | Interquartile Range |
|  |  | SGLT-2i-User | 2619.522 | 524.253 | 3006 | 748 |
|  |  | noSGLT-2i-User | 2276.917 | 727.082 | 2413 | 1281 |
| 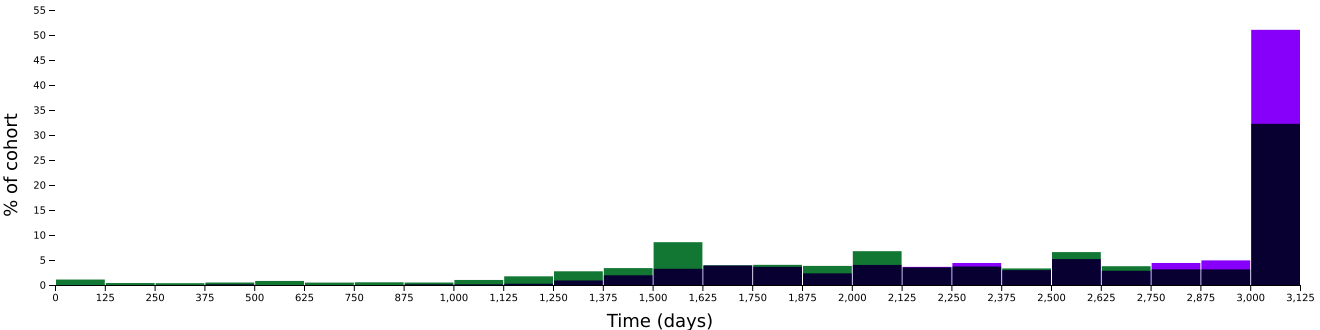 | | | | | | |
| **Follow-up Time (After Matching)** | | | | | | |
|  |  | Cohort | Mean Follow-up (Days) | Standard Deviation | Median Follow-up (Days) | Interquartile Range |
|  |  | SGLT-2i-User | 2619.556 | 524.593 | 3006 | 748 |
|  |  | noSGLT-2i-User | 2527.352 | 680.744 | 3006 | 898 |
| 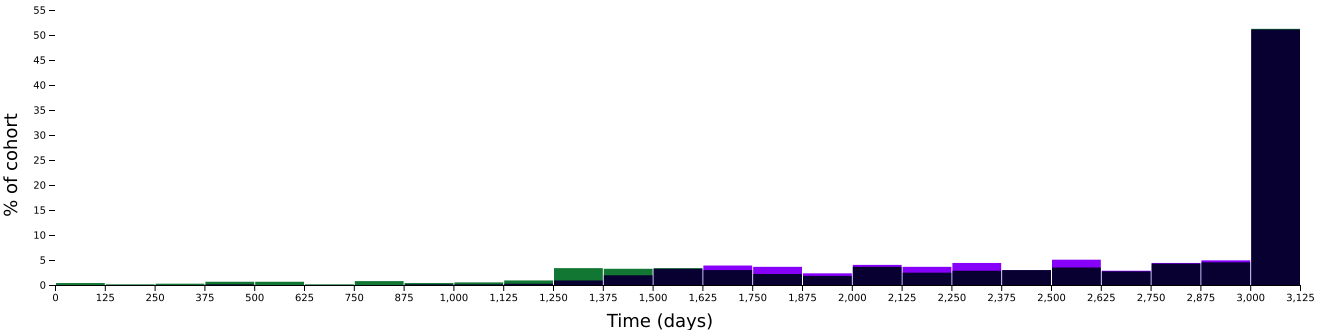 | | | | | | |

##### **Table S9. Risk and survival analyses for all-cause mortality.**

| **1 Mortality** | | | | | | | | | | | |
| --- | --- | --- | --- | --- | --- | --- | --- | --- | --- | --- | --- |
|  | | **Risk analysis** | | | | | | | | | |
|  |  | | | Cohort | | | Patients in cohort | Patients with outcome | Risk | | |
|  | | |  | 1 | | SGLT-2i-User | 867 | 35 | 0.040 | | |
|  | | |  | 2 | | noSGLT-2i-User | 867 | 59 | 0.068 | | |
|  | | |  |  | | |  | 95% CI | z | p |  |
|  | | |  | **Risk Difference** | | | -0.027 | (-0.049, -0.006) | -2.542 | 0.011 |  |
|  | | |  | **Risk Ratio** | | | 0.594 | (0.392, 0.891) | N/A | N/A |  |
|  | | |  | **Odds Ratio** | | | 0.552 | (0.375, 0.884) | N/A | N/A |  |
|  | |  | | | 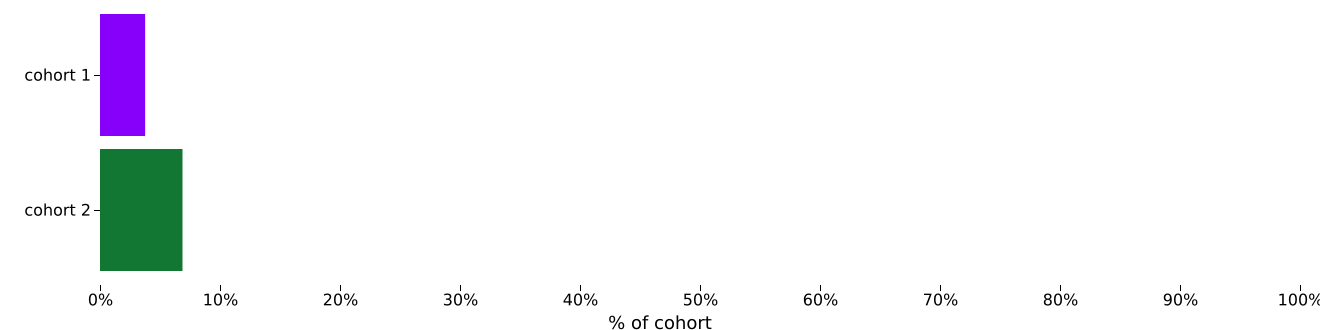 | | | | | | |
|  | | **Kaplan - Meier survival analysis** | | | | | | | | | |
|  | | |  | Cohort | | | Patients in cohort | Patients with outcome | Median survival (days) | Survival probability at end of time window | |
|  | | |  | 1 | | SGLT-2i-User | 867 | 35 | -- | 94.56% | |
|  | | |  | 2 | | noSGLT-2i-User | 867 | 59 | -- | 91.27% | |
|  | | |  |  | | | χ^2^ | df | p |  |  |
|  | | |  | **Log-Rank Test** | | | 7.629 | 1 | 0.005 |  |  |
|  | | |  |  | | | Hazard Ratio | 95% CI | χ^2^ | df | p |
|  | | |  | **Hazard Ratio and Proportionality** | | | 0.559 | (0.368, 0.850) | 3.687 | 1 | 0.055 |
|  | |  | | | 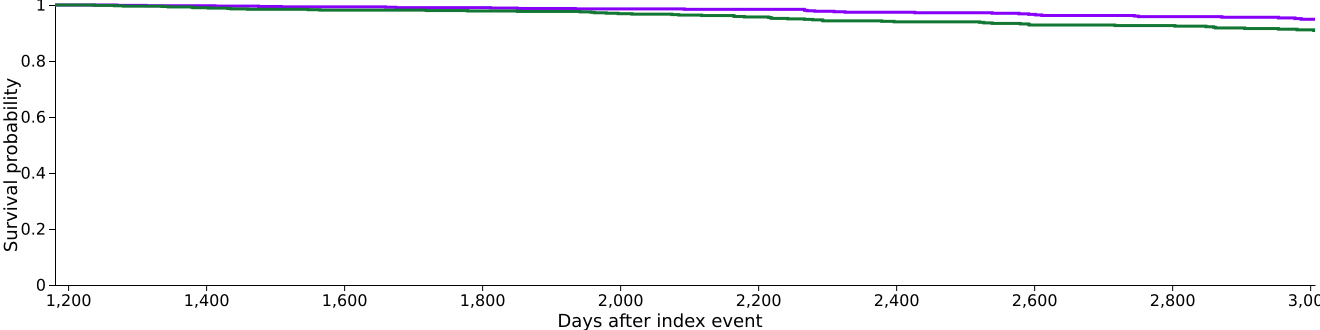 | | | | | | |
| **Table S10. Risk and survival analyses for MACE.** | | | | | | | | | | | |
| **2 MACE** | | | | | | | | | | | |
|  | | **Risk analysis excluding patients with outcome prior to the time window** | | | | | | | | | |
|  |  | | | Cohort | | | Patients in cohort | Patients with outcome | Risk | | |
|  | | |  | 1 | | SGLT-2i-User | 784 | 80 | 0.102 | | |
|  | | |  | 2 | | noSGLT-2i-User | 785 | 87 | 0.111 | | |
|  | | |  |  | | |  | 95% CI | z | p |  |
|  | | |  | **Risk Difference** | | | -0.009 | (-0.039, 0.022) | -0.564 | 0.573 |  |
|  | | |  | **Risk Ratio** | | | 0.921 | (0.691, 1.227) | N/A | N/A |  |
|  | | |  | **Odds Ratio** | | | 0.912 | (0.661, 1.257) | N/A | N/A |  |
|  | |  | | | 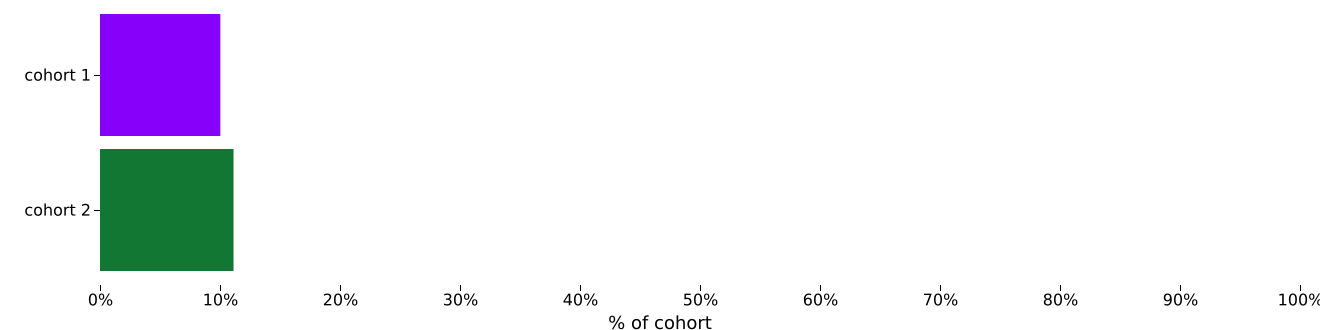 | | | | | | |
|  | |  | | | 83 patients in Cohort 1 and 82 patients in Cohort 2 were excluded from results because they had the outcome prior to the time window. | | | | | | |
|  | | **Kaplan - Meier survival analysis excluding patients with outcome prior to the time window** | | | | | | | | | |
|  | | |  | Cohort | | | Patients in cohort | Patients with outcome | Median survival (days) | Survival probability at end of time window | |
|  | | |  | 1 | | SGLT-2i-User | 784 | 80 | -- | 87.18% | |
|  | | |  | 2 | | noSGLT-2i-User | 785 | 87 | -- | 85.96% | |
|  | | |  |  | | | χ^2^ | df | p |  |  |
|  | | |  | **Log-Rank Test** | | | 0.836 | 1 | 0.361 |  |  |
|  | | |  |  | | | Hazard Ratio | 95% CI | χ^2^ | df | p |
|  | | |  | **Hazard Ratio and Proportionality** | | | 0.868 | (0.641, 1.176) | 2.521 | 1 | 0.112 |
|  | |  | | | 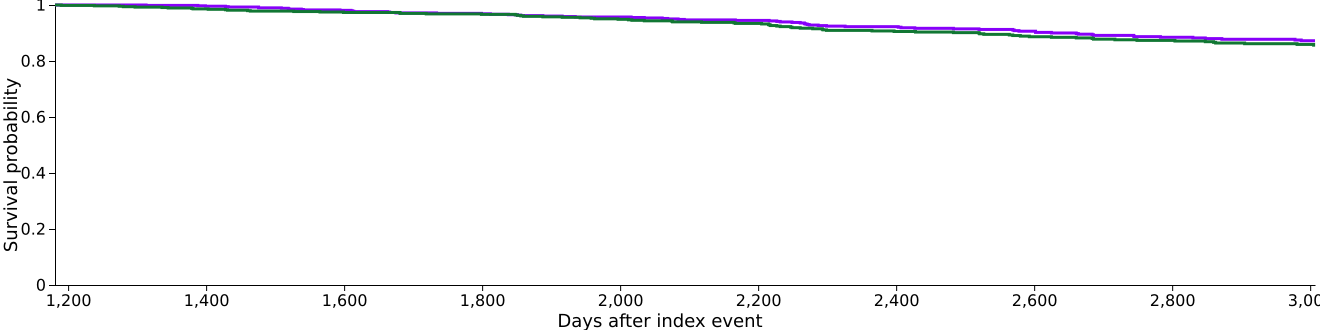 | | | | | | |
|  | |  | | | 83 patients in Cohort 1 and 82 patients in Cohort 2 were excluded from results because they had the outcome prior to the time window. | | | | | | |

##### **Table S11. Risk and survival analyses for MAKE.**

| **3 MAKE** | | | | | | | | | | | |
| --- | --- | --- | --- | --- | --- | --- | --- | --- | --- | --- | --- |
|  | | **Risk analysis excluding patients with outcome prior to the time window** | | | | | | | | | |
|  |  | | | Cohort | | | Patients in cohort | Patients with outcome | Risk | | |
|  | | |  | 1 | | SGLT-2i-User | 790 | 54 | 0.068 | | |
|  | | |  | 2 | | noSGLT-2i-User | 762 | 76 | 0.099 | | |
|  | | |  |  | | |  | 95% CI | z | p |  |
|  | | |  | **Risk Difference** | | | -0.031 | (-0.059, -0.004) | -2.231 | 0.026 |  |
|  | | |  | **Risk Ratio** | | | 0.685 | (0.491, 0.957) | N/A | N/A |  |
|  | | |  | **Odds Ratio** | | | 0.662 | (0.460, 0.953) | N/A | N/A |  |
|  | |  | | | 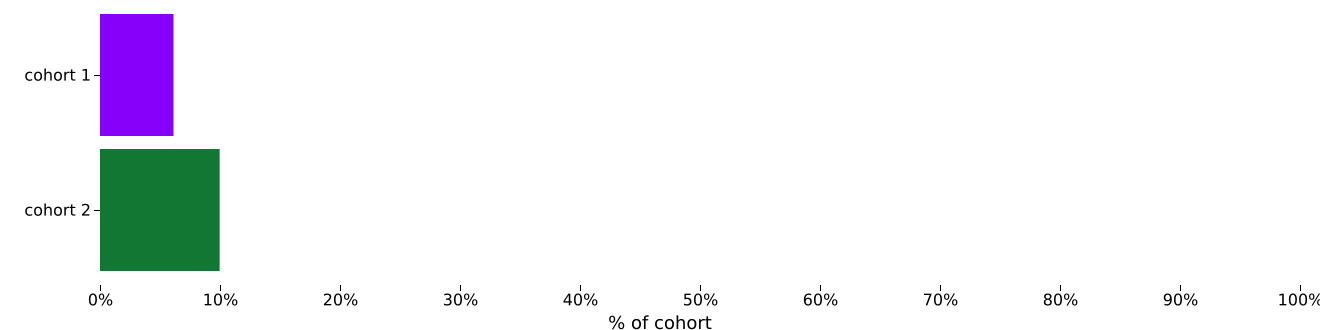 | | | | | | |
|  | |  | | | 77 patients in Cohort 1 and 105 patients in Cohort 2 were excluded from results because they had the outcome prior to the time window. | | | | | | |
|  | | **Kaplan - Meier survival analysis excluding patients with outcome prior to the time window** | | | | | | | | | |
|  | | |  | Cohort | | | Patients in cohort | Patients with outcome | Median survival (days) | Survival probability at end of time window | |
|  | | |  | 1 | | SGLT-2i-User | 790 | 54 | -- | 91.19% | |
|  | | |  | 2 | | noSGLT-2i-User | 762 | 76 | -- | 87.63% | |
|  | | |  |  | | | χ^2^ | df | p |  |  |
|  | | |  | **Log-Rank Test** | | | 6.055 | 1 | 0.014 |  |  |
|  | | |  |  | | | Hazard Ratio | 95% CI | χ^2^ | df | p |
|  | | |  | **Hazard Ratio and Proportionality** | | | 0.648 | (0.457, 0.918) | 4.174 | 1 | 0.041 |
|  | |  | | | 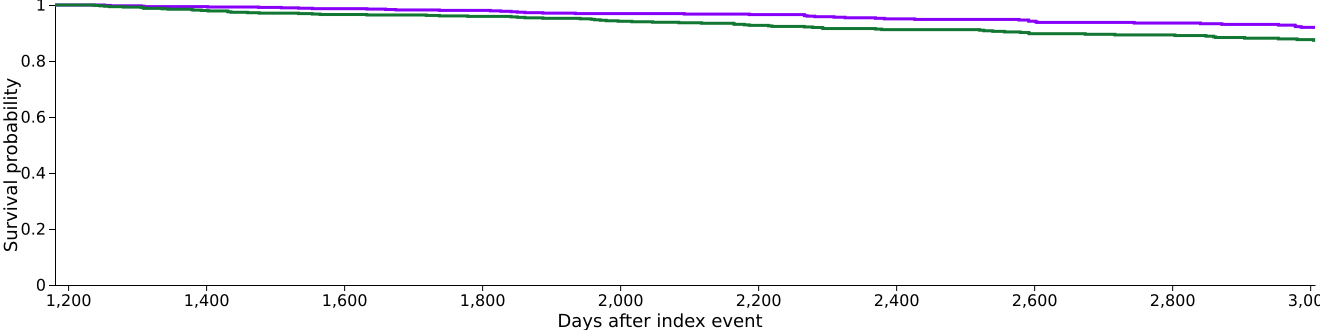 | | | | | | |
|  | |  | | | 77 patients in Cohort 1 and 105 patients in Cohort 2 were excluded from results because they had the outcome prior to the time window. | | | | | | |

### 6. **Specificity and safety analysis**

##### **Table S12. Outcome definitions for the specificity analysis.**

| AMI | | | | |
| --- | --- | --- | --- | --- |
|  | **Outcome definition** | | | |
|  | | Diagnosis | UMLS:ICD10CM:I21 | Acute myocardial infarction |
|  | **Settings for the performed analyses** | | | |
|  | | Kaplan - Meier survival analysis | | excluding patients with outcome prior to the time window |
| Stroke | | | | |
|  | **Outcome definition** | | | |
|  | | Diagnosis | UMLS:ICD10CM:I63 | Cerebral infarction |
|  | **Settings for the performed analyses** | | | |
|  | | Kaplan - Meier survival analysis | | excluding patients with outcome prior to the time window |
| Incident dialysis | | | | |
|  | **Outcome definition** | | | |
|  | | Procedure | UMLS:CPT:1012740 | Dialysis Services and Procedures |
|  | | Procedure | UMLS:CPT:90945 | Dialysis procedure other than hemodialysis (eg, peritoneal dialysis, hemofiltration, or other continuous renal replacement therapies), with single evaluation by a physician or other qualified health care professional |
|  | | Procedure | UMLS:CPT:90947 | Dialysis procedure other than hemodialysis (eg, peritoneal dialysis, hemofiltration, or other continuous renal replacement therapies) requiring repeated evaluations by a physician or other qualified health care professional, with or without substantial revision of dialysis prescription |
|  | | Procedure | UMLS:CPT:1012752 | Hemodialysis Procedures |
|  | | Procedure | UMLS:SNOMED:302497006 | Hemodialysis |
|  | | Procedure | UMLS:CPT:90935 | Hemodialysis procedure with single evaluation by a physician or other qualified health care professional |
|  | | Procedure | UMLS:CPT:90937 | Hemodialysis procedure requiring repeated evaluation(s) with or without substantial revision of dialysis prescription |
|  | **Settings for the performed analyses** | | | |
|  | | Kaplan - Meier survival analysis | | excluding patients with outcome prior to the time window |
| AKI | | | | |
|  | **Outcome definition** | | | |
|  | | Diagnosis | UMLS:ICD10CM:N17 | Acute kidney failure |
|  | **Settings for the performed analyses** | | | |
|  | | Kaplan - Meier survival analysis | | excluding patients with outcome prior to the time window |
| Heart failure | | | | |
|  | **Outcome definition** | | | |
|  | | Diagnosis | UMLS:ICD10CM:I50 | Heart failure |
|  | **Settings for the performed analyses** | | | |
|  | | Kaplan - Meier survival analysis | | excluding patients with outcome prior to the time window |

##### **Table S13. Outcome definitions for the safety analysis.**

| Urinary tract infection (UTI) | | | | |
| --- | --- | --- | --- | --- |
|  | **Outcome definition** | | | |
|  | | Diagnosis | UMLS:ICD10CM:N39.0 | Urinary tract infection, site not specified |
|  | | Diagnosis | UMLS:ICD10CM:N30 | Cystitis |
|  | | Diagnosis | UMLS:ICD10CM:N10 | Acute pyelonephritis |
|  | **Settings for the performed analyses** | | | |
|  | | Kaplan - Meier survival analysis | | excluding patients with outcome prior to the time window |
| Urogential candidiasis + UTI | | | | |
|  | **Outcome definition** | | | |
|  | | Diagnosis | UMLS:ICD10CM:B37.3 | Candidiasis of vulva and vagina |
|  | | Diagnosis | UMLS:ICD10CM:B37.4 | Candidiasis of other urogenital sites |
|  | | Diagnosis | UMLS:ICD10CM:B37.49 | Other urogenital candidiasis |
|  | | Diagnosis | UMLS:ICD10CM:N30 | Cystitis |
|  | | Diagnosis | UMLS:ICD10CM:N10 | Acute pyelonephritis |
|  | | Diagnosis | UMLS:ICD10CM:N39.0 | Urinary tract infection, site not specified |
|  | **Settings for the performed analyses** | | | |
|  | | Kaplan - Meier survival analysis | | excluding patients with outcome prior to the time window |

### 7. **Positive and negative Outcome control analyses**

##### **Table S14. Outcome definitions for the positive and negative outcome control analyses.**

| Volume depletion | | | | |
| --- | --- | --- | --- | --- |
|  | **Outcome definition** | | | |
|  | | Diagnosis | UMLS:ICD10CM:E86 | Volume depletion |
|  | **Settings for the performed analyses** | | | |
|  | | Kaplan - Meier survival analysis | | excluding patients with outcome prior to the time window |
| Proteinuria | | | | |
|  | **Outcome definition** | | | |
|  | | Diagnosis | UMLS:ICD10CM:R80 | Proteinuria |
|  | **Settings for the performed analyses** | | | |
|  | | Kaplan - Meier survival analysis | | excluding patients with outcome prior to the time window |
| Cholelithiasis | | | | |
|  | **Outcome definition** | | | |
|  | | Diagnosis | UMLS:ICD10CM:K80 | Cholelithiasis |
|  | **Settings for the performed analyses** | | | |
|  | | Kaplan - Meier survival analysis | | excluding patients with outcome prior to the time window |
| Osteoarthritis | | | | |
|  | **Outcome definition** | | | |
|  | | Diagnosis | UMLS:ICD10CM:M15-M19 | Osteoarthritis |
|  | **Settings for the performed analyses** | | | |
|  | | Kaplan - Meier survival analysis | | excluding patients with outcome prior to the time window |
| Disorders of lens | | | | |
|  | **Outcome definition** | | | |
|  | | Diagnosis | UMLS:ICD10CM:H25-H28 | Disorders of lens |
|  | **Settings for the performed analyses** | | | |
|  | | Kaplan - Meier survival analysis | | excluding patients with outcome prior to the time window |

### 8. **Positive and negative exposure control analyses**

#### Positive exposure control analysis

##### **Table S15. Code definitions and selection criteria for ARB users.**

| Ungrouped terms | | | | | |
| --- | --- | --- | --- | --- | --- |
|  | must have |  | demographics | Age | Age (at least 18 years (most recent occurrence)) |
|  | cannot have |  | diagnosis | UMLS:ICD10CM:E08-E13 | Diabetes mellitus |
| Group 1 | | | | | |
|  | **Kidney transplant** | | | | |
|  | must have | any of | diagnosis | UMLS:ICD10CM:Z94.0 | Kidney transplant status |
|  |  |  | procedure | UMLS:ICD10PCS:0TY00Z0 | Transplantation of Right Kidney, Allogeneic, Open Approach |
|  |  |  | procedure | UMLS:ICD10PCS:0TY10Z0 | Transplantation of Left Kidney, Allogeneic, Open Approach |
|  |  |  | procedure | UMLS:ICD10PCS:0TY10Z1 | Transplantation of Left Kidney, Syngeneic, Open Approach |
|  |  |  | procedure | UMLS:ICD10PCS:0TY10Z2 | Transplantation of Left Kidney, Zooplastic, Open Approach |
|  |  |  | procedure | UMLS:ICD10PCS:0TY00Z1 | Transplantation of Right Kidney, Syngeneic, Open Approach |
|  |  |  | procedure | UMLS:ICD10PCS:0TY00Z2 | Transplantation of Right Kidney, Zooplastic, Open Approach |
|  |  |  | procedure | UMLS:ICD10PCS:0TY0 | Urinary System / Transplantation / Kidney, Right |
|  |  |  | procedure | UMLS:ICD10PCS:0TY1 | Urinary System / Transplantation / Kidney, Left |
|  | date constraint | | The terms in this group occurred between Jan 1, 2015 and Jan 1, 2022 | | |
| Group 2 | | | | | |
|  | **Group 2A Kidney transplant** | | | | |
|  | must have | any of | diagnosis | UMLS:ICD10CM:Z94.0 | Kidney transplant status |
|  |  |  | procedure | UMLS:ICD10PCS:0TY00Z0 | Transplantation of Right Kidney, Allogeneic, Open Approach |
|  |  |  | procedure | UMLS:ICD10PCS:0TY10Z0 | Transplantation of Left Kidney, Allogeneic, Open Approach |
|  |  |  | procedure | UMLS:ICD10PCS:0TY10Z1 | Transplantation of Left Kidney, Syngeneic, Open Approach |
|  |  |  | procedure | UMLS:ICD10PCS:0TY10Z2 | Transplantation of Left Kidney, Zooplastic, Open Approach |
|  |  |  | procedure | UMLS:ICD10PCS:0TY00Z1 | Transplantation of Right Kidney, Syngeneic, Open Approach |
|  |  |  | procedure | UMLS:ICD10PCS:0TY00Z2 | Transplantation of Right Kidney, Zooplastic, Open Approach |
|  |  |  | procedure | UMLS:ICD10PCS:0TY0 | Urinary System / Transplantation / Kidney, Right |
|  |  |  | procedure | UMLS:ICD10PCS:0TY1 | Urinary System / Transplantation / Kidney, Left |
|  | date constraint | | The terms in this group occurred at any time | | |
|  | event relationship | | Any instance of noDialysis, Death occurred within 1 day and 999 days after the first instance of Kidney transplant | | |
|  | **Group 2B noDialysis, Death** | | | | |
|  | cannot have |  | procedure | UMLS:CPT:1012740 | Dialysis Services and Procedures |
|  |  | or | procedure | UMLS:CPT:90945 | Dialysis procedure other than hemodialysis (eg, peritoneal dialysis, hemofiltration, or other continuous renal replacement therapies), with single evaluation by a physician or other qualified health care professional |
|  |  | or | procedure | UMLS:CPT:90947 | Dialysis procedure other than hemodialysis (eg, peritoneal dialysis, hemofiltration, or other continuous renal replacement therapies) requiring repeated evaluations by a physician or other qualified health care professional, with or without substantial revision of dialysis prescription |
|  |  | or | procedure | UMLS:CPT:1012752 | Hemodialysis Procedures |
|  |  | or | procedure | UMLS:SNOMED:302497006 | Hemodialysis |
|  |  | or | procedure | UMLS:CPT:90937 | Hemodialysis procedure requiring repeated evaluation(s) with or without substantial revision of dialysis prescription |
|  |  | or | procedure | UMLS:CPT:90935 | Hemodialysis procedure with single evaluation by a physician or other qualified health care professional |
|  |  | or | demographics | Deceased | Deceased |
|  |  | or | diagnosis | UMLS:ICD10CM:R99 | Ill-defined and unknown cause of mortality |
| Group 3 | | | | | |
|  | **Group 3A Kidney transplant** | | | | |
|  | must have | any of | diagnosis | UMLS:ICD10CM:Z94.0 | Kidney transplant status |
|  |  |  | procedure | UMLS:ICD10PCS:0TY00Z0 | Transplantation of Right Kidney, Allogeneic, Open Approach |
|  |  |  | procedure | UMLS:ICD10PCS:0TY10Z0 | Transplantation of Left Kidney, Allogeneic, Open Approach |
|  |  |  | procedure | UMLS:ICD10PCS:0TY10Z1 | Transplantation of Left Kidney, Syngeneic, Open Approach |
|  |  |  | procedure | UMLS:ICD10PCS:0TY10Z2 | Transplantation of Left Kidney, Zooplastic, Open Approach |
|  |  |  | procedure | UMLS:ICD10PCS:0TY00Z1 | Transplantation of Right Kidney, Syngeneic, Open Approach |
|  |  |  | procedure | UMLS:ICD10PCS:0TY00Z2 | Transplantation of Right Kidney, Zooplastic, Open Approach |
|  |  |  | procedure | UMLS:ICD10PCS:0TY0 | Urinary System / Transplantation / Kidney, Right |
|  |  |  | procedure | UMLS:ICD10PCS:0TY1 | Urinary System / Transplantation / Kidney, Left |
|  | date constraint | | The terms in this group occurred at any time | | |
|  | event relationship | | Any instance of ARB occurred within 1000 days and 1180 days after the first instance of Kidney transplant | | |
|  | **Group 3B ARB** | | | | |
|  | must have |  | medication | NLM:VA:CV805 | ANGIOTENSIN II INHIBITOR |
| Group 4 | | | | | |
|  | **Group 4A Kidney transplant** | | | | |
|  | must have | any of | diagnosis | UMLS:ICD10CM:Z94.0 | Kidney transplant status |
|  |  |  | procedure | UMLS:ICD10PCS:0TY00Z0 | Transplantation of Right Kidney, Allogeneic, Open Approach |
|  |  |  | procedure | UMLS:ICD10PCS:0TY10Z0 | Transplantation of Left Kidney, Allogeneic, Open Approach |
|  |  |  | procedure | UMLS:ICD10PCS:0TY10Z1 | Transplantation of Left Kidney, Syngeneic, Open Approach |
|  |  |  | procedure | UMLS:ICD10PCS:0TY10Z2 | Transplantation of Left Kidney, Zooplastic, Open Approach |
|  |  |  | procedure | UMLS:ICD10PCS:0TY00Z1 | Transplantation of Right Kidney, Syngeneic, Open Approach |
|  |  |  | procedure | UMLS:ICD10PCS:0TY00Z2 | Transplantation of Right Kidney, Zooplastic, Open Approach |
|  |  |  | procedure | UMLS:ICD10PCS:0TY0 | Urinary System / Transplantation / Kidney, Right |
|  |  |  | procedure | UMLS:ICD10PCS:0TY1 | Urinary System / Transplantation / Kidney, Left |
|  | date constraint | | The terms in this group occurred at any time | | |
|  | event relationship | | Any instance of noARB occurred before or up to 999 days after the first instance of Kidney transplant | | |
|  | **Group 4B noARB** | | | | |
|  | cannot have |  | medication | NLM:VA:CV805 | ANGIOTENSIN II INHIBITOR |
| Group 5 | | | | | |
|  | **Group 5A Kidney transplant** | | | | |
|  | must have | any of | diagnosis | UMLS:ICD10CM:Z94.0 | Kidney transplant status |
|  |  |  | procedure | UMLS:ICD10PCS:0TY00Z0 | Transplantation of Right Kidney, Allogeneic, Open Approach |
|  |  |  | procedure | UMLS:ICD10PCS:0TY10Z0 | Transplantation of Left Kidney, Allogeneic, Open Approach |
|  |  |  | procedure | UMLS:ICD10PCS:0TY10Z1 | Transplantation of Left Kidney, Syngeneic, Open Approach |
|  |  |  | procedure | UMLS:ICD10PCS:0TY10Z2 | Transplantation of Left Kidney, Zooplastic, Open Approach |
|  |  |  | procedure | UMLS:ICD10PCS:0TY00Z1 | Transplantation of Right Kidney, Syngeneic, Open Approach |
|  |  |  | procedure | UMLS:ICD10PCS:0TY00Z2 | Transplantation of Right Kidney, Zooplastic, Open Approach |
|  |  |  | procedure | UMLS:ICD10PCS:0TY0 | Urinary System / Transplantation / Kidney, Right |
|  |  |  | procedure | UMLS:ICD10PCS:0TY1 | Urinary System / Transplantation / Kidney, Left |
|  | date constraint | | The terms in this group occurred at any time | | |
|  | event relationship | | Any instance of Visit occurred at least 1181 days after the first instance of Kidney transplant | | |
|  | **Group 5B Visit** | | | | |
|  | must have |  | visit | TNX:Visit | Visit |

##### **Table S16. Code definitions and selection criteria for ARB non-users.**

| Ungrouped terms | | | | | |
| --- | --- | --- | --- | --- | --- |
|  | must have |  | demographics | Age | Age (at least 18 years (most recent occurrence)) |
|  | cannot have |  | diagnosis | UMLS:ICD10CM:E08-E13 | Diabetes mellitus |
| Group 1 | | | | | |
|  | **Kidney transplant** | | | | |
|  | must have | any of | diagnosis | UMLS:ICD10CM:Z94.0 | Kidney transplant status |
|  |  |  | procedure | UMLS:ICD10PCS:0TY00Z0 | Transplantation of Right Kidney, Allogeneic, Open Approach |
|  |  |  | procedure | UMLS:ICD10PCS:0TY10Z0 | Transplantation of Left Kidney, Allogeneic, Open Approach |
|  |  |  | procedure | UMLS:ICD10PCS:0TY10Z1 | Transplantation of Left Kidney, Syngeneic, Open Approach |
|  |  |  | procedure | UMLS:ICD10PCS:0TY10Z2 | Transplantation of Left Kidney, Zooplastic, Open Approach |
|  |  |  | procedure | UMLS:ICD10PCS:0TY00Z1 | Transplantation of Right Kidney, Syngeneic, Open Approach |
|  |  |  | procedure | UMLS:ICD10PCS:0TY00Z2 | Transplantation of Right Kidney, Zooplastic, Open Approach |
|  |  |  | procedure | UMLS:ICD10PCS:0TY0 | Urinary System / Transplantation / Kidney, Right |
|  |  |  | procedure | UMLS:ICD10PCS:0TY1 | Urinary System / Transplantation / Kidney, Left |
|  | date constraint | | The terms in this group occurred between Jan 1, 2015 and Jan 1, 2022 | | |
| Group 2 | | | | | |
|  | **Group 2A Kidney transplant** | | | | |
|  | must have | any of | diagnosis | UMLS:ICD10CM:Z94.0 | Kidney transplant status |
|  |  |  | procedure | UMLS:ICD10PCS:0TY00Z0 | Transplantation of Right Kidney, Allogeneic, Open Approach |
|  |  |  | procedure | UMLS:ICD10PCS:0TY10Z0 | Transplantation of Left Kidney, Allogeneic, Open Approach |
|  |  |  | procedure | UMLS:ICD10PCS:0TY10Z1 | Transplantation of Left Kidney, Syngeneic, Open Approach |
|  |  |  | procedure | UMLS:ICD10PCS:0TY10Z2 | Transplantation of Left Kidney, Zooplastic, Open Approach |
|  |  |  | procedure | UMLS:ICD10PCS:0TY00Z1 | Transplantation of Right Kidney, Syngeneic, Open Approach |
|  |  |  | procedure | UMLS:ICD10PCS:0TY00Z2 | Transplantation of Right Kidney, Zooplastic, Open Approach |
|  |  |  | procedure | UMLS:ICD10PCS:0TY0 | Urinary System / Transplantation / Kidney, Right |
|  |  |  | procedure | UMLS:ICD10PCS:0TY1 | Urinary System / Transplantation / Kidney, Left |
|  | date constraint | | The terms in this group occurred at any time | | |
|  | event relationship | | Any instance of noDialysis, Death occurred within 1 day and 999 days after the first instance of Kidney transplant | | |
|  | **Group 2B noDialysis, Death** | | | | |
|  | cannot have |  | procedure | UMLS:CPT:1012740 | Dialysis Services and Procedures |
|  |  | or | procedure | UMLS:CPT:90945 | Dialysis procedure other than hemodialysis (eg, peritoneal dialysis, hemofiltration, or other continuous renal replacement therapies), with single evaluation by a physician or other qualified health care professional |
|  |  | or | procedure | UMLS:CPT:90947 | Dialysis procedure other than hemodialysis (eg, peritoneal dialysis, hemofiltration, or other continuous renal replacement therapies) requiring repeated evaluations by a physician or other qualified health care professional, with or without substantial revision of dialysis prescription |
|  |  | or | procedure | UMLS:CPT:1012752 | Hemodialysis Procedures |
|  |  | or | procedure | UMLS:SNOMED:302497006 | Hemodialysis |
|  |  | or | procedure | UMLS:CPT:90937 | Hemodialysis procedure requiring repeated evaluation(s) with or without substantial revision of dialysis prescription |
|  |  | or | procedure | UMLS:CPT:90935 | Hemodialysis procedure with single evaluation by a physician or other qualified health care professional |
|  |  | or | demographics | Deceased | Deceased |
|  |  | or | diagnosis | UMLS:ICD10CM:R99 | Ill-defined and unknown cause of mortality |
| Group 3 | | | | | |
|  | **Group 3A Kidney transplant** | | | | |
|  | must have | any of | diagnosis | UMLS:ICD10CM:Z94.0 | Kidney transplant status |
|  |  |  | procedure | UMLS:ICD10PCS:0TY00Z0 | Transplantation of Right Kidney, Allogeneic, Open Approach |
|  |  |  | procedure | UMLS:ICD10PCS:0TY10Z0 | Transplantation of Left Kidney, Allogeneic, Open Approach |
|  |  |  | procedure | UMLS:ICD10PCS:0TY10Z1 | Transplantation of Left Kidney, Syngeneic, Open Approach |
|  |  |  | procedure | UMLS:ICD10PCS:0TY10Z2 | Transplantation of Left Kidney, Zooplastic, Open Approach |
|  |  |  | procedure | UMLS:ICD10PCS:0TY00Z1 | Transplantation of Right Kidney, Syngeneic, Open Approach |
|  |  |  | procedure | UMLS:ICD10PCS:0TY00Z2 | Transplantation of Right Kidney, Zooplastic, Open Approach |
|  |  |  | procedure | UMLS:ICD10PCS:0TY0 | Urinary System / Transplantation / Kidney, Right |
|  |  |  | procedure | UMLS:ICD10PCS:0TY1 | Urinary System / Transplantation / Kidney, Left |
|  | date constraint | | The terms in this group occurred at any time | | |
|  | event relationship | | Any instance of ARB occurred within 1000 days and 3006 days after the first instance of Kidney transplant | | |
|  | **Group 3B noARB** | | | | |
|  | cannot have |  | medication | NLM:VA:CV805 | ANGIOTENSIN II INHIBITOR |
| Group 4 | | | | | |
|  | **Group 4A Kidney transplant** | | | | |
|  | must have | any of | diagnosis | UMLS:ICD10CM:Z94.0 | Kidney transplant status |
|  |  |  | procedure | UMLS:ICD10PCS:0TY00Z0 | Transplantation of Right Kidney, Allogeneic, Open Approach |
|  |  |  | procedure | UMLS:ICD10PCS:0TY10Z0 | Transplantation of Left Kidney, Allogeneic, Open Approach |
|  |  |  | procedure | UMLS:ICD10PCS:0TY10Z1 | Transplantation of Left Kidney, Syngeneic, Open Approach |
|  |  |  | procedure | UMLS:ICD10PCS:0TY10Z2 | Transplantation of Left Kidney, Zooplastic, Open Approach |
|  |  |  | procedure | UMLS:ICD10PCS:0TY00Z1 | Transplantation of Right Kidney, Syngeneic, Open Approach |
|  |  |  | procedure | UMLS:ICD10PCS:0TY00Z2 | Transplantation of Right Kidney, Zooplastic, Open Approach |
|  |  |  | procedure | UMLS:ICD10PCS:0TY0 | Urinary System / Transplantation / Kidney, Right |
|  |  |  | procedure | UMLS:ICD10PCS:0TY1 | Urinary System / Transplantation / Kidney, Left |
|  | date constraint | | The terms in this group occurred at any time | | |
|  | event relationship | | Any instance of noARB occurred before or up to 999 days after the first instance of Kidney transplant | | |
|  | **Group 4B noARB** | | | | |
|  | cannot have |  | medication | NLM:VA:CV805 | ANGIOTENSIN II INHIBITOR |
| Group 5 | | | | | |
|  | **Group 5A Kidney transplant** | | | | |
|  | must have | any of | diagnosis | UMLS:ICD10CM:Z94.0 | Kidney transplant status |
|  |  |  | procedure | UMLS:ICD10PCS:0TY00Z0 | Transplantation of Right Kidney, Allogeneic, Open Approach |
|  |  |  | procedure | UMLS:ICD10PCS:0TY10Z0 | Transplantation of Left Kidney, Allogeneic, Open Approach |
|  |  |  | procedure | UMLS:ICD10PCS:0TY10Z1 | Transplantation of Left Kidney, Syngeneic, Open Approach |
|  |  |  | procedure | UMLS:ICD10PCS:0TY10Z2 | Transplantation of Left Kidney, Zooplastic, Open Approach |
|  |  |  | procedure | UMLS:ICD10PCS:0TY00Z1 | Transplantation of Right Kidney, Syngeneic, Open Approach |
|  |  |  | procedure | UMLS:ICD10PCS:0TY00Z2 | Transplantation of Right Kidney, Zooplastic, Open Approach |
|  |  |  | procedure | UMLS:ICD10PCS:0TY0 | Urinary System / Transplantation / Kidney, Right |
|  |  |  | procedure | UMLS:ICD10PCS:0TY1 | Urinary System / Transplantation / Kidney, Left |
|  | date constraint | | The terms in this group occurred at any time | | |
|  | event relationship | | Any instance of Visit occurred at least 1181 days after the first instance of Kidney transplant | | |
|  | **Group 5B Visit** | | | | |
|  | must have |  | visit | TNX:Visit | Visit |

##### **Table S17. Code definitions and selection criteria for aspirin users.**

| Ungrouped terms | | | | | |
| --- | --- | --- | --- | --- | --- |
|  | must have |  | demographics | Age | Age (at least 18 years (most recent occurrence)) |
|  | cannot have |  | diagnosis | UMLS:ICD10CM:E08-E13 | Diabetes mellitus |
| Group 1 | | | | | |
|  | **Kidney transplant** | | | | |
|  | must have | any of | diagnosis | UMLS:ICD10CM:Z94.0 | Kidney transplant status |
|  |  |  | procedure | UMLS:ICD10PCS:0TY00Z0 | Transplantation of Right Kidney, Allogeneic, Open Approach |
|  |  |  | procedure | UMLS:ICD10PCS:0TY10Z0 | Transplantation of Left Kidney, Allogeneic, Open Approach |
|  |  |  | procedure | UMLS:ICD10PCS:0TY10Z1 | Transplantation of Left Kidney, Syngeneic, Open Approach |
|  |  |  | procedure | UMLS:ICD10PCS:0TY10Z2 | Transplantation of Left Kidney, Zooplastic, Open Approach |
|  |  |  | procedure | UMLS:ICD10PCS:0TY00Z1 | Transplantation of Right Kidney, Syngeneic, Open Approach |
|  |  |  | procedure | UMLS:ICD10PCS:0TY00Z2 | Transplantation of Right Kidney, Zooplastic, Open Approach |
|  |  |  | procedure | UMLS:ICD10PCS:0TY0 | Urinary System / Transplantation / Kidney, Right |
|  |  |  | procedure | UMLS:ICD10PCS:0TY1 | Urinary System / Transplantation / Kidney, Left |
|  | date constraint | | The terms in this group occurred between Jan 1, 2015 and Jan 1, 2022 | | |
| Group 2 | | | | | |
|  | **Group 2A Kidney transplant** | | | | |
|  | must have | any of | diagnosis | UMLS:ICD10CM:Z94.0 | Kidney transplant status |
|  |  |  | procedure | UMLS:ICD10PCS:0TY00Z0 | Transplantation of Right Kidney, Allogeneic, Open Approach |
|  |  |  | procedure | UMLS:ICD10PCS:0TY10Z0 | Transplantation of Left Kidney, Allogeneic, Open Approach |
|  |  |  | procedure | UMLS:ICD10PCS:0TY10Z1 | Transplantation of Left Kidney, Syngeneic, Open Approach |
|  |  |  | procedure | UMLS:ICD10PCS:0TY10Z2 | Transplantation of Left Kidney, Zooplastic, Open Approach |
|  |  |  | procedure | UMLS:ICD10PCS:0TY00Z1 | Transplantation of Right Kidney, Syngeneic, Open Approach |
|  |  |  | procedure | UMLS:ICD10PCS:0TY00Z2 | Transplantation of Right Kidney, Zooplastic, Open Approach |
|  |  |  | procedure | UMLS:ICD10PCS:0TY0 | Urinary System / Transplantation / Kidney, Right |
|  |  |  | procedure | UMLS:ICD10PCS:0TY1 | Urinary System / Transplantation / Kidney, Left |
|  | date constraint | | The terms in this group occurred at any time | | |
|  | event relationship | | Any instance of noDialysis, Death occurred within 1 day and 999 days after the first instance of Kidney transplant | | |
|  | **Group 2B noDialysis, Death** | | | | |
|  | cannot have |  | procedure | UMLS:CPT:1012740 | Dialysis Services and Procedures |
|  |  | or | procedure | UMLS:CPT:90945 | Dialysis procedure other than hemodialysis (eg, peritoneal dialysis, hemofiltration, or other continuous renal replacement therapies), with single evaluation by a physician or other qualified health care professional |
|  |  | or | procedure | UMLS:CPT:90947 | Dialysis procedure other than hemodialysis (eg, peritoneal dialysis, hemofiltration, or other continuous renal replacement therapies) requiring repeated evaluations by a physician or other qualified health care professional, with or without substantial revision of dialysis prescription |
|  |  | or | procedure | UMLS:CPT:1012752 | Hemodialysis Procedures |
|  |  | or | procedure | UMLS:SNOMED:302497006 | Hemodialysis |
|  |  | or | procedure | UMLS:CPT:90937 | Hemodialysis procedure requiring repeated evaluation(s) with or without substantial revision of dialysis prescription |
|  |  | or | procedure | UMLS:CPT:90935 | Hemodialysis procedure with single evaluation by a physician or other qualified health care professional |
|  |  | or | demographics | Deceased | Deceased |
|  |  | or | diagnosis | UMLS:ICD10CM:R99 | Ill-defined and unknown cause of mortality |
| Group 3 | | | | | |
|  | **Group 3A Kidney transplant** | | | | |
|  | must have | any of | diagnosis | UMLS:ICD10CM:Z94.0 | Kidney transplant status |
|  |  |  | procedure | UMLS:ICD10PCS:0TY00Z0 | Transplantation of Right Kidney, Allogeneic, Open Approach |
|  |  |  | procedure | UMLS:ICD10PCS:0TY10Z0 | Transplantation of Left Kidney, Allogeneic, Open Approach |
|  |  |  | procedure | UMLS:ICD10PCS:0TY10Z1 | Transplantation of Left Kidney, Syngeneic, Open Approach |
|  |  |  | procedure | UMLS:ICD10PCS:0TY10Z2 | Transplantation of Left Kidney, Zooplastic, Open Approach |
|  |  |  | procedure | UMLS:ICD10PCS:0TY00Z1 | Transplantation of Right Kidney, Syngeneic, Open Approach |
|  |  |  | procedure | UMLS:ICD10PCS:0TY00Z2 | Transplantation of Right Kidney, Zooplastic, Open Approach |
|  |  |  | procedure | UMLS:ICD10PCS:0TY0 | Urinary System / Transplantation / Kidney, Right |
|  |  |  | procedure | UMLS:ICD10PCS:0TY1 | Urinary System / Transplantation / Kidney, Left |
|  | date constraint | | The terms in this group occurred at any time | | |
|  | event relationship | | Any instance of Aspirin occurred within 1000 days and 1180 days after the first instance of Kidney transplant | | |
|  | **Group 3B Aspirin** | | | | |
|  | must have |  | medication | NLM:RXNORM:1191 | aspirin |
| Group 4 | | | | | |
|  | **Group 4A Kidney transplant** | | | | |
|  | must have | any of | diagnosis | UMLS:ICD10CM:Z94.0 | Kidney transplant status |
|  |  |  | procedure | UMLS:ICD10PCS:0TY00Z0 | Transplantation of Right Kidney, Allogeneic, Open Approach |
|  |  |  | procedure | UMLS:ICD10PCS:0TY10Z0 | Transplantation of Left Kidney, Allogeneic, Open Approach |
|  |  |  | procedure | UMLS:ICD10PCS:0TY10Z1 | Transplantation of Left Kidney, Syngeneic, Open Approach |
|  |  |  | procedure | UMLS:ICD10PCS:0TY10Z2 | Transplantation of Left Kidney, Zooplastic, Open Approach |
|  |  |  | procedure | UMLS:ICD10PCS:0TY00Z1 | Transplantation of Right Kidney, Syngeneic, Open Approach |
|  |  |  | procedure | UMLS:ICD10PCS:0TY00Z2 | Transplantation of Right Kidney, Zooplastic, Open Approach |
|  |  |  | procedure | UMLS:ICD10PCS:0TY0 | Urinary System / Transplantation / Kidney, Right |
|  |  |  | procedure | UMLS:ICD10PCS:0TY1 | Urinary System / Transplantation / Kidney, Left |
|  | date constraint | | The terms in this group occurred at any time | | |
|  | event relationship | | Any instance of noAspirin occurred before or up to 999 days after the first instance of Kidney transplant | | |
|  | **Group 4B noAspirin** | | | | |
|  | cannot have |  | medication | NLM:RXNORM:1191 | aspirin |
| Group 5 | | | | | |
|  | **Group 5A Kidney transplant** | | | | |
|  | must have | any of | diagnosis | UMLS:ICD10CM:Z94.0 | Kidney transplant status |
|  |  |  | procedure | UMLS:ICD10PCS:0TY00Z0 | Transplantation of Right Kidney, Allogeneic, Open Approach |
|  |  |  | procedure | UMLS:ICD10PCS:0TY10Z0 | Transplantation of Left Kidney, Allogeneic, Open Approach |
|  |  |  | procedure | UMLS:ICD10PCS:0TY10Z1 | Transplantation of Left Kidney, Syngeneic, Open Approach |
|  |  |  | procedure | UMLS:ICD10PCS:0TY10Z2 | Transplantation of Left Kidney, Zooplastic, Open Approach |
|  |  |  | procedure | UMLS:ICD10PCS:0TY00Z1 | Transplantation of Right Kidney, Syngeneic, Open Approach |
|  |  |  | procedure | UMLS:ICD10PCS:0TY00Z2 | Transplantation of Right Kidney, Zooplastic, Open Approach |
|  |  |  | procedure | UMLS:ICD10PCS:0TY0 | Urinary System / Transplantation / Kidney, Right |
|  |  |  | procedure | UMLS:ICD10PCS:0TY1 | Urinary System / Transplantation / Kidney, Left |
|  | date constraint | | The terms in this group occurred at any time | | |
|  | event relationship | | Any instance of Visit occurred at least 1181 days after the first instance of Kidney transplant | | |
|  | **Group 5B Visit** | | | | |
|  | must have |  | visit | TNX:Visit | Visit |

##### **Table S18. Code definitions and selection criteria for aspirin non-users.**

| Ungrouped terms | | | | | |
| --- | --- | --- | --- | --- | --- |
|  | must have |  | demographics | Age | Age (at least 18 years (most recent occurrence)) |
|  | cannot have |  | diagnosis | UMLS:ICD10CM:E08-E13 | Diabetes mellitus |
| Group 1 | | | | | |
|  | **Kidney transplant** | | | | |
|  | must have | any of | diagnosis | UMLS:ICD10CM:Z94.0 | Kidney transplant status |
|  |  |  | procedure | UMLS:ICD10PCS:0TY00Z0 | Transplantation of Right Kidney, Allogeneic, Open Approach |
|  |  |  | procedure | UMLS:ICD10PCS:0TY10Z0 | Transplantation of Left Kidney, Allogeneic, Open Approach |
|  |  |  | procedure | UMLS:ICD10PCS:0TY10Z1 | Transplantation of Left Kidney, Syngeneic, Open Approach |
|  |  |  | procedure | UMLS:ICD10PCS:0TY10Z2 | Transplantation of Left Kidney, Zooplastic, Open Approach |
|  |  |  | procedure | UMLS:ICD10PCS:0TY00Z1 | Transplantation of Right Kidney, Syngeneic, Open Approach |
|  |  |  | procedure | UMLS:ICD10PCS:0TY00Z2 | Transplantation of Right Kidney, Zooplastic, Open Approach |
|  |  |  | procedure | UMLS:ICD10PCS:0TY0 | Urinary System / Transplantation / Kidney, Right |
|  |  |  | procedure | UMLS:ICD10PCS:0TY1 | Urinary System / Transplantation / Kidney, Left |
|  | date constraint | | The terms in this group occurred between Jan 1, 2015 and Jan 1, 2022 | | |
| Group 2 | | | | | |
|  | **Group 2A Kidney transplant** | | | | |
|  | must have | any of | diagnosis | UMLS:ICD10CM:Z94.0 | Kidney transplant status |
|  |  |  | procedure | UMLS:ICD10PCS:0TY00Z0 | Transplantation of Right Kidney, Allogeneic, Open Approach |
|  |  |  | procedure | UMLS:ICD10PCS:0TY10Z0 | Transplantation of Left Kidney, Allogeneic, Open Approach |
|  |  |  | procedure | UMLS:ICD10PCS:0TY10Z1 | Transplantation of Left Kidney, Syngeneic, Open Approach |
|  |  |  | procedure | UMLS:ICD10PCS:0TY10Z2 | Transplantation of Left Kidney, Zooplastic, Open Approach |
|  |  |  | procedure | UMLS:ICD10PCS:0TY00Z1 | Transplantation of Right Kidney, Syngeneic, Open Approach |
|  |  |  | procedure | UMLS:ICD10PCS:0TY00Z2 | Transplantation of Right Kidney, Zooplastic, Open Approach |
|  |  |  | procedure | UMLS:ICD10PCS:0TY0 | Urinary System / Transplantation / Kidney, Right |
|  |  |  | procedure | UMLS:ICD10PCS:0TY1 | Urinary System / Transplantation / Kidney, Left |
|  | date constraint | | The terms in this group occurred at any time | | |
|  | event relationship | | Any instance of noDialysis, Death occurred within 1 day and 999 days after the first instance of Kidney transplant | | |
|  | **Group 2B noDialysis, Death** | | | | |
|  | cannot have |  | procedure | UMLS:CPT:1012740 | Dialysis Services and Procedures |
|  |  | or | procedure | UMLS:CPT:90945 | Dialysis procedure other than hemodialysis (eg, peritoneal dialysis, hemofiltration, or other continuous renal replacement therapies), with single evaluation by a physician or other qualified health care professional |
|  |  | or | procedure | UMLS:CPT:90947 | Dialysis procedure other than hemodialysis (eg, peritoneal dialysis, hemofiltration, or other continuous renal replacement therapies) requiring repeated evaluations by a physician or other qualified health care professional, with or without substantial revision of dialysis prescription |
|  |  | or | procedure | UMLS:CPT:1012752 | Hemodialysis Procedures |
|  |  | or | procedure | UMLS:SNOMED:302497006 | Hemodialysis |
|  |  | or | procedure | UMLS:CPT:90937 | Hemodialysis procedure requiring repeated evaluation(s) with or without substantial revision of dialysis prescription |
|  |  | or | procedure | UMLS:CPT:90935 | Hemodialysis procedure with single evaluation by a physician or other qualified health care professional |
|  |  | or | demographics | Deceased | Deceased |
|  |  | or | diagnosis | UMLS:ICD10CM:R99 | Ill-defined and unknown cause of mortality |
| Group 3 | | | | | |
|  | **Group 3A Kidney transplant** | | | | |
|  | must have | any of | diagnosis | UMLS:ICD10CM:Z94.0 | Kidney transplant status |
|  |  |  | procedure | UMLS:ICD10PCS:0TY00Z0 | Transplantation of Right Kidney, Allogeneic, Open Approach |
|  |  |  | procedure | UMLS:ICD10PCS:0TY10Z0 | Transplantation of Left Kidney, Allogeneic, Open Approach |
|  |  |  | procedure | UMLS:ICD10PCS:0TY10Z1 | Transplantation of Left Kidney, Syngeneic, Open Approach |
|  |  |  | procedure | UMLS:ICD10PCS:0TY10Z2 | Transplantation of Left Kidney, Zooplastic, Open Approach |
|  |  |  | procedure | UMLS:ICD10PCS:0TY00Z1 | Transplantation of Right Kidney, Syngeneic, Open Approach |
|  |  |  | procedure | UMLS:ICD10PCS:0TY00Z2 | Transplantation of Right Kidney, Zooplastic, Open Approach |
|  |  |  | procedure | UMLS:ICD10PCS:0TY0 | Urinary System / Transplantation / Kidney, Right |
|  |  |  | procedure | UMLS:ICD10PCS:0TY1 | Urinary System / Transplantation / Kidney, Left |
|  | date constraint | | The terms in this group occurred at any time | | |
|  | event relationship | | Any instance of noAspirin occurred within 1000 days and 3006 days after the first instance of Kidney transplant | | |
|  | **Group 3B noAspirin** | | | | |
|  | cannot have |  | medication | NLM:RXNORM:1191 | aspirin |
| Group 4 | | | | | |
|  | **Group 4A Kidney transplant** | | | | |
|  | must have | any of | diagnosis | UMLS:ICD10CM:Z94.0 | Kidney transplant status |
|  |  |  | procedure | UMLS:ICD10PCS:0TY00Z0 | Transplantation of Right Kidney, Allogeneic, Open Approach |
|  |  |  | procedure | UMLS:ICD10PCS:0TY10Z0 | Transplantation of Left Kidney, Allogeneic, Open Approach |
|  |  |  | procedure | UMLS:ICD10PCS:0TY10Z1 | Transplantation of Left Kidney, Syngeneic, Open Approach |
|  |  |  | procedure | UMLS:ICD10PCS:0TY10Z2 | Transplantation of Left Kidney, Zooplastic, Open Approach |
|  |  |  | procedure | UMLS:ICD10PCS:0TY00Z1 | Transplantation of Right Kidney, Syngeneic, Open Approach |
|  |  |  | procedure | UMLS:ICD10PCS:0TY00Z2 | Transplantation of Right Kidney, Zooplastic, Open Approach |
|  |  |  | procedure | UMLS:ICD10PCS:0TY0 | Urinary System / Transplantation / Kidney, Right |
|  |  |  | procedure | UMLS:ICD10PCS:0TY1 | Urinary System / Transplantation / Kidney, Left |
|  | date constraint | | The terms in this group occurred at any time | | |
|  | event relationship | | Any instance of noAspirin occurred before or up to 999 days after the first instance of Kidney transplant | | |
|  | **Group 4B noAspirin** | | | | |
|  | cannot have |  | medication | NLM:RXNORM:1191 | aspirin |
| Group 5 | | | | | |
|  | **Group 5A Kidney transplant** | | | | |
|  | must have | any of | diagnosis | UMLS:ICD10CM:Z94.0 | Kidney transplant status |
|  |  |  | procedure | UMLS:ICD10PCS:0TY00Z0 | Transplantation of Right Kidney, Allogeneic, Open Approach |
|  |  |  | procedure | UMLS:ICD10PCS:0TY10Z0 | Transplantation of Left Kidney, Allogeneic, Open Approach |
|  |  |  | procedure | UMLS:ICD10PCS:0TY10Z1 | Transplantation of Left Kidney, Syngeneic, Open Approach |
|  |  |  | procedure | UMLS:ICD10PCS:0TY10Z2 | Transplantation of Left Kidney, Zooplastic, Open Approach |
|  |  |  | procedure | UMLS:ICD10PCS:0TY00Z1 | Transplantation of Right Kidney, Syngeneic, Open Approach |
|  |  |  | procedure | UMLS:ICD10PCS:0TY00Z2 | Transplantation of Right Kidney, Zooplastic, Open Approach |
|  |  |  | procedure | UMLS:ICD10PCS:0TY0 | Urinary System / Transplantation / Kidney, Right |
|  |  |  | procedure | UMLS:ICD10PCS:0TY1 | Urinary System / Transplantation / Kidney, Left |
|  | date constraint | | The terms in this group occurred at any time | | |
|  | event relationship | | Any instance of Visit occurred at least 1181 days after the first instance of Kidney transplant | | |
|  | **Group 5B Visit** | | | | |
|  | must have |  | visit | TNX:Visit | Visit |

##### **Table S19. Code definitions and selection criteria for statin users.**

| Ungrouped terms | | | | | |
| --- | --- | --- | --- | --- | --- |
|  | must have |  | demographics | Age | Age (at least 18 years (most recent occurrence)) |
|  | cannot have |  | diagnosis | UMLS:ICD10CM:E08-E13 | Diabetes mellitus |
| Group 1 | | | | | |
|  | **Kidney transplant** | | | | |
|  | must have | any of | diagnosis | UMLS:ICD10CM:Z94.0 | Kidney transplant status |
|  |  |  | procedure | UMLS:ICD10PCS:0TY00Z0 | Transplantation of Right Kidney, Allogeneic, Open Approach |
|  |  |  | procedure | UMLS:ICD10PCS:0TY10Z0 | Transplantation of Left Kidney, Allogeneic, Open Approach |
|  |  |  | procedure | UMLS:ICD10PCS:0TY10Z1 | Transplantation of Left Kidney, Syngeneic, Open Approach |
|  |  |  | procedure | UMLS:ICD10PCS:0TY10Z2 | Transplantation of Left Kidney, Zooplastic, Open Approach |
|  |  |  | procedure | UMLS:ICD10PCS:0TY00Z1 | Transplantation of Right Kidney, Syngeneic, Open Approach |
|  |  |  | procedure | UMLS:ICD10PCS:0TY00Z2 | Transplantation of Right Kidney, Zooplastic, Open Approach |
|  |  |  | procedure | UMLS:ICD10PCS:0TY0 | Urinary System / Transplantation / Kidney, Right |
|  |  |  | procedure | UMLS:ICD10PCS:0TY1 | Urinary System / Transplantation / Kidney, Left |
|  | date constraint | | The terms in this group occurred between Jan 1, 2015 and Jan 1, 2022 | | |
| Group 2 | | | | | |
|  | **Group 2A Kidney transplant** | | | | |
|  | must have | any of | diagnosis | UMLS:ICD10CM:Z94.0 | Kidney transplant status |
|  |  |  | procedure | UMLS:ICD10PCS:0TY00Z0 | Transplantation of Right Kidney, Allogeneic, Open Approach |
|  |  |  | procedure | UMLS:ICD10PCS:0TY10Z0 | Transplantation of Left Kidney, Allogeneic, Open Approach |
|  |  |  | procedure | UMLS:ICD10PCS:0TY10Z1 | Transplantation of Left Kidney, Syngeneic, Open Approach |
|  |  |  | procedure | UMLS:ICD10PCS:0TY10Z2 | Transplantation of Left Kidney, Zooplastic, Open Approach |
|  |  |  | procedure | UMLS:ICD10PCS:0TY00Z1 | Transplantation of Right Kidney, Syngeneic, Open Approach |
|  |  |  | procedure | UMLS:ICD10PCS:0TY00Z2 | Transplantation of Right Kidney, Zooplastic, Open Approach |
|  |  |  | procedure | UMLS:ICD10PCS:0TY0 | Urinary System / Transplantation / Kidney, Right |
|  |  |  | procedure | UMLS:ICD10PCS:0TY1 | Urinary System / Transplantation / Kidney, Left |
|  | date constraint | | The terms in this group occurred at any time | | |
|  | event relationship | | Any instance of noDialysis, Death occurred within 1 day and 999 days after the first instance of Kidney transplant | | |
|  | **Group 2B noDialysis, Death** | | | | |
|  | cannot have |  | procedure | UMLS:CPT:1012740 | Dialysis Services and Procedures |
|  |  | or | procedure | UMLS:CPT:90945 | Dialysis procedure other than hemodialysis (eg, peritoneal dialysis, hemofiltration, or other continuous renal replacement therapies), with single evaluation by a physician or other qualified health care professional |
|  |  | or | procedure | UMLS:CPT:90947 | Dialysis procedure other than hemodialysis (eg, peritoneal dialysis, hemofiltration, or other continuous renal replacement therapies) requiring repeated evaluations by a physician or other qualified health care professional, with or without substantial revision of dialysis prescription |
|  |  | or | procedure | UMLS:CPT:1012752 | Hemodialysis Procedures |
|  |  | or | procedure | UMLS:SNOMED:302497006 | Hemodialysis |
|  |  | or | procedure | UMLS:CPT:90937 | Hemodialysis procedure requiring repeated evaluation(s) with or without substantial revision of dialysis prescription |
|  |  | or | procedure | UMLS:CPT:90935 | Hemodialysis procedure with single evaluation by a physician or other qualified health care professional |
|  |  | or | demographics | Deceased | Deceased |
|  |  | or | diagnosis | UMLS:ICD10CM:R99 | Ill-defined and unknown cause of mortality |
| Group 3 | | | | | |
|  | **Group 3A Kidney transplant** | | | | |
|  | must have | any of | diagnosis | UMLS:ICD10CM:Z94.0 | Kidney transplant status |
|  |  |  | procedure | UMLS:ICD10PCS:0TY00Z0 | Transplantation of Right Kidney, Allogeneic, Open Approach |
|  |  |  | procedure | UMLS:ICD10PCS:0TY10Z0 | Transplantation of Left Kidney, Allogeneic, Open Approach |
|  |  |  | procedure | UMLS:ICD10PCS:0TY10Z1 | Transplantation of Left Kidney, Syngeneic, Open Approach |
|  |  |  | procedure | UMLS:ICD10PCS:0TY10Z2 | Transplantation of Left Kidney, Zooplastic, Open Approach |
|  |  |  | procedure | UMLS:ICD10PCS:0TY00Z1 | Transplantation of Right Kidney, Syngeneic, Open Approach |
|  |  |  | procedure | UMLS:ICD10PCS:0TY00Z2 | Transplantation of Right Kidney, Zooplastic, Open Approach |
|  |  |  | procedure | UMLS:ICD10PCS:0TY0 | Urinary System / Transplantation / Kidney, Right |
|  |  |  | procedure | UMLS:ICD10PCS:0TY1 | Urinary System / Transplantation / Kidney, Left |
|  | date constraint | | The terms in this group occurred at any time | | |
|  | event relationship | | Any instance of Statins occurred within 1000 days and 1180 days after the first instance of Kidney transplant | | |
|  | **Group 3B Statins** | | | | |
|  | must have |  | medication | NLM:ATC:C10AA | HMG CoA reductase inhibitors |
| Group 4 | | | | | |
|  | **Group 4A Kidney transplant** | | | | |
|  | must have | any of | diagnosis | UMLS:ICD10CM:Z94.0 | Kidney transplant status |
|  |  |  | procedure | UMLS:ICD10PCS:0TY00Z0 | Transplantation of Right Kidney, Allogeneic, Open Approach |
|  |  |  | procedure | UMLS:ICD10PCS:0TY10Z0 | Transplantation of Left Kidney, Allogeneic, Open Approach |
|  |  |  | procedure | UMLS:ICD10PCS:0TY10Z1 | Transplantation of Left Kidney, Syngeneic, Open Approach |
|  |  |  | procedure | UMLS:ICD10PCS:0TY10Z2 | Transplantation of Left Kidney, Zooplastic, Open Approach |
|  |  |  | procedure | UMLS:ICD10PCS:0TY00Z1 | Transplantation of Right Kidney, Syngeneic, Open Approach |
|  |  |  | procedure | UMLS:ICD10PCS:0TY00Z2 | Transplantation of Right Kidney, Zooplastic, Open Approach |
|  |  |  | procedure | UMLS:ICD10PCS:0TY0 | Urinary System / Transplantation / Kidney, Right |
|  |  |  | procedure | UMLS:ICD10PCS:0TY1 | Urinary System / Transplantation / Kidney, Left |
|  | date constraint | | The terms in this group occurred at any time | | |
|  | event relationship | | Any instance of noStatins occurred before or up to 999 days after the first instance of Kidney transplant | | |
|  | **Group 4B noStatins** | | | | |
|  | cannot have |  | medication | NLM:ATC:C10AA | HMG CoA reductase inhibitors |
| Group 5 | | | | | |
|  | **Group 5A Kidney transplant** | | | | |
|  | must have | any of | diagnosis | UMLS:ICD10CM:Z94.0 | Kidney transplant status |
|  |  |  | procedure | UMLS:ICD10PCS:0TY00Z0 | Transplantation of Right Kidney, Allogeneic, Open Approach |
|  |  |  | procedure | UMLS:ICD10PCS:0TY10Z0 | Transplantation of Left Kidney, Allogeneic, Open Approach |
|  |  |  | procedure | UMLS:ICD10PCS:0TY10Z1 | Transplantation of Left Kidney, Syngeneic, Open Approach |
|  |  |  | procedure | UMLS:ICD10PCS:0TY10Z2 | Transplantation of Left Kidney, Zooplastic, Open Approach |
|  |  |  | procedure | UMLS:ICD10PCS:0TY00Z1 | Transplantation of Right Kidney, Syngeneic, Open Approach |
|  |  |  | procedure | UMLS:ICD10PCS:0TY00Z2 | Transplantation of Right Kidney, Zooplastic, Open Approach |
|  |  |  | procedure | UMLS:ICD10PCS:0TY0 | Urinary System / Transplantation / Kidney, Right |
|  |  |  | procedure | UMLS:ICD10PCS:0TY1 | Urinary System / Transplantation / Kidney, Left |
|  | date constraint | | The terms in this group occurred at any time | | |
|  | event relationship | | Any instance of Visit occurred at least 1181 days after the first instance of Kidney transplant | | |
|  | **Group 5B Visit** | | | | |
|  | must have |  | visit | TNX:Visit | Visit |

##### **Table S20. Code definitions and selection criteria for statin non-users.**

| Ungrouped terms | | | | | |
| --- | --- | --- | --- | --- | --- |
|  | must have |  | demographics | Age | Age (at least 18 years (most recent occurrence)) |
|  | cannot have |  | diagnosis | UMLS:ICD10CM:E08-E13 | Diabetes mellitus |
| Group 1 | | | | | |
|  | **Kidney transplant** | | | | |
|  | must have | any of | diagnosis | UMLS:ICD10CM:Z94.0 | Kidney transplant status |
|  |  |  | procedure | UMLS:ICD10PCS:0TY00Z0 | Transplantation of Right Kidney, Allogeneic, Open Approach |
|  |  |  | procedure | UMLS:ICD10PCS:0TY10Z0 | Transplantation of Left Kidney, Allogeneic, Open Approach |
|  |  |  | procedure | UMLS:ICD10PCS:0TY10Z1 | Transplantation of Left Kidney, Syngeneic, Open Approach |
|  |  |  | procedure | UMLS:ICD10PCS:0TY10Z2 | Transplantation of Left Kidney, Zooplastic, Open Approach |
|  |  |  | procedure | UMLS:ICD10PCS:0TY00Z1 | Transplantation of Right Kidney, Syngeneic, Open Approach |
|  |  |  | procedure | UMLS:ICD10PCS:0TY00Z2 | Transplantation of Right Kidney, Zooplastic, Open Approach |
|  |  |  | procedure | UMLS:ICD10PCS:0TY0 | Urinary System / Transplantation / Kidney, Right |
|  |  |  | procedure | UMLS:ICD10PCS:0TY1 | Urinary System / Transplantation / Kidney, Left |
|  | date constraint | | The terms in this group occurred between Jan 1, 2015 and Jan 1, 2022 | | |
| Group 2 | | | | | |
|  | **Group 2A Kidney transplant** | | | | |
|  | must have | any of | diagnosis | UMLS:ICD10CM:Z94.0 | Kidney transplant status |
|  |  |  | procedure | UMLS:ICD10PCS:0TY00Z0 | Transplantation of Right Kidney, Allogeneic, Open Approach |
|  |  |  | procedure | UMLS:ICD10PCS:0TY10Z0 | Transplantation of Left Kidney, Allogeneic, Open Approach |
|  |  |  | procedure | UMLS:ICD10PCS:0TY10Z1 | Transplantation of Left Kidney, Syngeneic, Open Approach |
|  |  |  | procedure | UMLS:ICD10PCS:0TY10Z2 | Transplantation of Left Kidney, Zooplastic, Open Approach |
|  |  |  | procedure | UMLS:ICD10PCS:0TY00Z1 | Transplantation of Right Kidney, Syngeneic, Open Approach |
|  |  |  | procedure | UMLS:ICD10PCS:0TY00Z2 | Transplantation of Right Kidney, Zooplastic, Open Approach |
|  |  |  | procedure | UMLS:ICD10PCS:0TY0 | Urinary System / Transplantation / Kidney, Right |
|  |  |  | procedure | UMLS:ICD10PCS:0TY1 | Urinary System / Transplantation / Kidney, Left |
|  | date constraint | | The terms in this group occurred at any time | | |
|  | event relationship | | Any instance of noDialysis, Death occurred within 1 day and 999 days after the first instance of Kidney transplant | | |
|  | **Group 2B noDialysis, Death** | | | | |
|  | cannot have |  | procedure | UMLS:CPT:1012740 | Dialysis Services and Procedures |
|  |  | or | procedure | UMLS:CPT:90945 | Dialysis procedure other than hemodialysis (eg, peritoneal dialysis, hemofiltration, or other continuous renal replacement therapies), with single evaluation by a physician or other qualified health care professional |
|  |  | or | procedure | UMLS:CPT:90947 | Dialysis procedure other than hemodialysis (eg, peritoneal dialysis, hemofiltration, or other continuous renal replacement therapies) requiring repeated evaluations by a physician or other qualified health care professional, with or without substantial revision of dialysis prescription |
|  |  | or | procedure | UMLS:CPT:1012752 | Hemodialysis Procedures |
|  |  | or | procedure | UMLS:SNOMED:302497006 | Hemodialysis |
|  |  | or | procedure | UMLS:CPT:90937 | Hemodialysis procedure requiring repeated evaluation(s) with or without substantial revision of dialysis prescription |
|  |  | or | procedure | UMLS:CPT:90935 | Hemodialysis procedure with single evaluation by a physician or other qualified health care professional |
|  |  | or | demographics | Deceased | Deceased |
|  |  | or | diagnosis | UMLS:ICD10CM:R99 | Ill-defined and unknown cause of mortality |
| Group 3 | | | | | |
|  | **Group 3A Kidney transplant** | | | | |
|  | must have | any of | diagnosis | UMLS:ICD10CM:Z94.0 | Kidney transplant status |
|  |  |  | procedure | UMLS:ICD10PCS:0TY00Z0 | Transplantation of Right Kidney, Allogeneic, Open Approach |
|  |  |  | procedure | UMLS:ICD10PCS:0TY10Z0 | Transplantation of Left Kidney, Allogeneic, Open Approach |
|  |  |  | procedure | UMLS:ICD10PCS:0TY10Z1 | Transplantation of Left Kidney, Syngeneic, Open Approach |
|  |  |  | procedure | UMLS:ICD10PCS:0TY10Z2 | Transplantation of Left Kidney, Zooplastic, Open Approach |
|  |  |  | procedure | UMLS:ICD10PCS:0TY00Z1 | Transplantation of Right Kidney, Syngeneic, Open Approach |
|  |  |  | procedure | UMLS:ICD10PCS:0TY00Z2 | Transplantation of Right Kidney, Zooplastic, Open Approach |
|  |  |  | procedure | UMLS:ICD10PCS:0TY0 | Urinary System / Transplantation / Kidney, Right |
|  |  |  | procedure | UMLS:ICD10PCS:0TY1 | Urinary System / Transplantation / Kidney, Left |
|  | date constraint | | The terms in this group occurred at any time | | |
|  | event relationship | | Any instance of noStatins occurred within 1000 days and 3006 days after the first instance of Kidney transplant | | |
|  | **Group 3B noStatins** | | | | |
|  | cannot have |  | medication | NLM:ATC:C10AA | HMG CoA reductase inhibitors |
| Group 4 | | | | | |
|  | **Group 4A Kidney transplant** | | | | |
|  | must have | any of | diagnosis | UMLS:ICD10CM:Z94.0 | Kidney transplant status |
|  |  |  | procedure | UMLS:ICD10PCS:0TY00Z0 | Transplantation of Right Kidney, Allogeneic, Open Approach |
|  |  |  | procedure | UMLS:ICD10PCS:0TY10Z0 | Transplantation of Left Kidney, Allogeneic, Open Approach |
|  |  |  | procedure | UMLS:ICD10PCS:0TY10Z1 | Transplantation of Left Kidney, Syngeneic, Open Approach |
|  |  |  | procedure | UMLS:ICD10PCS:0TY10Z2 | Transplantation of Left Kidney, Zooplastic, Open Approach |
|  |  |  | procedure | UMLS:ICD10PCS:0TY00Z1 | Transplantation of Right Kidney, Syngeneic, Open Approach |
|  |  |  | procedure | UMLS:ICD10PCS:0TY00Z2 | Transplantation of Right Kidney, Zooplastic, Open Approach |
|  |  |  | procedure | UMLS:ICD10PCS:0TY0 | Urinary System / Transplantation / Kidney, Right |
|  |  |  | procedure | UMLS:ICD10PCS:0TY1 | Urinary System / Transplantation / Kidney, Left |
|  | date constraint | | The terms in this group occurred at any time | | |
|  | event relationship | | Any instance of noStatins occurred before or up to 999 days after the first instance of Kidney transplant | | |
|  | **Group 4B noStatins** | | | | |
|  | cannot have |  | medication | NLM:ATC:C10AA | HMG CoA reductase inhibitors |
| Group 5 | | | | | |
|  | **Group 5A Kidney transplant** | | | | |
|  | must have | any of | diagnosis | UMLS:ICD10CM:Z94.0 | Kidney transplant status |
|  |  |  | procedure | UMLS:ICD10PCS:0TY00Z0 | Transplantation of Right Kidney, Allogeneic, Open Approach |
|  |  |  | procedure | UMLS:ICD10PCS:0TY10Z0 | Transplantation of Left Kidney, Allogeneic, Open Approach |
|  |  |  | procedure | UMLS:ICD10PCS:0TY10Z1 | Transplantation of Left Kidney, Syngeneic, Open Approach |
|  |  |  | procedure | UMLS:ICD10PCS:0TY10Z2 | Transplantation of Left Kidney, Zooplastic, Open Approach |
|  |  |  | procedure | UMLS:ICD10PCS:0TY00Z1 | Transplantation of Right Kidney, Syngeneic, Open Approach |
|  |  |  | procedure | UMLS:ICD10PCS:0TY00Z2 | Transplantation of Right Kidney, Zooplastic, Open Approach |
|  |  |  | procedure | UMLS:ICD10PCS:0TY0 | Urinary System / Transplantation / Kidney, Right |
|  |  |  | procedure | UMLS:ICD10PCS:0TY1 | Urinary System / Transplantation / Kidney, Left |
|  | date constraint | | The terms in this group occurred at any time | | |
|  | event relationship | | Any instance of Visit occurred at least 1181 days after the first instance of Kidney transplant | | |
|  | **Group 5B Visit** | | | | |
|  | must have |  | visit | TNX:Visit | Visit |

#### Negative exposure control analyses

##### **Table S21. Code definitions and selection criteria for amitriptyline users.**

| Ungrouped terms | | | | | |
| --- | --- | --- | --- | --- | --- |
|  | must have |  | demographics | Age | Age (at least 18 years (most recent occurrence)) |
|  | cannot have |  | diagnosis | UMLS:ICD10CM:E08-E13 | Diabetes mellitus |
| Group 1 | | | | | |
|  | **Kidney transplant** | | | | |
|  | must have | any of | diagnosis | UMLS:ICD10CM:Z94.0 | Kidney transplant status |
|  |  |  | procedure | UMLS:ICD10PCS:0TY00Z0 | Transplantation of Right Kidney, Allogeneic, Open Approach |
|  |  |  | procedure | UMLS:ICD10PCS:0TY10Z0 | Transplantation of Left Kidney, Allogeneic, Open Approach |
|  |  |  | procedure | UMLS:ICD10PCS:0TY10Z1 | Transplantation of Left Kidney, Syngeneic, Open Approach |
|  |  |  | procedure | UMLS:ICD10PCS:0TY10Z2 | Transplantation of Left Kidney, Zooplastic, Open Approach |
|  |  |  | procedure | UMLS:ICD10PCS:0TY00Z1 | Transplantation of Right Kidney, Syngeneic, Open Approach |
|  |  |  | procedure | UMLS:ICD10PCS:0TY00Z2 | Transplantation of Right Kidney, Zooplastic, Open Approach |
|  |  |  | procedure | UMLS:ICD10PCS:0TY0 | Urinary System / Transplantation / Kidney, Right |
|  |  |  | procedure | UMLS:ICD10PCS:0TY1 | Urinary System / Transplantation / Kidney, Left |
|  | date constraint | | The terms in this group occurred between Jan 1, 2015 and Jan 1, 2022 | | |
| Group 2 | | | | | |
|  | **Group 2A Kidney transplant** | | | | |
|  | must have | any of | diagnosis | UMLS:ICD10CM:Z94.0 | Kidney transplant status |
|  |  |  | procedure | UMLS:ICD10PCS:0TY00Z0 | Transplantation of Right Kidney, Allogeneic, Open Approach |
|  |  |  | procedure | UMLS:ICD10PCS:0TY10Z0 | Transplantation of Left Kidney, Allogeneic, Open Approach |
|  |  |  | procedure | UMLS:ICD10PCS:0TY10Z1 | Transplantation of Left Kidney, Syngeneic, Open Approach |
|  |  |  | procedure | UMLS:ICD10PCS:0TY10Z2 | Transplantation of Left Kidney, Zooplastic, Open Approach |
|  |  |  | procedure | UMLS:ICD10PCS:0TY00Z1 | Transplantation of Right Kidney, Syngeneic, Open Approach |
|  |  |  | procedure | UMLS:ICD10PCS:0TY00Z2 | Transplantation of Right Kidney, Zooplastic, Open Approach |
|  |  |  | procedure | UMLS:ICD10PCS:0TY0 | Urinary System / Transplantation / Kidney, Right |
|  |  |  | procedure | UMLS:ICD10PCS:0TY1 | Urinary System / Transplantation / Kidney, Left |
|  | date constraint | | The terms in this group occurred at any time | | |
|  | event relationship | | Any instance of noDialysis, Death occurred within 1 day and 999 days after the first instance of Kidney transplant | | |
|  | **Group 2B noDialysis, Death** | | | | |
|  | cannot have |  | procedure | UMLS:CPT:1012740 | Dialysis Services and Procedures |
|  |  | or | procedure | UMLS:CPT:90945 | Dialysis procedure other than hemodialysis (eg, peritoneal dialysis, hemofiltration, or other continuous renal replacement therapies), with single evaluation by a physician or other qualified health care professional |
|  |  | or | procedure | UMLS:CPT:90947 | Dialysis procedure other than hemodialysis (eg, peritoneal dialysis, hemofiltration, or other continuous renal replacement therapies) requiring repeated evaluations by a physician or other qualified health care professional, with or without substantial revision of dialysis prescription |
|  |  | or | procedure | UMLS:CPT:1012752 | Hemodialysis Procedures |
|  |  | or | procedure | UMLS:SNOMED:302497006 | Hemodialysis |
|  |  | or | procedure | UMLS:CPT:90937 | Hemodialysis procedure requiring repeated evaluation(s) with or without substantial revision of dialysis prescription |
|  |  | or | procedure | UMLS:CPT:90935 | Hemodialysis procedure with single evaluation by a physician or other qualified health care professional |
|  |  | or | demographics | Deceased | Deceased |
|  |  | or | diagnosis | UMLS:ICD10CM:R99 | Ill-defined and unknown cause of mortality |
| Group 3 | | | | | |
|  | **Group 3A Kidney transplant** | | | | |
|  | must have | any of | diagnosis | UMLS:ICD10CM:Z94.0 | Kidney transplant status |
|  |  |  | procedure | UMLS:ICD10PCS:0TY00Z0 | Transplantation of Right Kidney, Allogeneic, Open Approach |
|  |  |  | procedure | UMLS:ICD10PCS:0TY10Z0 | Transplantation of Left Kidney, Allogeneic, Open Approach |
|  |  |  | procedure | UMLS:ICD10PCS:0TY10Z1 | Transplantation of Left Kidney, Syngeneic, Open Approach |
|  |  |  | procedure | UMLS:ICD10PCS:0TY10Z2 | Transplantation of Left Kidney, Zooplastic, Open Approach |
|  |  |  | procedure | UMLS:ICD10PCS:0TY00Z1 | Transplantation of Right Kidney, Syngeneic, Open Approach |
|  |  |  | procedure | UMLS:ICD10PCS:0TY00Z2 | Transplantation of Right Kidney, Zooplastic, Open Approach |
|  |  |  | procedure | UMLS:ICD10PCS:0TY0 | Urinary System / Transplantation / Kidney, Right |
|  |  |  | procedure | UMLS:ICD10PCS:0TY1 | Urinary System / Transplantation / Kidney, Left |
|  | date constraint | | The terms in this group occurred at any time | | |
|  | event relationship | | Any instance of Amitriptyline occurred within 1000 days and 1180 days after the first instance of Kidney transplant | | |
|  | **Group 3B Amitriptyline** | | | | |
|  | must have |  | medication | NLM:RXNORM:704 | amitriptyline |
| Group 4 | | | | | |
|  | **Group 4A Kidney transplant** | | | | |
|  | must have | any of | diagnosis | UMLS:ICD10CM:Z94.0 | Kidney transplant status |
|  |  |  | procedure | UMLS:ICD10PCS:0TY00Z0 | Transplantation of Right Kidney, Allogeneic, Open Approach |
|  |  |  | procedure | UMLS:ICD10PCS:0TY10Z0 | Transplantation of Left Kidney, Allogeneic, Open Approach |
|  |  |  | procedure | UMLS:ICD10PCS:0TY10Z1 | Transplantation of Left Kidney, Syngeneic, Open Approach |
|  |  |  | procedure | UMLS:ICD10PCS:0TY10Z2 | Transplantation of Left Kidney, Zooplastic, Open Approach |
|  |  |  | procedure | UMLS:ICD10PCS:0TY00Z1 | Transplantation of Right Kidney, Syngeneic, Open Approach |
|  |  |  | procedure | UMLS:ICD10PCS:0TY00Z2 | Transplantation of Right Kidney, Zooplastic, Open Approach |
|  |  |  | procedure | UMLS:ICD10PCS:0TY0 | Urinary System / Transplantation / Kidney, Right |
|  |  |  | procedure | UMLS:ICD10PCS:0TY1 | Urinary System / Transplantation / Kidney, Left |
|  | date constraint | | The terms in this group occurred at any time | | |
|  | event relationship | | Any instance of Visit occurred at least 1181 days after the first instance of Kidney transplant | | |
|  | **Group 4B Visit** | | | | |
|  | must have |  | visit | TNX:Visit | Visit |

##### **Table S22. Code definitions and selection criteria for amitriptyline non-users.**

| Ungrouped terms | | | | | |
| --- | --- | --- | --- | --- | --- |
|  | must have |  | demographics | Age | Age (at least 18 years (most recent occurrence)) |
|  | cannot have |  | diagnosis | UMLS:ICD10CM:E08-E13 | Diabetes mellitus |
| Group 1 | | | | | |
|  | **Kidney transplant** | | | | |
|  | must have | any of | diagnosis | UMLS:ICD10CM:Z94.0 | Kidney transplant status |
|  |  |  | procedure | UMLS:ICD10PCS:0TY00Z0 | Transplantation of Right Kidney, Allogeneic, Open Approach |
|  |  |  | procedure | UMLS:ICD10PCS:0TY10Z0 | Transplantation of Left Kidney, Allogeneic, Open Approach |
|  |  |  | procedure | UMLS:ICD10PCS:0TY10Z1 | Transplantation of Left Kidney, Syngeneic, Open Approach |
|  |  |  | procedure | UMLS:ICD10PCS:0TY10Z2 | Transplantation of Left Kidney, Zooplastic, Open Approach |
|  |  |  | procedure | UMLS:ICD10PCS:0TY00Z1 | Transplantation of Right Kidney, Syngeneic, Open Approach |
|  |  |  | procedure | UMLS:ICD10PCS:0TY00Z2 | Transplantation of Right Kidney, Zooplastic, Open Approach |
|  |  |  | procedure | UMLS:ICD10PCS:0TY0 | Urinary System / Transplantation / Kidney, Right |
|  |  |  | procedure | UMLS:ICD10PCS:0TY1 | Urinary System / Transplantation / Kidney, Left |
|  | date constraint | | The terms in this group occurred between Jan 1, 2015 and Jan 1, 2022 | | |
| Group 2 | | | | | |
|  | **Group 2A Kidney transplant** | | | | |
|  | must have | any of | diagnosis | UMLS:ICD10CM:Z94.0 | Kidney transplant status |
|  |  |  | procedure | UMLS:ICD10PCS:0TY00Z0 | Transplantation of Right Kidney, Allogeneic, Open Approach |
|  |  |  | procedure | UMLS:ICD10PCS:0TY10Z0 | Transplantation of Left Kidney, Allogeneic, Open Approach |
|  |  |  | procedure | UMLS:ICD10PCS:0TY10Z1 | Transplantation of Left Kidney, Syngeneic, Open Approach |
|  |  |  | procedure | UMLS:ICD10PCS:0TY10Z2 | Transplantation of Left Kidney, Zooplastic, Open Approach |
|  |  |  | procedure | UMLS:ICD10PCS:0TY00Z1 | Transplantation of Right Kidney, Syngeneic, Open Approach |
|  |  |  | procedure | UMLS:ICD10PCS:0TY00Z2 | Transplantation of Right Kidney, Zooplastic, Open Approach |
|  |  |  | procedure | UMLS:ICD10PCS:0TY0 | Urinary System / Transplantation / Kidney, Right |
|  |  |  | procedure | UMLS:ICD10PCS:0TY1 | Urinary System / Transplantation / Kidney, Left |
|  | date constraint | | The terms in this group occurred at any time | | |
|  | event relationship | | Any instance of noDialysis, Death occurred within 1 day and 999 days after the first instance of Kidney transplant | | |
|  | **Group 2B noDialysis, Death** | | | | |
|  | cannot have |  | procedure | UMLS:CPT:1012740 | Dialysis Services and Procedures |
|  |  | or | procedure | UMLS:CPT:90945 | Dialysis procedure other than hemodialysis (eg, peritoneal dialysis, hemofiltration, or other continuous renal replacement therapies), with single evaluation by a physician or other qualified health care professional |
|  |  | or | procedure | UMLS:CPT:90947 | Dialysis procedure other than hemodialysis (eg, peritoneal dialysis, hemofiltration, or other continuous renal replacement therapies) requiring repeated evaluations by a physician or other qualified health care professional, with or without substantial revision of dialysis prescription |
|  |  | or | procedure | UMLS:CPT:1012752 | Hemodialysis Procedures |
|  |  | or | procedure | UMLS:SNOMED:302497006 | Hemodialysis |
|  |  | or | procedure | UMLS:CPT:90937 | Hemodialysis procedure requiring repeated evaluation(s) with or without substantial revision of dialysis prescription |
|  |  | or | procedure | UMLS:CPT:90935 | Hemodialysis procedure with single evaluation by a physician or other qualified health care professional |
|  |  | or | demographics | Deceased | Deceased |
|  |  | or | diagnosis | UMLS:ICD10CM:R99 | Ill-defined and unknown cause of mortality |
| Group 3 | | | | | |
|  | **Group 3A Kidney transplant** | | | | |
|  | must have | any of | diagnosis | UMLS:ICD10CM:Z94.0 | Kidney transplant status |
|  |  |  | procedure | UMLS:ICD10PCS:0TY00Z0 | Transplantation of Right Kidney, Allogeneic, Open Approach |
|  |  |  | procedure | UMLS:ICD10PCS:0TY10Z0 | Transplantation of Left Kidney, Allogeneic, Open Approach |
|  |  |  | procedure | UMLS:ICD10PCS:0TY10Z1 | Transplantation of Left Kidney, Syngeneic, Open Approach |
|  |  |  | procedure | UMLS:ICD10PCS:0TY10Z2 | Transplantation of Left Kidney, Zooplastic, Open Approach |
|  |  |  | procedure | UMLS:ICD10PCS:0TY00Z1 | Transplantation of Right Kidney, Syngeneic, Open Approach |
|  |  |  | procedure | UMLS:ICD10PCS:0TY00Z2 | Transplantation of Right Kidney, Zooplastic, Open Approach |
|  |  |  | procedure | UMLS:ICD10PCS:0TY0 | Urinary System / Transplantation / Kidney, Right |
|  |  |  | procedure | UMLS:ICD10PCS:0TY1 | Urinary System / Transplantation / Kidney, Left |
|  | date constraint | | The terms in this group occurred at any time | | |
|  | event relationship | | Any instance of noAmitriptyline occurred within 1000 days and 1180 days after the first instance of Kidney transplant | | |
|  | **Group 3B noAmitriptyline** | | | | |
|  | cannot have |  | medication | NLM:RXNORM:704 | amitriptyline |
| Group 4 | | | | | |
|  | **Group 4A Kidney transplant** | | | | |
|  | must have | any of | diagnosis | UMLS:ICD10CM:Z94.0 | Kidney transplant status |
|  |  |  | procedure | UMLS:ICD10PCS:0TY00Z0 | Transplantation of Right Kidney, Allogeneic, Open Approach |
|  |  |  | procedure | UMLS:ICD10PCS:0TY10Z0 | Transplantation of Left Kidney, Allogeneic, Open Approach |
|  |  |  | procedure | UMLS:ICD10PCS:0TY10Z1 | Transplantation of Left Kidney, Syngeneic, Open Approach |
|  |  |  | procedure | UMLS:ICD10PCS:0TY10Z2 | Transplantation of Left Kidney, Zooplastic, Open Approach |
|  |  |  | procedure | UMLS:ICD10PCS:0TY00Z1 | Transplantation of Right Kidney, Syngeneic, Open Approach |
|  |  |  | procedure | UMLS:ICD10PCS:0TY00Z2 | Transplantation of Right Kidney, Zooplastic, Open Approach |
|  |  |  | procedure | UMLS:ICD10PCS:0TY0 | Urinary System / Transplantation / Kidney, Right |
|  |  |  | procedure | UMLS:ICD10PCS:0TY1 | Urinary System / Transplantation / Kidney, Left |
|  | date constraint | | The terms in this group occurred at any time | | |
|  | event relationship | | Any instance of Visit occurred at least 1181 days after the first instance of Kidney transplant | | |
|  | **Group 4B Visit** | | | | |
|  | must have |  | visit | TNX:Visit | Visit |

##### **Table S23. Code definitions and selection criteria for mouthwash users.**

| Ungrouped terms | | | | | |
| --- | --- | --- | --- | --- | --- |
|  | must have |  | demographics | Age | Age (at least 18 years (most recent occurrence)) |
|  | cannot have |  | diagnosis | UMLS:ICD10CM:E08-E13 | Diabetes mellitus |
| Group 1 | | | | | |
|  | **Kidney transplant** | | | | |
|  | must have | any of | diagnosis | UMLS:ICD10CM:Z94.0 | Kidney transplant status |
|  |  |  | procedure | UMLS:ICD10PCS:0TY00Z0 | Transplantation of Right Kidney, Allogeneic, Open Approach |
|  |  |  | procedure | UMLS:ICD10PCS:0TY10Z0 | Transplantation of Left Kidney, Allogeneic, Open Approach |
|  |  |  | procedure | UMLS:ICD10PCS:0TY10Z1 | Transplantation of Left Kidney, Syngeneic, Open Approach |
|  |  |  | procedure | UMLS:ICD10PCS:0TY10Z2 | Transplantation of Left Kidney, Zooplastic, Open Approach |
|  |  |  | procedure | UMLS:ICD10PCS:0TY00Z1 | Transplantation of Right Kidney, Syngeneic, Open Approach |
|  |  |  | procedure | UMLS:ICD10PCS:0TY00Z2 | Transplantation of Right Kidney, Zooplastic, Open Approach |
|  |  |  | procedure | UMLS:ICD10PCS:0TY0 | Urinary System / Transplantation / Kidney, Right |
|  |  |  | procedure | UMLS:ICD10PCS:0TY1 | Urinary System / Transplantation / Kidney, Left |
|  | date constraint | | The terms in this group occurred between Jan 1, 2015 and Jan 1, 2022 | | |
| Group 2 | | | | | |
|  | **Group 2A Kidney transplant** | | | | |
|  | must have | any of | diagnosis | UMLS:ICD10CM:Z94.0 | Kidney transplant status |
|  |  |  | procedure | UMLS:ICD10PCS:0TY00Z0 | Transplantation of Right Kidney, Allogeneic, Open Approach |
|  |  |  | procedure | UMLS:ICD10PCS:0TY10Z0 | Transplantation of Left Kidney, Allogeneic, Open Approach |
|  |  |  | procedure | UMLS:ICD10PCS:0TY10Z1 | Transplantation of Left Kidney, Syngeneic, Open Approach |
|  |  |  | procedure | UMLS:ICD10PCS:0TY10Z2 | Transplantation of Left Kidney, Zooplastic, Open Approach |
|  |  |  | procedure | UMLS:ICD10PCS:0TY00Z1 | Transplantation of Right Kidney, Syngeneic, Open Approach |
|  |  |  | procedure | UMLS:ICD10PCS:0TY00Z2 | Transplantation of Right Kidney, Zooplastic, Open Approach |
|  |  |  | procedure | UMLS:ICD10PCS:0TY0 | Urinary System / Transplantation / Kidney, Right |
|  |  |  | procedure | UMLS:ICD10PCS:0TY1 | Urinary System / Transplantation / Kidney, Left |
|  | date constraint | | The terms in this group occurred at any time | | |
|  | event relationship | | Any instance of noDialysis, Death occurred within 1 day and 999 days after the first instance of Kidney transplant | | |
|  | **Group 2B noDialysis, Death** | | | | |
|  | cannot have |  | procedure | UMLS:CPT:1012740 | Dialysis Services and Procedures |
|  |  | or | procedure | UMLS:CPT:90945 | Dialysis procedure other than hemodialysis (eg, peritoneal dialysis, hemofiltration, or other continuous renal replacement therapies), with single evaluation by a physician or other qualified health care professional |
|  |  | or | procedure | UMLS:CPT:90947 | Dialysis procedure other than hemodialysis (eg, peritoneal dialysis, hemofiltration, or other continuous renal replacement therapies) requiring repeated evaluations by a physician or other qualified health care professional, with or without substantial revision of dialysis prescription |
|  |  | or | procedure | UMLS:CPT:1012752 | Hemodialysis Procedures |
|  |  | or | procedure | UMLS:SNOMED:302497006 | Hemodialysis |
|  |  | or | procedure | UMLS:CPT:90937 | Hemodialysis procedure requiring repeated evaluation(s) with or without substantial revision of dialysis prescription |
|  |  | or | procedure | UMLS:CPT:90935 | Hemodialysis procedure with single evaluation by a physician or other qualified health care professional |
|  |  | or | demographics | Deceased | Deceased |
|  |  | or | diagnosis | UMLS:ICD10CM:R99 | Ill-defined and unknown cause of mortality |
| Group 3 | | | | | |
|  | **Group 3A Kidney transplant** | | | | |
|  | must have | any of | diagnosis | UMLS:ICD10CM:Z94.0 | Kidney transplant status |
|  |  |  | procedure | UMLS:ICD10PCS:0TY00Z0 | Transplantation of Right Kidney, Allogeneic, Open Approach |
|  |  |  | procedure | UMLS:ICD10PCS:0TY10Z0 | Transplantation of Left Kidney, Allogeneic, Open Approach |
|  |  |  | procedure | UMLS:ICD10PCS:0TY10Z1 | Transplantation of Left Kidney, Syngeneic, Open Approach |
|  |  |  | procedure | UMLS:ICD10PCS:0TY10Z2 | Transplantation of Left Kidney, Zooplastic, Open Approach |
|  |  |  | procedure | UMLS:ICD10PCS:0TY00Z1 | Transplantation of Right Kidney, Syngeneic, Open Approach |
|  |  |  | procedure | UMLS:ICD10PCS:0TY00Z2 | Transplantation of Right Kidney, Zooplastic, Open Approach |
|  |  |  | procedure | UMLS:ICD10PCS:0TY0 | Urinary System / Transplantation / Kidney, Right |
|  |  |  | procedure | UMLS:ICD10PCS:0TY1 | Urinary System / Transplantation / Kidney, Left |
|  | date constraint | | The terms in this group occurred at any time | | |
|  | event relationship | | Any instance of Mouthwash occurred within 1000 days and 1180 days after the first instance of Kidney transplant | | |
|  | **Group 3B Mouthwash** | | | | |
|  | must have |  | medication | NLM:VA:OR500 | MOUTHWASHES |
| Group 4 | | | | | |
|  | **Group 4A Kidney transplant** | | | | |
|  | must have | any of | diagnosis | UMLS:ICD10CM:Z94.0 | Kidney transplant status |
|  |  |  | procedure | UMLS:ICD10PCS:0TY00Z0 | Transplantation of Right Kidney, Allogeneic, Open Approach |
|  |  |  | procedure | UMLS:ICD10PCS:0TY10Z0 | Transplantation of Left Kidney, Allogeneic, Open Approach |
|  |  |  | procedure | UMLS:ICD10PCS:0TY10Z1 | Transplantation of Left Kidney, Syngeneic, Open Approach |
|  |  |  | procedure | UMLS:ICD10PCS:0TY10Z2 | Transplantation of Left Kidney, Zooplastic, Open Approach |
|  |  |  | procedure | UMLS:ICD10PCS:0TY00Z1 | Transplantation of Right Kidney, Syngeneic, Open Approach |
|  |  |  | procedure | UMLS:ICD10PCS:0TY00Z2 | Transplantation of Right Kidney, Zooplastic, Open Approach |
|  |  |  | procedure | UMLS:ICD10PCS:0TY0 | Urinary System / Transplantation / Kidney, Right |
|  |  |  | procedure | UMLS:ICD10PCS:0TY1 | Urinary System / Transplantation / Kidney, Left |
|  | date constraint | | The terms in this group occurred at any time | | |
|  | event relationship | | Any instance of Visit occurred at least 1181 days after the first instance of Kidney transplant | | |
|  | **Group 4B Visit** | | | | |
|  | must have |  | visit | TNX:Visit | Visit |

##### **Table S24. Code definitions and selection criteria for mouthwash non-users.**

| Ungrouped terms | | | | | |
| --- | --- | --- | --- | --- | --- |
|  | must have |  | demographics | Age | Age (at least 18 years (most recent occurrence)) |
|  | cannot have |  | diagnosis | UMLS:ICD10CM:E08-E13 | Diabetes mellitus |
| Group 1 | | | | | |
|  | **Kidney transplant** | | | | |
|  | must have | any of | diagnosis | UMLS:ICD10CM:Z94.0 | Kidney transplant status |
|  |  |  | procedure | UMLS:ICD10PCS:0TY00Z0 | Transplantation of Right Kidney, Allogeneic, Open Approach |
|  |  |  | procedure | UMLS:ICD10PCS:0TY10Z0 | Transplantation of Left Kidney, Allogeneic, Open Approach |
|  |  |  | procedure | UMLS:ICD10PCS:0TY10Z1 | Transplantation of Left Kidney, Syngeneic, Open Approach |
|  |  |  | procedure | UMLS:ICD10PCS:0TY10Z2 | Transplantation of Left Kidney, Zooplastic, Open Approach |
|  |  |  | procedure | UMLS:ICD10PCS:0TY00Z1 | Transplantation of Right Kidney, Syngeneic, Open Approach |
|  |  |  | procedure | UMLS:ICD10PCS:0TY00Z2 | Transplantation of Right Kidney, Zooplastic, Open Approach |
|  |  |  | procedure | UMLS:ICD10PCS:0TY0 | Urinary System / Transplantation / Kidney, Right |
|  |  |  | procedure | UMLS:ICD10PCS:0TY1 | Urinary System / Transplantation / Kidney, Left |
|  | date constraint | | The terms in this group occurred between Jan 1, 2015 and Jan 1, 2022 | | |
| Group 2 | | | | | |
|  | **Group 2A Kidney transplant** | | | | |
|  | must have | any of | diagnosis | UMLS:ICD10CM:Z94.0 | Kidney transplant status |
|  |  |  | procedure | UMLS:ICD10PCS:0TY00Z0 | Transplantation of Right Kidney, Allogeneic, Open Approach |
|  |  |  | procedure | UMLS:ICD10PCS:0TY10Z0 | Transplantation of Left Kidney, Allogeneic, Open Approach |
|  |  |  | procedure | UMLS:ICD10PCS:0TY10Z1 | Transplantation of Left Kidney, Syngeneic, Open Approach |
|  |  |  | procedure | UMLS:ICD10PCS:0TY10Z2 | Transplantation of Left Kidney, Zooplastic, Open Approach |
|  |  |  | procedure | UMLS:ICD10PCS:0TY00Z1 | Transplantation of Right Kidney, Syngeneic, Open Approach |
|  |  |  | procedure | UMLS:ICD10PCS:0TY00Z2 | Transplantation of Right Kidney, Zooplastic, Open Approach |
|  |  |  | procedure | UMLS:ICD10PCS:0TY0 | Urinary System / Transplantation / Kidney, Right |
|  |  |  | procedure | UMLS:ICD10PCS:0TY1 | Urinary System / Transplantation / Kidney, Left |
|  | date constraint | | The terms in this group occurred at any time | | |
|  | event relationship | | Any instance of noDialysis, Death occurred within 1 day and 999 days after the first instance of Kidney transplant | | |
|  | **Group 2B noDialysis, Death** | | | | |
|  | cannot have |  | procedure | UMLS:CPT:1012740 | Dialysis Services and Procedures |
|  |  | or | procedure | UMLS:CPT:90945 | Dialysis procedure other than hemodialysis (eg, peritoneal dialysis, hemofiltration, or other continuous renal replacement therapies), with single evaluation by a physician or other qualified health care professional |
|  |  | or | procedure | UMLS:CPT:90947 | Dialysis procedure other than hemodialysis (eg, peritoneal dialysis, hemofiltration, or other continuous renal replacement therapies) requiring repeated evaluations by a physician or other qualified health care professional, with or without substantial revision of dialysis prescription |
|  |  | or | procedure | UMLS:CPT:1012752 | Hemodialysis Procedures |
|  |  | or | procedure | UMLS:SNOMED:302497006 | Hemodialysis |
|  |  | or | procedure | UMLS:CPT:90937 | Hemodialysis procedure requiring repeated evaluation(s) with or without substantial revision of dialysis prescription |
|  |  | or | procedure | UMLS:CPT:90935 | Hemodialysis procedure with single evaluation by a physician or other qualified health care professional |
|  |  | or | demographics | Deceased | Deceased |
|  |  | or | diagnosis | UMLS:ICD10CM:R99 | Ill-defined and unknown cause of mortality |
| Group 3 | | | | | |
|  | **Group 3A Kidney transplant** | | | | |
|  | must have | any of | diagnosis | UMLS:ICD10CM:Z94.0 | Kidney transplant status |
|  |  |  | procedure | UMLS:ICD10PCS:0TY00Z0 | Transplantation of Right Kidney, Allogeneic, Open Approach |
|  |  |  | procedure | UMLS:ICD10PCS:0TY10Z0 | Transplantation of Left Kidney, Allogeneic, Open Approach |
|  |  |  | procedure | UMLS:ICD10PCS:0TY10Z1 | Transplantation of Left Kidney, Syngeneic, Open Approach |
|  |  |  | procedure | UMLS:ICD10PCS:0TY10Z2 | Transplantation of Left Kidney, Zooplastic, Open Approach |
|  |  |  | procedure | UMLS:ICD10PCS:0TY00Z1 | Transplantation of Right Kidney, Syngeneic, Open Approach |
|  |  |  | procedure | UMLS:ICD10PCS:0TY00Z2 | Transplantation of Right Kidney, Zooplastic, Open Approach |
|  |  |  | procedure | UMLS:ICD10PCS:0TY0 | Urinary System / Transplantation / Kidney, Right |
|  |  |  | procedure | UMLS:ICD10PCS:0TY1 | Urinary System / Transplantation / Kidney, Left |
|  | date constraint | | The terms in this group occurred at any time | | |
|  | event relationship | | Any instance of noMouthwash occurred within 1000 days and 3006 days after the first instance of Kidney transplant | | |
|  | **Group 3B noMouthwash** | | | | |
|  | cannot have |  | medication | NLM:VA:OR500 | MOUTHWASHES |
| Group 4 | | | | | |
|  | **Group 4A Kidney transplant** | | | | |
|  | must have | any of | diagnosis | UMLS:ICD10CM:Z94.0 | Kidney transplant status |
|  |  |  | procedure | UMLS:ICD10PCS:0TY00Z0 | Transplantation of Right Kidney, Allogeneic, Open Approach |
|  |  |  | procedure | UMLS:ICD10PCS:0TY10Z0 | Transplantation of Left Kidney, Allogeneic, Open Approach |
|  |  |  | procedure | UMLS:ICD10PCS:0TY10Z1 | Transplantation of Left Kidney, Syngeneic, Open Approach |
|  |  |  | procedure | UMLS:ICD10PCS:0TY10Z2 | Transplantation of Left Kidney, Zooplastic, Open Approach |
|  |  |  | procedure | UMLS:ICD10PCS:0TY00Z1 | Transplantation of Right Kidney, Syngeneic, Open Approach |
|  |  |  | procedure | UMLS:ICD10PCS:0TY00Z2 | Transplantation of Right Kidney, Zooplastic, Open Approach |
|  |  |  | procedure | UMLS:ICD10PCS:0TY0 | Urinary System / Transplantation / Kidney, Right |
|  |  |  | procedure | UMLS:ICD10PCS:0TY1 | Urinary System / Transplantation / Kidney, Left |
|  | date constraint | | The terms in this group occurred at any time | | |
|  | event relationship | | Any instance of Visit occurred at least 1181 days after the first instance of Kidney transplant | | |
|  | **Group 4B Visit** | | | | |
|  | must have |  | visit | TNX:Visit | Visit |

### 9. **Subgroup analyses**

#### Demographics

##### **Table S25. Code definitions and selection criteria for SGLT2i users – female.**

| Ungrouped terms | | | | | |
| --- | --- | --- | --- | --- | --- |
|  | must have |  | demographics | Age | Age (at least 18 years (most recent occurrence)) |
|  |  | and any of | demographics | UMLS:HL7V3.0:Gender:F | Female |
|  |  |  | demographics | UMLS:HL7V3.0:Gender:UN | Unknown |
|  | cannot have |  | diagnosis | UMLS:ICD10CM:E08-E13 | Diabetes mellitus |
| Group 1 | | | | | |
|  | **Kidney transplant** | | | | |
|  | must have | any of | diagnosis | UMLS:ICD10CM:Z94.0 | Kidney transplant status |
|  |  |  | procedure | UMLS:ICD10PCS:0TY00Z0 | Transplantation of Right Kidney, Allogeneic, Open Approach |
|  |  |  | procedure | UMLS:ICD10PCS:0TY10Z0 | Transplantation of Left Kidney, Allogeneic, Open Approach |
|  |  |  | procedure | UMLS:ICD10PCS:0TY10Z1 | Transplantation of Left Kidney, Syngeneic, Open Approach |
|  |  |  | procedure | UMLS:ICD10PCS:0TY10Z2 | Transplantation of Left Kidney, Zooplastic, Open Approach |
|  |  |  | procedure | UMLS:ICD10PCS:0TY00Z1 | Transplantation of Right Kidney, Syngeneic, Open Approach |
|  |  |  | procedure | UMLS:ICD10PCS:0TY00Z2 | Transplantation of Right Kidney, Zooplastic, Open Approach |
|  |  |  | procedure | UMLS:ICD10PCS:0TY0 | Urinary System / Transplantation / Kidney, Right |
|  |  |  | procedure | UMLS:ICD10PCS:0TY1 | Urinary System / Transplantation / Kidney, Left |
|  | date constraint | | The terms in this group occurred between Jan 1, 2015 and Jan 1, 2022 | | |
| Group 2 | | | | | |
|  | **Group 2A Kidney transplant** | | | | |
|  | must have | any of | diagnosis | UMLS:ICD10CM:Z94.0 | Kidney transplant status |
|  |  |  | procedure | UMLS:ICD10PCS:0TY00Z0 | Transplantation of Right Kidney, Allogeneic, Open Approach |
|  |  |  | procedure | UMLS:ICD10PCS:0TY10Z0 | Transplantation of Left Kidney, Allogeneic, Open Approach |
|  |  |  | procedure | UMLS:ICD10PCS:0TY10Z1 | Transplantation of Left Kidney, Syngeneic, Open Approach |
|  |  |  | procedure | UMLS:ICD10PCS:0TY10Z2 | Transplantation of Left Kidney, Zooplastic, Open Approach |
|  |  |  | procedure | UMLS:ICD10PCS:0TY00Z1 | Transplantation of Right Kidney, Syngeneic, Open Approach |
|  |  |  | procedure | UMLS:ICD10PCS:0TY00Z2 | Transplantation of Right Kidney, Zooplastic, Open Approach |
|  |  |  | procedure | UMLS:ICD10PCS:0TY0 | Urinary System / Transplantation / Kidney, Right |
|  |  |  | procedure | UMLS:ICD10PCS:0TY1 | Urinary System / Transplantation / Kidney, Left |
|  | date constraint | | The terms in this group occurred at any time | | |
|  | event relationship | | Any instance of noDialysis, Death, SGLT2 occurred within 1 day and 999 days after the first instance of Kidney transplant | | |
|  | **Group 2B noDialysis, Death, SGLT2i** | | | | |
|  | cannot have |  | procedure | UMLS:CPT:1012740 | Dialysis Services and Procedures |
|  |  | or | procedure | UMLS:CPT:90945 | Dialysis procedure other than hemodialysis (eg, peritoneal dialysis, hemofiltration, or other continuous renal replacement therapies), with single evaluation by a physician or other qualified health care professional |
|  |  | or | procedure | UMLS:CPT:90947 | Dialysis procedure other than hemodialysis (eg, peritoneal dialysis, hemofiltration, or other continuous renal replacement therapies) requiring repeated evaluations by a physician or other qualified health care professional, with or without substantial revision of dialysis prescription |
|  |  | or | procedure | UMLS:CPT:1012752 | Hemodialysis Procedures |
|  |  | or | procedure | UMLS:SNOMED:302497006 | Hemodialysis |
|  |  | or | procedure | UMLS:CPT:90937 | Hemodialysis procedure requiring repeated evaluation(s) with or without substantial revision of dialysis prescription |
|  |  | or | procedure | UMLS:CPT:90935 | Hemodialysis procedure with single evaluation by a physician or other qualified health care professional |
|  |  | or | demographics | Deceased | Deceased |
|  |  | or | diagnosis | UMLS:ICD10CM:R99 | Ill-defined and unknown cause of mortality |
|  |  | or | medication | NLM:ATC:A10BK | Sodium-glucose co-transporter 2 (SGLT2) inhibitors |
| Group 3 | | | | | |
|  | **Group 3A Kidney transplant** | | | | |
|  | must have | any of | diagnosis | UMLS:ICD10CM:Z94.0 | Kidney transplant status |
|  |  |  | procedure | UMLS:ICD10PCS:0TY00Z0 | Transplantation of Right Kidney, Allogeneic, Open Approach |
|  |  |  | procedure | UMLS:ICD10PCS:0TY10Z0 | Transplantation of Left Kidney, Allogeneic, Open Approach |
|  |  |  | procedure | UMLS:ICD10PCS:0TY10Z1 | Transplantation of Left Kidney, Syngeneic, Open Approach |
|  |  |  | procedure | UMLS:ICD10PCS:0TY10Z2 | Transplantation of Left Kidney, Zooplastic, Open Approach |
|  |  |  | procedure | UMLS:ICD10PCS:0TY00Z1 | Transplantation of Right Kidney, Syngeneic, Open Approach |
|  |  |  | procedure | UMLS:ICD10PCS:0TY00Z2 | Transplantation of Right Kidney, Zooplastic, Open Approach |
|  |  |  | procedure | UMLS:ICD10PCS:0TY0 | Urinary System / Transplantation / Kidney, Right |
|  |  |  | procedure | UMLS:ICD10PCS:0TY1 | Urinary System / Transplantation / Kidney, Left |
|  | date constraint | | The terms in this group occurred at any time | | |
|  | event relationship | | Any instance of SGLT2i occurred at least 1000 days after the first instance of Kidney transplant | | |
|  | **Group 3B SGLT2i** | | | | |
|  | must have |  | medication | NLM:ATC:A10BK | Sodium-glucose co-transporter 2 (SGLT2) inhibitors |
| Group 4 | | | | | |
|  | **Group 4A Kidney transplant** | | | | |
|  | must have | any of | diagnosis | UMLS:ICD10CM:Z94.0 | Kidney transplant status |
|  |  |  | procedure | UMLS:ICD10PCS:0TY00Z0 | Transplantation of Right Kidney, Allogeneic, Open Approach |
|  |  |  | procedure | UMLS:ICD10PCS:0TY10Z0 | Transplantation of Left Kidney, Allogeneic, Open Approach |
|  |  |  | procedure | UMLS:ICD10PCS:0TY10Z1 | Transplantation of Left Kidney, Syngeneic, Open Approach |
|  |  |  | procedure | UMLS:ICD10PCS:0TY10Z2 | Transplantation of Left Kidney, Zooplastic, Open Approach |
|  |  |  | procedure | UMLS:ICD10PCS:0TY00Z1 | Transplantation of Right Kidney, Syngeneic, Open Approach |
|  |  |  | procedure | UMLS:ICD10PCS:0TY00Z2 | Transplantation of Right Kidney, Zooplastic, Open Approach |
|  |  |  | procedure | UMLS:ICD10PCS:0TY0 | Urinary System / Transplantation / Kidney, Right |
|  |  |  | procedure | UMLS:ICD10PCS:0TY1 | Urinary System / Transplantation / Kidney, Left |
|  | date constraint | | The terms in this group occurred at any time | | |
|  | event relationship | | Any instance of noSGLT2i occurred at least 1 day before the first instance of Kidney transplant | | |
|  | **Group 4B noSGLT2i** | | | | |
|  | cannot have |  | medication | NLM:ATC:A10BK | Sodium-glucose co-transporter 2 (SGLT2) inhibitors |
| Group 5 | | | | | |
|  | **Group 5A Kidney transplant** | | | | |
|  | must have | any of | diagnosis | UMLS:ICD10CM:Z94.0 | Kidney transplant status |
|  |  |  | procedure | UMLS:ICD10PCS:0TY00Z0 | Transplantation of Right Kidney, Allogeneic, Open Approach |
|  |  |  | procedure | UMLS:ICD10PCS:0TY10Z0 | Transplantation of Left Kidney, Allogeneic, Open Approach |
|  |  |  | procedure | UMLS:ICD10PCS:0TY10Z1 | Transplantation of Left Kidney, Syngeneic, Open Approach |
|  |  |  | procedure | UMLS:ICD10PCS:0TY10Z2 | Transplantation of Left Kidney, Zooplastic, Open Approach |
|  |  |  | procedure | UMLS:ICD10PCS:0TY00Z1 | Transplantation of Right Kidney, Syngeneic, Open Approach |
|  |  |  | procedure | UMLS:ICD10PCS:0TY00Z2 | Transplantation of Right Kidney, Zooplastic, Open Approach |
|  |  |  | procedure | UMLS:ICD10PCS:0TY0 | Urinary System / Transplantation / Kidney, Right |
|  |  |  | procedure | UMLS:ICD10PCS:0TY1 | Urinary System / Transplantation / Kidney, Left |
|  | date constraint | | The terms in this group occurred at any time | | |
|  | event relationship | | Any instance of Visit occurred at least 1181 days after the first instance of Kidney transplant | | |
|  | **Group 5B Visit** | | | | |
|  | must have |  | visit | TNX:Visit | Visit |

##### **Table S26. Code definitions and selection criteria for SGLT2i non-users – female.**

| Ungrouped terms | | | | | |
| --- | --- | --- | --- | --- | --- |
|  | must have |  | demographics | Age | Age (at least 18 years (most recent occurrence)) |
|  |  | and any of | demographics | UMLS:HL7V3.0:Gender:F | Female |
|  |  |  | demographics | UMLS:HL7V3.0:Gender:UN | Unknown |
|  | cannot have |  | diagnosis | UMLS:ICD10CM:E08-E13 | Diabetes mellitus |
| Group 1 | | | | | |
|  | **Kidney transplant** | | | | |
|  | must have | any of | diagnosis | UMLS:ICD10CM:Z94.0 | Kidney transplant status |
|  |  |  | procedure | UMLS:ICD10PCS:0TY00Z0 | Transplantation of Right Kidney, Allogeneic, Open Approach |
|  |  |  | procedure | UMLS:ICD10PCS:0TY10Z0 | Transplantation of Left Kidney, Allogeneic, Open Approach |
|  |  |  | procedure | UMLS:ICD10PCS:0TY10Z1 | Transplantation of Left Kidney, Syngeneic, Open Approach |
|  |  |  | procedure | UMLS:ICD10PCS:0TY10Z2 | Transplantation of Left Kidney, Zooplastic, Open Approach |
|  |  |  | procedure | UMLS:ICD10PCS:0TY00Z1 | Transplantation of Right Kidney, Syngeneic, Open Approach |
|  |  |  | procedure | UMLS:ICD10PCS:0TY00Z2 | Transplantation of Right Kidney, Zooplastic, Open Approach |
|  |  |  | procedure | UMLS:ICD10PCS:0TY0 | Urinary System / Transplantation / Kidney, Right |
|  |  |  | procedure | UMLS:ICD10PCS:0TY1 | Urinary System / Transplantation / Kidney, Left |
|  | date constraint | | The terms in this group occurred between Jan 1, 2015 and Jan 1, 2022 | | |
| Group 2 | | | | | |
|  | **Group 2A Kidney transplant** | | | | |
|  | must have | any of | diagnosis | UMLS:ICD10CM:Z94.0 | Kidney transplant status |
|  |  |  | procedure | UMLS:ICD10PCS:0TY00Z0 | Transplantation of Right Kidney, Allogeneic, Open Approach |
|  |  |  | procedure | UMLS:ICD10PCS:0TY10Z0 | Transplantation of Left Kidney, Allogeneic, Open Approach |
|  |  |  | procedure | UMLS:ICD10PCS:0TY10Z1 | Transplantation of Left Kidney, Syngeneic, Open Approach |
|  |  |  | procedure | UMLS:ICD10PCS:0TY10Z2 | Transplantation of Left Kidney, Zooplastic, Open Approach |
|  |  |  | procedure | UMLS:ICD10PCS:0TY00Z1 | Transplantation of Right Kidney, Syngeneic, Open Approach |
|  |  |  | procedure | UMLS:ICD10PCS:0TY00Z2 | Transplantation of Right Kidney, Zooplastic, Open Approach |
|  |  |  | procedure | UMLS:ICD10PCS:0TY0 | Urinary System / Transplantation / Kidney, Right |
|  |  |  | procedure | UMLS:ICD10PCS:0TY1 | Urinary System / Transplantation / Kidney, Left |
|  | date constraint | | The terms in this group occurred at any time | | |
|  | event relationship | | Any instance of noDialysis, Death, SGLT2i occurred within 1 day and 999 days after the first instance of Kidney transplant | | |
|  | **Group 2B noDialysis, Death, SGLT2i** | | | | |
|  | cannot have |  | procedure | UMLS:CPT:1012740 | Dialysis Services and Procedures |
|  |  | or | procedure | UMLS:CPT:90945 | Dialysis procedure other than hemodialysis (eg, peritoneal dialysis, hemofiltration, or other continuous renal replacement therapies), with single evaluation by a physician or other qualified health care professional |
|  |  | or | procedure | UMLS:CPT:90947 | Dialysis procedure other than hemodialysis (eg, peritoneal dialysis, hemofiltration, or other continuous renal replacement therapies) requiring repeated evaluations by a physician or other qualified health care professional, with or without substantial revision of dialysis prescription |
|  |  | or | procedure | UMLS:CPT:1012752 | Hemodialysis Procedures |
|  |  | or | procedure | UMLS:SNOMED:302497006 | Hemodialysis |
|  |  | or | procedure | UMLS:CPT:90937 | Hemodialysis procedure requiring repeated evaluation(s) with or without substantial revision of dialysis prescription |
|  |  | or | procedure | UMLS:CPT:90935 | Hemodialysis procedure with single evaluation by a physician or other qualified health care professional |
|  |  | or | demographics | Deceased | Deceased |
|  |  | or | diagnosis | UMLS:ICD10CM:R99 | Ill-defined and unknown cause of mortality |
|  |  | or | medication | NLM:ATC:A10BK | Sodium-glucose co-transporter 2 (SGLT2) inhibitors |
| Group 3 | | | | | |
|  | **Group 3A Kidney transplant** | | | | |
|  | must have | any of | diagnosis | UMLS:ICD10CM:Z94.0 | Kidney transplant status |
|  |  |  | procedure | UMLS:ICD10PCS:0TY00Z0 | Transplantation of Right Kidney, Allogeneic, Open Approach |
|  |  |  | procedure | UMLS:ICD10PCS:0TY10Z0 | Transplantation of Left Kidney, Allogeneic, Open Approach |
|  |  |  | procedure | UMLS:ICD10PCS:0TY10Z1 | Transplantation of Left Kidney, Syngeneic, Open Approach |
|  |  |  | procedure | UMLS:ICD10PCS:0TY10Z2 | Transplantation of Left Kidney, Zooplastic, Open Approach |
|  |  |  | procedure | UMLS:ICD10PCS:0TY00Z1 | Transplantation of Right Kidney, Syngeneic, Open Approach |
|  |  |  | procedure | UMLS:ICD10PCS:0TY00Z2 | Transplantation of Right Kidney, Zooplastic, Open Approach |
|  |  |  | procedure | UMLS:ICD10PCS:0TY0 | Urinary System / Transplantation / Kidney, Right |
|  |  |  | procedure | UMLS:ICD10PCS:0TY1 | Urinary System / Transplantation / Kidney, Left |
|  | date constraint | | The terms in this group occurred at any time | | |
|  | event relationship | | Any instance of SGLT2i occurred within 1000 days and 3006 days after the first instance of Kidney transplant | | |
|  | **Group 3B noSGLT2i** | | | | |
|  | cannot have |  | medication | NLM:ATC:A10BK | Sodium-glucose co-transporter 2 (SGLT2) inhibitors |
| Group 4 | | | | | |
|  | **Group 4A Kidney transplant** | | | | |
|  | must have | any of | diagnosis | UMLS:ICD10CM:Z94.0 | Kidney transplant status |
|  |  |  | procedure | UMLS:ICD10PCS:0TY00Z0 | Transplantation of Right Kidney, Allogeneic, Open Approach |
|  |  |  | procedure | UMLS:ICD10PCS:0TY10Z0 | Transplantation of Left Kidney, Allogeneic, Open Approach |
|  |  |  | procedure | UMLS:ICD10PCS:0TY10Z1 | Transplantation of Left Kidney, Syngeneic, Open Approach |
|  |  |  | procedure | UMLS:ICD10PCS:0TY10Z2 | Transplantation of Left Kidney, Zooplastic, Open Approach |
|  |  |  | procedure | UMLS:ICD10PCS:0TY00Z1 | Transplantation of Right Kidney, Syngeneic, Open Approach |
|  |  |  | procedure | UMLS:ICD10PCS:0TY00Z2 | Transplantation of Right Kidney, Zooplastic, Open Approach |
|  |  |  | procedure | UMLS:ICD10PCS:0TY0 | Urinary System / Transplantation / Kidney, Right |
|  |  |  | procedure | UMLS:ICD10PCS:0TY1 | Urinary System / Transplantation / Kidney, Left |
|  | date constraint | | The terms in this group occurred at any time | | |
|  | event relationship | | Any instance of noSGLT2i occurred at least 1 day before the first instance of Kidney transplant | | |
|  | **Group 4B noSGLT2i** | | | | |
|  | cannot have |  | medication | NLM:ATC:A10BK | Sodium-glucose co-transporter 2 (SGLT2) inhibitors |
| Group 5 | | | | | |
|  | **Group 5A Kidney transplant** | | | | |
|  | must have | any of | diagnosis | UMLS:ICD10CM:Z94.0 | Kidney transplant status |
|  |  |  | procedure | UMLS:ICD10PCS:0TY00Z0 | Transplantation of Right Kidney, Allogeneic, Open Approach |
|  |  |  | procedure | UMLS:ICD10PCS:0TY10Z0 | Transplantation of Left Kidney, Allogeneic, Open Approach |
|  |  |  | procedure | UMLS:ICD10PCS:0TY10Z1 | Transplantation of Left Kidney, Syngeneic, Open Approach |
|  |  |  | procedure | UMLS:ICD10PCS:0TY10Z2 | Transplantation of Left Kidney, Zooplastic, Open Approach |
|  |  |  | procedure | UMLS:ICD10PCS:0TY00Z1 | Transplantation of Right Kidney, Syngeneic, Open Approach |
|  |  |  | procedure | UMLS:ICD10PCS:0TY00Z2 | Transplantation of Right Kidney, Zooplastic, Open Approach |
|  |  |  | procedure | UMLS:ICD10PCS:0TY0 | Urinary System / Transplantation / Kidney, Right |
|  |  |  | procedure | UMLS:ICD10PCS:0TY1 | Urinary System / Transplantation / Kidney, Left |
|  | date constraint | | The terms in this group occurred at any time | | |
|  | event relationship | | Any instance of Visit occurred at least 1181 days after the first instance of Kidney transplant | | |
|  | **Group 5B Visit** | | | | |
|  | must have |  | visit | TNX:Visit | Visit |

##### **Table S27. Code definitions and selection criteria for SGLT2i users – male.**

| Ungrouped terms | | | | | |
| --- | --- | --- | --- | --- | --- |
|  | must have |  | demographics | Age | Age (at least 18 years (most recent occurrence)) |
|  |  | and any of | demographics | UMLS:HL7V3.0:Gender:M | Male |
|  |  |  | demographics | UMLS:HL7V3.0:Gender:UN | Unknown |
|  | cannot have |  | diagnosis | UMLS:ICD10CM:E08-E13 | Diabetes mellitus |
| Group 1 | | | | | |
|  | **Kidney transplant** | | | | |
|  | must have | any of | diagnosis | UMLS:ICD10CM:Z94.0 | Kidney transplant status |
|  |  |  | procedure | UMLS:ICD10PCS:0TY00Z0 | Transplantation of Right Kidney, Allogeneic, Open Approach |
|  |  |  | procedure | UMLS:ICD10PCS:0TY10Z0 | Transplantation of Left Kidney, Allogeneic, Open Approach |
|  |  |  | procedure | UMLS:ICD10PCS:0TY10Z1 | Transplantation of Left Kidney, Syngeneic, Open Approach |
|  |  |  | procedure | UMLS:ICD10PCS:0TY10Z2 | Transplantation of Left Kidney, Zooplastic, Open Approach |
|  |  |  | procedure | UMLS:ICD10PCS:0TY00Z1 | Transplantation of Right Kidney, Syngeneic, Open Approach |
|  |  |  | procedure | UMLS:ICD10PCS:0TY00Z2 | Transplantation of Right Kidney, Zooplastic, Open Approach |
|  |  |  | procedure | UMLS:ICD10PCS:0TY0 | Urinary System / Transplantation / Kidney, Right |
|  |  |  | procedure | UMLS:ICD10PCS:0TY1 | Urinary System / Transplantation / Kidney, Left |
|  | date constraint | | The terms in this group occurred between Jan 1, 2015 and Jan 1, 2022 | | |
| Group 2 | | | | | |
|  | **Group 2A Kidney transplant** | | | | |
|  | must have | any of | diagnosis | UMLS:ICD10CM:Z94.0 | Kidney transplant status |
|  |  |  | procedure | UMLS:ICD10PCS:0TY00Z0 | Transplantation of Right Kidney, Allogeneic, Open Approach |
|  |  |  | procedure | UMLS:ICD10PCS:0TY10Z0 | Transplantation of Left Kidney, Allogeneic, Open Approach |
|  |  |  | procedure | UMLS:ICD10PCS:0TY10Z1 | Transplantation of Left Kidney, Syngeneic, Open Approach |
|  |  |  | procedure | UMLS:ICD10PCS:0TY10Z2 | Transplantation of Left Kidney, Zooplastic, Open Approach |
|  |  |  | procedure | UMLS:ICD10PCS:0TY00Z1 | Transplantation of Right Kidney, Syngeneic, Open Approach |
|  |  |  | procedure | UMLS:ICD10PCS:0TY00Z2 | Transplantation of Right Kidney, Zooplastic, Open Approach |
|  |  |  | procedure | UMLS:ICD10PCS:0TY0 | Urinary System / Transplantation / Kidney, Right |
|  |  |  | procedure | UMLS:ICD10PCS:0TY1 | Urinary System / Transplantation / Kidney, Left |
|  | date constraint | | The terms in this group occurred at any time | | |
|  | event relationship | | Any instance of noDialysis, Death, SGLT2i occurred within 1 day and 999 days after the first instance of Kidney transplant | | |
|  | **Group 2B noDialysis, Death, SGLT2i** | | | | |
|  | cannot have |  | procedure | UMLS:CPT:1012740 | Dialysis Services and Procedures |
|  |  | or | procedure | UMLS:CPT:90945 | Dialysis procedure other than hemodialysis (eg, peritoneal dialysis, hemofiltration, or other continuous renal replacement therapies), with single evaluation by a physician or other qualified health care professional |
|  |  | or | procedure | UMLS:CPT:90947 | Dialysis procedure other than hemodialysis (eg, peritoneal dialysis, hemofiltration, or other continuous renal replacement therapies) requiring repeated evaluations by a physician or other qualified health care professional, with or without substantial revision of dialysis prescription |
|  |  | or | procedure | UMLS:CPT:1012752 | Hemodialysis Procedures |
|  |  | or | procedure | UMLS:SNOMED:302497006 | Hemodialysis |
|  |  | or | procedure | UMLS:CPT:90937 | Hemodialysis procedure requiring repeated evaluation(s) with or without substantial revision of dialysis prescription |
|  |  | or | procedure | UMLS:CPT:90935 | Hemodialysis procedure with single evaluation by a physician or other qualified health care professional |
|  |  | or | demographics | Deceased | Deceased |
|  |  | or | diagnosis | UMLS:ICD10CM:R99 | Ill-defined and unknown cause of mortality |
|  |  | or | medication | NLM:ATC:A10BK | Sodium-glucose co-transporter 2 (SGLT2) inhibitors |
| Group 3 | | | | | |
|  | **Group 3A Kidney transplant** | | | | |
|  | must have | any of | diagnosis | UMLS:ICD10CM:Z94.0 | Kidney transplant status |
|  |  |  | procedure | UMLS:ICD10PCS:0TY00Z0 | Transplantation of Right Kidney, Allogeneic, Open Approach |
|  |  |  | procedure | UMLS:ICD10PCS:0TY10Z0 | Transplantation of Left Kidney, Allogeneic, Open Approach |
|  |  |  | procedure | UMLS:ICD10PCS:0TY10Z1 | Transplantation of Left Kidney, Syngeneic, Open Approach |
|  |  |  | procedure | UMLS:ICD10PCS:0TY10Z2 | Transplantation of Left Kidney, Zooplastic, Open Approach |
|  |  |  | procedure | UMLS:ICD10PCS:0TY00Z1 | Transplantation of Right Kidney, Syngeneic, Open Approach |
|  |  |  | procedure | UMLS:ICD10PCS:0TY00Z2 | Transplantation of Right Kidney, Zooplastic, Open Approach |
|  |  |  | procedure | UMLS:ICD10PCS:0TY0 | Urinary System / Transplantation / Kidney, Right |
|  |  |  | procedure | UMLS:ICD10PCS:0TY1 | Urinary System / Transplantation / Kidney, Left |
|  | date constraint | | The terms in this group occurred at any time | | |
|  | event relationship | | Any instance of SGLT2i occurred at least 1000 days after the first instance of Kidney transplant | | |
|  | **Group 3B SGLT2i** | | | | |
|  | must have |  | medication | NLM:ATC:A10BK | Sodium-glucose co-transporter 2 (SGLT2) inhibitors |
| Group 4 | | | | | |
|  | **Group 4A Kidney transplant** | | | | |
|  | must have | any of | diagnosis | UMLS:ICD10CM:Z94.0 | Kidney transplant status |
|  |  |  | procedure | UMLS:ICD10PCS:0TY00Z0 | Transplantation of Right Kidney, Allogeneic, Open Approach |
|  |  |  | procedure | UMLS:ICD10PCS:0TY10Z0 | Transplantation of Left Kidney, Allogeneic, Open Approach |
|  |  |  | procedure | UMLS:ICD10PCS:0TY10Z1 | Transplantation of Left Kidney, Syngeneic, Open Approach |
|  |  |  | procedure | UMLS:ICD10PCS:0TY10Z2 | Transplantation of Left Kidney, Zooplastic, Open Approach |
|  |  |  | procedure | UMLS:ICD10PCS:0TY00Z1 | Transplantation of Right Kidney, Syngeneic, Open Approach |
|  |  |  | procedure | UMLS:ICD10PCS:0TY00Z2 | Transplantation of Right Kidney, Zooplastic, Open Approach |
|  |  |  | procedure | UMLS:ICD10PCS:0TY0 | Urinary System / Transplantation / Kidney, Right |
|  |  |  | procedure | UMLS:ICD10PCS:0TY1 | Urinary System / Transplantation / Kidney, Left |
|  | date constraint | | The terms in this group occurred at any time | | |
|  | event relationship | | Any instance of noSGLT2i occurred at least 1 day before the first instance of Kidney transplant | | |
|  | **Group 4B noSGLT2i** | | | | |
|  | cannot have |  | medication | NLM:ATC:A10BK | Sodium-glucose co-transporter 2 (SGLT2) inhibitors |
| Group 5 | | | | | |
|  | **Group 5A Kidney transplant** | | | | |
|  | must have | any of | diagnosis | UMLS:ICD10CM:Z94.0 | Kidney transplant status |
|  |  |  | procedure | UMLS:ICD10PCS:0TY00Z0 | Transplantation of Right Kidney, Allogeneic, Open Approach |
|  |  |  | procedure | UMLS:ICD10PCS:0TY10Z0 | Transplantation of Left Kidney, Allogeneic, Open Approach |
|  |  |  | procedure | UMLS:ICD10PCS:0TY10Z1 | Transplantation of Left Kidney, Syngeneic, Open Approach |
|  |  |  | procedure | UMLS:ICD10PCS:0TY10Z2 | Transplantation of Left Kidney, Zooplastic, Open Approach |
|  |  |  | procedure | UMLS:ICD10PCS:0TY00Z1 | Transplantation of Right Kidney, Syngeneic, Open Approach |
|  |  |  | procedure | UMLS:ICD10PCS:0TY00Z2 | Transplantation of Right Kidney, Zooplastic, Open Approach |
|  |  |  | procedure | UMLS:ICD10PCS:0TY0 | Urinary System / Transplantation / Kidney, Right |
|  |  |  | procedure | UMLS:ICD10PCS:0TY1 | Urinary System / Transplantation / Kidney, Left |
|  | date constraint | | The terms in this group occurred at any time | | |
|  | event relationship | | Any instance of Visit occurred at least 1181 days after the first instance of Kidney transplant | | |
|  | **Group 5B Visit** | | | | |
|  | must have |  | visit | TNX:Visit | Visit |

##### **Table S28. Code definitions and selection criteria for SGLT2i non-users – male.**

| Ungrouped terms | | | | | |
| --- | --- | --- | --- | --- | --- |
|  | must have |  | demographics | Age | Age (at least 18 years (most recent occurrence)) |
|  |  | and any of | demographics | UMLS:HL7V3.0:Gender:M | Male |
|  |  |  | demographics | UMLS:HL7V3.0:Gender:UN | Unknown |
|  | cannot have |  | diagnosis | UMLS:ICD10CM:E08-E13 | Diabetes mellitus |
| Group 1 | | | | | |
|  | **Kidney transplant** | | | | |
|  | must have | any of | diagnosis | UMLS:ICD10CM:Z94.0 | Kidney transplant status |
|  |  |  | procedure | UMLS:ICD10PCS:0TY00Z0 | Transplantation of Right Kidney, Allogeneic, Open Approach |
|  |  |  | procedure | UMLS:ICD10PCS:0TY10Z0 | Transplantation of Left Kidney, Allogeneic, Open Approach |
|  |  |  | procedure | UMLS:ICD10PCS:0TY10Z1 | Transplantation of Left Kidney, Syngeneic, Open Approach |
|  |  |  | procedure | UMLS:ICD10PCS:0TY10Z2 | Transplantation of Left Kidney, Zooplastic, Open Approach |
|  |  |  | procedure | UMLS:ICD10PCS:0TY00Z1 | Transplantation of Right Kidney, Syngeneic, Open Approach |
|  |  |  | procedure | UMLS:ICD10PCS:0TY00Z2 | Transplantation of Right Kidney, Zooplastic, Open Approach |
|  |  |  | procedure | UMLS:ICD10PCS:0TY0 | Urinary System / Transplantation / Kidney, Right |
|  |  |  | procedure | UMLS:ICD10PCS:0TY1 | Urinary System / Transplantation / Kidney, Left |
|  | date constraint | | The terms in this group occurred between Jan 1, 2015 and Jan 1, 2022 | | |
| Group 2 | | | | | |
|  | **Group 2A Kidney transplant** | | | | |
|  | must have | any of | diagnosis | UMLS:ICD10CM:Z94.0 | Kidney transplant status |
|  |  |  | procedure | UMLS:ICD10PCS:0TY00Z0 | Transplantation of Right Kidney, Allogeneic, Open Approach |
|  |  |  | procedure | UMLS:ICD10PCS:0TY10Z0 | Transplantation of Left Kidney, Allogeneic, Open Approach |
|  |  |  | procedure | UMLS:ICD10PCS:0TY10Z1 | Transplantation of Left Kidney, Syngeneic, Open Approach |
|  |  |  | procedure | UMLS:ICD10PCS:0TY10Z2 | Transplantation of Left Kidney, Zooplastic, Open Approach |
|  |  |  | procedure | UMLS:ICD10PCS:0TY00Z1 | Transplantation of Right Kidney, Syngeneic, Open Approach |
|  |  |  | procedure | UMLS:ICD10PCS:0TY00Z2 | Transplantation of Right Kidney, Zooplastic, Open Approach |
|  |  |  | procedure | UMLS:ICD10PCS:0TY0 | Urinary System / Transplantation / Kidney, Right |
|  |  |  | procedure | UMLS:ICD10PCS:0TY1 | Urinary System / Transplantation / Kidney, Left |
|  | date constraint | | The terms in this group occurred at any time | | |
|  | event relationship | | Any instance of noDialysis, Death, SGLT2i occurred within 1 day and 999 days after the first instance of Kidney transplant | | |
|  | **Group 2B noDialysis, Death, SGLT2i** | | | | |
|  | cannot have |  | procedure | UMLS:CPT:1012740 | Dialysis Services and Procedures |
|  |  | or | procedure | UMLS:CPT:90945 | Dialysis procedure other than hemodialysis (eg, peritoneal dialysis, hemofiltration, or other continuous renal replacement therapies), with single evaluation by a physician or other qualified health care professional |
|  |  | or | procedure | UMLS:CPT:90947 | Dialysis procedure other than hemodialysis (eg, peritoneal dialysis, hemofiltration, or other continuous renal replacement therapies) requiring repeated evaluations by a physician or other qualified health care professional, with or without substantial revision of dialysis prescription |
|  |  | or | procedure | UMLS:CPT:1012752 | Hemodialysis Procedures |
|  |  | or | procedure | UMLS:SNOMED:302497006 | Hemodialysis |
|  |  | or | procedure | UMLS:CPT:90937 | Hemodialysis procedure requiring repeated evaluation(s) with or without substantial revision of dialysis prescription |
|  |  | or | procedure | UMLS:CPT:90935 | Hemodialysis procedure with single evaluation by a physician or other qualified health care professional |
|  |  | or | demographics | Deceased | Deceased |
|  |  | or | diagnosis | UMLS:ICD10CM:R99 | Ill-defined and unknown cause of mortality |
|  |  | or | medication | NLM:ATC:A10BK | Sodium-glucose co-transporter 2 (SGLT2) inhibitors |
| Group 3 | | | | | |
|  | **Group 3A Kidney transplant** | | | | |
|  | must have | any of | diagnosis | UMLS:ICD10CM:Z94.0 | Kidney transplant status |
|  |  |  | procedure | UMLS:ICD10PCS:0TY00Z0 | Transplantation of Right Kidney, Allogeneic, Open Approach |
|  |  |  | procedure | UMLS:ICD10PCS:0TY10Z0 | Transplantation of Left Kidney, Allogeneic, Open Approach |
|  |  |  | procedure | UMLS:ICD10PCS:0TY10Z1 | Transplantation of Left Kidney, Syngeneic, Open Approach |
|  |  |  | procedure | UMLS:ICD10PCS:0TY10Z2 | Transplantation of Left Kidney, Zooplastic, Open Approach |
|  |  |  | procedure | UMLS:ICD10PCS:0TY00Z1 | Transplantation of Right Kidney, Syngeneic, Open Approach |
|  |  |  | procedure | UMLS:ICD10PCS:0TY00Z2 | Transplantation of Right Kidney, Zooplastic, Open Approach |
|  |  |  | procedure | UMLS:ICD10PCS:0TY0 | Urinary System / Transplantation / Kidney, Right |
|  |  |  | procedure | UMLS:ICD10PCS:0TY1 | Urinary System / Transplantation / Kidney, Left |
|  | date constraint | | The terms in this group occurred at any time | | |
|  | event relationship | | Any instance of SGLT2i occurred within 1000 days and 3006 days after the first instance of Kidney transplant | | |
|  | **Group 3B noSGLT2i** | | | | |
|  | cannot have |  | medication | NLM:ATC:A10BK | Sodium-glucose co-transporter 2 (SGLT2) inhibitors |
| Group 4 | | | | | |
|  | **Group 4A Kidney transplant** | | | | |
|  | must have | any of | diagnosis | UMLS:ICD10CM:Z94.0 | Kidney transplant status |
|  |  |  | procedure | UMLS:ICD10PCS:0TY00Z0 | Transplantation of Right Kidney, Allogeneic, Open Approach |
|  |  |  | procedure | UMLS:ICD10PCS:0TY10Z0 | Transplantation of Left Kidney, Allogeneic, Open Approach |
|  |  |  | procedure | UMLS:ICD10PCS:0TY10Z1 | Transplantation of Left Kidney, Syngeneic, Open Approach |
|  |  |  | procedure | UMLS:ICD10PCS:0TY10Z2 | Transplantation of Left Kidney, Zooplastic, Open Approach |
|  |  |  | procedure | UMLS:ICD10PCS:0TY00Z1 | Transplantation of Right Kidney, Syngeneic, Open Approach |
|  |  |  | procedure | UMLS:ICD10PCS:0TY00Z2 | Transplantation of Right Kidney, Zooplastic, Open Approach |
|  |  |  | procedure | UMLS:ICD10PCS:0TY0 | Urinary System / Transplantation / Kidney, Right |
|  |  |  | procedure | UMLS:ICD10PCS:0TY1 | Urinary System / Transplantation / Kidney, Left |
|  | date constraint | | The terms in this group occurred at any time | | |
|  | event relationship | | Any instance of noSGLT2i occurred at least 1 day before the first instance of Kidney transplant | | |
|  | **Group 4B noSGLT2i** | | | | |
|  | cannot have |  | medication | NLM:ATC:A10BK | Sodium-glucose co-transporter 2 (SGLT2) inhibitors |
| Group 5 | | | | | |
|  | **Group 5A Kidney transplant** | | | | |
|  | must have | any of | diagnosis | UMLS:ICD10CM:Z94.0 | Kidney transplant status |
|  |  |  | procedure | UMLS:ICD10PCS:0TY00Z0 | Transplantation of Right Kidney, Allogeneic, Open Approach |
|  |  |  | procedure | UMLS:ICD10PCS:0TY10Z0 | Transplantation of Left Kidney, Allogeneic, Open Approach |
|  |  |  | procedure | UMLS:ICD10PCS:0TY10Z1 | Transplantation of Left Kidney, Syngeneic, Open Approach |
|  |  |  | procedure | UMLS:ICD10PCS:0TY10Z2 | Transplantation of Left Kidney, Zooplastic, Open Approach |
|  |  |  | procedure | UMLS:ICD10PCS:0TY00Z1 | Transplantation of Right Kidney, Syngeneic, Open Approach |
|  |  |  | procedure | UMLS:ICD10PCS:0TY00Z2 | Transplantation of Right Kidney, Zooplastic, Open Approach |
|  |  |  | procedure | UMLS:ICD10PCS:0TY0 | Urinary System / Transplantation / Kidney, Right |
|  |  |  | procedure | UMLS:ICD10PCS:0TY1 | Urinary System / Transplantation / Kidney, Left |
|  | date constraint | | The terms in this group occurred at any time | | |
|  | event relationship | | Any instance of Visit occurred at least 1181 days after the first instance of Kidney transplant | | |
|  | **Group 5B Visit** | | | | |
|  | must have |  | visit | TNX:Visit | Visit |

##### **Table S29. Code definitions and selection criteria for SGLT2i users – 18-59 years.**

| Ungrouped terms | | | | | |
| --- | --- | --- | --- | --- | --- |
|  | must have |  | demographics | Age | Age (between 18 and 59 years (most recent occurrence)) |
|  | cannot have |  | diagnosis | UMLS:ICD10CM:E08-E13 | Diabetes mellitus |
| Group 1 | | | | | |
|  | **Kidney transplant** | | | | |
|  | must have | any of | diagnosis | UMLS:ICD10CM:Z94.0 | Kidney transplant status |
|  |  |  | procedure | UMLS:ICD10PCS:0TY00Z0 | Transplantation of Right Kidney, Allogeneic, Open Approach |
|  |  |  | procedure | UMLS:ICD10PCS:0TY10Z0 | Transplantation of Left Kidney, Allogeneic, Open Approach |
|  |  |  | procedure | UMLS:ICD10PCS:0TY10Z1 | Transplantation of Left Kidney, Syngeneic, Open Approach |
|  |  |  | procedure | UMLS:ICD10PCS:0TY10Z2 | Transplantation of Left Kidney, Zooplastic, Open Approach |
|  |  |  | procedure | UMLS:ICD10PCS:0TY00Z1 | Transplantation of Right Kidney, Syngeneic, Open Approach |
|  |  |  | procedure | UMLS:ICD10PCS:0TY00Z2 | Transplantation of Right Kidney, Zooplastic, Open Approach |
|  |  |  | procedure | UMLS:ICD10PCS:0TY0 | Urinary System / Transplantation / Kidney, Right |
|  |  |  | procedure | UMLS:ICD10PCS:0TY1 | Urinary System / Transplantation / Kidney, Left |
|  | date constraint | | The terms in this group occurred between Jan 1, 2015 and Jan 1, 2022 | | |
| Group 2 | | | | | |
|  | **Group 2A Kidney transplant** | | | | |
|  | must have | any of | diagnosis | UMLS:ICD10CM:Z94.0 | Kidney transplant status |
|  |  |  | procedure | UMLS:ICD10PCS:0TY00Z0 | Transplantation of Right Kidney, Allogeneic, Open Approach |
|  |  |  | procedure | UMLS:ICD10PCS:0TY10Z0 | Transplantation of Left Kidney, Allogeneic, Open Approach |
|  |  |  | procedure | UMLS:ICD10PCS:0TY10Z1 | Transplantation of Left Kidney, Syngeneic, Open Approach |
|  |  |  | procedure | UMLS:ICD10PCS:0TY10Z2 | Transplantation of Left Kidney, Zooplastic, Open Approach |
|  |  |  | procedure | UMLS:ICD10PCS:0TY00Z1 | Transplantation of Right Kidney, Syngeneic, Open Approach |
|  |  |  | procedure | UMLS:ICD10PCS:0TY00Z2 | Transplantation of Right Kidney, Zooplastic, Open Approach |
|  |  |  | procedure | UMLS:ICD10PCS:0TY0 | Urinary System / Transplantation / Kidney, Right |
|  |  |  | procedure | UMLS:ICD10PCS:0TY1 | Urinary System / Transplantation / Kidney, Left |
|  | date constraint | | The terms in this group occurred at any time | | |
|  | event relationship | | Any instance of noDialysis, Death, SGLT2i occurred within 1 day and 999 days after the first instance of Kidney transplant | | |
|  | **Group 2B noDialysis, Death, SGLT2i** | | | | |
|  | cannot have |  | procedure | UMLS:CPT:1012740 | Dialysis Services and Procedures |
|  |  | or | procedure | UMLS:CPT:90945 | Dialysis procedure other than hemodialysis (eg, peritoneal dialysis, hemofiltration, or other continuous renal replacement therapies), with single evaluation by a physician or other qualified health care professional |
|  |  | or | procedure | UMLS:CPT:90947 | Dialysis procedure other than hemodialysis (eg, peritoneal dialysis, hemofiltration, or other continuous renal replacement therapies) requiring repeated evaluations by a physician or other qualified health care professional, with or without substantial revision of dialysis prescription |
|  |  | or | procedure | UMLS:CPT:1012752 | Hemodialysis Procedures |
|  |  | or | procedure | UMLS:SNOMED:302497006 | Hemodialysis |
|  |  | or | procedure | UMLS:CPT:90937 | Hemodialysis procedure requiring repeated evaluation(s) with or without substantial revision of dialysis prescription |
|  |  | or | procedure | UMLS:CPT:90935 | Hemodialysis procedure with single evaluation by a physician or other qualified health care professional |
|  |  | or | demographics | Deceased | Deceased |
|  |  | or | diagnosis | UMLS:ICD10CM:R99 | Ill-defined and unknown cause of mortality |
|  |  | or | medication | NLM:ATC:A10BK | Sodium-glucose co-transporter 2 (SGLT2) inhibitors |
| Group 3 | | | | | |
|  | **Group 3A Kidney transplant** | | | | |
|  | must have | any of | diagnosis | UMLS:ICD10CM:Z94.0 | Kidney transplant status |
|  |  |  | procedure | UMLS:ICD10PCS:0TY00Z0 | Transplantation of Right Kidney, Allogeneic, Open Approach |
|  |  |  | procedure | UMLS:ICD10PCS:0TY10Z0 | Transplantation of Left Kidney, Allogeneic, Open Approach |
|  |  |  | procedure | UMLS:ICD10PCS:0TY10Z1 | Transplantation of Left Kidney, Syngeneic, Open Approach |
|  |  |  | procedure | UMLS:ICD10PCS:0TY10Z2 | Transplantation of Left Kidney, Zooplastic, Open Approach |
|  |  |  | procedure | UMLS:ICD10PCS:0TY00Z1 | Transplantation of Right Kidney, Syngeneic, Open Approach |
|  |  |  | procedure | UMLS:ICD10PCS:0TY00Z2 | Transplantation of Right Kidney, Zooplastic, Open Approach |
|  |  |  | procedure | UMLS:ICD10PCS:0TY0 | Urinary System / Transplantation / Kidney, Right |
|  |  |  | procedure | UMLS:ICD10PCS:0TY1 | Urinary System / Transplantation / Kidney, Left |
|  | date constraint | | The terms in this group occurred at any time | | |
|  | event relationship | | Any instance of SGLT2i occurred at least 1000 days after the first instance of Kidney transplant | | |
|  | **Group 3B SGLT2i** | | | | |
|  | must have |  | medication | NLM:ATC:A10BK | Sodium-glucose co-transporter 2 (SGLT2) inhibitors |
| Group 4 | | | | | |
|  | **Group 4A Kidney transplant** | | | | |
|  | must have | any of | diagnosis | UMLS:ICD10CM:Z94.0 | Kidney transplant status |
|  |  |  | procedure | UMLS:ICD10PCS:0TY00Z0 | Transplantation of Right Kidney, Allogeneic, Open Approach |
|  |  |  | procedure | UMLS:ICD10PCS:0TY10Z0 | Transplantation of Left Kidney, Allogeneic, Open Approach |
|  |  |  | procedure | UMLS:ICD10PCS:0TY10Z1 | Transplantation of Left Kidney, Syngeneic, Open Approach |
|  |  |  | procedure | UMLS:ICD10PCS:0TY10Z2 | Transplantation of Left Kidney, Zooplastic, Open Approach |
|  |  |  | procedure | UMLS:ICD10PCS:0TY00Z1 | Transplantation of Right Kidney, Syngeneic, Open Approach |
|  |  |  | procedure | UMLS:ICD10PCS:0TY00Z2 | Transplantation of Right Kidney, Zooplastic, Open Approach |
|  |  |  | procedure | UMLS:ICD10PCS:0TY0 | Urinary System / Transplantation / Kidney, Right |
|  |  |  | procedure | UMLS:ICD10PCS:0TY1 | Urinary System / Transplantation / Kidney, Left |
|  | date constraint | | The terms in this group occurred at any time | | |
|  | event relationship | | Any instance of noSGLT2i occurred at least 1 day before the first instance of Kidney transplant | | |
|  | **Group 4B noSGLT2i** | | | | |
|  | cannot have |  | medication | NLM:ATC:A10BK | Sodium-glucose co-transporter 2 (SGLT2) inhibitors |
| Group 5 | | | | | |
|  | **Group 5A Kidney transplant** | | | | |
|  | must have | any of | diagnosis | UMLS:ICD10CM:Z94.0 | Kidney transplant status |
|  |  |  | procedure | UMLS:ICD10PCS:0TY00Z0 | Transplantation of Right Kidney, Allogeneic, Open Approach |
|  |  |  | procedure | UMLS:ICD10PCS:0TY10Z0 | Transplantation of Left Kidney, Allogeneic, Open Approach |
|  |  |  | procedure | UMLS:ICD10PCS:0TY10Z1 | Transplantation of Left Kidney, Syngeneic, Open Approach |
|  |  |  | procedure | UMLS:ICD10PCS:0TY10Z2 | Transplantation of Left Kidney, Zooplastic, Open Approach |
|  |  |  | procedure | UMLS:ICD10PCS:0TY00Z1 | Transplantation of Right Kidney, Syngeneic, Open Approach |
|  |  |  | procedure | UMLS:ICD10PCS:0TY00Z2 | Transplantation of Right Kidney, Zooplastic, Open Approach |
|  |  |  | procedure | UMLS:ICD10PCS:0TY0 | Urinary System / Transplantation / Kidney, Right |
|  |  |  | procedure | UMLS:ICD10PCS:0TY1 | Urinary System / Transplantation / Kidney, Left |
|  | date constraint | | The terms in this group occurred at any time | | |
|  | event relationship | | Any instance of Visit occurred at least 1181 days after the first instance of Kidney transplant | | |
|  | **Group 5B Visit** | | | | |
|  | must have |  | visit | TNX:Visit | Visit |

##### **Table S30. Code definitions and selection criteria for SGLT2i non-users – 18-59 years.**

| Ungrouped terms | | | | | |
| --- | --- | --- | --- | --- | --- |
|  | must have |  | demographics | Age | Age (between 18 and 59 years (most recent occurrence)) |
|  | cannot have |  | diagnosis | UMLS:ICD10CM:E08-E13 | Diabetes mellitus |
| Group 1 | | | | | |
|  | **Kidney transplant** | | | | |
|  | must have | any of | diagnosis | UMLS:ICD10CM:Z94.0 | Kidney transplant status |
|  |  |  | procedure | UMLS:ICD10PCS:0TY00Z0 | Transplantation of Right Kidney, Allogeneic, Open Approach |
|  |  |  | procedure | UMLS:ICD10PCS:0TY10Z0 | Transplantation of Left Kidney, Allogeneic, Open Approach |
|  |  |  | procedure | UMLS:ICD10PCS:0TY10Z1 | Transplantation of Left Kidney, Syngeneic, Open Approach |
|  |  |  | procedure | UMLS:ICD10PCS:0TY10Z2 | Transplantation of Left Kidney, Zooplastic, Open Approach |
|  |  |  | procedure | UMLS:ICD10PCS:0TY00Z1 | Transplantation of Right Kidney, Syngeneic, Open Approach |
|  |  |  | procedure | UMLS:ICD10PCS:0TY00Z2 | Transplantation of Right Kidney, Zooplastic, Open Approach |
|  |  |  | procedure | UMLS:ICD10PCS:0TY0 | Urinary System / Transplantation / Kidney, Right |
|  |  |  | procedure | UMLS:ICD10PCS:0TY1 | Urinary System / Transplantation / Kidney, Left |
|  | date constraint | | The terms in this group occurred between Jan 1, 2015 and Jan 1, 2022 | | |
| Group 2 | | | | | |
|  | **Group 2A Kidney transplant** | | | | |
|  | must have | any of | diagnosis | UMLS:ICD10CM:Z94.0 | Kidney transplant status |
|  |  |  | procedure | UMLS:ICD10PCS:0TY00Z0 | Transplantation of Right Kidney, Allogeneic, Open Approach |
|  |  |  | procedure | UMLS:ICD10PCS:0TY10Z0 | Transplantation of Left Kidney, Allogeneic, Open Approach |
|  |  |  | procedure | UMLS:ICD10PCS:0TY10Z1 | Transplantation of Left Kidney, Syngeneic, Open Approach |
|  |  |  | procedure | UMLS:ICD10PCS:0TY10Z2 | Transplantation of Left Kidney, Zooplastic, Open Approach |
|  |  |  | procedure | UMLS:ICD10PCS:0TY00Z1 | Transplantation of Right Kidney, Syngeneic, Open Approach |
|  |  |  | procedure | UMLS:ICD10PCS:0TY00Z2 | Transplantation of Right Kidney, Zooplastic, Open Approach |
|  |  |  | procedure | UMLS:ICD10PCS:0TY0 | Urinary System / Transplantation / Kidney, Right |
|  |  |  | procedure | UMLS:ICD10PCS:0TY1 | Urinary System / Transplantation / Kidney, Left |
|  | date constraint | | The terms in this group occurred at any time | | |
|  | event relationship | | Any instance of noDialysis, Death, SGLT2i occurred within 1 day and 999 days after the first instance of Kidney transplant | | |
|  | **Group 2B noDialysis, Death, SGLT2i** | | | | |
|  | cannot have |  | procedure | UMLS:CPT:1012740 | Dialysis Services and Procedures |
|  |  | or | procedure | UMLS:CPT:90945 | Dialysis procedure other than hemodialysis (eg, peritoneal dialysis, hemofiltration, or other continuous renal replacement therapies), with single evaluation by a physician or other qualified health care professional |
|  |  | or | procedure | UMLS:CPT:90947 | Dialysis procedure other than hemodialysis (eg, peritoneal dialysis, hemofiltration, or other continuous renal replacement therapies) requiring repeated evaluations by a physician or other qualified health care professional, with or without substantial revision of dialysis prescription |
|  |  | or | procedure | UMLS:CPT:1012752 | Hemodialysis Procedures |
|  |  | or | procedure | UMLS:SNOMED:302497006 | Hemodialysis |
|  |  | or | procedure | UMLS:CPT:90937 | Hemodialysis procedure requiring repeated evaluation(s) with or without substantial revision of dialysis prescription |
|  |  | or | procedure | UMLS:CPT:90935 | Hemodialysis procedure with single evaluation by a physician or other qualified health care professional |
|  |  | or | demographics | Deceased | Deceased |
|  |  | or | diagnosis | UMLS:ICD10CM:R99 | Ill-defined and unknown cause of mortality |
|  |  | or | medication | NLM:ATC:A10BK | Sodium-glucose co-transporter 2 (SGLT2) inhibitors |
| Group 3 | | | | | |
|  | **Group 3A Kidney transplant** | | | | |
|  | must have | any of | diagnosis | UMLS:ICD10CM:Z94.0 | Kidney transplant status |
|  |  |  | procedure | UMLS:ICD10PCS:0TY00Z0 | Transplantation of Right Kidney, Allogeneic, Open Approach |
|  |  |  | procedure | UMLS:ICD10PCS:0TY10Z0 | Transplantation of Left Kidney, Allogeneic, Open Approach |
|  |  |  | procedure | UMLS:ICD10PCS:0TY10Z1 | Transplantation of Left Kidney, Syngeneic, Open Approach |
|  |  |  | procedure | UMLS:ICD10PCS:0TY10Z2 | Transplantation of Left Kidney, Zooplastic, Open Approach |
|  |  |  | procedure | UMLS:ICD10PCS:0TY00Z1 | Transplantation of Right Kidney, Syngeneic, Open Approach |
|  |  |  | procedure | UMLS:ICD10PCS:0TY00Z2 | Transplantation of Right Kidney, Zooplastic, Open Approach |
|  |  |  | procedure | UMLS:ICD10PCS:0TY0 | Urinary System / Transplantation / Kidney, Right |
|  |  |  | procedure | UMLS:ICD10PCS:0TY1 | Urinary System / Transplantation / Kidney, Left |
|  | date constraint | | The terms in this group occurred at any time | | |
|  | event relationship | | Any instance of SGLT2i occurred within 1000 days and 3006 days after the first instance of Kidney transplant | | |
|  | **Group 3B noSGLT2i** | | | | |
|  | cannot have |  | medication | NLM:ATC:A10BK | Sodium-glucose co-transporter 2 (SGLT2) inhibitors |
| Group 4 | | | | | |
|  | **Group 4A Kidney transplant** | | | | |
|  | must have | any of | diagnosis | UMLS:ICD10CM:Z94.0 | Kidney transplant status |
|  |  |  | procedure | UMLS:ICD10PCS:0TY00Z0 | Transplantation of Right Kidney, Allogeneic, Open Approach |
|  |  |  | procedure | UMLS:ICD10PCS:0TY10Z0 | Transplantation of Left Kidney, Allogeneic, Open Approach |
|  |  |  | procedure | UMLS:ICD10PCS:0TY10Z1 | Transplantation of Left Kidney, Syngeneic, Open Approach |
|  |  |  | procedure | UMLS:ICD10PCS:0TY10Z2 | Transplantation of Left Kidney, Zooplastic, Open Approach |
|  |  |  | procedure | UMLS:ICD10PCS:0TY00Z1 | Transplantation of Right Kidney, Syngeneic, Open Approach |
|  |  |  | procedure | UMLS:ICD10PCS:0TY00Z2 | Transplantation of Right Kidney, Zooplastic, Open Approach |
|  |  |  | procedure | UMLS:ICD10PCS:0TY0 | Urinary System / Transplantation / Kidney, Right |
|  |  |  | procedure | UMLS:ICD10PCS:0TY1 | Urinary System / Transplantation / Kidney, Left |
|  | date constraint | | The terms in this group occurred at any time | | |
|  | event relationship | | Any instance of noSGLT2i occurred at least 1 day before the first instance of Kidney transplant | | |
|  | **Group 4B noSGLT2i** | | | | |
|  | cannot have |  | medication | NLM:ATC:A10BK | Sodium-glucose co-transporter 2 (SGLT2) inhibitors |
| Group 5 | | | | | |
|  | **Group 5A Kidney transplant** | | | | |
|  | must have | any of | diagnosis | UMLS:ICD10CM:Z94.0 | Kidney transplant status |
|  |  |  | procedure | UMLS:ICD10PCS:0TY00Z0 | Transplantation of Right Kidney, Allogeneic, Open Approach |
|  |  |  | procedure | UMLS:ICD10PCS:0TY10Z0 | Transplantation of Left Kidney, Allogeneic, Open Approach |
|  |  |  | procedure | UMLS:ICD10PCS:0TY10Z1 | Transplantation of Left Kidney, Syngeneic, Open Approach |
|  |  |  | procedure | UMLS:ICD10PCS:0TY10Z2 | Transplantation of Left Kidney, Zooplastic, Open Approach |
|  |  |  | procedure | UMLS:ICD10PCS:0TY00Z1 | Transplantation of Right Kidney, Syngeneic, Open Approach |
|  |  |  | procedure | UMLS:ICD10PCS:0TY00Z2 | Transplantation of Right Kidney, Zooplastic, Open Approach |
|  |  |  | procedure | UMLS:ICD10PCS:0TY0 | Urinary System / Transplantation / Kidney, Right |
|  |  |  | procedure | UMLS:ICD10PCS:0TY1 | Urinary System / Transplantation / Kidney, Left |
|  | date constraint | | The terms in this group occurred at any time | | |
|  | event relationship | | Any instance of Visit occurred at least 1181 days after the first instance of Kidney transplant | | |
|  | **Group 5B Visit** | | | | |
|  | must have |  | visit | TNX:Visit | Visit |

##### **Table S31. Code definitions and selection criteria for SGLT2i users – ≥ 60 years.**

| Ungrouped terms | | | | | |
| --- | --- | --- | --- | --- | --- |
|  | must have |  | demographics | Age | Age (at least 60 years (most recent occurrence)) |
|  | cannot have |  | diagnosis | UMLS:ICD10CM:E08-E13 | Diabetes mellitus |
| Group 1 | | | | | |
|  | **Kidney transplant** | | | | |
|  | must have | any of | diagnosis | UMLS:ICD10CM:Z94.0 | Kidney transplant status |
|  |  |  | procedure | UMLS:ICD10PCS:0TY00Z0 | Transplantation of Right Kidney, Allogeneic, Open Approach |
|  |  |  | procedure | UMLS:ICD10PCS:0TY10Z0 | Transplantation of Left Kidney, Allogeneic, Open Approach |
|  |  |  | procedure | UMLS:ICD10PCS:0TY10Z1 | Transplantation of Left Kidney, Syngeneic, Open Approach |
|  |  |  | procedure | UMLS:ICD10PCS:0TY10Z2 | Transplantation of Left Kidney, Zooplastic, Open Approach |
|  |  |  | procedure | UMLS:ICD10PCS:0TY00Z1 | Transplantation of Right Kidney, Syngeneic, Open Approach |
|  |  |  | procedure | UMLS:ICD10PCS:0TY00Z2 | Transplantation of Right Kidney, Zooplastic, Open Approach |
|  |  |  | procedure | UMLS:ICD10PCS:0TY0 | Urinary System / Transplantation / Kidney, Right |
|  |  |  | procedure | UMLS:ICD10PCS:0TY1 | Urinary System / Transplantation / Kidney, Left |
|  | date constraint | | The terms in this group occurred between Jan 1, 2015 and Jan 1, 2022 | | |
| Group 2 | | | | | |
|  | **Group 2A Kidney transplant** | | | | |
|  | must have | any of | diagnosis | UMLS:ICD10CM:Z94.0 | Kidney transplant status |
|  |  |  | procedure | UMLS:ICD10PCS:0TY00Z0 | Transplantation of Right Kidney, Allogeneic, Open Approach |
|  |  |  | procedure | UMLS:ICD10PCS:0TY10Z0 | Transplantation of Left Kidney, Allogeneic, Open Approach |
|  |  |  | procedure | UMLS:ICD10PCS:0TY10Z1 | Transplantation of Left Kidney, Syngeneic, Open Approach |
|  |  |  | procedure | UMLS:ICD10PCS:0TY10Z2 | Transplantation of Left Kidney, Zooplastic, Open Approach |
|  |  |  | procedure | UMLS:ICD10PCS:0TY00Z1 | Transplantation of Right Kidney, Syngeneic, Open Approach |
|  |  |  | procedure | UMLS:ICD10PCS:0TY00Z2 | Transplantation of Right Kidney, Zooplastic, Open Approach |
|  |  |  | procedure | UMLS:ICD10PCS:0TY0 | Urinary System / Transplantation / Kidney, Right |
|  |  |  | procedure | UMLS:ICD10PCS:0TY1 | Urinary System / Transplantation / Kidney, Left |
|  | date constraint | | The terms in this group occurred at any time | | |
|  | event relationship | | Any instance of noDialysis, Death, SGLT2i occurred within 1 day and 999 days after the first instance of Kidney transplant | | |
|  | **Group 2B noDialysis, Death, SGLT2i** | | | | |
|  | cannot have |  | procedure | UMLS:CPT:1012740 | Dialysis Services and Procedures |
|  |  | or | procedure | UMLS:CPT:90945 | Dialysis procedure other than hemodialysis (eg, peritoneal dialysis, hemofiltration, or other continuous renal replacement therapies), with single evaluation by a physician or other qualified health care professional |
|  |  | or | procedure | UMLS:CPT:90947 | Dialysis procedure other than hemodialysis (eg, peritoneal dialysis, hemofiltration, or other continuous renal replacement therapies) requiring repeated evaluations by a physician or other qualified health care professional, with or without substantial revision of dialysis prescription |
|  |  | or | procedure | UMLS:CPT:1012752 | Hemodialysis Procedures |
|  |  | or | procedure | UMLS:SNOMED:302497006 | Hemodialysis |
|  |  | or | procedure | UMLS:CPT:90937 | Hemodialysis procedure requiring repeated evaluation(s) with or without substantial revision of dialysis prescription |
|  |  | or | procedure | UMLS:CPT:90935 | Hemodialysis procedure with single evaluation by a physician or other qualified health care professional |
|  |  | or | demographics | Deceased | Deceased |
|  |  | or | diagnosis | UMLS:ICD10CM:R99 | Ill-defined and unknown cause of mortality |
|  |  | or | medication | NLM:ATC:A10BK | Sodium-glucose co-transporter 2 (SGLT2) inhibitors |
| Group 3 | | | | | |
|  | **Group 3A Kidney transplant** | | | | |
|  | must have | any of | diagnosis | UMLS:ICD10CM:Z94.0 | Kidney transplant status |
|  |  |  | procedure | UMLS:ICD10PCS:0TY00Z0 | Transplantation of Right Kidney, Allogeneic, Open Approach |
|  |  |  | procedure | UMLS:ICD10PCS:0TY10Z0 | Transplantation of Left Kidney, Allogeneic, Open Approach |
|  |  |  | procedure | UMLS:ICD10PCS:0TY10Z1 | Transplantation of Left Kidney, Syngeneic, Open Approach |
|  |  |  | procedure | UMLS:ICD10PCS:0TY10Z2 | Transplantation of Left Kidney, Zooplastic, Open Approach |
|  |  |  | procedure | UMLS:ICD10PCS:0TY00Z1 | Transplantation of Right Kidney, Syngeneic, Open Approach |
|  |  |  | procedure | UMLS:ICD10PCS:0TY00Z2 | Transplantation of Right Kidney, Zooplastic, Open Approach |
|  |  |  | procedure | UMLS:ICD10PCS:0TY0 | Urinary System / Transplantation / Kidney, Right |
|  |  |  | procedure | UMLS:ICD10PCS:0TY1 | Urinary System / Transplantation / Kidney, Left |
|  | date constraint | | The terms in this group occurred at any time | | |
|  | event relationship | | Any instance of SGLT2i occurred at least 1000 days after the first instance of Kidney transplant | | |
|  | **Group 3B SGLT2i** | | | | |
|  | must have |  | medication | NLM:ATC:A10BK | Sodium-glucose co-transporter 2 (SGLT2) inhibitors |
| Group 4 | | | | | |
|  | **Group 4A Kidney transplant** | | | | |
|  | must have | any of | diagnosis | UMLS:ICD10CM:Z94.0 | Kidney transplant status |
|  |  |  | procedure | UMLS:ICD10PCS:0TY00Z0 | Transplantation of Right Kidney, Allogeneic, Open Approach |
|  |  |  | procedure | UMLS:ICD10PCS:0TY10Z0 | Transplantation of Left Kidney, Allogeneic, Open Approach |
|  |  |  | procedure | UMLS:ICD10PCS:0TY10Z1 | Transplantation of Left Kidney, Syngeneic, Open Approach |
|  |  |  | procedure | UMLS:ICD10PCS:0TY10Z2 | Transplantation of Left Kidney, Zooplastic, Open Approach |
|  |  |  | procedure | UMLS:ICD10PCS:0TY00Z1 | Transplantation of Right Kidney, Syngeneic, Open Approach |
|  |  |  | procedure | UMLS:ICD10PCS:0TY00Z2 | Transplantation of Right Kidney, Zooplastic, Open Approach |
|  |  |  | procedure | UMLS:ICD10PCS:0TY0 | Urinary System / Transplantation / Kidney, Right |
|  |  |  | procedure | UMLS:ICD10PCS:0TY1 | Urinary System / Transplantation / Kidney, Left |
|  | date constraint | | The terms in this group occurred at any time | | |
|  | event relationship | | Any instance of noSGLT2i occurred at least 1 day before the first instance of Kidney transplant | | |
|  | **Group 4B noSGLT2i** | | | | |
|  | cannot have |  | medication | NLM:ATC:A10BK | Sodium-glucose co-transporter 2 (SGLT2) inhibitors |
| Group 5 | | | | | |
|  | **Group 5A Kidney transplant** | | | | |
|  | must have | any of | diagnosis | UMLS:ICD10CM:Z94.0 | Kidney transplant status |
|  |  |  | procedure | UMLS:ICD10PCS:0TY00Z0 | Transplantation of Right Kidney, Allogeneic, Open Approach |
|  |  |  | procedure | UMLS:ICD10PCS:0TY10Z0 | Transplantation of Left Kidney, Allogeneic, Open Approach |
|  |  |  | procedure | UMLS:ICD10PCS:0TY10Z1 | Transplantation of Left Kidney, Syngeneic, Open Approach |
|  |  |  | procedure | UMLS:ICD10PCS:0TY10Z2 | Transplantation of Left Kidney, Zooplastic, Open Approach |
|  |  |  | procedure | UMLS:ICD10PCS:0TY00Z1 | Transplantation of Right Kidney, Syngeneic, Open Approach |
|  |  |  | procedure | UMLS:ICD10PCS:0TY00Z2 | Transplantation of Right Kidney, Zooplastic, Open Approach |
|  |  |  | procedure | UMLS:ICD10PCS:0TY0 | Urinary System / Transplantation / Kidney, Right |
|  |  |  | procedure | UMLS:ICD10PCS:0TY1 | Urinary System / Transplantation / Kidney, Left |
|  | date constraint | | The terms in this group occurred at any time | | |
|  | event relationship | | Any instance of Visit occurred at least 1181 days after the first instance of Kidney transplant | | |
|  | **Group 5B Visit** | | | | |
|  | must have |  | visit | TNX:Visit | Visit |

##### **Table S32. Code definitions and selection criteria for SGLT2i non-users – ≥ 60 years.**

| Ungrouped terms | | | | | |
| --- | --- | --- | --- | --- | --- |
|  | must have |  | demographics | Age | Age (at least 60 years (most recent occurrence)) |
|  | cannot have |  | diagnosis | UMLS:ICD10CM:E08-E13 | Diabetes mellitus |
| Group 1 | | | | | |
|  | **Kidney transplant** | | | | |
|  | must have | any of | diagnosis | UMLS:ICD10CM:Z94.0 | Kidney transplant status |
|  |  |  | procedure | UMLS:ICD10PCS:0TY00Z0 | Transplantation of Right Kidney, Allogeneic, Open Approach |
|  |  |  | procedure | UMLS:ICD10PCS:0TY10Z0 | Transplantation of Left Kidney, Allogeneic, Open Approach |
|  |  |  | procedure | UMLS:ICD10PCS:0TY10Z1 | Transplantation of Left Kidney, Syngeneic, Open Approach |
|  |  |  | procedure | UMLS:ICD10PCS:0TY10Z2 | Transplantation of Left Kidney, Zooplastic, Open Approach |
|  |  |  | procedure | UMLS:ICD10PCS:0TY00Z1 | Transplantation of Right Kidney, Syngeneic, Open Approach |
|  |  |  | procedure | UMLS:ICD10PCS:0TY00Z2 | Transplantation of Right Kidney, Zooplastic, Open Approach |
|  |  |  | procedure | UMLS:ICD10PCS:0TY0 | Urinary System / Transplantation / Kidney, Right |
|  |  |  | procedure | UMLS:ICD10PCS:0TY1 | Urinary System / Transplantation / Kidney, Left |
|  | date constraint | | The terms in this group occurred between Jan 1, 2015 and Jan 1, 2022 | | |
| Group 2 | | | | | |
|  | **Group 2A Kidney transplant** | | | | |
|  | must have | any of | diagnosis | UMLS:ICD10CM:Z94.0 | Kidney transplant status |
|  |  |  | procedure | UMLS:ICD10PCS:0TY00Z0 | Transplantation of Right Kidney, Allogeneic, Open Approach |
|  |  |  | procedure | UMLS:ICD10PCS:0TY10Z0 | Transplantation of Left Kidney, Allogeneic, Open Approach |
|  |  |  | procedure | UMLS:ICD10PCS:0TY10Z1 | Transplantation of Left Kidney, Syngeneic, Open Approach |
|  |  |  | procedure | UMLS:ICD10PCS:0TY10Z2 | Transplantation of Left Kidney, Zooplastic, Open Approach |
|  |  |  | procedure | UMLS:ICD10PCS:0TY00Z1 | Transplantation of Right Kidney, Syngeneic, Open Approach |
|  |  |  | procedure | UMLS:ICD10PCS:0TY00Z2 | Transplantation of Right Kidney, Zooplastic, Open Approach |
|  |  |  | procedure | UMLS:ICD10PCS:0TY0 | Urinary System / Transplantation / Kidney, Right |
|  |  |  | procedure | UMLS:ICD10PCS:0TY1 | Urinary System / Transplantation / Kidney, Left |
|  | date constraint | | The terms in this group occurred at any time | | |
|  | event relationship | | Any instance of noDialysis, Death, SGLT2i occurred within 1 day and 999 days after the first instance of Kidney transplant | | |
|  | **Group 2B noDialysis, Death, SGLT2i** | | | | |
|  | cannot have |  | procedure | UMLS:CPT:1012740 | Dialysis Services and Procedures |
|  |  | or | procedure | UMLS:CPT:90945 | Dialysis procedure other than hemodialysis (eg, peritoneal dialysis, hemofiltration, or other continuous renal replacement therapies), with single evaluation by a physician or other qualified health care professional |
|  |  | or | procedure | UMLS:CPT:90947 | Dialysis procedure other than hemodialysis (eg, peritoneal dialysis, hemofiltration, or other continuous renal replacement therapies) requiring repeated evaluations by a physician or other qualified health care professional, with or without substantial revision of dialysis prescription |
|  |  | or | procedure | UMLS:CPT:1012752 | Hemodialysis Procedures |
|  |  | or | procedure | UMLS:SNOMED:302497006 | Hemodialysis |
|  |  | or | procedure | UMLS:CPT:90937 | Hemodialysis procedure requiring repeated evaluation(s) with or without substantial revision of dialysis prescription |
|  |  | or | procedure | UMLS:CPT:90935 | Hemodialysis procedure with single evaluation by a physician or other qualified health care professional |
|  |  | or | demographics | Deceased | Deceased |
|  |  | or | diagnosis | UMLS:ICD10CM:R99 | Ill-defined and unknown cause of mortality |
|  |  | or | medication | NLM:ATC:A10BK | Sodium-glucose co-transporter 2 (SGLT2) inhibitors |
| Group 3 | | | | | |
|  | **Group 3A Kidney transplant** | | | | |
|  | must have | any of | diagnosis | UMLS:ICD10CM:Z94.0 | Kidney transplant status |
|  |  |  | procedure | UMLS:ICD10PCS:0TY00Z0 | Transplantation of Right Kidney, Allogeneic, Open Approach |
|  |  |  | procedure | UMLS:ICD10PCS:0TY10Z0 | Transplantation of Left Kidney, Allogeneic, Open Approach |
|  |  |  | procedure | UMLS:ICD10PCS:0TY10Z1 | Transplantation of Left Kidney, Syngeneic, Open Approach |
|  |  |  | procedure | UMLS:ICD10PCS:0TY10Z2 | Transplantation of Left Kidney, Zooplastic, Open Approach |
|  |  |  | procedure | UMLS:ICD10PCS:0TY00Z1 | Transplantation of Right Kidney, Syngeneic, Open Approach |
|  |  |  | procedure | UMLS:ICD10PCS:0TY00Z2 | Transplantation of Right Kidney, Zooplastic, Open Approach |
|  |  |  | procedure | UMLS:ICD10PCS:0TY0 | Urinary System / Transplantation / Kidney, Right |
|  |  |  | procedure | UMLS:ICD10PCS:0TY1 | Urinary System / Transplantation / Kidney, Left |
|  | date constraint | | The terms in this group occurred at any time | | |
|  | event relationship | | Any instance of SGLT2i occurred within 1000 days and 3006 days after the first instance of Kidney transplant | | |
|  | **Group 3B noSGLT2i** | | | | |
|  | cannot have |  | medication | NLM:ATC:A10BK | Sodium-glucose co-transporter 2 (SGLT2) inhibitors |
| Group 4 | | | | | |
|  | **Group 4A Kidney transplant** | | | | |
|  | must have | any of | diagnosis | UMLS:ICD10CM:Z94.0 | Kidney transplant status |
|  |  |  | procedure | UMLS:ICD10PCS:0TY00Z0 | Transplantation of Right Kidney, Allogeneic, Open Approach |
|  |  |  | procedure | UMLS:ICD10PCS:0TY10Z0 | Transplantation of Left Kidney, Allogeneic, Open Approach |
|  |  |  | procedure | UMLS:ICD10PCS:0TY10Z1 | Transplantation of Left Kidney, Syngeneic, Open Approach |
|  |  |  | procedure | UMLS:ICD10PCS:0TY10Z2 | Transplantation of Left Kidney, Zooplastic, Open Approach |
|  |  |  | procedure | UMLS:ICD10PCS:0TY00Z1 | Transplantation of Right Kidney, Syngeneic, Open Approach |
|  |  |  | procedure | UMLS:ICD10PCS:0TY00Z2 | Transplantation of Right Kidney, Zooplastic, Open Approach |
|  |  |  | procedure | UMLS:ICD10PCS:0TY0 | Urinary System / Transplantation / Kidney, Right |
|  |  |  | procedure | UMLS:ICD10PCS:0TY1 | Urinary System / Transplantation / Kidney, Left |
|  | date constraint | | The terms in this group occurred at any time | | |
|  | event relationship | | Any instance of noSGLT2i occurred at least 1 day before the first instance of Kidney transplant | | |
|  | **Group 4B noSGLT2i** | | | | |
|  | cannot have |  | medication | NLM:ATC:A10BK | Sodium-glucose co-transporter 2 (SGLT2) inhibitors |
| Group 5 | | | | | |
|  | **Group 5A Kidney transplant** | | | | |
|  | must have | any of | diagnosis | UMLS:ICD10CM:Z94.0 | Kidney transplant status |
|  |  |  | procedure | UMLS:ICD10PCS:0TY00Z0 | Transplantation of Right Kidney, Allogeneic, Open Approach |
|  |  |  | procedure | UMLS:ICD10PCS:0TY10Z0 | Transplantation of Left Kidney, Allogeneic, Open Approach |
|  |  |  | procedure | UMLS:ICD10PCS:0TY10Z1 | Transplantation of Left Kidney, Syngeneic, Open Approach |
|  |  |  | procedure | UMLS:ICD10PCS:0TY10Z2 | Transplantation of Left Kidney, Zooplastic, Open Approach |
|  |  |  | procedure | UMLS:ICD10PCS:0TY00Z1 | Transplantation of Right Kidney, Syngeneic, Open Approach |
|  |  |  | procedure | UMLS:ICD10PCS:0TY00Z2 | Transplantation of Right Kidney, Zooplastic, Open Approach |
|  |  |  | procedure | UMLS:ICD10PCS:0TY0 | Urinary System / Transplantation / Kidney, Right |
|  |  |  | procedure | UMLS:ICD10PCS:0TY1 | Urinary System / Transplantation / Kidney, Left |
|  | date constraint | | The terms in this group occurred at any time | | |
|  | event relationship | | Any instance of Visit occurred at least 1181 days after the first instance of Kidney transplant | | |
|  | **Group 5B Visit** | | | | |
|  | must have |  | visit | TNX:Visit | Visit |

#### Comorbidities

##### **Table S33. Code definitions and selection criteria for SGLT2i users – hypertension.**

| Ungrouped terms | | | | | |
| --- | --- | --- | --- | --- | --- |
|  | must have |  | demographics | Age | Age (at least 18 years (most recent occurrence)) |
|  | cannot have |  | diagnosis | UMLS:ICD10CM:E08-E13 | Diabetes mellitus |
| Group 1 | | | | | |
|  | **Kidney transplant** | | | | |
|  | must have | any of | diagnosis | UMLS:ICD10CM:Z94.0 | Kidney transplant status |
|  |  |  | procedure | UMLS:ICD10PCS:0TY00Z0 | Transplantation of Right Kidney, Allogeneic, Open Approach |
|  |  |  | procedure | UMLS:ICD10PCS:0TY10Z0 | Transplantation of Left Kidney, Allogeneic, Open Approach |
|  |  |  | procedure | UMLS:ICD10PCS:0TY10Z1 | Transplantation of Left Kidney, Syngeneic, Open Approach |
|  |  |  | procedure | UMLS:ICD10PCS:0TY10Z2 | Transplantation of Left Kidney, Zooplastic, Open Approach |
|  |  |  | procedure | UMLS:ICD10PCS:0TY00Z1 | Transplantation of Right Kidney, Syngeneic, Open Approach |
|  |  |  | procedure | UMLS:ICD10PCS:0TY00Z2 | Transplantation of Right Kidney, Zooplastic, Open Approach |
|  |  |  | procedure | UMLS:ICD10PCS:0TY0 | Urinary System / Transplantation / Kidney, Right |
|  |  |  | procedure | UMLS:ICD10PCS:0TY1 | Urinary System / Transplantation / Kidney, Left |
|  | date constraint | | The terms in this group occurred between Jan 1, 2015 and Jan 1, 2022 | | |
| Group 2 | | | | | |
|  | **Group 2A Kidney transplant** | | | | |
|  | must have | any of | diagnosis | UMLS:ICD10CM:Z94.0 | Kidney transplant status |
|  |  |  | procedure | UMLS:ICD10PCS:0TY00Z0 | Transplantation of Right Kidney, Allogeneic, Open Approach |
|  |  |  | procedure | UMLS:ICD10PCS:0TY10Z0 | Transplantation of Left Kidney, Allogeneic, Open Approach |
|  |  |  | procedure | UMLS:ICD10PCS:0TY10Z1 | Transplantation of Left Kidney, Syngeneic, Open Approach |
|  |  |  | procedure | UMLS:ICD10PCS:0TY10Z2 | Transplantation of Left Kidney, Zooplastic, Open Approach |
|  |  |  | procedure | UMLS:ICD10PCS:0TY00Z1 | Transplantation of Right Kidney, Syngeneic, Open Approach |
|  |  |  | procedure | UMLS:ICD10PCS:0TY00Z2 | Transplantation of Right Kidney, Zooplastic, Open Approach |
|  |  |  | procedure | UMLS:ICD10PCS:0TY0 | Urinary System / Transplantation / Kidney, Right |
|  |  |  | procedure | UMLS:ICD10PCS:0TY1 | Urinary System / Transplantation / Kidney, Left |
|  | date constraint | | The terms in this group occurred at any time | | |
|  | event relationship | | Any instance of noDialysis, Death, SGLT2i occurred within 1 day and 999 days after the first instance of Kidney transplant | | |
|  | **Group 2B noDialysis, Death, SGLT2i** | | | | |
|  | cannot have |  | procedure | UMLS:CPT:1012740 | Dialysis Services and Procedures |
|  |  | or | procedure | UMLS:CPT:90945 | Dialysis procedure other than hemodialysis (eg, peritoneal dialysis, hemofiltration, or other continuous renal replacement therapies), with single evaluation by a physician or other qualified health care professional |
|  |  | or | procedure | UMLS:CPT:90947 | Dialysis procedure other than hemodialysis (eg, peritoneal dialysis, hemofiltration, or other continuous renal replacement therapies) requiring repeated evaluations by a physician or other qualified health care professional, with or without substantial revision of dialysis prescription |
|  |  | or | procedure | UMLS:CPT:1012752 | Hemodialysis Procedures |
|  |  | or | procedure | UMLS:SNOMED:302497006 | Hemodialysis |
|  |  | or | procedure | UMLS:CPT:90937 | Hemodialysis procedure requiring repeated evaluation(s) with or without substantial revision of dialysis prescription |
|  |  | or | procedure | UMLS:CPT:90935 | Hemodialysis procedure with single evaluation by a physician or other qualified health care professional |
|  |  | or | demographics | Deceased | Deceased |
|  |  | or | diagnosis | UMLS:ICD10CM:R99 | Ill-defined and unknown cause of mortality |
|  |  | or | medication | NLM:ATC:A10BK | Sodium-glucose co-transporter 2 (SGLT2) inhibitors |
| Group 3 | | | | | |
|  | **Group 3A Kidney transplant** | | | | |
|  | must have | any of | diagnosis | UMLS:ICD10CM:Z94.0 | Kidney transplant status |
|  |  |  | procedure | UMLS:ICD10PCS:0TY00Z0 | Transplantation of Right Kidney, Allogeneic, Open Approach |
|  |  |  | procedure | UMLS:ICD10PCS:0TY10Z0 | Transplantation of Left Kidney, Allogeneic, Open Approach |
|  |  |  | procedure | UMLS:ICD10PCS:0TY10Z1 | Transplantation of Left Kidney, Syngeneic, Open Approach |
|  |  |  | procedure | UMLS:ICD10PCS:0TY10Z2 | Transplantation of Left Kidney, Zooplastic, Open Approach |
|  |  |  | procedure | UMLS:ICD10PCS:0TY00Z1 | Transplantation of Right Kidney, Syngeneic, Open Approach |
|  |  |  | procedure | UMLS:ICD10PCS:0TY00Z2 | Transplantation of Right Kidney, Zooplastic, Open Approach |
|  |  |  | procedure | UMLS:ICD10PCS:0TY0 | Urinary System / Transplantation / Kidney, Right |
|  |  |  | procedure | UMLS:ICD10PCS:0TY1 | Urinary System / Transplantation / Kidney, Left |
|  | date constraint | | The terms in this group occurred at any time | | |
|  | event relationship | | Any instance of SGLT2i occurred at least 1000 days after the first instance of Kidney transplant | | |
|  | **Group 3B SGLT2i** | | | | |
|  | must have |  | medication | NLM:ATC:A10BK | Sodium-glucose co-transporter 2 (SGLT2) inhibitors |
| Group 4 | | | | | |
|  | **Group 4A Kidney transplant** | | | | |
|  | must have | any of | diagnosis | UMLS:ICD10CM:Z94.0 | Kidney transplant status |
|  |  |  | procedure | UMLS:ICD10PCS:0TY00Z0 | Transplantation of Right Kidney, Allogeneic, Open Approach |
|  |  |  | procedure | UMLS:ICD10PCS:0TY10Z0 | Transplantation of Left Kidney, Allogeneic, Open Approach |
|  |  |  | procedure | UMLS:ICD10PCS:0TY10Z1 | Transplantation of Left Kidney, Syngeneic, Open Approach |
|  |  |  | procedure | UMLS:ICD10PCS:0TY10Z2 | Transplantation of Left Kidney, Zooplastic, Open Approach |
|  |  |  | procedure | UMLS:ICD10PCS:0TY00Z1 | Transplantation of Right Kidney, Syngeneic, Open Approach |
|  |  |  | procedure | UMLS:ICD10PCS:0TY00Z2 | Transplantation of Right Kidney, Zooplastic, Open Approach |
|  |  |  | procedure | UMLS:ICD10PCS:0TY0 | Urinary System / Transplantation / Kidney, Right |
|  |  |  | procedure | UMLS:ICD10PCS:0TY1 | Urinary System / Transplantation / Kidney, Left |
|  | date constraint | | The terms in this group occurred at any time | | |
|  | event relationship | | Any instance of noSGLT2i occurred at least 1 day before the first instance of Kidney transplant | | |
|  | **Group 4B noSGLT2i** | | | | |
|  | cannot have |  | medication | NLM:ATC:A10BK | Sodium-glucose co-transporter 2 (SGLT2) inhibitors |
| Group 5 | | | | | |
|  | **Group 5A Kidney transplant** | | | | |
|  | must have | any of | diagnosis | UMLS:ICD10CM:Z94.0 | Kidney transplant status |
|  |  |  | procedure | UMLS:ICD10PCS:0TY00Z0 | Transplantation of Right Kidney, Allogeneic, Open Approach |
|  |  |  | procedure | UMLS:ICD10PCS:0TY10Z0 | Transplantation of Left Kidney, Allogeneic, Open Approach |
|  |  |  | procedure | UMLS:ICD10PCS:0TY10Z1 | Transplantation of Left Kidney, Syngeneic, Open Approach |
|  |  |  | procedure | UMLS:ICD10PCS:0TY10Z2 | Transplantation of Left Kidney, Zooplastic, Open Approach |
|  |  |  | procedure | UMLS:ICD10PCS:0TY00Z1 | Transplantation of Right Kidney, Syngeneic, Open Approach |
|  |  |  | procedure | UMLS:ICD10PCS:0TY00Z2 | Transplantation of Right Kidney, Zooplastic, Open Approach |
|  |  |  | procedure | UMLS:ICD10PCS:0TY0 | Urinary System / Transplantation / Kidney, Right |
|  |  |  | procedure | UMLS:ICD10PCS:0TY1 | Urinary System / Transplantation / Kidney, Left |
|  | date constraint | | The terms in this group occurred at any time | | |
|  | event relationship | | Any instance of Visit occurred at least 1181 days after the first instance of Kidney transplant | | |
|  | **Group 5B Visit** | | | | |
|  | must have |  | visit | TNX:Visit | Visit |
| Group 6 | | | | | |
|  | **Group 6A Kidney transplant** | | | | |
|  | must have | any of | diagnosis | UMLS:ICD10CM:Z94.0 | Kidney transplant status |
|  |  |  | procedure | UMLS:ICD10PCS:0TY00Z0 | Transplantation of Right Kidney, Allogeneic, Open Approach |
|  |  |  | procedure | UMLS:ICD10PCS:0TY10Z0 | Transplantation of Left Kidney, Allogeneic, Open Approach |
|  |  |  | procedure | UMLS:ICD10PCS:0TY10Z1 | Transplantation of Left Kidney, Syngeneic, Open Approach |
|  |  |  | procedure | UMLS:ICD10PCS:0TY10Z2 | Transplantation of Left Kidney, Zooplastic, Open Approach |
|  |  |  | procedure | UMLS:ICD10PCS:0TY00Z1 | Transplantation of Right Kidney, Syngeneic, Open Approach |
|  |  |  | procedure | UMLS:ICD10PCS:0TY00Z2 | Transplantation of Right Kidney, Zooplastic, Open Approach |
|  |  |  | procedure | UMLS:ICD10PCS:0TY0 | Urinary System / Transplantation / Kidney, Right |
|  |  |  | procedure | UMLS:ICD10PCS:0TY1 | Urinary System / Transplantation / Kidney, Left |
|  | date constraint | | The terms in this group occurred at any time | | |
|  | event relationship | | Any instance of Hypertension occurred within 1 day and 999 days after the first instance of Kidney transplant | | |
|  | **Group 6B Hypertension** | | | | |
|  | must have |  | diagnosis | UMLS:ICD10CM:I10 | Essential (primary) hypertension |

##### **Table S34. Code definitions and selection criteria for SGLT2i non-users – hypertension.**

| Ungrouped terms | | | | | |
| --- | --- | --- | --- | --- | --- |
|  | must have |  | demographics | Age | Age (at least 18 years (most recent occurrence)) |
|  | cannot have |  | diagnosis | UMLS:ICD10CM:E08-E13 | Diabetes mellitus |
| Group 1 | | | | | |
|  | **Kidney transplant** | | | | |
|  | must have | any of | diagnosis | UMLS:ICD10CM:Z94.0 | Kidney transplant status |
|  |  |  | procedure | UMLS:ICD10PCS:0TY00Z0 | Transplantation of Right Kidney, Allogeneic, Open Approach |
|  |  |  | procedure | UMLS:ICD10PCS:0TY10Z0 | Transplantation of Left Kidney, Allogeneic, Open Approach |
|  |  |  | procedure | UMLS:ICD10PCS:0TY10Z1 | Transplantation of Left Kidney, Syngeneic, Open Approach |
|  |  |  | procedure | UMLS:ICD10PCS:0TY10Z2 | Transplantation of Left Kidney, Zooplastic, Open Approach |
|  |  |  | procedure | UMLS:ICD10PCS:0TY00Z1 | Transplantation of Right Kidney, Syngeneic, Open Approach |
|  |  |  | procedure | UMLS:ICD10PCS:0TY00Z2 | Transplantation of Right Kidney, Zooplastic, Open Approach |
|  |  |  | procedure | UMLS:ICD10PCS:0TY0 | Urinary System / Transplantation / Kidney, Right |
|  |  |  | procedure | UMLS:ICD10PCS:0TY1 | Urinary System / Transplantation / Kidney, Left |
|  | date constraint | | The terms in this group occurred between Jan 1, 2015 and Jan 1, 2022 | | |
| Group 2 | | | | | |
|  | **Group 2A Kidney transplant** | | | | |
|  | must have | any of | diagnosis | UMLS:ICD10CM:Z94.0 | Kidney transplant status |
|  |  |  | procedure | UMLS:ICD10PCS:0TY00Z0 | Transplantation of Right Kidney, Allogeneic, Open Approach |
|  |  |  | procedure | UMLS:ICD10PCS:0TY10Z0 | Transplantation of Left Kidney, Allogeneic, Open Approach |
|  |  |  | procedure | UMLS:ICD10PCS:0TY10Z1 | Transplantation of Left Kidney, Syngeneic, Open Approach |
|  |  |  | procedure | UMLS:ICD10PCS:0TY10Z2 | Transplantation of Left Kidney, Zooplastic, Open Approach |
|  |  |  | procedure | UMLS:ICD10PCS:0TY00Z1 | Transplantation of Right Kidney, Syngeneic, Open Approach |
|  |  |  | procedure | UMLS:ICD10PCS:0TY00Z2 | Transplantation of Right Kidney, Zooplastic, Open Approach |
|  |  |  | procedure | UMLS:ICD10PCS:0TY0 | Urinary System / Transplantation / Kidney, Right |
|  |  |  | procedure | UMLS:ICD10PCS:0TY1 | Urinary System / Transplantation / Kidney, Left |
|  | date constraint | | The terms in this group occurred at any time | | |
|  | event relationship | | Any instance of noDialysis, Death, SGLT2i occurred within 1 day and 999 days after the first instance of Kidney transplant | | |
|  | **Group 2B noDialysis, Death, SGLT2i** | | | | |
|  | cannot have |  | procedure | UMLS:CPT:1012740 | Dialysis Services and Procedures |
|  |  | or | procedure | UMLS:CPT:90945 | Dialysis procedure other than hemodialysis (eg, peritoneal dialysis, hemofiltration, or other continuous renal replacement therapies), with single evaluation by a physician or other qualified health care professional |
|  |  | or | procedure | UMLS:CPT:90947 | Dialysis procedure other than hemodialysis (eg, peritoneal dialysis, hemofiltration, or other continuous renal replacement therapies) requiring repeated evaluations by a physician or other qualified health care professional, with or without substantial revision of dialysis prescription |
|  |  | or | procedure | UMLS:CPT:1012752 | Hemodialysis Procedures |
|  |  | or | procedure | UMLS:SNOMED:302497006 | Hemodialysis |
|  |  | or | procedure | UMLS:CPT:90937 | Hemodialysis procedure requiring repeated evaluation(s) with or without substantial revision of dialysis prescription |
|  |  | or | procedure | UMLS:CPT:90935 | Hemodialysis procedure with single evaluation by a physician or other qualified health care professional |
|  |  | or | demographics | Deceased | Deceased |
|  |  | or | diagnosis | UMLS:ICD10CM:R99 | Ill-defined and unknown cause of mortality |
|  |  | or | medication | NLM:ATC:A10BK | Sodium-glucose co-transporter 2 (SGLT2) inhibitors |
| Group 3 | | | | | |
|  | **Group 3A Kidney transplant** | | | | |
|  | must have | any of | diagnosis | UMLS:ICD10CM:Z94.0 | Kidney transplant status |
|  |  |  | procedure | UMLS:ICD10PCS:0TY00Z0 | Transplantation of Right Kidney, Allogeneic, Open Approach |
|  |  |  | procedure | UMLS:ICD10PCS:0TY10Z0 | Transplantation of Left Kidney, Allogeneic, Open Approach |
|  |  |  | procedure | UMLS:ICD10PCS:0TY10Z1 | Transplantation of Left Kidney, Syngeneic, Open Approach |
|  |  |  | procedure | UMLS:ICD10PCS:0TY10Z2 | Transplantation of Left Kidney, Zooplastic, Open Approach |
|  |  |  | procedure | UMLS:ICD10PCS:0TY00Z1 | Transplantation of Right Kidney, Syngeneic, Open Approach |
|  |  |  | procedure | UMLS:ICD10PCS:0TY00Z2 | Transplantation of Right Kidney, Zooplastic, Open Approach |
|  |  |  | procedure | UMLS:ICD10PCS:0TY0 | Urinary System / Transplantation / Kidney, Right |
|  |  |  | procedure | UMLS:ICD10PCS:0TY1 | Urinary System / Transplantation / Kidney, Left |
|  | date constraint | | The terms in this group occurred at any time | | |
|  | event relationship | | Any instance of SGLT2i occurred within 1000 days and 3006 days after the first instance of Kidney transplant | | |
|  | **Group 3B noSGLT2i** | | | | |
|  | cannot have |  | medication | NLM:ATC:A10BK | Sodium-glucose co-transporter 2 (SGLT2) inhibitors |
| Group 4 | | | | | |
|  | **Group 4A Kidney transplant** | | | | |
|  | must have | any of | diagnosis | UMLS:ICD10CM:Z94.0 | Kidney transplant status |
|  |  |  | procedure | UMLS:ICD10PCS:0TY00Z0 | Transplantation of Right Kidney, Allogeneic, Open Approach |
|  |  |  | procedure | UMLS:ICD10PCS:0TY10Z0 | Transplantation of Left Kidney, Allogeneic, Open Approach |
|  |  |  | procedure | UMLS:ICD10PCS:0TY10Z1 | Transplantation of Left Kidney, Syngeneic, Open Approach |
|  |  |  | procedure | UMLS:ICD10PCS:0TY10Z2 | Transplantation of Left Kidney, Zooplastic, Open Approach |
|  |  |  | procedure | UMLS:ICD10PCS:0TY00Z1 | Transplantation of Right Kidney, Syngeneic, Open Approach |
|  |  |  | procedure | UMLS:ICD10PCS:0TY00Z2 | Transplantation of Right Kidney, Zooplastic, Open Approach |
|  |  |  | procedure | UMLS:ICD10PCS:0TY0 | Urinary System / Transplantation / Kidney, Right |
|  |  |  | procedure | UMLS:ICD10PCS:0TY1 | Urinary System / Transplantation / Kidney, Left |
|  | date constraint | | The terms in this group occurred at any time | | |
|  | event relationship | | Any instance of noSGLT2i occurred at least 1 day before the first instance of Kidney transplant | | |
|  | **Group 4B noSGLT2i** | | | | |
|  | cannot have |  | medication | NLM:ATC:A10BK | Sodium-glucose co-transporter 2 (SGLT2) inhibitors |
| Group 5 | | | | | |
|  | **Group 5A Kidney transplant** | | | | |
|  | must have | any of | diagnosis | UMLS:ICD10CM:Z94.0 | Kidney transplant status |
|  |  |  | procedure | UMLS:ICD10PCS:0TY00Z0 | Transplantation of Right Kidney, Allogeneic, Open Approach |
|  |  |  | procedure | UMLS:ICD10PCS:0TY10Z0 | Transplantation of Left Kidney, Allogeneic, Open Approach |
|  |  |  | procedure | UMLS:ICD10PCS:0TY10Z1 | Transplantation of Left Kidney, Syngeneic, Open Approach |
|  |  |  | procedure | UMLS:ICD10PCS:0TY10Z2 | Transplantation of Left Kidney, Zooplastic, Open Approach |
|  |  |  | procedure | UMLS:ICD10PCS:0TY00Z1 | Transplantation of Right Kidney, Syngeneic, Open Approach |
|  |  |  | procedure | UMLS:ICD10PCS:0TY00Z2 | Transplantation of Right Kidney, Zooplastic, Open Approach |
|  |  |  | procedure | UMLS:ICD10PCS:0TY0 | Urinary System / Transplantation / Kidney, Right |
|  |  |  | procedure | UMLS:ICD10PCS:0TY1 | Urinary System / Transplantation / Kidney, Left |
|  | date constraint | | The terms in this group occurred at any time | | |
|  | event relationship | | Any instance of Visit occurred at least 1181 days after the first instance of Kidney transplant | | |
|  | **Group 5B Visit** | | | | |
|  | must have |  | visit | TNX:Visit | Visit |
| Group 6 | | | | | |
|  | **Group 6A Kidney transplant** | | | | |
|  | must have | any of | diagnosis | UMLS:ICD10CM:Z94.0 | Kidney transplant status |
|  |  |  | procedure | UMLS:ICD10PCS:0TY00Z0 | Transplantation of Right Kidney, Allogeneic, Open Approach |
|  |  |  | procedure | UMLS:ICD10PCS:0TY10Z0 | Transplantation of Left Kidney, Allogeneic, Open Approach |
|  |  |  | procedure | UMLS:ICD10PCS:0TY10Z1 | Transplantation of Left Kidney, Syngeneic, Open Approach |
|  |  |  | procedure | UMLS:ICD10PCS:0TY10Z2 | Transplantation of Left Kidney, Zooplastic, Open Approach |
|  |  |  | procedure | UMLS:ICD10PCS:0TY00Z1 | Transplantation of Right Kidney, Syngeneic, Open Approach |
|  |  |  | procedure | UMLS:ICD10PCS:0TY00Z2 | Transplantation of Right Kidney, Zooplastic, Open Approach |
|  |  |  | procedure | UMLS:ICD10PCS:0TY0 | Urinary System / Transplantation / Kidney, Right |
|  |  |  | procedure | UMLS:ICD10PCS:0TY1 | Urinary System / Transplantation / Kidney, Left |
|  | date constraint | | The terms in this group occurred at any time | | |
|  | event relationship | | Any instance of Hypertension occurred within 1 day and 999 days after the first instance of Kidney transplant | | |
|  | **Group 6B Hypertension** | | | | |
|  | must have |  | diagnosis | UMLS:ICD10CM:I10 | Essential (primary) hypertension |

##### **Table S35. Code definitions and selection criteria for SGLT2i users – no hypertension.**

| Ungrouped terms | | | | | |
| --- | --- | --- | --- | --- | --- |
|  | must have |  | demographics | Age | Age (at least 18 years (most recent occurrence)) |
|  | cannot have |  | diagnosis | UMLS:ICD10CM:E08-E13 | Diabetes mellitus |
| Group 1 | | | | | |
|  | **Kidney transplant** | | | | |
|  | must have | any of | diagnosis | UMLS:ICD10CM:Z94.0 | Kidney transplant status |
|  |  |  | procedure | UMLS:ICD10PCS:0TY00Z0 | Transplantation of Right Kidney, Allogeneic, Open Approach |
|  |  |  | procedure | UMLS:ICD10PCS:0TY10Z0 | Transplantation of Left Kidney, Allogeneic, Open Approach |
|  |  |  | procedure | UMLS:ICD10PCS:0TY10Z1 | Transplantation of Left Kidney, Syngeneic, Open Approach |
|  |  |  | procedure | UMLS:ICD10PCS:0TY10Z2 | Transplantation of Left Kidney, Zooplastic, Open Approach |
|  |  |  | procedure | UMLS:ICD10PCS:0TY00Z1 | Transplantation of Right Kidney, Syngeneic, Open Approach |
|  |  |  | procedure | UMLS:ICD10PCS:0TY00Z2 | Transplantation of Right Kidney, Zooplastic, Open Approach |
|  |  |  | procedure | UMLS:ICD10PCS:0TY0 | Urinary System / Transplantation / Kidney, Right |
|  |  |  | procedure | UMLS:ICD10PCS:0TY1 | Urinary System / Transplantation / Kidney, Left |
|  | date constraint | | The terms in this group occurred between Jan 1, 2015 and Jan 1, 2022 | | |
| Group 2 | | | | | |
|  | **Group 2A Kidney transplant** | | | | |
|  | must have | any of | diagnosis | UMLS:ICD10CM:Z94.0 | Kidney transplant status |
|  |  |  | procedure | UMLS:ICD10PCS:0TY00Z0 | Transplantation of Right Kidney, Allogeneic, Open Approach |
|  |  |  | procedure | UMLS:ICD10PCS:0TY10Z0 | Transplantation of Left Kidney, Allogeneic, Open Approach |
|  |  |  | procedure | UMLS:ICD10PCS:0TY10Z1 | Transplantation of Left Kidney, Syngeneic, Open Approach |
|  |  |  | procedure | UMLS:ICD10PCS:0TY10Z2 | Transplantation of Left Kidney, Zooplastic, Open Approach |
|  |  |  | procedure | UMLS:ICD10PCS:0TY00Z1 | Transplantation of Right Kidney, Syngeneic, Open Approach |
|  |  |  | procedure | UMLS:ICD10PCS:0TY00Z2 | Transplantation of Right Kidney, Zooplastic, Open Approach |
|  |  |  | procedure | UMLS:ICD10PCS:0TY0 | Urinary System / Transplantation / Kidney, Right |
|  |  |  | procedure | UMLS:ICD10PCS:0TY1 | Urinary System / Transplantation / Kidney, Left |
|  | date constraint | | The terms in this group occurred at any time | | |
|  | event relationship | | Any instance of noDialysis, Death, SGLT2i occurred within 1 day and 999 days after the first instance of Kidney transplant | | |
|  | **Group 2B noDialysis, Death, SGLT2i** | | | | |
|  | cannot have |  | procedure | UMLS:CPT:1012740 | Dialysis Services and Procedures |
|  |  | or | procedure | UMLS:CPT:90945 | Dialysis procedure other than hemodialysis (eg, peritoneal dialysis, hemofiltration, or other continuous renal replacement therapies), with single evaluation by a physician or other qualified health care professional |
|  |  | or | procedure | UMLS:CPT:90947 | Dialysis procedure other than hemodialysis (eg, peritoneal dialysis, hemofiltration, or other continuous renal replacement therapies) requiring repeated evaluations by a physician or other qualified health care professional, with or without substantial revision of dialysis prescription |
|  |  | or | procedure | UMLS:CPT:1012752 | Hemodialysis Procedures |
|  |  | or | procedure | UMLS:SNOMED:302497006 | Hemodialysis |
|  |  | or | procedure | UMLS:CPT:90937 | Hemodialysis procedure requiring repeated evaluation(s) with or without substantial revision of dialysis prescription |
|  |  | or | procedure | UMLS:CPT:90935 | Hemodialysis procedure with single evaluation by a physician or other qualified health care professional |
|  |  | or | demographics | Deceased | Deceased |
|  |  | or | diagnosis | UMLS:ICD10CM:R99 | Ill-defined and unknown cause of mortality |
|  |  | or | medication | NLM:ATC:A10BK | Sodium-glucose co-transporter 2 (SGLT2) inhibitors |
| Group 3 | | | | | |
|  | **Group 3A Kidney transplant** | | | | |
|  | must have | any of | diagnosis | UMLS:ICD10CM:Z94.0 | Kidney transplant status |
|  |  |  | procedure | UMLS:ICD10PCS:0TY00Z0 | Transplantation of Right Kidney, Allogeneic, Open Approach |
|  |  |  | procedure | UMLS:ICD10PCS:0TY10Z0 | Transplantation of Left Kidney, Allogeneic, Open Approach |
|  |  |  | procedure | UMLS:ICD10PCS:0TY10Z1 | Transplantation of Left Kidney, Syngeneic, Open Approach |
|  |  |  | procedure | UMLS:ICD10PCS:0TY10Z2 | Transplantation of Left Kidney, Zooplastic, Open Approach |
|  |  |  | procedure | UMLS:ICD10PCS:0TY00Z1 | Transplantation of Right Kidney, Syngeneic, Open Approach |
|  |  |  | procedure | UMLS:ICD10PCS:0TY00Z2 | Transplantation of Right Kidney, Zooplastic, Open Approach |
|  |  |  | procedure | UMLS:ICD10PCS:0TY0 | Urinary System / Transplantation / Kidney, Right |
|  |  |  | procedure | UMLS:ICD10PCS:0TY1 | Urinary System / Transplantation / Kidney, Left |
|  | date constraint | | The terms in this group occurred at any time | | |
|  | event relationship | | Any instance of SGLT2i occurred at least 1000 days after the first instance of Kidney transplant | | |
|  | **Group 3B SGLT2i** | | | | |
|  | must have |  | medication | NLM:ATC:A10BK | Sodium-glucose co-transporter 2 (SGLT2) inhibitors |
| Group 4 | | | | | |
|  | **Group 4A Kidney transplant** | | | | |
|  | must have | any of | diagnosis | UMLS:ICD10CM:Z94.0 | Kidney transplant status |
|  |  |  | procedure | UMLS:ICD10PCS:0TY00Z0 | Transplantation of Right Kidney, Allogeneic, Open Approach |
|  |  |  | procedure | UMLS:ICD10PCS:0TY10Z0 | Transplantation of Left Kidney, Allogeneic, Open Approach |
|  |  |  | procedure | UMLS:ICD10PCS:0TY10Z1 | Transplantation of Left Kidney, Syngeneic, Open Approach |
|  |  |  | procedure | UMLS:ICD10PCS:0TY10Z2 | Transplantation of Left Kidney, Zooplastic, Open Approach |
|  |  |  | procedure | UMLS:ICD10PCS:0TY00Z1 | Transplantation of Right Kidney, Syngeneic, Open Approach |
|  |  |  | procedure | UMLS:ICD10PCS:0TY00Z2 | Transplantation of Right Kidney, Zooplastic, Open Approach |
|  |  |  | procedure | UMLS:ICD10PCS:0TY0 | Urinary System / Transplantation / Kidney, Right |
|  |  |  | procedure | UMLS:ICD10PCS:0TY1 | Urinary System / Transplantation / Kidney, Left |
|  | date constraint | | The terms in this group occurred at any time | | |
|  | event relationship | | Any instance of noSGLT2i occurred at least 1 day before the first instance of Kidney transplant | | |
|  | **Group 4B noSGLT2i** | | | | |
|  | cannot have |  | medication | NLM:ATC:A10BK | Sodium-glucose co-transporter 2 (SGLT2) inhibitors |
| Group 5 | | | | | |
|  | **Group 5A Kidney transplant** | | | | |
|  | must have | any of | diagnosis | UMLS:ICD10CM:Z94.0 | Kidney transplant status |
|  |  |  | procedure | UMLS:ICD10PCS:0TY00Z0 | Transplantation of Right Kidney, Allogeneic, Open Approach |
|  |  |  | procedure | UMLS:ICD10PCS:0TY10Z0 | Transplantation of Left Kidney, Allogeneic, Open Approach |
|  |  |  | procedure | UMLS:ICD10PCS:0TY10Z1 | Transplantation of Left Kidney, Syngeneic, Open Approach |
|  |  |  | procedure | UMLS:ICD10PCS:0TY10Z2 | Transplantation of Left Kidney, Zooplastic, Open Approach |
|  |  |  | procedure | UMLS:ICD10PCS:0TY00Z1 | Transplantation of Right Kidney, Syngeneic, Open Approach |
|  |  |  | procedure | UMLS:ICD10PCS:0TY00Z2 | Transplantation of Right Kidney, Zooplastic, Open Approach |
|  |  |  | procedure | UMLS:ICD10PCS:0TY0 | Urinary System / Transplantation / Kidney, Right |
|  |  |  | procedure | UMLS:ICD10PCS:0TY1 | Urinary System / Transplantation / Kidney, Left |
|  | date constraint | | The terms in this group occurred at any time | | |
|  | event relationship | | Any instance of Visit occurred at least 1181 days after the first instance of Kidney transplant | | |
|  | **Group 5B Visit** | | | | |
|  | must have |  | visit | TNX:Visit | Visit |
| Group 6 | | | | | |
|  | **Group 6A Kidney transplant** | | | | |
|  | must have | any of | diagnosis | UMLS:ICD10CM:Z94.0 | Kidney transplant status |
|  |  |  | procedure | UMLS:ICD10PCS:0TY00Z0 | Transplantation of Right Kidney, Allogeneic, Open Approach |
|  |  |  | procedure | UMLS:ICD10PCS:0TY10Z0 | Transplantation of Left Kidney, Allogeneic, Open Approach |
|  |  |  | procedure | UMLS:ICD10PCS:0TY10Z1 | Transplantation of Left Kidney, Syngeneic, Open Approach |
|  |  |  | procedure | UMLS:ICD10PCS:0TY10Z2 | Transplantation of Left Kidney, Zooplastic, Open Approach |
|  |  |  | procedure | UMLS:ICD10PCS:0TY00Z1 | Transplantation of Right Kidney, Syngeneic, Open Approach |
|  |  |  | procedure | UMLS:ICD10PCS:0TY00Z2 | Transplantation of Right Kidney, Zooplastic, Open Approach |
|  |  |  | procedure | UMLS:ICD10PCS:0TY0 | Urinary System / Transplantation / Kidney, Right |
|  |  |  | procedure | UMLS:ICD10PCS:0TY1 | Urinary System / Transplantation / Kidney, Left |
|  | date constraint | | The terms in this group occurred at any time | | |
|  | event relationship | | Any instance of noHypertension occurred within 1 day and 999 days after the first instance of Kidney transplant | | |
|  | **Group 6B noHypertension** | | | | |
|  | cannot have |  | diagnosis | UMLS:ICD10CM:I10 | Essential (primary) hypertension |

##### **Table S36. Code definitions and selection criteria for SGLT2i non-users – no hypertension.**

| Ungrouped terms | | | | | |
| --- | --- | --- | --- | --- | --- |
|  | must have |  | demographics | Age | Age (at least 18 years (most recent occurrence)) |
|  | cannot have |  | diagnosis | UMLS:ICD10CM:E08-E13 | Diabetes mellitus |
| Group 1 | | | | | |
|  | **Kidney transplant** | | | | |
|  | must have | any of | diagnosis | UMLS:ICD10CM:Z94.0 | Kidney transplant status |
|  |  |  | procedure | UMLS:ICD10PCS:0TY00Z0 | Transplantation of Right Kidney, Allogeneic, Open Approach |
|  |  |  | procedure | UMLS:ICD10PCS:0TY10Z0 | Transplantation of Left Kidney, Allogeneic, Open Approach |
|  |  |  | procedure | UMLS:ICD10PCS:0TY10Z1 | Transplantation of Left Kidney, Syngeneic, Open Approach |
|  |  |  | procedure | UMLS:ICD10PCS:0TY10Z2 | Transplantation of Left Kidney, Zooplastic, Open Approach |
|  |  |  | procedure | UMLS:ICD10PCS:0TY00Z1 | Transplantation of Right Kidney, Syngeneic, Open Approach |
|  |  |  | procedure | UMLS:ICD10PCS:0TY00Z2 | Transplantation of Right Kidney, Zooplastic, Open Approach |
|  |  |  | procedure | UMLS:ICD10PCS:0TY0 | Urinary System / Transplantation / Kidney, Right |
|  |  |  | procedure | UMLS:ICD10PCS:0TY1 | Urinary System / Transplantation / Kidney, Left |
|  | date constraint | | The terms in this group occurred between Jan 1, 2015 and Jan 1, 2022 | | |
| Group 2 | | | | | |
|  | **Group 2A Kidney transplant** | | | | |
|  | must have | any of | diagnosis | UMLS:ICD10CM:Z94.0 | Kidney transplant status |
|  |  |  | procedure | UMLS:ICD10PCS:0TY00Z0 | Transplantation of Right Kidney, Allogeneic, Open Approach |
|  |  |  | procedure | UMLS:ICD10PCS:0TY10Z0 | Transplantation of Left Kidney, Allogeneic, Open Approach |
|  |  |  | procedure | UMLS:ICD10PCS:0TY10Z1 | Transplantation of Left Kidney, Syngeneic, Open Approach |
|  |  |  | procedure | UMLS:ICD10PCS:0TY10Z2 | Transplantation of Left Kidney, Zooplastic, Open Approach |
|  |  |  | procedure | UMLS:ICD10PCS:0TY00Z1 | Transplantation of Right Kidney, Syngeneic, Open Approach |
|  |  |  | procedure | UMLS:ICD10PCS:0TY00Z2 | Transplantation of Right Kidney, Zooplastic, Open Approach |
|  |  |  | procedure | UMLS:ICD10PCS:0TY0 | Urinary System / Transplantation / Kidney, Right |
|  |  |  | procedure | UMLS:ICD10PCS:0TY1 | Urinary System / Transplantation / Kidney, Left |
|  | date constraint | | The terms in this group occurred at any time | | |
|  | event relationship | | Any instance of noDialysis, Death, SGLT2i occurred within 1 day and 999 days after the first instance of Kidney transplant | | |
|  | **Group 2B noDialysis, Death, SGLT2i** | | | | |
|  | cannot have |  | procedure | UMLS:CPT:1012740 | Dialysis Services and Procedures |
|  |  | or | procedure | UMLS:CPT:90945 | Dialysis procedure other than hemodialysis (eg, peritoneal dialysis, hemofiltration, or other continuous renal replacement therapies), with single evaluation by a physician or other qualified health care professional |
|  |  | or | procedure | UMLS:CPT:90947 | Dialysis procedure other than hemodialysis (eg, peritoneal dialysis, hemofiltration, or other continuous renal replacement therapies) requiring repeated evaluations by a physician or other qualified health care professional, with or without substantial revision of dialysis prescription |
|  |  | or | procedure | UMLS:CPT:1012752 | Hemodialysis Procedures |
|  |  | or | procedure | UMLS:SNOMED:302497006 | Hemodialysis |
|  |  | or | procedure | UMLS:CPT:90937 | Hemodialysis procedure requiring repeated evaluation(s) with or without substantial revision of dialysis prescription |
|  |  | or | procedure | UMLS:CPT:90935 | Hemodialysis procedure with single evaluation by a physician or other qualified health care professional |
|  |  | or | demographics | Deceased | Deceased |
|  |  | or | diagnosis | UMLS:ICD10CM:R99 | Ill-defined and unknown cause of mortality |
|  |  | or | medication | NLM:ATC:A10BK | Sodium-glucose co-transporter 2 (SGLT2) inhibitors |
| Group 3 | | | | | |
|  | **Group 3A Kidney transplant** | | | | |
|  | must have | any of | diagnosis | UMLS:ICD10CM:Z94.0 | Kidney transplant status |
|  |  |  | procedure | UMLS:ICD10PCS:0TY00Z0 | Transplantation of Right Kidney, Allogeneic, Open Approach |
|  |  |  | procedure | UMLS:ICD10PCS:0TY10Z0 | Transplantation of Left Kidney, Allogeneic, Open Approach |
|  |  |  | procedure | UMLS:ICD10PCS:0TY10Z1 | Transplantation of Left Kidney, Syngeneic, Open Approach |
|  |  |  | procedure | UMLS:ICD10PCS:0TY10Z2 | Transplantation of Left Kidney, Zooplastic, Open Approach |
|  |  |  | procedure | UMLS:ICD10PCS:0TY00Z1 | Transplantation of Right Kidney, Syngeneic, Open Approach |
|  |  |  | procedure | UMLS:ICD10PCS:0TY00Z2 | Transplantation of Right Kidney, Zooplastic, Open Approach |
|  |  |  | procedure | UMLS:ICD10PCS:0TY0 | Urinary System / Transplantation / Kidney, Right |
|  |  |  | procedure | UMLS:ICD10PCS:0TY1 | Urinary System / Transplantation / Kidney, Left |
|  | date constraint | | The terms in this group occurred at any time | | |
|  | event relationship | | Any instance of SGLT2i occurred within 1000 days and 3006 days after the first instance of Kidney transplant | | |
|  | **Group 3B noSGLT2i** | | | | |
|  | cannot have |  | medication | NLM:ATC:A10BK | Sodium-glucose co-transporter 2 (SGLT2) inhibitors |
| Group 4 | | | | | |
|  | **Group 4A Kidney transplant** | | | | |
|  | must have | any of | diagnosis | UMLS:ICD10CM:Z94.0 | Kidney transplant status |
|  |  |  | procedure | UMLS:ICD10PCS:0TY00Z0 | Transplantation of Right Kidney, Allogeneic, Open Approach |
|  |  |  | procedure | UMLS:ICD10PCS:0TY10Z0 | Transplantation of Left Kidney, Allogeneic, Open Approach |
|  |  |  | procedure | UMLS:ICD10PCS:0TY10Z1 | Transplantation of Left Kidney, Syngeneic, Open Approach |
|  |  |  | procedure | UMLS:ICD10PCS:0TY10Z2 | Transplantation of Left Kidney, Zooplastic, Open Approach |
|  |  |  | procedure | UMLS:ICD10PCS:0TY00Z1 | Transplantation of Right Kidney, Syngeneic, Open Approach |
|  |  |  | procedure | UMLS:ICD10PCS:0TY00Z2 | Transplantation of Right Kidney, Zooplastic, Open Approach |
|  |  |  | procedure | UMLS:ICD10PCS:0TY0 | Urinary System / Transplantation / Kidney, Right |
|  |  |  | procedure | UMLS:ICD10PCS:0TY1 | Urinary System / Transplantation / Kidney, Left |
|  | date constraint | | The terms in this group occurred at any time | | |
|  | event relationship | | Any instance of noSGLT2i occurred at least 1 day before the first instance of Kidney transplant | | |
|  | **Group 4B noSGLT2i** | | | | |
|  | cannot have |  | medication | NLM:ATC:A10BK | Sodium-glucose co-transporter 2 (SGLT2) inhibitors |
| Group 5 | | | | | |
|  | **Group 5A Kidney transplant** | | | | |
|  | must have | any of | diagnosis | UMLS:ICD10CM:Z94.0 | Kidney transplant status |
|  |  |  | procedure | UMLS:ICD10PCS:0TY00Z0 | Transplantation of Right Kidney, Allogeneic, Open Approach |
|  |  |  | procedure | UMLS:ICD10PCS:0TY10Z0 | Transplantation of Left Kidney, Allogeneic, Open Approach |
|  |  |  | procedure | UMLS:ICD10PCS:0TY10Z1 | Transplantation of Left Kidney, Syngeneic, Open Approach |
|  |  |  | procedure | UMLS:ICD10PCS:0TY10Z2 | Transplantation of Left Kidney, Zooplastic, Open Approach |
|  |  |  | procedure | UMLS:ICD10PCS:0TY00Z1 | Transplantation of Right Kidney, Syngeneic, Open Approach |
|  |  |  | procedure | UMLS:ICD10PCS:0TY00Z2 | Transplantation of Right Kidney, Zooplastic, Open Approach |
|  |  |  | procedure | UMLS:ICD10PCS:0TY0 | Urinary System / Transplantation / Kidney, Right |
|  |  |  | procedure | UMLS:ICD10PCS:0TY1 | Urinary System / Transplantation / Kidney, Left |
|  | date constraint | | The terms in this group occurred at any time | | |
|  | event relationship | | Any instance of Visit occurred at least 1181 days after the first instance of Kidney transplant | | |
|  | **Group 5B Visit** | | | | |
|  | must have |  | visit | TNX:Visit | Visit |
| Group 6 | | | | | |
|  | **Group 6A Kidney transplant** | | | | |
|  | must have | any of | diagnosis | UMLS:ICD10CM:Z94.0 | Kidney transplant status |
|  |  |  | procedure | UMLS:ICD10PCS:0TY00Z0 | Transplantation of Right Kidney, Allogeneic, Open Approach |
|  |  |  | procedure | UMLS:ICD10PCS:0TY10Z0 | Transplantation of Left Kidney, Allogeneic, Open Approach |
|  |  |  | procedure | UMLS:ICD10PCS:0TY10Z1 | Transplantation of Left Kidney, Syngeneic, Open Approach |
|  |  |  | procedure | UMLS:ICD10PCS:0TY10Z2 | Transplantation of Left Kidney, Zooplastic, Open Approach |
|  |  |  | procedure | UMLS:ICD10PCS:0TY00Z1 | Transplantation of Right Kidney, Syngeneic, Open Approach |
|  |  |  | procedure | UMLS:ICD10PCS:0TY00Z2 | Transplantation of Right Kidney, Zooplastic, Open Approach |
|  |  |  | procedure | UMLS:ICD10PCS:0TY0 | Urinary System / Transplantation / Kidney, Right |
|  |  |  | procedure | UMLS:ICD10PCS:0TY1 | Urinary System / Transplantation / Kidney, Left |
|  | date constraint | | The terms in this group occurred at any time | | |
|  | event relationship | | Any instance of noHypertension occurred within 1 day and 999 days after the first instance of Kidney transplant | | |
|  | **Group 6B noHypertension** | | | | |
|  | cannot have |  | diagnosis | UMLS:ICD10CM:I10 | Essential (primary) hypertension |

##### **Table S37. Code definitions and selection criteria for SGLT2i users – dyslipidemia.**

| Ungrouped terms | | | | | |
| --- | --- | --- | --- | --- | --- |
|  | must have |  | demographics | Age | Age (at least 18 years (most recent occurrence)) |
|  | cannot have |  | diagnosis | UMLS:ICD10CM:E08-E13 | Diabetes mellitus |
| Group 1 | | | | | |
|  | **Kidney transplant** | | | | |
|  | must have | any of | diagnosis | UMLS:ICD10CM:Z94.0 | Kidney transplant status |
|  |  |  | procedure | UMLS:ICD10PCS:0TY00Z0 | Transplantation of Right Kidney, Allogeneic, Open Approach |
|  |  |  | procedure | UMLS:ICD10PCS:0TY10Z0 | Transplantation of Left Kidney, Allogeneic, Open Approach |
|  |  |  | procedure | UMLS:ICD10PCS:0TY10Z1 | Transplantation of Left Kidney, Syngeneic, Open Approach |
|  |  |  | procedure | UMLS:ICD10PCS:0TY10Z2 | Transplantation of Left Kidney, Zooplastic, Open Approach |
|  |  |  | procedure | UMLS:ICD10PCS:0TY00Z1 | Transplantation of Right Kidney, Syngeneic, Open Approach |
|  |  |  | procedure | UMLS:ICD10PCS:0TY00Z2 | Transplantation of Right Kidney, Zooplastic, Open Approach |
|  |  |  | procedure | UMLS:ICD10PCS:0TY0 | Urinary System / Transplantation / Kidney, Right |
|  |  |  | procedure | UMLS:ICD10PCS:0TY1 | Urinary System / Transplantation / Kidney, Left |
|  | date constraint | | The terms in this group occurred between Jan 1, 2015 and Jan 1, 2022 | | |
| Group 2 | | | | | |
|  | **Group 2A Kidney transplant** | | | | |
|  | must have | any of | diagnosis | UMLS:ICD10CM:Z94.0 | Kidney transplant status |
|  |  |  | procedure | UMLS:ICD10PCS:0TY00Z0 | Transplantation of Right Kidney, Allogeneic, Open Approach |
|  |  |  | procedure | UMLS:ICD10PCS:0TY10Z0 | Transplantation of Left Kidney, Allogeneic, Open Approach |
|  |  |  | procedure | UMLS:ICD10PCS:0TY10Z1 | Transplantation of Left Kidney, Syngeneic, Open Approach |
|  |  |  | procedure | UMLS:ICD10PCS:0TY10Z2 | Transplantation of Left Kidney, Zooplastic, Open Approach |
|  |  |  | procedure | UMLS:ICD10PCS:0TY00Z1 | Transplantation of Right Kidney, Syngeneic, Open Approach |
|  |  |  | procedure | UMLS:ICD10PCS:0TY00Z2 | Transplantation of Right Kidney, Zooplastic, Open Approach |
|  |  |  | procedure | UMLS:ICD10PCS:0TY0 | Urinary System / Transplantation / Kidney, Right |
|  |  |  | procedure | UMLS:ICD10PCS:0TY1 | Urinary System / Transplantation / Kidney, Left |
|  | date constraint | | The terms in this group occurred at any time | | |
|  | event relationship | | Any instance of noDialysis, Death, SGLT2i occurred within 1 day and 999 days after the first instance of Kidney transplant | | |
|  | **Group 2B noDialysis, Death, SGLT2i** | | | | |
|  | cannot have |  | procedure | UMLS:CPT:1012740 | Dialysis Services and Procedures |
|  |  | or | procedure | UMLS:CPT:90945 | Dialysis procedure other than hemodialysis (eg, peritoneal dialysis, hemofiltration, or other continuous renal replacement therapies), with single evaluation by a physician or other qualified health care professional |
|  |  | or | procedure | UMLS:CPT:90947 | Dialysis procedure other than hemodialysis (eg, peritoneal dialysis, hemofiltration, or other continuous renal replacement therapies) requiring repeated evaluations by a physician or other qualified health care professional, with or without substantial revision of dialysis prescription |
|  |  | or | procedure | UMLS:CPT:1012752 | Hemodialysis Procedures |
|  |  | or | procedure | UMLS:SNOMED:302497006 | Hemodialysis |
|  |  | or | procedure | UMLS:CPT:90937 | Hemodialysis procedure requiring repeated evaluation(s) with or without substantial revision of dialysis prescription |
|  |  | or | procedure | UMLS:CPT:90935 | Hemodialysis procedure with single evaluation by a physician or other qualified health care professional |
|  |  | or | demographics | Deceased | Deceased |
|  |  | or | diagnosis | UMLS:ICD10CM:R99 | Ill-defined and unknown cause of mortality |
|  |  | or | medication | NLM:ATC:A10BK | Sodium-glucose co-transporter 2 (SGLT2) inhibitors |
| Group 3 | | | | | |
|  | **Group 3A Kidney transplant** | | | | |
|  | must have | any of | diagnosis | UMLS:ICD10CM:Z94.0 | Kidney transplant status |
|  |  |  | procedure | UMLS:ICD10PCS:0TY00Z0 | Transplantation of Right Kidney, Allogeneic, Open Approach |
|  |  |  | procedure | UMLS:ICD10PCS:0TY10Z0 | Transplantation of Left Kidney, Allogeneic, Open Approach |
|  |  |  | procedure | UMLS:ICD10PCS:0TY10Z1 | Transplantation of Left Kidney, Syngeneic, Open Approach |
|  |  |  | procedure | UMLS:ICD10PCS:0TY10Z2 | Transplantation of Left Kidney, Zooplastic, Open Approach |
|  |  |  | procedure | UMLS:ICD10PCS:0TY00Z1 | Transplantation of Right Kidney, Syngeneic, Open Approach |
|  |  |  | procedure | UMLS:ICD10PCS:0TY00Z2 | Transplantation of Right Kidney, Zooplastic, Open Approach |
|  |  |  | procedure | UMLS:ICD10PCS:0TY0 | Urinary System / Transplantation / Kidney, Right |
|  |  |  | procedure | UMLS:ICD10PCS:0TY1 | Urinary System / Transplantation / Kidney, Left |
|  | date constraint | | The terms in this group occurred at any time | | |
|  | event relationship | | Any instance of SGLT2i occurred at least 1000 days after the first instance of Kidney transplant | | |
|  | **Group 3B SGLT2i** | | | | |
|  | must have |  | medication | NLM:ATC:A10BK | Sodium-glucose co-transporter 2 (SGLT2) inhibitors |
| Group 4 | | | | | |
|  | **Group 4A Kidney transplant** | | | | |
|  | must have | any of | diagnosis | UMLS:ICD10CM:Z94.0 | Kidney transplant status |
|  |  |  | procedure | UMLS:ICD10PCS:0TY00Z0 | Transplantation of Right Kidney, Allogeneic, Open Approach |
|  |  |  | procedure | UMLS:ICD10PCS:0TY10Z0 | Transplantation of Left Kidney, Allogeneic, Open Approach |
|  |  |  | procedure | UMLS:ICD10PCS:0TY10Z1 | Transplantation of Left Kidney, Syngeneic, Open Approach |
|  |  |  | procedure | UMLS:ICD10PCS:0TY10Z2 | Transplantation of Left Kidney, Zooplastic, Open Approach |
|  |  |  | procedure | UMLS:ICD10PCS:0TY00Z1 | Transplantation of Right Kidney, Syngeneic, Open Approach |
|  |  |  | procedure | UMLS:ICD10PCS:0TY00Z2 | Transplantation of Right Kidney, Zooplastic, Open Approach |
|  |  |  | procedure | UMLS:ICD10PCS:0TY0 | Urinary System / Transplantation / Kidney, Right |
|  |  |  | procedure | UMLS:ICD10PCS:0TY1 | Urinary System / Transplantation / Kidney, Left |
|  | date constraint | | The terms in this group occurred at any time | | |
|  | event relationship | | Any instance of noSGLT2i occurred at least 1 day before the first instance of Kidney transplant | | |
|  | **Group 4B noSGLT2i** | | | | |
|  | cannot have |  | medication | NLM:ATC:A10BK | Sodium-glucose co-transporter 2 (SGLT2) inhibitors |
| Group 5 | | | | | |
|  | **Group 5A Kidney transplant** | | | | |
|  | must have | any of | diagnosis | UMLS:ICD10CM:Z94.0 | Kidney transplant status |
|  |  |  | procedure | UMLS:ICD10PCS:0TY00Z0 | Transplantation of Right Kidney, Allogeneic, Open Approach |
|  |  |  | procedure | UMLS:ICD10PCS:0TY10Z0 | Transplantation of Left Kidney, Allogeneic, Open Approach |
|  |  |  | procedure | UMLS:ICD10PCS:0TY10Z1 | Transplantation of Left Kidney, Syngeneic, Open Approach |
|  |  |  | procedure | UMLS:ICD10PCS:0TY10Z2 | Transplantation of Left Kidney, Zooplastic, Open Approach |
|  |  |  | procedure | UMLS:ICD10PCS:0TY00Z1 | Transplantation of Right Kidney, Syngeneic, Open Approach |
|  |  |  | procedure | UMLS:ICD10PCS:0TY00Z2 | Transplantation of Right Kidney, Zooplastic, Open Approach |
|  |  |  | procedure | UMLS:ICD10PCS:0TY0 | Urinary System / Transplantation / Kidney, Right |
|  |  |  | procedure | UMLS:ICD10PCS:0TY1 | Urinary System / Transplantation / Kidney, Left |
|  | date constraint | | The terms in this group occurred at any time | | |
|  | event relationship | | Any instance of Visit occurred at least 1181 days after the first instance of Kidney transplant | | |
|  | **Group 5B Visit** | | | | |
|  | must have |  | visit | TNX:Visit | Visit |
| Group 6 | | | | | |
|  | **Group 6A Kidney transplant** | | | | |
|  | must have | any of | diagnosis | UMLS:ICD10CM:Z94.0 | Kidney transplant status |
|  |  |  | procedure | UMLS:ICD10PCS:0TY00Z0 | Transplantation of Right Kidney, Allogeneic, Open Approach |
|  |  |  | procedure | UMLS:ICD10PCS:0TY10Z0 | Transplantation of Left Kidney, Allogeneic, Open Approach |
|  |  |  | procedure | UMLS:ICD10PCS:0TY10Z1 | Transplantation of Left Kidney, Syngeneic, Open Approach |
|  |  |  | procedure | UMLS:ICD10PCS:0TY10Z2 | Transplantation of Left Kidney, Zooplastic, Open Approach |
|  |  |  | procedure | UMLS:ICD10PCS:0TY00Z1 | Transplantation of Right Kidney, Syngeneic, Open Approach |
|  |  |  | procedure | UMLS:ICD10PCS:0TY00Z2 | Transplantation of Right Kidney, Zooplastic, Open Approach |
|  |  |  | procedure | UMLS:ICD10PCS:0TY0 | Urinary System / Transplantation / Kidney, Right |
|  |  |  | procedure | UMLS:ICD10PCS:0TY1 | Urinary System / Transplantation / Kidney, Left |
|  | date constraint | | The terms in this group occurred at any time | | |
|  | event relationship | | Any instance of Dyslipidemia occurred within 1 day and 999 days after the first instance of Kidney transplant | | |
|  | **Group 6B Dyslipidemia** | | | | |
|  | must have |  | diagnosis | UMLS:ICD10CM:E78 | Disorders of lipoprotein metabolism and other lipidemias |

##### **Table S38. Code definitions and selection criteria for SGLT2i non-users – dyslipidemia.**

| Ungrouped terms | | | | | |
| --- | --- | --- | --- | --- | --- |
|  | must have |  | demographics | Age | Age (at least 18 years (most recent occurrence)) |
|  | cannot have |  | diagnosis | UMLS:ICD10CM:E08-E13 | Diabetes mellitus |
| Group 1 | | | | | |
|  | **Kidney transplant** | | | | |
|  | must have | any of | diagnosis | UMLS:ICD10CM:Z94.0 | Kidney transplant status |
|  |  |  | procedure | UMLS:ICD10PCS:0TY00Z0 | Transplantation of Right Kidney, Allogeneic, Open Approach |
|  |  |  | procedure | UMLS:ICD10PCS:0TY10Z0 | Transplantation of Left Kidney, Allogeneic, Open Approach |
|  |  |  | procedure | UMLS:ICD10PCS:0TY10Z1 | Transplantation of Left Kidney, Syngeneic, Open Approach |
|  |  |  | procedure | UMLS:ICD10PCS:0TY10Z2 | Transplantation of Left Kidney, Zooplastic, Open Approach |
|  |  |  | procedure | UMLS:ICD10PCS:0TY00Z1 | Transplantation of Right Kidney, Syngeneic, Open Approach |
|  |  |  | procedure | UMLS:ICD10PCS:0TY00Z2 | Transplantation of Right Kidney, Zooplastic, Open Approach |
|  |  |  | procedure | UMLS:ICD10PCS:0TY0 | Urinary System / Transplantation / Kidney, Right |
|  |  |  | procedure | UMLS:ICD10PCS:0TY1 | Urinary System / Transplantation / Kidney, Left |
|  | date constraint | | The terms in this group occurred between Jan 1, 2015 and Jan 1, 2022 | | |
| Group 2 | | | | | |
|  | **Group 2A Kidney transplant** | | | | |
|  | must have | any of | diagnosis | UMLS:ICD10CM:Z94.0 | Kidney transplant status |
|  |  |  | procedure | UMLS:ICD10PCS:0TY00Z0 | Transplantation of Right Kidney, Allogeneic, Open Approach |
|  |  |  | procedure | UMLS:ICD10PCS:0TY10Z0 | Transplantation of Left Kidney, Allogeneic, Open Approach |
|  |  |  | procedure | UMLS:ICD10PCS:0TY10Z1 | Transplantation of Left Kidney, Syngeneic, Open Approach |
|  |  |  | procedure | UMLS:ICD10PCS:0TY10Z2 | Transplantation of Left Kidney, Zooplastic, Open Approach |
|  |  |  | procedure | UMLS:ICD10PCS:0TY00Z1 | Transplantation of Right Kidney, Syngeneic, Open Approach |
|  |  |  | procedure | UMLS:ICD10PCS:0TY00Z2 | Transplantation of Right Kidney, Zooplastic, Open Approach |
|  |  |  | procedure | UMLS:ICD10PCS:0TY0 | Urinary System / Transplantation / Kidney, Right |
|  |  |  | procedure | UMLS:ICD10PCS:0TY1 | Urinary System / Transplantation / Kidney, Left |
|  | date constraint | | The terms in this group occurred at any time | | |
|  | event relationship | | Any instance of noDialysis, Death, SGLT2i occurred within 1 day and 999 days after the first instance of Kidney transplant | | |
|  | **Group 2B noDialysis, Death, SGLT2i** | | | | |
|  | cannot have |  | procedure | UMLS:CPT:1012740 | Dialysis Services and Procedures |
|  |  | or | procedure | UMLS:CPT:90945 | Dialysis procedure other than hemodialysis (eg, peritoneal dialysis, hemofiltration, or other continuous renal replacement therapies), with single evaluation by a physician or other qualified health care professional |
|  |  | or | procedure | UMLS:CPT:90947 | Dialysis procedure other than hemodialysis (eg, peritoneal dialysis, hemofiltration, or other continuous renal replacement therapies) requiring repeated evaluations by a physician or other qualified health care professional, with or without substantial revision of dialysis prescription |
|  |  | or | procedure | UMLS:CPT:1012752 | Hemodialysis Procedures |
|  |  | or | procedure | UMLS:SNOMED:302497006 | Hemodialysis |
|  |  | or | procedure | UMLS:CPT:90937 | Hemodialysis procedure requiring repeated evaluation(s) with or without substantial revision of dialysis prescription |
|  |  | or | procedure | UMLS:CPT:90935 | Hemodialysis procedure with single evaluation by a physician or other qualified health care professional |
|  |  | or | demographics | Deceased | Deceased |
|  |  | or | diagnosis | UMLS:ICD10CM:R99 | Ill-defined and unknown cause of mortality |
|  |  | or | medication | NLM:ATC:A10BK | Sodium-glucose co-transporter 2 (SGLT2) inhibitors |
| Group 3 | | | | | |
|  | **Group 3A Kidney transplant** | | | | |
|  | must have | any of | diagnosis | UMLS:ICD10CM:Z94.0 | Kidney transplant status |
|  |  |  | procedure | UMLS:ICD10PCS:0TY00Z0 | Transplantation of Right Kidney, Allogeneic, Open Approach |
|  |  |  | procedure | UMLS:ICD10PCS:0TY10Z0 | Transplantation of Left Kidney, Allogeneic, Open Approach |
|  |  |  | procedure | UMLS:ICD10PCS:0TY10Z1 | Transplantation of Left Kidney, Syngeneic, Open Approach |
|  |  |  | procedure | UMLS:ICD10PCS:0TY10Z2 | Transplantation of Left Kidney, Zooplastic, Open Approach |
|  |  |  | procedure | UMLS:ICD10PCS:0TY00Z1 | Transplantation of Right Kidney, Syngeneic, Open Approach |
|  |  |  | procedure | UMLS:ICD10PCS:0TY00Z2 | Transplantation of Right Kidney, Zooplastic, Open Approach |
|  |  |  | procedure | UMLS:ICD10PCS:0TY0 | Urinary System / Transplantation / Kidney, Right |
|  |  |  | procedure | UMLS:ICD10PCS:0TY1 | Urinary System / Transplantation / Kidney, Left |
|  | date constraint | | The terms in this group occurred at any time | | |
|  | event relationship | | Any instance of SGLT2i occurred within 1000 days and 3006 days after the first instance of Kidney transplant | | |
|  | **Group 3B SGLT2i** | | | | |
|  | cannot have |  | medication | NLM:ATC:A10BK | Sodium-glucose co-transporter 2 (SGLT2) inhibitors |
| Group 4 | | | | | |
|  | **Group 4A Kidney transplant** | | | | |
|  | must have | any of | diagnosis | UMLS:ICD10CM:Z94.0 | Kidney transplant status |
|  |  |  | procedure | UMLS:ICD10PCS:0TY00Z0 | Transplantation of Right Kidney, Allogeneic, Open Approach |
|  |  |  | procedure | UMLS:ICD10PCS:0TY10Z0 | Transplantation of Left Kidney, Allogeneic, Open Approach |
|  |  |  | procedure | UMLS:ICD10PCS:0TY10Z1 | Transplantation of Left Kidney, Syngeneic, Open Approach |
|  |  |  | procedure | UMLS:ICD10PCS:0TY10Z2 | Transplantation of Left Kidney, Zooplastic, Open Approach |
|  |  |  | procedure | UMLS:ICD10PCS:0TY00Z1 | Transplantation of Right Kidney, Syngeneic, Open Approach |
|  |  |  | procedure | UMLS:ICD10PCS:0TY00Z2 | Transplantation of Right Kidney, Zooplastic, Open Approach |
|  |  |  | procedure | UMLS:ICD10PCS:0TY0 | Urinary System / Transplantation / Kidney, Right |
|  |  |  | procedure | UMLS:ICD10PCS:0TY1 | Urinary System / Transplantation / Kidney, Left |
|  | date constraint | | The terms in this group occurred at any time | | |
|  | event relationship | | Any instance of noSGLT2i occurred at least 1 day before the first instance of Kidney transplant | | |
|  | **Group 4B noSGLT2i** | | | | |
|  | cannot have |  | medication | NLM:ATC:A10BK | Sodium-glucose co-transporter 2 (SGLT2) inhibitors |
| Group 5 | | | | | |
|  | **Group 5A Kidney transplant** | | | | |
|  | must have | any of | diagnosis | UMLS:ICD10CM:Z94.0 | Kidney transplant status |
|  |  |  | procedure | UMLS:ICD10PCS:0TY00Z0 | Transplantation of Right Kidney, Allogeneic, Open Approach |
|  |  |  | procedure | UMLS:ICD10PCS:0TY10Z0 | Transplantation of Left Kidney, Allogeneic, Open Approach |
|  |  |  | procedure | UMLS:ICD10PCS:0TY10Z1 | Transplantation of Left Kidney, Syngeneic, Open Approach |
|  |  |  | procedure | UMLS:ICD10PCS:0TY10Z2 | Transplantation of Left Kidney, Zooplastic, Open Approach |
|  |  |  | procedure | UMLS:ICD10PCS:0TY00Z1 | Transplantation of Right Kidney, Syngeneic, Open Approach |
|  |  |  | procedure | UMLS:ICD10PCS:0TY00Z2 | Transplantation of Right Kidney, Zooplastic, Open Approach |
|  |  |  | procedure | UMLS:ICD10PCS:0TY0 | Urinary System / Transplantation / Kidney, Right |
|  |  |  | procedure | UMLS:ICD10PCS:0TY1 | Urinary System / Transplantation / Kidney, Left |
|  | date constraint | | The terms in this group occurred at any time | | |
|  | event relationship | | Any instance of Visit occurred at least 1181 days after the first instance of Kidney transplant | | |
|  | **Group 5B Visit** | | | | |
|  | must have |  | visit | TNX:Visit | Visit |
| Group 6 | | | | | |
|  | **Group 6A Kidney transplant** | | | | |
|  | must have | any of | diagnosis | UMLS:ICD10CM:Z94.0 | Kidney transplant status |
|  |  |  | procedure | UMLS:ICD10PCS:0TY00Z0 | Transplantation of Right Kidney, Allogeneic, Open Approach |
|  |  |  | procedure | UMLS:ICD10PCS:0TY10Z0 | Transplantation of Left Kidney, Allogeneic, Open Approach |
|  |  |  | procedure | UMLS:ICD10PCS:0TY10Z1 | Transplantation of Left Kidney, Syngeneic, Open Approach |
|  |  |  | procedure | UMLS:ICD10PCS:0TY10Z2 | Transplantation of Left Kidney, Zooplastic, Open Approach |
|  |  |  | procedure | UMLS:ICD10PCS:0TY00Z1 | Transplantation of Right Kidney, Syngeneic, Open Approach |
|  |  |  | procedure | UMLS:ICD10PCS:0TY00Z2 | Transplantation of Right Kidney, Zooplastic, Open Approach |
|  |  |  | procedure | UMLS:ICD10PCS:0TY0 | Urinary System / Transplantation / Kidney, Right |
|  |  |  | procedure | UMLS:ICD10PCS:0TY1 | Urinary System / Transplantation / Kidney, Left |
|  | date constraint | | The terms in this group occurred at any time | | |
|  | event relationship | | Any instance of Dyslipidemia occurred within 1 day and 999 days after the first instance of Kidney transplant | | |
|  | **Group 6B Dyslipidemia** | | | | |
|  | must have |  | diagnosis | UMLS:ICD10CM:E78 | Disorders of lipoprotein metabolism and other lipidemias |

##### **Table S39. Code definitions and selection criteria for SGLT2i users – no dyslipidemia.**

| Ungrouped terms | | | | | |
| --- | --- | --- | --- | --- | --- |
|  | must have |  | demographics | Age | Age (at least 18 years (most recent occurrence)) |
|  | cannot have |  | diagnosis | UMLS:ICD10CM:E08-E13 | Diabetes mellitus |
| Group 1 | | | | | |
|  | **Kidney transplant** | | | | |
|  | must have | any of | diagnosis | UMLS:ICD10CM:Z94.0 | Kidney transplant status |
|  |  |  | procedure | UMLS:ICD10PCS:0TY00Z0 | Transplantation of Right Kidney, Allogeneic, Open Approach |
|  |  |  | procedure | UMLS:ICD10PCS:0TY10Z0 | Transplantation of Left Kidney, Allogeneic, Open Approach |
|  |  |  | procedure | UMLS:ICD10PCS:0TY10Z1 | Transplantation of Left Kidney, Syngeneic, Open Approach |
|  |  |  | procedure | UMLS:ICD10PCS:0TY10Z2 | Transplantation of Left Kidney, Zooplastic, Open Approach |
|  |  |  | procedure | UMLS:ICD10PCS:0TY00Z1 | Transplantation of Right Kidney, Syngeneic, Open Approach |
|  |  |  | procedure | UMLS:ICD10PCS:0TY00Z2 | Transplantation of Right Kidney, Zooplastic, Open Approach |
|  |  |  | procedure | UMLS:ICD10PCS:0TY0 | Urinary System / Transplantation / Kidney, Right |
|  |  |  | procedure | UMLS:ICD10PCS:0TY1 | Urinary System / Transplantation / Kidney, Left |
|  | date constraint | | The terms in this group occurred between Jan 1, 2015 and Jan 1, 2022 | | |
| Group 2 | | | | | |
|  | **Group 2A Kidney transplant** | | | | |
|  | must have | any of | diagnosis | UMLS:ICD10CM:Z94.0 | Kidney transplant status |
|  |  |  | procedure | UMLS:ICD10PCS:0TY00Z0 | Transplantation of Right Kidney, Allogeneic, Open Approach |
|  |  |  | procedure | UMLS:ICD10PCS:0TY10Z0 | Transplantation of Left Kidney, Allogeneic, Open Approach |
|  |  |  | procedure | UMLS:ICD10PCS:0TY10Z1 | Transplantation of Left Kidney, Syngeneic, Open Approach |
|  |  |  | procedure | UMLS:ICD10PCS:0TY10Z2 | Transplantation of Left Kidney, Zooplastic, Open Approach |
|  |  |  | procedure | UMLS:ICD10PCS:0TY00Z1 | Transplantation of Right Kidney, Syngeneic, Open Approach |
|  |  |  | procedure | UMLS:ICD10PCS:0TY00Z2 | Transplantation of Right Kidney, Zooplastic, Open Approach |
|  |  |  | procedure | UMLS:ICD10PCS:0TY0 | Urinary System / Transplantation / Kidney, Right |
|  |  |  | procedure | UMLS:ICD10PCS:0TY1 | Urinary System / Transplantation / Kidney, Left |
|  | date constraint | | The terms in this group occurred at any time | | |
|  | event relationship | | Any instance of noDialysis, Death, SGLT2i occurred within 1 day and 999 days after the first instance of Kidney transplant | | |
|  | **Group 2B noDialysis, Death, SGLT2i** | | | | |
|  | cannot have |  | procedure | UMLS:CPT:1012740 | Dialysis Services and Procedures |
|  |  | or | procedure | UMLS:CPT:90945 | Dialysis procedure other than hemodialysis (eg, peritoneal dialysis, hemofiltration, or other continuous renal replacement therapies), with single evaluation by a physician or other qualified health care professional |
|  |  | or | procedure | UMLS:CPT:90947 | Dialysis procedure other than hemodialysis (eg, peritoneal dialysis, hemofiltration, or other continuous renal replacement therapies) requiring repeated evaluations by a physician or other qualified health care professional, with or without substantial revision of dialysis prescription |
|  |  | or | procedure | UMLS:CPT:1012752 | Hemodialysis Procedures |
|  |  | or | procedure | UMLS:SNOMED:302497006 | Hemodialysis |
|  |  | or | procedure | UMLS:CPT:90937 | Hemodialysis procedure requiring repeated evaluation(s) with or without substantial revision of dialysis prescription |
|  |  | or | procedure | UMLS:CPT:90935 | Hemodialysis procedure with single evaluation by a physician or other qualified health care professional |
|  |  | or | demographics | Deceased | Deceased |
|  |  | or | diagnosis | UMLS:ICD10CM:R99 | Ill-defined and unknown cause of mortality |
|  |  | or | medication | NLM:ATC:A10BK | Sodium-glucose co-transporter 2 (SGLT2) inhibitors |
| Group 3 | | | | | |
|  | **Group 3A Kidney transplant** | | | | |
|  | must have | any of | diagnosis | UMLS:ICD10CM:Z94.0 | Kidney transplant status |
|  |  |  | procedure | UMLS:ICD10PCS:0TY00Z0 | Transplantation of Right Kidney, Allogeneic, Open Approach |
|  |  |  | procedure | UMLS:ICD10PCS:0TY10Z0 | Transplantation of Left Kidney, Allogeneic, Open Approach |
|  |  |  | procedure | UMLS:ICD10PCS:0TY10Z1 | Transplantation of Left Kidney, Syngeneic, Open Approach |
|  |  |  | procedure | UMLS:ICD10PCS:0TY10Z2 | Transplantation of Left Kidney, Zooplastic, Open Approach |
|  |  |  | procedure | UMLS:ICD10PCS:0TY00Z1 | Transplantation of Right Kidney, Syngeneic, Open Approach |
|  |  |  | procedure | UMLS:ICD10PCS:0TY00Z2 | Transplantation of Right Kidney, Zooplastic, Open Approach |
|  |  |  | procedure | UMLS:ICD10PCS:0TY0 | Urinary System / Transplantation / Kidney, Right |
|  |  |  | procedure | UMLS:ICD10PCS:0TY1 | Urinary System / Transplantation / Kidney, Left |
|  | date constraint | | The terms in this group occurred at any time | | |
|  | event relationship | | Any instance of SGLT2i occurred at least 1000 days after the first instance of Kidney transplant | | |
|  | **Group 3B SGLT2i** | | | | |
|  | must have |  | medication | NLM:ATC:A10BK | Sodium-glucose co-transporter 2 (SGLT2) inhibitors |
| Group 4 | | | | | |
|  | **Group 4A Kidney transplant** | | | | |
|  | must have | any of | diagnosis | UMLS:ICD10CM:Z94.0 | Kidney transplant status |
|  |  |  | procedure | UMLS:ICD10PCS:0TY00Z0 | Transplantation of Right Kidney, Allogeneic, Open Approach |
|  |  |  | procedure | UMLS:ICD10PCS:0TY10Z0 | Transplantation of Left Kidney, Allogeneic, Open Approach |
|  |  |  | procedure | UMLS:ICD10PCS:0TY10Z1 | Transplantation of Left Kidney, Syngeneic, Open Approach |
|  |  |  | procedure | UMLS:ICD10PCS:0TY10Z2 | Transplantation of Left Kidney, Zooplastic, Open Approach |
|  |  |  | procedure | UMLS:ICD10PCS:0TY00Z1 | Transplantation of Right Kidney, Syngeneic, Open Approach |
|  |  |  | procedure | UMLS:ICD10PCS:0TY00Z2 | Transplantation of Right Kidney, Zooplastic, Open Approach |
|  |  |  | procedure | UMLS:ICD10PCS:0TY0 | Urinary System / Transplantation / Kidney, Right |
|  |  |  | procedure | UMLS:ICD10PCS:0TY1 | Urinary System / Transplantation / Kidney, Left |
|  | date constraint | | The terms in this group occurred at any time | | |
|  | event relationship | | Any instance of noSGLT2i occurred at least 1 day before the first instance of Kidney transplant | | |
|  | **Group 4B noSGLT2i** | | | | |
|  | cannot have |  | medication | NLM:ATC:A10BK | Sodium-glucose co-transporter 2 (SGLT2) inhibitors |
| Group 5 | | | | | |
|  | **Group 5A Kidney transplant** | | | | |
|  | must have | any of | diagnosis | UMLS:ICD10CM:Z94.0 | Kidney transplant status |
|  |  |  | procedure | UMLS:ICD10PCS:0TY00Z0 | Transplantation of Right Kidney, Allogeneic, Open Approach |
|  |  |  | procedure | UMLS:ICD10PCS:0TY10Z0 | Transplantation of Left Kidney, Allogeneic, Open Approach |
|  |  |  | procedure | UMLS:ICD10PCS:0TY10Z1 | Transplantation of Left Kidney, Syngeneic, Open Approach |
|  |  |  | procedure | UMLS:ICD10PCS:0TY10Z2 | Transplantation of Left Kidney, Zooplastic, Open Approach |
|  |  |  | procedure | UMLS:ICD10PCS:0TY00Z1 | Transplantation of Right Kidney, Syngeneic, Open Approach |
|  |  |  | procedure | UMLS:ICD10PCS:0TY00Z2 | Transplantation of Right Kidney, Zooplastic, Open Approach |
|  |  |  | procedure | UMLS:ICD10PCS:0TY0 | Urinary System / Transplantation / Kidney, Right |
|  |  |  | procedure | UMLS:ICD10PCS:0TY1 | Urinary System / Transplantation / Kidney, Left |
|  | date constraint | | The terms in this group occurred at any time | | |
|  | event relationship | | Any instance of Visit occurred at least 1181 days after the first instance of Kidney transplant | | |
|  | **Group 5B Visit** | | | | |
|  | must have |  | visit | TNX:Visit | Visit |
| Group 6 | | | | | |
|  | **Group 6A Kidney transplant** | | | | |
|  | must have | any of | diagnosis | UMLS:ICD10CM:Z94.0 | Kidney transplant status |
|  |  |  | procedure | UMLS:ICD10PCS:0TY00Z0 | Transplantation of Right Kidney, Allogeneic, Open Approach |
|  |  |  | procedure | UMLS:ICD10PCS:0TY10Z0 | Transplantation of Left Kidney, Allogeneic, Open Approach |
|  |  |  | procedure | UMLS:ICD10PCS:0TY10Z1 | Transplantation of Left Kidney, Syngeneic, Open Approach |
|  |  |  | procedure | UMLS:ICD10PCS:0TY10Z2 | Transplantation of Left Kidney, Zooplastic, Open Approach |
|  |  |  | procedure | UMLS:ICD10PCS:0TY00Z1 | Transplantation of Right Kidney, Syngeneic, Open Approach |
|  |  |  | procedure | UMLS:ICD10PCS:0TY00Z2 | Transplantation of Right Kidney, Zooplastic, Open Approach |
|  |  |  | procedure | UMLS:ICD10PCS:0TY0 | Urinary System / Transplantation / Kidney, Right |
|  |  |  | procedure | UMLS:ICD10PCS:0TY1 | Urinary System / Transplantation / Kidney, Left |
|  | date constraint | | The terms in this group occurred at any time | | |
|  | event relationship | | Any instance of noDyslipidemia occurred within 1 day and 999 days after the first instance of Kidney transplant | | |
|  | **Group 6B noDyslipidemia** | | | | |
|  | cannot have |  | diagnosis | UMLS:ICD10CM:E78 | Disorders of lipoprotein metabolism and other lipidemias |

##### **Table S40. Code definitions and selection criteria for SGLT2i non-users – no dyslipidemia.**

| Ungrouped terms | | | | | |
| --- | --- | --- | --- | --- | --- |
|  | must have |  | demographics | Age | Age (at least 18 years (most recent occurrence)) |
|  | cannot have |  | diagnosis | UMLS:ICD10CM:E08-E13 | Diabetes mellitus |
| Group 1 | | | | | |
|  | **Kidney transplant** | | | | |
|  | must have | any of | diagnosis | UMLS:ICD10CM:Z94.0 | Kidney transplant status |
|  |  |  | procedure | UMLS:ICD10PCS:0TY00Z0 | Transplantation of Right Kidney, Allogeneic, Open Approach |
|  |  |  | procedure | UMLS:ICD10PCS:0TY10Z0 | Transplantation of Left Kidney, Allogeneic, Open Approach |
|  |  |  | procedure | UMLS:ICD10PCS:0TY10Z1 | Transplantation of Left Kidney, Syngeneic, Open Approach |
|  |  |  | procedure | UMLS:ICD10PCS:0TY10Z2 | Transplantation of Left Kidney, Zooplastic, Open Approach |
|  |  |  | procedure | UMLS:ICD10PCS:0TY00Z1 | Transplantation of Right Kidney, Syngeneic, Open Approach |
|  |  |  | procedure | UMLS:ICD10PCS:0TY00Z2 | Transplantation of Right Kidney, Zooplastic, Open Approach |
|  |  |  | procedure | UMLS:ICD10PCS:0TY0 | Urinary System / Transplantation / Kidney, Right |
|  |  |  | procedure | UMLS:ICD10PCS:0TY1 | Urinary System / Transplantation / Kidney, Left |
|  | date constraint | | The terms in this group occurred between Jan 1, 2015 and Jan 1, 2022 | | |
| Group 2 | | | | | |
|  | **Group 2A Kidney transplant** | | | | |
|  | must have | any of | diagnosis | UMLS:ICD10CM:Z94.0 | Kidney transplant status |
|  |  |  | procedure | UMLS:ICD10PCS:0TY00Z0 | Transplantation of Right Kidney, Allogeneic, Open Approach |
|  |  |  | procedure | UMLS:ICD10PCS:0TY10Z0 | Transplantation of Left Kidney, Allogeneic, Open Approach |
|  |  |  | procedure | UMLS:ICD10PCS:0TY10Z1 | Transplantation of Left Kidney, Syngeneic, Open Approach |
|  |  |  | procedure | UMLS:ICD10PCS:0TY10Z2 | Transplantation of Left Kidney, Zooplastic, Open Approach |
|  |  |  | procedure | UMLS:ICD10PCS:0TY00Z1 | Transplantation of Right Kidney, Syngeneic, Open Approach |
|  |  |  | procedure | UMLS:ICD10PCS:0TY00Z2 | Transplantation of Right Kidney, Zooplastic, Open Approach |
|  |  |  | procedure | UMLS:ICD10PCS:0TY0 | Urinary System / Transplantation / Kidney, Right |
|  |  |  | procedure | UMLS:ICD10PCS:0TY1 | Urinary System / Transplantation / Kidney, Left |
|  | date constraint | | The terms in this group occurred at any time | | |
|  | event relationship | | Any instance of noDialysis, Death, SGLT2i occurred within 1 day and 999 days after the first instance of Kidney transplant | | |
|  | **Group 2B noDialysis, Death, SGLT2i** | | | | |
|  | cannot have |  | procedure | UMLS:CPT:1012740 | Dialysis Services and Procedures |
|  |  | or | procedure | UMLS:CPT:90945 | Dialysis procedure other than hemodialysis (eg, peritoneal dialysis, hemofiltration, or other continuous renal replacement therapies), with single evaluation by a physician or other qualified health care professional |
|  |  | or | procedure | UMLS:CPT:90947 | Dialysis procedure other than hemodialysis (eg, peritoneal dialysis, hemofiltration, or other continuous renal replacement therapies) requiring repeated evaluations by a physician or other qualified health care professional, with or without substantial revision of dialysis prescription |
|  |  | or | procedure | UMLS:CPT:1012752 | Hemodialysis Procedures |
|  |  | or | procedure | UMLS:SNOMED:302497006 | Hemodialysis |
|  |  | or | procedure | UMLS:CPT:90937 | Hemodialysis procedure requiring repeated evaluation(s) with or without substantial revision of dialysis prescription |
|  |  | or | procedure | UMLS:CPT:90935 | Hemodialysis procedure with single evaluation by a physician or other qualified health care professional |
|  |  | or | demographics | Deceased | Deceased |
|  |  | or | diagnosis | UMLS:ICD10CM:R99 | Ill-defined and unknown cause of mortality |
|  |  | or | medication | NLM:ATC:A10BK | Sodium-glucose co-transporter 2 (SGLT2) inhibitors |
| Group 3 | | | | | |
|  | **Group 3A Kidney transplant** | | | | |
|  | must have | any of | diagnosis | UMLS:ICD10CM:Z94.0 | Kidney transplant status |
|  |  |  | procedure | UMLS:ICD10PCS:0TY00Z0 | Transplantation of Right Kidney, Allogeneic, Open Approach |
|  |  |  | procedure | UMLS:ICD10PCS:0TY10Z0 | Transplantation of Left Kidney, Allogeneic, Open Approach |
|  |  |  | procedure | UMLS:ICD10PCS:0TY10Z1 | Transplantation of Left Kidney, Syngeneic, Open Approach |
|  |  |  | procedure | UMLS:ICD10PCS:0TY10Z2 | Transplantation of Left Kidney, Zooplastic, Open Approach |
|  |  |  | procedure | UMLS:ICD10PCS:0TY00Z1 | Transplantation of Right Kidney, Syngeneic, Open Approach |
|  |  |  | procedure | UMLS:ICD10PCS:0TY00Z2 | Transplantation of Right Kidney, Zooplastic, Open Approach |
|  |  |  | procedure | UMLS:ICD10PCS:0TY0 | Urinary System / Transplantation / Kidney, Right |
|  |  |  | procedure | UMLS:ICD10PCS:0TY1 | Urinary System / Transplantation / Kidney, Left |
|  | date constraint | | The terms in this group occurred at any time | | |
|  | event relationship | | Any instance of SGLT2i occurred within 1000 days and 3006 days after the first instance of Kidney transplant | | |
|  | **Group 3B SGLT2i** | | | | |
|  | cannot have |  | medication | NLM:ATC:A10BK | Sodium-glucose co-transporter 2 (SGLT2) inhibitors |
| Group 4 | | | | | |
|  | **Group 4A Kidney transplant** | | | | |
|  | must have | any of | diagnosis | UMLS:ICD10CM:Z94.0 | Kidney transplant status |
|  |  |  | procedure | UMLS:ICD10PCS:0TY00Z0 | Transplantation of Right Kidney, Allogeneic, Open Approach |
|  |  |  | procedure | UMLS:ICD10PCS:0TY10Z0 | Transplantation of Left Kidney, Allogeneic, Open Approach |
|  |  |  | procedure | UMLS:ICD10PCS:0TY10Z1 | Transplantation of Left Kidney, Syngeneic, Open Approach |
|  |  |  | procedure | UMLS:ICD10PCS:0TY10Z2 | Transplantation of Left Kidney, Zooplastic, Open Approach |
|  |  |  | procedure | UMLS:ICD10PCS:0TY00Z1 | Transplantation of Right Kidney, Syngeneic, Open Approach |
|  |  |  | procedure | UMLS:ICD10PCS:0TY00Z2 | Transplantation of Right Kidney, Zooplastic, Open Approach |
|  |  |  | procedure | UMLS:ICD10PCS:0TY0 | Urinary System / Transplantation / Kidney, Right |
|  |  |  | procedure | UMLS:ICD10PCS:0TY1 | Urinary System / Transplantation / Kidney, Left |
|  | date constraint | | The terms in this group occurred at any time | | |
|  | event relationship | | Any instance of noSGLT2 occurred at least 1 day before the first instance of Kidney transplant | | |
|  | **Group 4B noSGLT2i** | | | | |
|  | cannot have |  | medication | NLM:ATC:A10BK | Sodium-glucose co-transporter 2 (SGLT2) inhibitors |
| Group 5 | | | | | |
|  | **Group 5A Kidney transplant** | | | | |
|  | must have | any of | diagnosis | UMLS:ICD10CM:Z94.0 | Kidney transplant status |
|  |  |  | procedure | UMLS:ICD10PCS:0TY00Z0 | Transplantation of Right Kidney, Allogeneic, Open Approach |
|  |  |  | procedure | UMLS:ICD10PCS:0TY10Z0 | Transplantation of Left Kidney, Allogeneic, Open Approach |
|  |  |  | procedure | UMLS:ICD10PCS:0TY10Z1 | Transplantation of Left Kidney, Syngeneic, Open Approach |
|  |  |  | procedure | UMLS:ICD10PCS:0TY10Z2 | Transplantation of Left Kidney, Zooplastic, Open Approach |
|  |  |  | procedure | UMLS:ICD10PCS:0TY00Z1 | Transplantation of Right Kidney, Syngeneic, Open Approach |
|  |  |  | procedure | UMLS:ICD10PCS:0TY00Z2 | Transplantation of Right Kidney, Zooplastic, Open Approach |
|  |  |  | procedure | UMLS:ICD10PCS:0TY0 | Urinary System / Transplantation / Kidney, Right |
|  |  |  | procedure | UMLS:ICD10PCS:0TY1 | Urinary System / Transplantation / Kidney, Left |
|  | date constraint | | The terms in this group occurred at any time | | |
|  | event relationship | | Any instance of Visit occurred at least 1181 days after the first instance of Kidney transplant | | |
|  | **Group 5B Visit** | | | | |
|  | must have |  | visit | TNX:Visit | Visit |
| Group 6 | | | | | |
|  | **Group 6A Kidney transplant** | | | | |
|  | must have | any of | diagnosis | UMLS:ICD10CM:Z94.0 | Kidney transplant status |
|  |  |  | procedure | UMLS:ICD10PCS:0TY00Z0 | Transplantation of Right Kidney, Allogeneic, Open Approach |
|  |  |  | procedure | UMLS:ICD10PCS:0TY10Z0 | Transplantation of Left Kidney, Allogeneic, Open Approach |
|  |  |  | procedure | UMLS:ICD10PCS:0TY10Z1 | Transplantation of Left Kidney, Syngeneic, Open Approach |
|  |  |  | procedure | UMLS:ICD10PCS:0TY10Z2 | Transplantation of Left Kidney, Zooplastic, Open Approach |
|  |  |  | procedure | UMLS:ICD10PCS:0TY00Z1 | Transplantation of Right Kidney, Syngeneic, Open Approach |
|  |  |  | procedure | UMLS:ICD10PCS:0TY00Z2 | Transplantation of Right Kidney, Zooplastic, Open Approach |
|  |  |  | procedure | UMLS:ICD10PCS:0TY0 | Urinary System / Transplantation / Kidney, Right |
|  |  |  | procedure | UMLS:ICD10PCS:0TY1 | Urinary System / Transplantation / Kidney, Left |
|  | date constraint | | The terms in this group occurred at any time | | |
|  | event relationship | | Any instance of noDyslipidemia occurred within 1 day and 999 days after the first instance of Kidney transplant | | |
|  | **Group 6B noDyslipidemia** | | | | |
|  | cannot have |  | diagnosis | UMLS:ICD10CM:E78 | Disorders of lipoprotein metabolism and other lipidemias |

##### **Table S41. Code definitions and selection criteria for SGLT2i users – heart failure.**

| Ungrouped terms | | | | | |
| --- | --- | --- | --- | --- | --- |
|  | must have |  | demographics | Age | Age (at least 18 years (most recent occurrence)) |
|  | cannot have |  | diagnosis | UMLS:ICD10CM:E08-E13 | Diabetes mellitus |
| Group 1 | | | | | |
|  | **Kidney transplant** | | | | |
|  | must have | any of | diagnosis | UMLS:ICD10CM:Z94.0 | Kidney transplant status |
|  |  |  | procedure | UMLS:ICD10PCS:0TY00Z0 | Transplantation of Right Kidney, Allogeneic, Open Approach |
|  |  |  | procedure | UMLS:ICD10PCS:0TY10Z0 | Transplantation of Left Kidney, Allogeneic, Open Approach |
|  |  |  | procedure | UMLS:ICD10PCS:0TY10Z1 | Transplantation of Left Kidney, Syngeneic, Open Approach |
|  |  |  | procedure | UMLS:ICD10PCS:0TY10Z2 | Transplantation of Left Kidney, Zooplastic, Open Approach |
|  |  |  | procedure | UMLS:ICD10PCS:0TY00Z1 | Transplantation of Right Kidney, Syngeneic, Open Approach |
|  |  |  | procedure | UMLS:ICD10PCS:0TY00Z2 | Transplantation of Right Kidney, Zooplastic, Open Approach |
|  |  |  | procedure | UMLS:ICD10PCS:0TY0 | Urinary System / Transplantation / Kidney, Right |
|  |  |  | procedure | UMLS:ICD10PCS:0TY1 | Urinary System / Transplantation / Kidney, Left |
|  | date constraint | | The terms in this group occurred between Jan 1, 2015 and Jan 1, 2022 | | |
| Group 2 | | | | | |
|  | **Group 2A Kidney transplant** | | | | |
|  | must have | any of | diagnosis | UMLS:ICD10CM:Z94.0 | Kidney transplant status |
|  |  |  | procedure | UMLS:ICD10PCS:0TY00Z0 | Transplantation of Right Kidney, Allogeneic, Open Approach |
|  |  |  | procedure | UMLS:ICD10PCS:0TY10Z0 | Transplantation of Left Kidney, Allogeneic, Open Approach |
|  |  |  | procedure | UMLS:ICD10PCS:0TY10Z1 | Transplantation of Left Kidney, Syngeneic, Open Approach |
|  |  |  | procedure | UMLS:ICD10PCS:0TY10Z2 | Transplantation of Left Kidney, Zooplastic, Open Approach |
|  |  |  | procedure | UMLS:ICD10PCS:0TY00Z1 | Transplantation of Right Kidney, Syngeneic, Open Approach |
|  |  |  | procedure | UMLS:ICD10PCS:0TY00Z2 | Transplantation of Right Kidney, Zooplastic, Open Approach |
|  |  |  | procedure | UMLS:ICD10PCS:0TY0 | Urinary System / Transplantation / Kidney, Right |
|  |  |  | procedure | UMLS:ICD10PCS:0TY1 | Urinary System / Transplantation / Kidney, Left |
|  | date constraint | | The terms in this group occurred at any time | | |
|  | event relationship | | Any instance of noDialysis, Death, SGLT2i occurred within 1 day and 999 days after the first instance of Kidney transplant | | |
|  | **Group 2B noDialysis, Death, SGLT2i** | | | | |
|  | cannot have |  | procedure | UMLS:CPT:1012740 | Dialysis Services and Procedures |
|  |  | or | procedure | UMLS:CPT:90945 | Dialysis procedure other than hemodialysis (eg, peritoneal dialysis, hemofiltration, or other continuous renal replacement therapies), with single evaluation by a physician or other qualified health care professional |
|  |  | or | procedure | UMLS:CPT:90947 | Dialysis procedure other than hemodialysis (eg, peritoneal dialysis, hemofiltration, or other continuous renal replacement therapies) requiring repeated evaluations by a physician or other qualified health care professional, with or without substantial revision of dialysis prescription |
|  |  | or | procedure | UMLS:CPT:1012752 | Hemodialysis Procedures |
|  |  | or | procedure | UMLS:SNOMED:302497006 | Hemodialysis |
|  |  | or | procedure | UMLS:CPT:90937 | Hemodialysis procedure requiring repeated evaluation(s) with or without substantial revision of dialysis prescription |
|  |  | or | procedure | UMLS:CPT:90935 | Hemodialysis procedure with single evaluation by a physician or other qualified health care professional |
|  |  | or | demographics | Deceased | Deceased |
|  |  | or | diagnosis | UMLS:ICD10CM:R99 | Ill-defined and unknown cause of mortality |
|  |  | or | medication | NLM:ATC:A10BK | Sodium-glucose co-transporter 2 (SGLT2) inhibitors |
| Group 3 | | | | | |
|  | **Group 3A Kidney transplant** | | | | |
|  | must have | any of | diagnosis | UMLS:ICD10CM:Z94.0 | Kidney transplant status |
|  |  |  | procedure | UMLS:ICD10PCS:0TY00Z0 | Transplantation of Right Kidney, Allogeneic, Open Approach |
|  |  |  | procedure | UMLS:ICD10PCS:0TY10Z0 | Transplantation of Left Kidney, Allogeneic, Open Approach |
|  |  |  | procedure | UMLS:ICD10PCS:0TY10Z1 | Transplantation of Left Kidney, Syngeneic, Open Approach |
|  |  |  | procedure | UMLS:ICD10PCS:0TY10Z2 | Transplantation of Left Kidney, Zooplastic, Open Approach |
|  |  |  | procedure | UMLS:ICD10PCS:0TY00Z1 | Transplantation of Right Kidney, Syngeneic, Open Approach |
|  |  |  | procedure | UMLS:ICD10PCS:0TY00Z2 | Transplantation of Right Kidney, Zooplastic, Open Approach |
|  |  |  | procedure | UMLS:ICD10PCS:0TY0 | Urinary System / Transplantation / Kidney, Right |
|  |  |  | procedure | UMLS:ICD10PCS:0TY1 | Urinary System / Transplantation / Kidney, Left |
|  | date constraint | | The terms in this group occurred at any time | | |
|  | event relationship | | Any instance of SGLT2i occurred at least 1000 days after the first instance of Kidney transplant | | |
|  | **Group 3B SGLT2i** | | | | |
|  | must have |  | medication | NLM:ATC:A10BK | Sodium-glucose co-transporter 2 (SGLT2) inhibitors |
| Group 4 | | | | | |
|  | **Group 4A Kidney transplant** | | | | |
|  | must have | any of | diagnosis | UMLS:ICD10CM:Z94.0 | Kidney transplant status |
|  |  |  | procedure | UMLS:ICD10PCS:0TY00Z0 | Transplantation of Right Kidney, Allogeneic, Open Approach |
|  |  |  | procedure | UMLS:ICD10PCS:0TY10Z0 | Transplantation of Left Kidney, Allogeneic, Open Approach |
|  |  |  | procedure | UMLS:ICD10PCS:0TY10Z1 | Transplantation of Left Kidney, Syngeneic, Open Approach |
|  |  |  | procedure | UMLS:ICD10PCS:0TY10Z2 | Transplantation of Left Kidney, Zooplastic, Open Approach |
|  |  |  | procedure | UMLS:ICD10PCS:0TY00Z1 | Transplantation of Right Kidney, Syngeneic, Open Approach |
|  |  |  | procedure | UMLS:ICD10PCS:0TY00Z2 | Transplantation of Right Kidney, Zooplastic, Open Approach |
|  |  |  | procedure | UMLS:ICD10PCS:0TY0 | Urinary System / Transplantation / Kidney, Right |
|  |  |  | procedure | UMLS:ICD10PCS:0TY1 | Urinary System / Transplantation / Kidney, Left |
|  | date constraint | | The terms in this group occurred at any time | | |
|  | event relationship | | Any instance of noSGLT2i occurred at least 1 day before the first instance of Kidney transplant | | |
|  | **Group 4B noSGLT2i** | | | | |
|  | cannot have |  | medication | NLM:ATC:A10BK | Sodium-glucose co-transporter 2 (SGLT2) inhibitors |
| Group 5 | | | | | |
|  | **Group 5A Kidney transplant** | | | | |
|  | must have | any of | diagnosis | UMLS:ICD10CM:Z94.0 | Kidney transplant status |
|  |  |  | procedure | UMLS:ICD10PCS:0TY00Z0 | Transplantation of Right Kidney, Allogeneic, Open Approach |
|  |  |  | procedure | UMLS:ICD10PCS:0TY10Z0 | Transplantation of Left Kidney, Allogeneic, Open Approach |
|  |  |  | procedure | UMLS:ICD10PCS:0TY10Z1 | Transplantation of Left Kidney, Syngeneic, Open Approach |
|  |  |  | procedure | UMLS:ICD10PCS:0TY10Z2 | Transplantation of Left Kidney, Zooplastic, Open Approach |
|  |  |  | procedure | UMLS:ICD10PCS:0TY00Z1 | Transplantation of Right Kidney, Syngeneic, Open Approach |
|  |  |  | procedure | UMLS:ICD10PCS:0TY00Z2 | Transplantation of Right Kidney, Zooplastic, Open Approach |
|  |  |  | procedure | UMLS:ICD10PCS:0TY0 | Urinary System / Transplantation / Kidney, Right |
|  |  |  | procedure | UMLS:ICD10PCS:0TY1 | Urinary System / Transplantation / Kidney, Left |
|  | date constraint | | The terms in this group occurred at any time | | |
|  | event relationship | | Any instance of Visit occurred at least 1181 days after the first instance of Kidney transplant | | |
|  | **Group 5B Visit** | | | | |
|  | must have |  | visit | TNX:Visit | Visit |
| Group 6 | | | | | |
|  | **Group 6A Kidney transplant** | | | | |
|  | must have | any of | diagnosis | UMLS:ICD10CM:Z94.0 | Kidney transplant status |
|  |  |  | procedure | UMLS:ICD10PCS:0TY00Z0 | Transplantation of Right Kidney, Allogeneic, Open Approach |
|  |  |  | procedure | UMLS:ICD10PCS:0TY10Z0 | Transplantation of Left Kidney, Allogeneic, Open Approach |
|  |  |  | procedure | UMLS:ICD10PCS:0TY10Z1 | Transplantation of Left Kidney, Syngeneic, Open Approach |
|  |  |  | procedure | UMLS:ICD10PCS:0TY10Z2 | Transplantation of Left Kidney, Zooplastic, Open Approach |
|  |  |  | procedure | UMLS:ICD10PCS:0TY00Z1 | Transplantation of Right Kidney, Syngeneic, Open Approach |
|  |  |  | procedure | UMLS:ICD10PCS:0TY00Z2 | Transplantation of Right Kidney, Zooplastic, Open Approach |
|  |  |  | procedure | UMLS:ICD10PCS:0TY0 | Urinary System / Transplantation / Kidney, Right |
|  |  |  | procedure | UMLS:ICD10PCS:0TY1 | Urinary System / Transplantation / Kidney, Left |
|  | date constraint | | The terms in this group occurred at any time | | |
|  | event relationship | | Any instance of Heart failure occurred within 1 year before or up to 1180 days after the first instance of Kidney transplant | | |
|  | **Group 6B Heart failure** | | | | |
|  | must have |  | diagnosis | UMLS:ICD10CM:I50 | Heart failure |

##### **Table S42. Code definitions and selection criteria for SGLT2i non-users – heart failure.**

| Ungrouped terms | | | | | |
| --- | --- | --- | --- | --- | --- |
|  | must have |  | demographics | Age | Age (at least 18 years (most recent occurrence)) |
|  | cannot have |  | diagnosis | UMLS:ICD10CM:E08-E13 | Diabetes mellitus |
| Group 1 | | | | | |
|  | **Kidney transplant** | | | | |
|  | must have | any of | diagnosis | UMLS:ICD10CM:Z94.0 | Kidney transplant status |
|  |  |  | procedure | UMLS:ICD10PCS:0TY00Z0 | Transplantation of Right Kidney, Allogeneic, Open Approach |
|  |  |  | procedure | UMLS:ICD10PCS:0TY10Z0 | Transplantation of Left Kidney, Allogeneic, Open Approach |
|  |  |  | procedure | UMLS:ICD10PCS:0TY10Z1 | Transplantation of Left Kidney, Syngeneic, Open Approach |
|  |  |  | procedure | UMLS:ICD10PCS:0TY10Z2 | Transplantation of Left Kidney, Zooplastic, Open Approach |
|  |  |  | procedure | UMLS:ICD10PCS:0TY00Z1 | Transplantation of Right Kidney, Syngeneic, Open Approach |
|  |  |  | procedure | UMLS:ICD10PCS:0TY00Z2 | Transplantation of Right Kidney, Zooplastic, Open Approach |
|  |  |  | procedure | UMLS:ICD10PCS:0TY0 | Urinary System / Transplantation / Kidney, Right |
|  |  |  | procedure | UMLS:ICD10PCS:0TY1 | Urinary System / Transplantation / Kidney, Left |
|  | date constraint | | The terms in this group occurred between Jan 1, 2015 and Jan 1, 2022 | | |
| Group 2 | | | | | |
|  | **Group 2A Kidney transplant** | | | | |
|  | must have | any of | diagnosis | UMLS:ICD10CM:Z94.0 | Kidney transplant status |
|  |  |  | procedure | UMLS:ICD10PCS:0TY00Z0 | Transplantation of Right Kidney, Allogeneic, Open Approach |
|  |  |  | procedure | UMLS:ICD10PCS:0TY10Z0 | Transplantation of Left Kidney, Allogeneic, Open Approach |
|  |  |  | procedure | UMLS:ICD10PCS:0TY10Z1 | Transplantation of Left Kidney, Syngeneic, Open Approach |
|  |  |  | procedure | UMLS:ICD10PCS:0TY10Z2 | Transplantation of Left Kidney, Zooplastic, Open Approach |
|  |  |  | procedure | UMLS:ICD10PCS:0TY00Z1 | Transplantation of Right Kidney, Syngeneic, Open Approach |
|  |  |  | procedure | UMLS:ICD10PCS:0TY00Z2 | Transplantation of Right Kidney, Zooplastic, Open Approach |
|  |  |  | procedure | UMLS:ICD10PCS:0TY0 | Urinary System / Transplantation / Kidney, Right |
|  |  |  | procedure | UMLS:ICD10PCS:0TY1 | Urinary System / Transplantation / Kidney, Left |
|  | date constraint | | The terms in this group occurred at any time | | |
|  | event relationship | | Any instance of noDialysis, Death, SGLT2i occurred within 1 day and 999 days after the first instance of Kidney transplant | | |
|  | **Group 2B noDialysis, Death, SGLT2i** | | | | |
|  | cannot have |  | procedure | UMLS:CPT:1012740 | Dialysis Services and Procedures |
|  |  | or | procedure | UMLS:CPT:90945 | Dialysis procedure other than hemodialysis (eg, peritoneal dialysis, hemofiltration, or other continuous renal replacement therapies), with single evaluation by a physician or other qualified health care professional |
|  |  | or | procedure | UMLS:CPT:90947 | Dialysis procedure other than hemodialysis (eg, peritoneal dialysis, hemofiltration, or other continuous renal replacement therapies) requiring repeated evaluations by a physician or other qualified health care professional, with or without substantial revision of dialysis prescription |
|  |  | or | procedure | UMLS:CPT:1012752 | Hemodialysis Procedures |
|  |  | or | procedure | UMLS:SNOMED:302497006 | Hemodialysis |
|  |  | or | procedure | UMLS:CPT:90937 | Hemodialysis procedure requiring repeated evaluation(s) with or without substantial revision of dialysis prescription |
|  |  | or | procedure | UMLS:CPT:90935 | Hemodialysis procedure with single evaluation by a physician or other qualified health care professional |
|  |  | or | demographics | Deceased | Deceased |
|  |  | or | diagnosis | UMLS:ICD10CM:R99 | Ill-defined and unknown cause of mortality |
|  |  | or | medication | NLM:ATC:A10BK | Sodium-glucose co-transporter 2 (SGLT2) inhibitors |
| Group 3 | | | | | |
|  | **Group 3A Kidney transplant** | | | | |
|  | must have | any of | diagnosis | UMLS:ICD10CM:Z94.0 | Kidney transplant status |
|  |  |  | procedure | UMLS:ICD10PCS:0TY00Z0 | Transplantation of Right Kidney, Allogeneic, Open Approach |
|  |  |  | procedure | UMLS:ICD10PCS:0TY10Z0 | Transplantation of Left Kidney, Allogeneic, Open Approach |
|  |  |  | procedure | UMLS:ICD10PCS:0TY10Z1 | Transplantation of Left Kidney, Syngeneic, Open Approach |
|  |  |  | procedure | UMLS:ICD10PCS:0TY10Z2 | Transplantation of Left Kidney, Zooplastic, Open Approach |
|  |  |  | procedure | UMLS:ICD10PCS:0TY00Z1 | Transplantation of Right Kidney, Syngeneic, Open Approach |
|  |  |  | procedure | UMLS:ICD10PCS:0TY00Z2 | Transplantation of Right Kidney, Zooplastic, Open Approach |
|  |  |  | procedure | UMLS:ICD10PCS:0TY0 | Urinary System / Transplantation / Kidney, Right |
|  |  |  | procedure | UMLS:ICD10PCS:0TY1 | Urinary System / Transplantation / Kidney, Left |
|  | date constraint | | The terms in this group occurred at any time | | |
|  | event relationship | | Any instance of noSGLT2i occurred within 1000 days and 3006 days after the first instance of Kidney transplant | | |
|  | **Group 3B noSGLT2i** | | | | |
|  | cannot have |  | medication | NLM:ATC:A10BK | Sodium-glucose co-transporter 2 (SGLT2) inhibitors |
| Group 4 | | | | | |
|  | **Group 4A Kidney transplant** | | | | |
|  | must have | any of | diagnosis | UMLS:ICD10CM:Z94.0 | Kidney transplant status |
|  |  |  | procedure | UMLS:ICD10PCS:0TY00Z0 | Transplantation of Right Kidney, Allogeneic, Open Approach |
|  |  |  | procedure | UMLS:ICD10PCS:0TY10Z0 | Transplantation of Left Kidney, Allogeneic, Open Approach |
|  |  |  | procedure | UMLS:ICD10PCS:0TY10Z1 | Transplantation of Left Kidney, Syngeneic, Open Approach |
|  |  |  | procedure | UMLS:ICD10PCS:0TY10Z2 | Transplantation of Left Kidney, Zooplastic, Open Approach |
|  |  |  | procedure | UMLS:ICD10PCS:0TY00Z1 | Transplantation of Right Kidney, Syngeneic, Open Approach |
|  |  |  | procedure | UMLS:ICD10PCS:0TY00Z2 | Transplantation of Right Kidney, Zooplastic, Open Approach |
|  |  |  | procedure | UMLS:ICD10PCS:0TY0 | Urinary System / Transplantation / Kidney, Right |
|  |  |  | procedure | UMLS:ICD10PCS:0TY1 | Urinary System / Transplantation / Kidney, Left |
|  | date constraint | | The terms in this group occurred at any time | | |
|  | event relationship | | Any instance of noSGLT2i occurred at least 1 day before the first instance of Kidney transplant | | |
|  | **Group 4B noSGLT2i** | | | | |
|  | cannot have |  | medication | NLM:ATC:A10BK | Sodium-glucose co-transporter 2 (SGLT2) inhibitors |
| Group 5 | | | | | |
|  | **Group 5A Kidney transplant** | | | | |
|  | must have | any of | diagnosis | UMLS:ICD10CM:Z94.0 | Kidney transplant status |
|  |  |  | procedure | UMLS:ICD10PCS:0TY00Z0 | Transplantation of Right Kidney, Allogeneic, Open Approach |
|  |  |  | procedure | UMLS:ICD10PCS:0TY10Z0 | Transplantation of Left Kidney, Allogeneic, Open Approach |
|  |  |  | procedure | UMLS:ICD10PCS:0TY10Z1 | Transplantation of Left Kidney, Syngeneic, Open Approach |
|  |  |  | procedure | UMLS:ICD10PCS:0TY10Z2 | Transplantation of Left Kidney, Zooplastic, Open Approach |
|  |  |  | procedure | UMLS:ICD10PCS:0TY00Z1 | Transplantation of Right Kidney, Syngeneic, Open Approach |
|  |  |  | procedure | UMLS:ICD10PCS:0TY00Z2 | Transplantation of Right Kidney, Zooplastic, Open Approach |
|  |  |  | procedure | UMLS:ICD10PCS:0TY0 | Urinary System / Transplantation / Kidney, Right |
|  |  |  | procedure | UMLS:ICD10PCS:0TY1 | Urinary System / Transplantation / Kidney, Left |
|  | date constraint | | The terms in this group occurred at any time | | |
|  | event relationship | | Any instance of Visit occurred at least 1181 days after the first instance of Kidney transplant | | |
|  | **Group 5B Visit** | | | | |
|  | must have |  | visit | TNX:Visit | Visit |
| Group 6 | | | | | |
|  | **Group 6A Kidney transplant** | | | | |
|  | must have | any of | diagnosis | UMLS:ICD10CM:Z94.0 | Kidney transplant status |
|  |  |  | procedure | UMLS:ICD10PCS:0TY00Z0 | Transplantation of Right Kidney, Allogeneic, Open Approach |
|  |  |  | procedure | UMLS:ICD10PCS:0TY10Z0 | Transplantation of Left Kidney, Allogeneic, Open Approach |
|  |  |  | procedure | UMLS:ICD10PCS:0TY10Z1 | Transplantation of Left Kidney, Syngeneic, Open Approach |
|  |  |  | procedure | UMLS:ICD10PCS:0TY10Z2 | Transplantation of Left Kidney, Zooplastic, Open Approach |
|  |  |  | procedure | UMLS:ICD10PCS:0TY00Z1 | Transplantation of Right Kidney, Syngeneic, Open Approach |
|  |  |  | procedure | UMLS:ICD10PCS:0TY00Z2 | Transplantation of Right Kidney, Zooplastic, Open Approach |
|  |  |  | procedure | UMLS:ICD10PCS:0TY0 | Urinary System / Transplantation / Kidney, Right |
|  |  |  | procedure | UMLS:ICD10PCS:0TY1 | Urinary System / Transplantation / Kidney, Left |
|  | date constraint | | The terms in this group occurred at any time | | |
|  | event relationship | | Any instance of Heart failure occurred within 1 year before or up to 1180 days after the first instance of Kidney transplant | | |
|  | **Group 6B Heart failure** | | | | |
|  | must have |  | diagnosis | UMLS:ICD10CM:I50 | Heart failure |

##### **Table S43. Code definitions and selection criteria for SGLT2i users – no heart failure.**

| Ungrouped terms | | | | | |
| --- | --- | --- | --- | --- | --- |
|  | must have |  | demographics | Age | Age (at least 18 years (most recent occurrence)) |
|  | cannot have |  | diagnosis | UMLS:ICD10CM:E08-E13 | Diabetes mellitus |
| Group 1 | | | | | |
|  | **Kidney transplant** | | | | |
|  | must have | any of | diagnosis | UMLS:ICD10CM:Z94.0 | Kidney transplant status |
|  |  |  | procedure | UMLS:ICD10PCS:0TY00Z0 | Transplantation of Right Kidney, Allogeneic, Open Approach |
|  |  |  | procedure | UMLS:ICD10PCS:0TY10Z0 | Transplantation of Left Kidney, Allogeneic, Open Approach |
|  |  |  | procedure | UMLS:ICD10PCS:0TY10Z1 | Transplantation of Left Kidney, Syngeneic, Open Approach |
|  |  |  | procedure | UMLS:ICD10PCS:0TY10Z2 | Transplantation of Left Kidney, Zooplastic, Open Approach |
|  |  |  | procedure | UMLS:ICD10PCS:0TY00Z1 | Transplantation of Right Kidney, Syngeneic, Open Approach |
|  |  |  | procedure | UMLS:ICD10PCS:0TY00Z2 | Transplantation of Right Kidney, Zooplastic, Open Approach |
|  |  |  | procedure | UMLS:ICD10PCS:0TY0 | Urinary System / Transplantation / Kidney, Right |
|  |  |  | procedure | UMLS:ICD10PCS:0TY1 | Urinary System / Transplantation / Kidney, Left |
|  | date constraint | | The terms in this group occurred between Jan 1, 2015 and Jan 1, 2022 | | |
| Group 2 | | | | | |
|  | **Group 2A Kidney transplant** | | | | |
|  | must have | any of | diagnosis | UMLS:ICD10CM:Z94.0 | Kidney transplant status |
|  |  |  | procedure | UMLS:ICD10PCS:0TY00Z0 | Transplantation of Right Kidney, Allogeneic, Open Approach |
|  |  |  | procedure | UMLS:ICD10PCS:0TY10Z0 | Transplantation of Left Kidney, Allogeneic, Open Approach |
|  |  |  | procedure | UMLS:ICD10PCS:0TY10Z1 | Transplantation of Left Kidney, Syngeneic, Open Approach |
|  |  |  | procedure | UMLS:ICD10PCS:0TY10Z2 | Transplantation of Left Kidney, Zooplastic, Open Approach |
|  |  |  | procedure | UMLS:ICD10PCS:0TY00Z1 | Transplantation of Right Kidney, Syngeneic, Open Approach |
|  |  |  | procedure | UMLS:ICD10PCS:0TY00Z2 | Transplantation of Right Kidney, Zooplastic, Open Approach |
|  |  |  | procedure | UMLS:ICD10PCS:0TY0 | Urinary System / Transplantation / Kidney, Right |
|  |  |  | procedure | UMLS:ICD10PCS:0TY1 | Urinary System / Transplantation / Kidney, Left |
|  | date constraint | | The terms in this group occurred at any time | | |
|  | event relationship | | Any instance of noDialysis, Death, SGLT2i occurred within 1 day and 999 days after the first instance of Kidney transplant | | |
|  | **Group 2B noDialysis, Death, SGLT2i** | | | | |
|  | cannot have |  | procedure | UMLS:CPT:1012740 | Dialysis Services and Procedures |
|  |  | or | procedure | UMLS:CPT:90945 | Dialysis procedure other than hemodialysis (eg, peritoneal dialysis, hemofiltration, or other continuous renal replacement therapies), with single evaluation by a physician or other qualified health care professional |
|  |  | or | procedure | UMLS:CPT:90947 | Dialysis procedure other than hemodialysis (eg, peritoneal dialysis, hemofiltration, or other continuous renal replacement therapies) requiring repeated evaluations by a physician or other qualified health care professional, with or without substantial revision of dialysis prescription |
|  |  | or | procedure | UMLS:CPT:1012752 | Hemodialysis Procedures |
|  |  | or | procedure | UMLS:SNOMED:302497006 | Hemodialysis |
|  |  | or | procedure | UMLS:CPT:90937 | Hemodialysis procedure requiring repeated evaluation(s) with or without substantial revision of dialysis prescription |
|  |  | or | procedure | UMLS:CPT:90935 | Hemodialysis procedure with single evaluation by a physician or other qualified health care professional |
|  |  | or | demographics | Deceased | Deceased |
|  |  | or | diagnosis | UMLS:ICD10CM:R99 | Ill-defined and unknown cause of mortality |
|  |  | or | medication | NLM:ATC:A10BK | Sodium-glucose co-transporter 2 (SGLT2) inhibitors |
| Group 3 | | | | | |
|  | **Group 3A Kidney transplant** | | | | |
|  | must have | any of | diagnosis | UMLS:ICD10CM:Z94.0 | Kidney transplant status |
|  |  |  | procedure | UMLS:ICD10PCS:0TY00Z0 | Transplantation of Right Kidney, Allogeneic, Open Approach |
|  |  |  | procedure | UMLS:ICD10PCS:0TY10Z0 | Transplantation of Left Kidney, Allogeneic, Open Approach |
|  |  |  | procedure | UMLS:ICD10PCS:0TY10Z1 | Transplantation of Left Kidney, Syngeneic, Open Approach |
|  |  |  | procedure | UMLS:ICD10PCS:0TY10Z2 | Transplantation of Left Kidney, Zooplastic, Open Approach |
|  |  |  | procedure | UMLS:ICD10PCS:0TY00Z1 | Transplantation of Right Kidney, Syngeneic, Open Approach |
|  |  |  | procedure | UMLS:ICD10PCS:0TY00Z2 | Transplantation of Right Kidney, Zooplastic, Open Approach |
|  |  |  | procedure | UMLS:ICD10PCS:0TY0 | Urinary System / Transplantation / Kidney, Right |
|  |  |  | procedure | UMLS:ICD10PCS:0TY1 | Urinary System / Transplantation / Kidney, Left |
|  | date constraint | | The terms in this group occurred at any time | | |
|  | event relationship | | Any instance of SGLT2i occurred at least 1000 days after the first instance of Kidney transplant | | |
|  | **Group 3B SGLT2i** | | | | |
|  | must have |  | medication | NLM:ATC:A10BK | Sodium-glucose co-transporter 2 (SGLT2) inhibitors |
| Group 4 | | | | | |
|  | **Group 4A Kidney transplant** | | | | |
|  | must have | any of | diagnosis | UMLS:ICD10CM:Z94.0 | Kidney transplant status |
|  |  |  | procedure | UMLS:ICD10PCS:0TY00Z0 | Transplantation of Right Kidney, Allogeneic, Open Approach |
|  |  |  | procedure | UMLS:ICD10PCS:0TY10Z0 | Transplantation of Left Kidney, Allogeneic, Open Approach |
|  |  |  | procedure | UMLS:ICD10PCS:0TY10Z1 | Transplantation of Left Kidney, Syngeneic, Open Approach |
|  |  |  | procedure | UMLS:ICD10PCS:0TY10Z2 | Transplantation of Left Kidney, Zooplastic, Open Approach |
|  |  |  | procedure | UMLS:ICD10PCS:0TY00Z1 | Transplantation of Right Kidney, Syngeneic, Open Approach |
|  |  |  | procedure | UMLS:ICD10PCS:0TY00Z2 | Transplantation of Right Kidney, Zooplastic, Open Approach |
|  |  |  | procedure | UMLS:ICD10PCS:0TY0 | Urinary System / Transplantation / Kidney, Right |
|  |  |  | procedure | UMLS:ICD10PCS:0TY1 | Urinary System / Transplantation / Kidney, Left |
|  | date constraint | | The terms in this group occurred at any time | | |
|  | event relationship | | Any instance of noSGLT2i occurred at least 1 day before the first instance of Kidney transplant | | |
|  | **Group 4B noSGLT2i** | | | | |
|  | cannot have |  | medication | NLM:ATC:A10BK | Sodium-glucose co-transporter 2 (SGLT2) inhibitors |
| Group 5 | | | | | |
|  | **Group 5A Kidney transplant** | | | | |
|  | must have | any of | diagnosis | UMLS:ICD10CM:Z94.0 | Kidney transplant status |
|  |  |  | procedure | UMLS:ICD10PCS:0TY00Z0 | Transplantation of Right Kidney, Allogeneic, Open Approach |
|  |  |  | procedure | UMLS:ICD10PCS:0TY10Z0 | Transplantation of Left Kidney, Allogeneic, Open Approach |
|  |  |  | procedure | UMLS:ICD10PCS:0TY10Z1 | Transplantation of Left Kidney, Syngeneic, Open Approach |
|  |  |  | procedure | UMLS:ICD10PCS:0TY10Z2 | Transplantation of Left Kidney, Zooplastic, Open Approach |
|  |  |  | procedure | UMLS:ICD10PCS:0TY00Z1 | Transplantation of Right Kidney, Syngeneic, Open Approach |
|  |  |  | procedure | UMLS:ICD10PCS:0TY00Z2 | Transplantation of Right Kidney, Zooplastic, Open Approach |
|  |  |  | procedure | UMLS:ICD10PCS:0TY0 | Urinary System / Transplantation / Kidney, Right |
|  |  |  | procedure | UMLS:ICD10PCS:0TY1 | Urinary System / Transplantation / Kidney, Left |
|  | date constraint | | The terms in this group occurred at any time | | |
|  | event relationship | | Any instance of Visit occurred at least 1181 days after the first instance of Kidney transplant | | |
|  | **Group 5B Visit** | | | | |
|  | must have |  | visit | TNX:Visit | Visit |
| Group 6 | | | | | |
|  | **Group 6A Kidney transplant** | | | | |
|  | must have | any of | diagnosis | UMLS:ICD10CM:Z94.0 | Kidney transplant status |
|  |  |  | procedure | UMLS:ICD10PCS:0TY00Z0 | Transplantation of Right Kidney, Allogeneic, Open Approach |
|  |  |  | procedure | UMLS:ICD10PCS:0TY10Z0 | Transplantation of Left Kidney, Allogeneic, Open Approach |
|  |  |  | procedure | UMLS:ICD10PCS:0TY10Z1 | Transplantation of Left Kidney, Syngeneic, Open Approach |
|  |  |  | procedure | UMLS:ICD10PCS:0TY10Z2 | Transplantation of Left Kidney, Zooplastic, Open Approach |
|  |  |  | procedure | UMLS:ICD10PCS:0TY00Z1 | Transplantation of Right Kidney, Syngeneic, Open Approach |
|  |  |  | procedure | UMLS:ICD10PCS:0TY00Z2 | Transplantation of Right Kidney, Zooplastic, Open Approach |
|  |  |  | procedure | UMLS:ICD10PCS:0TY0 | Urinary System / Transplantation / Kidney, Right |
|  |  |  | procedure | UMLS:ICD10PCS:0TY1 | Urinary System / Transplantation / Kidney, Left |
|  | date constraint | | The terms in this group occurred at any time | | |
|  | event relationship | | Any instance of noHeart failure occurred within 1 day and 999 days after the first instance of Kidney transplant | | |
|  | **Group 6B noHeart failure** | | | | |
|  | cannot have |  | diagnosis | UMLS:ICD10CM:I50 | Heart failure |

##### **Table S44. Code definitions and selection criteria for SGLT2i non-users – no heart failure.**

| Ungrouped terms | | | | | |
| --- | --- | --- | --- | --- | --- |
|  | must have |  | demographics | Age | Age (at least 18 years (most recent occurrence)) |
|  | cannot have |  | diagnosis | UMLS:ICD10CM:E08-E13 | Diabetes mellitus |
| Group 1 | | | | | |
|  | **Kidney transplant** | | | | |
|  | must have | any of | diagnosis | UMLS:ICD10CM:Z94.0 | Kidney transplant status |
|  |  |  | procedure | UMLS:ICD10PCS:0TY00Z0 | Transplantation of Right Kidney, Allogeneic, Open Approach |
|  |  |  | procedure | UMLS:ICD10PCS:0TY10Z0 | Transplantation of Left Kidney, Allogeneic, Open Approach |
|  |  |  | procedure | UMLS:ICD10PCS:0TY10Z1 | Transplantation of Left Kidney, Syngeneic, Open Approach |
|  |  |  | procedure | UMLS:ICD10PCS:0TY10Z2 | Transplantation of Left Kidney, Zooplastic, Open Approach |
|  |  |  | procedure | UMLS:ICD10PCS:0TY00Z1 | Transplantation of Right Kidney, Syngeneic, Open Approach |
|  |  |  | procedure | UMLS:ICD10PCS:0TY00Z2 | Transplantation of Right Kidney, Zooplastic, Open Approach |
|  |  |  | procedure | UMLS:ICD10PCS:0TY0 | Urinary System / Transplantation / Kidney, Right |
|  |  |  | procedure | UMLS:ICD10PCS:0TY1 | Urinary System / Transplantation / Kidney, Left |
|  | date constraint | | The terms in this group occurred between Jan 1, 2015 and Jan 1, 2022 | | |
| Group 2 | | | | | |
|  | **Group 2A Kidney transplant** | | | | |
|  | must have | any of | diagnosis | UMLS:ICD10CM:Z94.0 | Kidney transplant status |
|  |  |  | procedure | UMLS:ICD10PCS:0TY00Z0 | Transplantation of Right Kidney, Allogeneic, Open Approach |
|  |  |  | procedure | UMLS:ICD10PCS:0TY10Z0 | Transplantation of Left Kidney, Allogeneic, Open Approach |
|  |  |  | procedure | UMLS:ICD10PCS:0TY10Z1 | Transplantation of Left Kidney, Syngeneic, Open Approach |
|  |  |  | procedure | UMLS:ICD10PCS:0TY10Z2 | Transplantation of Left Kidney, Zooplastic, Open Approach |
|  |  |  | procedure | UMLS:ICD10PCS:0TY00Z1 | Transplantation of Right Kidney, Syngeneic, Open Approach |
|  |  |  | procedure | UMLS:ICD10PCS:0TY00Z2 | Transplantation of Right Kidney, Zooplastic, Open Approach |
|  |  |  | procedure | UMLS:ICD10PCS:0TY0 | Urinary System / Transplantation / Kidney, Right |
|  |  |  | procedure | UMLS:ICD10PCS:0TY1 | Urinary System / Transplantation / Kidney, Left |
|  | date constraint | | The terms in this group occurred at any time | | |
|  | event relationship | | Any instance of noDialysis, Death, SGLT2i occurred within 1 day and 999 days after the first instance of Kidney transplant | | |
|  | **Group 2B noDialysis, Death, SGLT2i** | | | | |
|  | cannot have |  | procedure | UMLS:CPT:1012740 | Dialysis Services and Procedures |
|  |  | or | procedure | UMLS:CPT:90945 | Dialysis procedure other than hemodialysis (eg, peritoneal dialysis, hemofiltration, or other continuous renal replacement therapies), with single evaluation by a physician or other qualified health care professional |
|  |  | or | procedure | UMLS:CPT:90947 | Dialysis procedure other than hemodialysis (eg, peritoneal dialysis, hemofiltration, or other continuous renal replacement therapies) requiring repeated evaluations by a physician or other qualified health care professional, with or without substantial revision of dialysis prescription |
|  |  | or | procedure | UMLS:CPT:1012752 | Hemodialysis Procedures |
|  |  | or | procedure | UMLS:SNOMED:302497006 | Hemodialysis |
|  |  | or | procedure | UMLS:CPT:90937 | Hemodialysis procedure requiring repeated evaluation(s) with or without substantial revision of dialysis prescription |
|  |  | or | procedure | UMLS:CPT:90935 | Hemodialysis procedure with single evaluation by a physician or other qualified health care professional |
|  |  | or | demographics | Deceased | Deceased |
|  |  | or | diagnosis | UMLS:ICD10CM:R99 | Ill-defined and unknown cause of mortality |
|  |  | or | medication | NLM:ATC:A10BK | Sodium-glucose co-transporter 2 (SGLT2) inhibitors |
| Group 3 | | | | | |
|  | **Group 3A Kidney transplant** | | | | |
|  | must have | any of | diagnosis | UMLS:ICD10CM:Z94.0 | Kidney transplant status |
|  |  |  | procedure | UMLS:ICD10PCS:0TY00Z0 | Transplantation of Right Kidney, Allogeneic, Open Approach |
|  |  |  | procedure | UMLS:ICD10PCS:0TY10Z0 | Transplantation of Left Kidney, Allogeneic, Open Approach |
|  |  |  | procedure | UMLS:ICD10PCS:0TY10Z1 | Transplantation of Left Kidney, Syngeneic, Open Approach |
|  |  |  | procedure | UMLS:ICD10PCS:0TY10Z2 | Transplantation of Left Kidney, Zooplastic, Open Approach |
|  |  |  | procedure | UMLS:ICD10PCS:0TY00Z1 | Transplantation of Right Kidney, Syngeneic, Open Approach |
|  |  |  | procedure | UMLS:ICD10PCS:0TY00Z2 | Transplantation of Right Kidney, Zooplastic, Open Approach |
|  |  |  | procedure | UMLS:ICD10PCS:0TY0 | Urinary System / Transplantation / Kidney, Right |
|  |  |  | procedure | UMLS:ICD10PCS:0TY1 | Urinary System / Transplantation / Kidney, Left |
|  | date constraint | | The terms in this group occurred at any time | | |
|  | event relationship | | Any instance of SGLT2i occurred within 1000 days and 3006 days after the first instance of Kidney transplant | | |
|  | **Group 3B SGLT2i** | | | | |
|  | cannot have |  | medication | NLM:ATC:A10BK | Sodium-glucose co-transporter 2 (SGLT2) inhibitors |
| Group 4 | | | | | |
|  | **Group 4A Kidney transplant** | | | | |
|  | must have | any of | diagnosis | UMLS:ICD10CM:Z94.0 | Kidney transplant status |
|  |  |  | procedure | UMLS:ICD10PCS:0TY00Z0 | Transplantation of Right Kidney, Allogeneic, Open Approach |
|  |  |  | procedure | UMLS:ICD10PCS:0TY10Z0 | Transplantation of Left Kidney, Allogeneic, Open Approach |
|  |  |  | procedure | UMLS:ICD10PCS:0TY10Z1 | Transplantation of Left Kidney, Syngeneic, Open Approach |
|  |  |  | procedure | UMLS:ICD10PCS:0TY10Z2 | Transplantation of Left Kidney, Zooplastic, Open Approach |
|  |  |  | procedure | UMLS:ICD10PCS:0TY00Z1 | Transplantation of Right Kidney, Syngeneic, Open Approach |
|  |  |  | procedure | UMLS:ICD10PCS:0TY00Z2 | Transplantation of Right Kidney, Zooplastic, Open Approach |
|  |  |  | procedure | UMLS:ICD10PCS:0TY0 | Urinary System / Transplantation / Kidney, Right |
|  |  |  | procedure | UMLS:ICD10PCS:0TY1 | Urinary System / Transplantation / Kidney, Left |
|  | date constraint | | The terms in this group occurred at any time | | |
|  | event relationship | | Any instance of noSGLT2i occurred at least 1 day before the first instance of Kidney transplant | | |
|  | **Group 4B noSGLT2i** | | | | |
|  | cannot have |  | medication | NLM:ATC:A10BK | Sodium-glucose co-transporter 2 (SGLT2) inhibitors |
| Group 5 | | | | | |
|  | **Group 5A Kidney transplant** | | | | |
|  | must have | any of | diagnosis | UMLS:ICD10CM:Z94.0 | Kidney transplant status |
|  |  |  | procedure | UMLS:ICD10PCS:0TY00Z0 | Transplantation of Right Kidney, Allogeneic, Open Approach |
|  |  |  | procedure | UMLS:ICD10PCS:0TY10Z0 | Transplantation of Left Kidney, Allogeneic, Open Approach |
|  |  |  | procedure | UMLS:ICD10PCS:0TY10Z1 | Transplantation of Left Kidney, Syngeneic, Open Approach |
|  |  |  | procedure | UMLS:ICD10PCS:0TY10Z2 | Transplantation of Left Kidney, Zooplastic, Open Approach |
|  |  |  | procedure | UMLS:ICD10PCS:0TY00Z1 | Transplantation of Right Kidney, Syngeneic, Open Approach |
|  |  |  | procedure | UMLS:ICD10PCS:0TY00Z2 | Transplantation of Right Kidney, Zooplastic, Open Approach |
|  |  |  | procedure | UMLS:ICD10PCS:0TY0 | Urinary System / Transplantation / Kidney, Right |
|  |  |  | procedure | UMLS:ICD10PCS:0TY1 | Urinary System / Transplantation / Kidney, Left |
|  | date constraint | | The terms in this group occurred at any time | | |
|  | event relationship | | Any instance of Visit occurred at least 1181 days after the first instance of Kidney transplant | | |
|  | **Group 5B Visit** | | | | |
|  | must have |  | visit | TNX:Visit | Visit |
| Group 6 | | | | | |
|  | **Group 6A Kidney transplant** | | | | |
|  | must have | any of | diagnosis | UMLS:ICD10CM:Z94.0 | Kidney transplant status |
|  |  |  | procedure | UMLS:ICD10PCS:0TY00Z0 | Transplantation of Right Kidney, Allogeneic, Open Approach |
|  |  |  | procedure | UMLS:ICD10PCS:0TY10Z0 | Transplantation of Left Kidney, Allogeneic, Open Approach |
|  |  |  | procedure | UMLS:ICD10PCS:0TY10Z1 | Transplantation of Left Kidney, Syngeneic, Open Approach |
|  |  |  | procedure | UMLS:ICD10PCS:0TY10Z2 | Transplantation of Left Kidney, Zooplastic, Open Approach |
|  |  |  | procedure | UMLS:ICD10PCS:0TY00Z1 | Transplantation of Right Kidney, Syngeneic, Open Approach |
|  |  |  | procedure | UMLS:ICD10PCS:0TY00Z2 | Transplantation of Right Kidney, Zooplastic, Open Approach |
|  |  |  | procedure | UMLS:ICD10PCS:0TY0 | Urinary System / Transplantation / Kidney, Right |
|  |  |  | procedure | UMLS:ICD10PCS:0TY1 | Urinary System / Transplantation / Kidney, Left |
|  | date constraint | | The terms in this group occurred at any time | | |
|  | event relationship | | Any instance of noHeart failure occurred within 1 day and 999 days after the first instance of Kidney transplant | | |
|  | **Group 6B noHeart failure** | | | | |
|  | cannot have |  | diagnosis | UMLS:ICD10CM:I50 | Heart failure |

#### Laboratory parameters

##### **Table S45. Code definitions and selection criteria for SGLT2i users – <30 BMI.**

| Ungrouped terms | | | | | |
| --- | --- | --- | --- | --- | --- |
|  | must have |  | demographics | Age | Age (at least 18 years (most recent occurrence)) |
|  | cannot have |  | diagnosis | UMLS:ICD10CM:E08-E13 | Diabetes mellitus |
| Group 1 | | | | | |
|  | **Kidney transplant** | | | | |
|  | must have | any of | diagnosis | UMLS:ICD10CM:Z94.0 | Kidney transplant status |
|  |  |  | procedure | UMLS:ICD10PCS:0TY00Z0 | Transplantation of Right Kidney, Allogeneic, Open Approach |
|  |  |  | procedure | UMLS:ICD10PCS:0TY10Z0 | Transplantation of Left Kidney, Allogeneic, Open Approach |
|  |  |  | procedure | UMLS:ICD10PCS:0TY10Z1 | Transplantation of Left Kidney, Syngeneic, Open Approach |
|  |  |  | procedure | UMLS:ICD10PCS:0TY10Z2 | Transplantation of Left Kidney, Zooplastic, Open Approach |
|  |  |  | procedure | UMLS:ICD10PCS:0TY00Z1 | Transplantation of Right Kidney, Syngeneic, Open Approach |
|  |  |  | procedure | UMLS:ICD10PCS:0TY00Z2 | Transplantation of Right Kidney, Zooplastic, Open Approach |
|  |  |  | procedure | UMLS:ICD10PCS:0TY0 | Urinary System / Transplantation / Kidney, Right |
|  |  |  | procedure | UMLS:ICD10PCS:0TY1 | Urinary System / Transplantation / Kidney, Left |
|  | date constraint | | The terms in this group occurred between Jan 1, 2015 and Jan 1, 2022 | | |
| Group 2 | | | | | |
|  | **Group 2A Kidney transplant** | | | | |
|  | must have | any of | diagnosis | UMLS:ICD10CM:Z94.0 | Kidney transplant status |
|  |  |  | procedure | UMLS:ICD10PCS:0TY00Z0 | Transplantation of Right Kidney, Allogeneic, Open Approach |
|  |  |  | procedure | UMLS:ICD10PCS:0TY10Z0 | Transplantation of Left Kidney, Allogeneic, Open Approach |
|  |  |  | procedure | UMLS:ICD10PCS:0TY10Z1 | Transplantation of Left Kidney, Syngeneic, Open Approach |
|  |  |  | procedure | UMLS:ICD10PCS:0TY10Z2 | Transplantation of Left Kidney, Zooplastic, Open Approach |
|  |  |  | procedure | UMLS:ICD10PCS:0TY00Z1 | Transplantation of Right Kidney, Syngeneic, Open Approach |
|  |  |  | procedure | UMLS:ICD10PCS:0TY00Z2 | Transplantation of Right Kidney, Zooplastic, Open Approach |
|  |  |  | procedure | UMLS:ICD10PCS:0TY0 | Urinary System / Transplantation / Kidney, Right |
|  |  |  | procedure | UMLS:ICD10PCS:0TY1 | Urinary System / Transplantation / Kidney, Left |
|  | date constraint | | The terms in this group occurred at any time | | |
|  | event relationship | | Any instance of noDialysis, Death, SGLT2i occurred within 1 day and 999 days after the first instance of Kidney transplant | | |
|  | **Group 2B noDialysis, Death, SGLT2i** | | | | |
|  | cannot have |  | procedure | UMLS:CPT:1012740 | Dialysis Services and Procedures |
|  |  | or | procedure | UMLS:CPT:90945 | Dialysis procedure other than hemodialysis (eg, peritoneal dialysis, hemofiltration, or other continuous renal replacement therapies), with single evaluation by a physician or other qualified health care professional |
|  |  | or | procedure | UMLS:CPT:90947 | Dialysis procedure other than hemodialysis (eg, peritoneal dialysis, hemofiltration, or other continuous renal replacement therapies) requiring repeated evaluations by a physician or other qualified health care professional, with or without substantial revision of dialysis prescription |
|  |  | or | procedure | UMLS:CPT:1012752 | Hemodialysis Procedures |
|  |  | or | procedure | UMLS:SNOMED:302497006 | Hemodialysis |
|  |  | or | procedure | UMLS:CPT:90937 | Hemodialysis procedure requiring repeated evaluation(s) with or without substantial revision of dialysis prescription |
|  |  | or | procedure | UMLS:CPT:90935 | Hemodialysis procedure with single evaluation by a physician or other qualified health care professional |
|  |  | or | demographics | Deceased | Deceased |
|  |  | or | diagnosis | UMLS:ICD10CM:R99 | Ill-defined and unknown cause of mortality |
|  |  | or | medication | NLM:ATC:A10BK | Sodium-glucose co-transporter 2 (SGLT2) inhibitors |
| Group 3 | | | | | |
|  | **Group 3A Kidney transplant** | | | | |
|  | must have | any of | diagnosis | UMLS:ICD10CM:Z94.0 | Kidney transplant status |
|  |  |  | procedure | UMLS:ICD10PCS:0TY00Z0 | Transplantation of Right Kidney, Allogeneic, Open Approach |
|  |  |  | procedure | UMLS:ICD10PCS:0TY10Z0 | Transplantation of Left Kidney, Allogeneic, Open Approach |
|  |  |  | procedure | UMLS:ICD10PCS:0TY10Z1 | Transplantation of Left Kidney, Syngeneic, Open Approach |
|  |  |  | procedure | UMLS:ICD10PCS:0TY10Z2 | Transplantation of Left Kidney, Zooplastic, Open Approach |
|  |  |  | procedure | UMLS:ICD10PCS:0TY00Z1 | Transplantation of Right Kidney, Syngeneic, Open Approach |
|  |  |  | procedure | UMLS:ICD10PCS:0TY00Z2 | Transplantation of Right Kidney, Zooplastic, Open Approach |
|  |  |  | procedure | UMLS:ICD10PCS:0TY0 | Urinary System / Transplantation / Kidney, Right |
|  |  |  | procedure | UMLS:ICD10PCS:0TY1 | Urinary System / Transplantation / Kidney, Left |
|  | date constraint | | The terms in this group occurred at any time | | |
|  | event relationship | | Any instance of SGLT2i occurred at least 1000 days after the first instance of Kidney transplant | | |
|  | **Group 3B SGLT2i** | | | | |
|  | must have |  | medication | NLM:ATC:A10BK | Sodium-glucose co-transporter 2 (SGLT2) inhibitors |
| Group 4 | | | | | |
|  | **Group 4A Kidney transplant** | | | | |
|  | must have | any of | diagnosis | UMLS:ICD10CM:Z94.0 | Kidney transplant status |
|  |  |  | procedure | UMLS:ICD10PCS:0TY00Z0 | Transplantation of Right Kidney, Allogeneic, Open Approach |
|  |  |  | procedure | UMLS:ICD10PCS:0TY10Z0 | Transplantation of Left Kidney, Allogeneic, Open Approach |
|  |  |  | procedure | UMLS:ICD10PCS:0TY10Z1 | Transplantation of Left Kidney, Syngeneic, Open Approach |
|  |  |  | procedure | UMLS:ICD10PCS:0TY10Z2 | Transplantation of Left Kidney, Zooplastic, Open Approach |
|  |  |  | procedure | UMLS:ICD10PCS:0TY00Z1 | Transplantation of Right Kidney, Syngeneic, Open Approach |
|  |  |  | procedure | UMLS:ICD10PCS:0TY00Z2 | Transplantation of Right Kidney, Zooplastic, Open Approach |
|  |  |  | procedure | UMLS:ICD10PCS:0TY0 | Urinary System / Transplantation / Kidney, Right |
|  |  |  | procedure | UMLS:ICD10PCS:0TY1 | Urinary System / Transplantation / Kidney, Left |
|  | date constraint | | The terms in this group occurred at any time | | |
|  | event relationship | | Any instance of noSGLT2i occurred at least 1 day before the first instance of Kidney transplant | | |
|  | **Group 4B noSGLT2i** | | | | |
|  | cannot have |  | medication | NLM:ATC:A10BK | Sodium-glucose co-transporter 2 (SGLT2) inhibitors |
| Group 5 | | | | | |
|  | **Group 5A Kidney transplant** | | | | |
|  | must have | any of | diagnosis | UMLS:ICD10CM:Z94.0 | Kidney transplant status |
|  |  |  | procedure | UMLS:ICD10PCS:0TY00Z0 | Transplantation of Right Kidney, Allogeneic, Open Approach |
|  |  |  | procedure | UMLS:ICD10PCS:0TY10Z0 | Transplantation of Left Kidney, Allogeneic, Open Approach |
|  |  |  | procedure | UMLS:ICD10PCS:0TY10Z1 | Transplantation of Left Kidney, Syngeneic, Open Approach |
|  |  |  | procedure | UMLS:ICD10PCS:0TY10Z2 | Transplantation of Left Kidney, Zooplastic, Open Approach |
|  |  |  | procedure | UMLS:ICD10PCS:0TY00Z1 | Transplantation of Right Kidney, Syngeneic, Open Approach |
|  |  |  | procedure | UMLS:ICD10PCS:0TY00Z2 | Transplantation of Right Kidney, Zooplastic, Open Approach |
|  |  |  | procedure | UMLS:ICD10PCS:0TY0 | Urinary System / Transplantation / Kidney, Right |
|  |  |  | procedure | UMLS:ICD10PCS:0TY1 | Urinary System / Transplantation / Kidney, Left |
|  | date constraint | | The terms in this group occurred at any time | | |
|  | event relationship | | Any instance of Visit occurred at least 1181 days after the first instance of Kidney transplant | | |
|  | **Group 5B Visit** | | | | |
|  | must have |  | visit | TNX:Visit | Visit |
| Group 6 | | | | | |
|  | **Group 6A Kidney transplant** | | | | |
|  | must have | any of | diagnosis | UMLS:ICD10CM:Z94.0 | Kidney transplant status |
|  |  |  | procedure | UMLS:ICD10PCS:0TY00Z0 | Transplantation of Right Kidney, Allogeneic, Open Approach |
|  |  |  | procedure | UMLS:ICD10PCS:0TY10Z0 | Transplantation of Left Kidney, Allogeneic, Open Approach |
|  |  |  | procedure | UMLS:ICD10PCS:0TY10Z1 | Transplantation of Left Kidney, Syngeneic, Open Approach |
|  |  |  | procedure | UMLS:ICD10PCS:0TY10Z2 | Transplantation of Left Kidney, Zooplastic, Open Approach |
|  |  |  | procedure | UMLS:ICD10PCS:0TY00Z1 | Transplantation of Right Kidney, Syngeneic, Open Approach |
|  |  |  | procedure | UMLS:ICD10PCS:0TY00Z2 | Transplantation of Right Kidney, Zooplastic, Open Approach |
|  |  |  | procedure | UMLS:ICD10PCS:0TY0 | Urinary System / Transplantation / Kidney, Right |
|  |  |  | procedure | UMLS:ICD10PCS:0TY1 | Urinary System / Transplantation / Kidney, Left |
|  | date constraint | | The terms in this group occurred at any time | | |
|  | event relationship | | Any instance of <30 BMI occurred within 1 day and 999 days after the first instance of Kidney transplant | | |
|  | **Group 6B <30 BMI** | | | | |
|  | must have |  | laboratory | TNX:9083 | BMI (between 0.00 and 30.00 kg/m2) |

##### **Table S46. Code definitions and selection criteria for SGLT2i non-users – <30 BMI.**

| Ungrouped terms | | | | | |
| --- | --- | --- | --- | --- | --- |
|  | must have |  | demographics | Age | Age (at least 18 years (most recent occurrence)) |
|  | cannot have |  | diagnosis | UMLS:ICD10CM:E08-E13 | Diabetes mellitus |
| Group 1 | | | | | |
|  | **Kidney transplant** | | | | |
|  | must have | any of | diagnosis | UMLS:ICD10CM:Z94.0 | Kidney transplant status |
|  |  |  | procedure | UMLS:ICD10PCS:0TY00Z0 | Transplantation of Right Kidney, Allogeneic, Open Approach |
|  |  |  | procedure | UMLS:ICD10PCS:0TY10Z0 | Transplantation of Left Kidney, Allogeneic, Open Approach |
|  |  |  | procedure | UMLS:ICD10PCS:0TY10Z1 | Transplantation of Left Kidney, Syngeneic, Open Approach |
|  |  |  | procedure | UMLS:ICD10PCS:0TY10Z2 | Transplantation of Left Kidney, Zooplastic, Open Approach |
|  |  |  | procedure | UMLS:ICD10PCS:0TY00Z1 | Transplantation of Right Kidney, Syngeneic, Open Approach |
|  |  |  | procedure | UMLS:ICD10PCS:0TY00Z2 | Transplantation of Right Kidney, Zooplastic, Open Approach |
|  |  |  | procedure | UMLS:ICD10PCS:0TY0 | Urinary System / Transplantation / Kidney, Right |
|  |  |  | procedure | UMLS:ICD10PCS:0TY1 | Urinary System / Transplantation / Kidney, Left |
|  | date constraint | | The terms in this group occurred between Jan 1, 2015 and Jan 1, 2022 | | |
| Group 2 | | | | | |
|  | **Group 2A Kidney transplant** | | | | |
|  | must have | any of | diagnosis | UMLS:ICD10CM:Z94.0 | Kidney transplant status |
|  |  |  | procedure | UMLS:ICD10PCS:0TY00Z0 | Transplantation of Right Kidney, Allogeneic, Open Approach |
|  |  |  | procedure | UMLS:ICD10PCS:0TY10Z0 | Transplantation of Left Kidney, Allogeneic, Open Approach |
|  |  |  | procedure | UMLS:ICD10PCS:0TY10Z1 | Transplantation of Left Kidney, Syngeneic, Open Approach |
|  |  |  | procedure | UMLS:ICD10PCS:0TY10Z2 | Transplantation of Left Kidney, Zooplastic, Open Approach |
|  |  |  | procedure | UMLS:ICD10PCS:0TY00Z1 | Transplantation of Right Kidney, Syngeneic, Open Approach |
|  |  |  | procedure | UMLS:ICD10PCS:0TY00Z2 | Transplantation of Right Kidney, Zooplastic, Open Approach |
|  |  |  | procedure | UMLS:ICD10PCS:0TY0 | Urinary System / Transplantation / Kidney, Right |
|  |  |  | procedure | UMLS:ICD10PCS:0TY1 | Urinary System / Transplantation / Kidney, Left |
|  | date constraint | | The terms in this group occurred at any time | | |
|  | event relationship | | Any instance of noDialysis, Death, SGLT2i occurred within 1 day and 999 days after the first instance of Kidney transplant | | |
|  | **Group 2B noDialysis, Death, SGLT2i** | | | | |
|  | cannot have |  | procedure | UMLS:CPT:1012740 | Dialysis Services and Procedures |
|  |  | or | procedure | UMLS:CPT:90945 | Dialysis procedure other than hemodialysis (eg, peritoneal dialysis, hemofiltration, or other continuous renal replacement therapies), with single evaluation by a physician or other qualified health care professional |
|  |  | or | procedure | UMLS:CPT:90947 | Dialysis procedure other than hemodialysis (eg, peritoneal dialysis, hemofiltration, or other continuous renal replacement therapies) requiring repeated evaluations by a physician or other qualified health care professional, with or without substantial revision of dialysis prescription |
|  |  | or | procedure | UMLS:CPT:1012752 | Hemodialysis Procedures |
|  |  | or | procedure | UMLS:SNOMED:302497006 | Hemodialysis |
|  |  | or | procedure | UMLS:CPT:90937 | Hemodialysis procedure requiring repeated evaluation(s) with or without substantial revision of dialysis prescription |
|  |  | or | procedure | UMLS:CPT:90935 | Hemodialysis procedure with single evaluation by a physician or other qualified health care professional |
|  |  | or | demographics | Deceased | Deceased |
|  |  | or | diagnosis | UMLS:ICD10CM:R99 | Ill-defined and unknown cause of mortality |
|  |  | or | medication | NLM:ATC:A10BK | Sodium-glucose co-transporter 2 (SGLT2) inhibitors |
| Group 3 | | | | | |
|  | **Group 3A Kidney transplant** | | | | |
|  | must have | any of | diagnosis | UMLS:ICD10CM:Z94.0 | Kidney transplant status |
|  |  |  | procedure | UMLS:ICD10PCS:0TY00Z0 | Transplantation of Right Kidney, Allogeneic, Open Approach |
|  |  |  | procedure | UMLS:ICD10PCS:0TY10Z0 | Transplantation of Left Kidney, Allogeneic, Open Approach |
|  |  |  | procedure | UMLS:ICD10PCS:0TY10Z1 | Transplantation of Left Kidney, Syngeneic, Open Approach |
|  |  |  | procedure | UMLS:ICD10PCS:0TY10Z2 | Transplantation of Left Kidney, Zooplastic, Open Approach |
|  |  |  | procedure | UMLS:ICD10PCS:0TY00Z1 | Transplantation of Right Kidney, Syngeneic, Open Approach |
|  |  |  | procedure | UMLS:ICD10PCS:0TY00Z2 | Transplantation of Right Kidney, Zooplastic, Open Approach |
|  |  |  | procedure | UMLS:ICD10PCS:0TY0 | Urinary System / Transplantation / Kidney, Right |
|  |  |  | procedure | UMLS:ICD10PCS:0TY1 | Urinary System / Transplantation / Kidney, Left |
|  | date constraint | | The terms in this group occurred at any time | | |
|  | event relationship | | Any instance of SGLT2i occurred within 1000 days and 3006 days after the first instance of Kidney transplant | | |
|  | **Group 3B SGLT2i** | | | | |
|  | cannot have |  | medication | NLM:ATC:A10BK | Sodium-glucose co-transporter 2 (SGLT2) inhibitors |
| Group 4 | | | | | |
|  | **Group 4A Kidney transplant** | | | | |
|  | must have | any of | diagnosis | UMLS:ICD10CM:Z94.0 | Kidney transplant status |
|  |  |  | procedure | UMLS:ICD10PCS:0TY00Z0 | Transplantation of Right Kidney, Allogeneic, Open Approach |
|  |  |  | procedure | UMLS:ICD10PCS:0TY10Z0 | Transplantation of Left Kidney, Allogeneic, Open Approach |
|  |  |  | procedure | UMLS:ICD10PCS:0TY10Z1 | Transplantation of Left Kidney, Syngeneic, Open Approach |
|  |  |  | procedure | UMLS:ICD10PCS:0TY10Z2 | Transplantation of Left Kidney, Zooplastic, Open Approach |
|  |  |  | procedure | UMLS:ICD10PCS:0TY00Z1 | Transplantation of Right Kidney, Syngeneic, Open Approach |
|  |  |  | procedure | UMLS:ICD10PCS:0TY00Z2 | Transplantation of Right Kidney, Zooplastic, Open Approach |
|  |  |  | procedure | UMLS:ICD10PCS:0TY0 | Urinary System / Transplantation / Kidney, Right |
|  |  |  | procedure | UMLS:ICD10PCS:0TY1 | Urinary System / Transplantation / Kidney, Left |
|  | date constraint | | The terms in this group occurred at any time | | |
|  | event relationship | | Any instance of noSGLT2i occurred at least 1 day before the first instance of Kidney transplant | | |
|  | **Group 4B noSGLT2i** | | | | |
|  | cannot have |  | medication | NLM:ATC:A10BK | Sodium-glucose co-transporter 2 (SGLT2) inhibitors |
| Group 5 | | | | | |
|  | **Group 5A Kidney transplant** | | | | |
|  | must have | any of | diagnosis | UMLS:ICD10CM:Z94.0 | Kidney transplant status |
|  |  |  | procedure | UMLS:ICD10PCS:0TY00Z0 | Transplantation of Right Kidney, Allogeneic, Open Approach |
|  |  |  | procedure | UMLS:ICD10PCS:0TY10Z0 | Transplantation of Left Kidney, Allogeneic, Open Approach |
|  |  |  | procedure | UMLS:ICD10PCS:0TY10Z1 | Transplantation of Left Kidney, Syngeneic, Open Approach |
|  |  |  | procedure | UMLS:ICD10PCS:0TY10Z2 | Transplantation of Left Kidney, Zooplastic, Open Approach |
|  |  |  | procedure | UMLS:ICD10PCS:0TY00Z1 | Transplantation of Right Kidney, Syngeneic, Open Approach |
|  |  |  | procedure | UMLS:ICD10PCS:0TY00Z2 | Transplantation of Right Kidney, Zooplastic, Open Approach |
|  |  |  | procedure | UMLS:ICD10PCS:0TY0 | Urinary System / Transplantation / Kidney, Right |
|  |  |  | procedure | UMLS:ICD10PCS:0TY1 | Urinary System / Transplantation / Kidney, Left |
|  | date constraint | | The terms in this group occurred at any time | | |
|  | event relationship | | Any instance of Visit occurred at least 1181 days after the first instance of Kidney transplant | | |
|  | **Group 5B Visit** | | | | |
|  | must have |  | visit | TNX:Visit | Visit |
| Group 6 | | | | | |
|  | **Group 6A Kidney transplant** | | | | |
|  | must have | any of | diagnosis | UMLS:ICD10CM:Z94.0 | Kidney transplant status |
|  |  |  | procedure | UMLS:ICD10PCS:0TY00Z0 | Transplantation of Right Kidney, Allogeneic, Open Approach |
|  |  |  | procedure | UMLS:ICD10PCS:0TY10Z0 | Transplantation of Left Kidney, Allogeneic, Open Approach |
|  |  |  | procedure | UMLS:ICD10PCS:0TY10Z1 | Transplantation of Left Kidney, Syngeneic, Open Approach |
|  |  |  | procedure | UMLS:ICD10PCS:0TY10Z2 | Transplantation of Left Kidney, Zooplastic, Open Approach |
|  |  |  | procedure | UMLS:ICD10PCS:0TY00Z1 | Transplantation of Right Kidney, Syngeneic, Open Approach |
|  |  |  | procedure | UMLS:ICD10PCS:0TY00Z2 | Transplantation of Right Kidney, Zooplastic, Open Approach |
|  |  |  | procedure | UMLS:ICD10PCS:0TY0 | Urinary System / Transplantation / Kidney, Right |
|  |  |  | procedure | UMLS:ICD10PCS:0TY1 | Urinary System / Transplantation / Kidney, Left |
|  | date constraint | | The terms in this group occurred at any time | | |
|  | event relationship | | Any instance of <30 BMI occurred within 1 day and 999 days after the first instance of Kidney transplant | | |
|  | **Group 6B <30 BMI** | | | | |
|  | must have |  | laboratory | TNX:9083 | BMI (between 0.00 and 30.00 kg/m2) |

##### **Table S47. Code definitions and selection criteria for SGLT2i users – ≥30 BMI.**

| Ungrouped terms | | | | | |
| --- | --- | --- | --- | --- | --- |
|  | must have |  | demographics | Age | Age (at least 18 years (most recent occurrence)) |
|  | cannot have |  | diagnosis | UMLS:ICD10CM:E08-E13 | Diabetes mellitus |
| Group 1 | | | | | |
|  | **Kidney transplant** | | | | |
|  | must have | any of | diagnosis | UMLS:ICD10CM:Z94.0 | Kidney transplant status |
|  |  |  | procedure | UMLS:ICD10PCS:0TY00Z0 | Transplantation of Right Kidney, Allogeneic, Open Approach |
|  |  |  | procedure | UMLS:ICD10PCS:0TY10Z0 | Transplantation of Left Kidney, Allogeneic, Open Approach |
|  |  |  | procedure | UMLS:ICD10PCS:0TY10Z1 | Transplantation of Left Kidney, Syngeneic, Open Approach |
|  |  |  | procedure | UMLS:ICD10PCS:0TY10Z2 | Transplantation of Left Kidney, Zooplastic, Open Approach |
|  |  |  | procedure | UMLS:ICD10PCS:0TY00Z1 | Transplantation of Right Kidney, Syngeneic, Open Approach |
|  |  |  | procedure | UMLS:ICD10PCS:0TY00Z2 | Transplantation of Right Kidney, Zooplastic, Open Approach |
|  |  |  | procedure | UMLS:ICD10PCS:0TY0 | Urinary System / Transplantation / Kidney, Right |
|  |  |  | procedure | UMLS:ICD10PCS:0TY1 | Urinary System / Transplantation / Kidney, Left |
|  | date constraint | | The terms in this group occurred between Jan 1, 2015 and Jan 1, 2022 | | |
| Group 2 | | | | | |
|  | **Group 2A Kidney transplant** | | | | |
|  | must have | any of | diagnosis | UMLS:ICD10CM:Z94.0 | Kidney transplant status |
|  |  |  | procedure | UMLS:ICD10PCS:0TY00Z0 | Transplantation of Right Kidney, Allogeneic, Open Approach |
|  |  |  | procedure | UMLS:ICD10PCS:0TY10Z0 | Transplantation of Left Kidney, Allogeneic, Open Approach |
|  |  |  | procedure | UMLS:ICD10PCS:0TY10Z1 | Transplantation of Left Kidney, Syngeneic, Open Approach |
|  |  |  | procedure | UMLS:ICD10PCS:0TY10Z2 | Transplantation of Left Kidney, Zooplastic, Open Approach |
|  |  |  | procedure | UMLS:ICD10PCS:0TY00Z1 | Transplantation of Right Kidney, Syngeneic, Open Approach |
|  |  |  | procedure | UMLS:ICD10PCS:0TY00Z2 | Transplantation of Right Kidney, Zooplastic, Open Approach |
|  |  |  | procedure | UMLS:ICD10PCS:0TY0 | Urinary System / Transplantation / Kidney, Right |
|  |  |  | procedure | UMLS:ICD10PCS:0TY1 | Urinary System / Transplantation / Kidney, Left |
|  | date constraint | | The terms in this group occurred at any time | | |
|  | event relationship | | Any instance of noDialysis, Death, SGLT2i occurred within 1 day and 999 days after the first instance of Kidney transplant | | |
|  | **Group 2B noDialysis, Death, SGLT2i** | | | | |
|  | cannot have |  | procedure | UMLS:CPT:1012740 | Dialysis Services and Procedures |
|  |  | or | procedure | UMLS:CPT:90945 | Dialysis procedure other than hemodialysis (eg, peritoneal dialysis, hemofiltration, or other continuous renal replacement therapies), with single evaluation by a physician or other qualified health care professional |
|  |  | or | procedure | UMLS:CPT:90947 | Dialysis procedure other than hemodialysis (eg, peritoneal dialysis, hemofiltration, or other continuous renal replacement therapies) requiring repeated evaluations by a physician or other qualified health care professional, with or without substantial revision of dialysis prescription |
|  |  | or | procedure | UMLS:CPT:1012752 | Hemodialysis Procedures |
|  |  | or | procedure | UMLS:SNOMED:302497006 | Hemodialysis |
|  |  | or | procedure | UMLS:CPT:90937 | Hemodialysis procedure requiring repeated evaluation(s) with or without substantial revision of dialysis prescription |
|  |  | or | procedure | UMLS:CPT:90935 | Hemodialysis procedure with single evaluation by a physician or other qualified health care professional |
|  |  | or | demographics | Deceased | Deceased |
|  |  | or | diagnosis | UMLS:ICD10CM:R99 | Ill-defined and unknown cause of mortality |
|  |  | or | medication | NLM:ATC:A10BK | Sodium-glucose co-transporter 2 (SGLT2) inhibitors |
| Group 3 | | | | | |
|  | **Group 3A Kidney transplant** | | | | |
|  | must have | any of | diagnosis | UMLS:ICD10CM:Z94.0 | Kidney transplant status |
|  |  |  | procedure | UMLS:ICD10PCS:0TY00Z0 | Transplantation of Right Kidney, Allogeneic, Open Approach |
|  |  |  | procedure | UMLS:ICD10PCS:0TY10Z0 | Transplantation of Left Kidney, Allogeneic, Open Approach |
|  |  |  | procedure | UMLS:ICD10PCS:0TY10Z1 | Transplantation of Left Kidney, Syngeneic, Open Approach |
|  |  |  | procedure | UMLS:ICD10PCS:0TY10Z2 | Transplantation of Left Kidney, Zooplastic, Open Approach |
|  |  |  | procedure | UMLS:ICD10PCS:0TY00Z1 | Transplantation of Right Kidney, Syngeneic, Open Approach |
|  |  |  | procedure | UMLS:ICD10PCS:0TY00Z2 | Transplantation of Right Kidney, Zooplastic, Open Approach |
|  |  |  | procedure | UMLS:ICD10PCS:0TY0 | Urinary System / Transplantation / Kidney, Right |
|  |  |  | procedure | UMLS:ICD10PCS:0TY1 | Urinary System / Transplantation / Kidney, Left |
|  | date constraint | | The terms in this group occurred at any time | | |
|  | event relationship | | Any instance of SGLT2i occurred at least 1000 days after the first instance of Kidney transplant | | |
|  | **Group 3B SGLT2i** | | | | |
|  | must have |  | medication | NLM:ATC:A10BK | Sodium-glucose co-transporter 2 (SGLT2) inhibitors |
| Group 4 | | | | | |
|  | **Group 4A Kidney transplant** | | | | |
|  | must have | any of | diagnosis | UMLS:ICD10CM:Z94.0 | Kidney transplant status |
|  |  |  | procedure | UMLS:ICD10PCS:0TY00Z0 | Transplantation of Right Kidney, Allogeneic, Open Approach |
|  |  |  | procedure | UMLS:ICD10PCS:0TY10Z0 | Transplantation of Left Kidney, Allogeneic, Open Approach |
|  |  |  | procedure | UMLS:ICD10PCS:0TY10Z1 | Transplantation of Left Kidney, Syngeneic, Open Approach |
|  |  |  | procedure | UMLS:ICD10PCS:0TY10Z2 | Transplantation of Left Kidney, Zooplastic, Open Approach |
|  |  |  | procedure | UMLS:ICD10PCS:0TY00Z1 | Transplantation of Right Kidney, Syngeneic, Open Approach |
|  |  |  | procedure | UMLS:ICD10PCS:0TY00Z2 | Transplantation of Right Kidney, Zooplastic, Open Approach |
|  |  |  | procedure | UMLS:ICD10PCS:0TY0 | Urinary System / Transplantation / Kidney, Right |
|  |  |  | procedure | UMLS:ICD10PCS:0TY1 | Urinary System / Transplantation / Kidney, Left |
|  | date constraint | | The terms in this group occurred at any time | | |
|  | event relationship | | Any instance of noSGLT2i occurred at least 1 day before the first instance of Kidney transplant | | |
|  | **Group 4B noSGLT2i** | | | | |
|  | cannot have |  | medication | NLM:ATC:A10BK | Sodium-glucose co-transporter 2 (SGLT2) inhibitors |
| Group 5 | | | | | |
|  | **Group 5A Kidney transplant** | | | | |
|  | must have | any of | diagnosis | UMLS:ICD10CM:Z94.0 | Kidney transplant status |
|  |  |  | procedure | UMLS:ICD10PCS:0TY00Z0 | Transplantation of Right Kidney, Allogeneic, Open Approach |
|  |  |  | procedure | UMLS:ICD10PCS:0TY10Z0 | Transplantation of Left Kidney, Allogeneic, Open Approach |
|  |  |  | procedure | UMLS:ICD10PCS:0TY10Z1 | Transplantation of Left Kidney, Syngeneic, Open Approach |
|  |  |  | procedure | UMLS:ICD10PCS:0TY10Z2 | Transplantation of Left Kidney, Zooplastic, Open Approach |
|  |  |  | procedure | UMLS:ICD10PCS:0TY00Z1 | Transplantation of Right Kidney, Syngeneic, Open Approach |
|  |  |  | procedure | UMLS:ICD10PCS:0TY00Z2 | Transplantation of Right Kidney, Zooplastic, Open Approach |
|  |  |  | procedure | UMLS:ICD10PCS:0TY0 | Urinary System / Transplantation / Kidney, Right |
|  |  |  | procedure | UMLS:ICD10PCS:0TY1 | Urinary System / Transplantation / Kidney, Left |
|  | date constraint | | The terms in this group occurred at any time | | |
|  | event relationship | | Any instance of Visit occurred at least 1181 days after the first instance of Kidney transplant | | |
|  | **Group 5B Visit** | | | | |
|  | must have |  | visit | TNX:Visit | Visit |
| Group 6 | | | | | |
|  | **Group 6A Kidney transplant** | | | | |
|  | must have | any of | diagnosis | UMLS:ICD10CM:Z94.0 | Kidney transplant status |
|  |  |  | procedure | UMLS:ICD10PCS:0TY00Z0 | Transplantation of Right Kidney, Allogeneic, Open Approach |
|  |  |  | procedure | UMLS:ICD10PCS:0TY10Z0 | Transplantation of Left Kidney, Allogeneic, Open Approach |
|  |  |  | procedure | UMLS:ICD10PCS:0TY10Z1 | Transplantation of Left Kidney, Syngeneic, Open Approach |
|  |  |  | procedure | UMLS:ICD10PCS:0TY10Z2 | Transplantation of Left Kidney, Zooplastic, Open Approach |
|  |  |  | procedure | UMLS:ICD10PCS:0TY00Z1 | Transplantation of Right Kidney, Syngeneic, Open Approach |
|  |  |  | procedure | UMLS:ICD10PCS:0TY00Z2 | Transplantation of Right Kidney, Zooplastic, Open Approach |
|  |  |  | procedure | UMLS:ICD10PCS:0TY0 | Urinary System / Transplantation / Kidney, Right |
|  |  |  | procedure | UMLS:ICD10PCS:0TY1 | Urinary System / Transplantation / Kidney, Left |
|  | date constraint | | The terms in this group occurred at any time | | |
|  | event relationship | | Any instance of >29 BMI occurred within 1 day and 999 days after the first instance of Kidney transplant | | |
|  | **Group 6B >29 BMI** | | | | |
|  | must have |  | laboratory | TNX:9083 | BMI (at least 30.00 kg/m2) |

##### **Table S48. Code definitions and selection criteria for SGLT2i non-users – ≥30 BMI.**

| Ungrouped terms | | | | | |
| --- | --- | --- | --- | --- | --- |
|  | must have |  | demographics | Age | Age (at least 18 years (most recent occurrence)) |
|  | cannot have |  | diagnosis | UMLS:ICD10CM:E08-E13 | Diabetes mellitus |
| Group 1 | | | | | |
|  | **Kidney transplant** | | | | |
|  | must have | any of | diagnosis | UMLS:ICD10CM:Z94.0 | Kidney transplant status |
|  |  |  | procedure | UMLS:ICD10PCS:0TY00Z0 | Transplantation of Right Kidney, Allogeneic, Open Approach |
|  |  |  | procedure | UMLS:ICD10PCS:0TY10Z0 | Transplantation of Left Kidney, Allogeneic, Open Approach |
|  |  |  | procedure | UMLS:ICD10PCS:0TY10Z1 | Transplantation of Left Kidney, Syngeneic, Open Approach |
|  |  |  | procedure | UMLS:ICD10PCS:0TY10Z2 | Transplantation of Left Kidney, Zooplastic, Open Approach |
|  |  |  | procedure | UMLS:ICD10PCS:0TY00Z1 | Transplantation of Right Kidney, Syngeneic, Open Approach |
|  |  |  | procedure | UMLS:ICD10PCS:0TY00Z2 | Transplantation of Right Kidney, Zooplastic, Open Approach |
|  |  |  | procedure | UMLS:ICD10PCS:0TY0 | Urinary System / Transplantation / Kidney, Right |
|  |  |  | procedure | UMLS:ICD10PCS:0TY1 | Urinary System / Transplantation / Kidney, Left |
|  | date constraint | | The terms in this group occurred between Jan 1, 2015 and Jan 1, 2022 | | |
| Group 2 | | | | | |
|  | **Group 2A Kidney transplant** | | | | |
|  | must have | any of | diagnosis | UMLS:ICD10CM:Z94.0 | Kidney transplant status |
|  |  |  | procedure | UMLS:ICD10PCS:0TY00Z0 | Transplantation of Right Kidney, Allogeneic, Open Approach |
|  |  |  | procedure | UMLS:ICD10PCS:0TY10Z0 | Transplantation of Left Kidney, Allogeneic, Open Approach |
|  |  |  | procedure | UMLS:ICD10PCS:0TY10Z1 | Transplantation of Left Kidney, Syngeneic, Open Approach |
|  |  |  | procedure | UMLS:ICD10PCS:0TY10Z2 | Transplantation of Left Kidney, Zooplastic, Open Approach |
|  |  |  | procedure | UMLS:ICD10PCS:0TY00Z1 | Transplantation of Right Kidney, Syngeneic, Open Approach |
|  |  |  | procedure | UMLS:ICD10PCS:0TY00Z2 | Transplantation of Right Kidney, Zooplastic, Open Approach |
|  |  |  | procedure | UMLS:ICD10PCS:0TY0 | Urinary System / Transplantation / Kidney, Right |
|  |  |  | procedure | UMLS:ICD10PCS:0TY1 | Urinary System / Transplantation / Kidney, Left |
|  | date constraint | | The terms in this group occurred at any time | | |
|  | event relationship | | Any instance of noDialysis, Death, SGLT2i occurred within 1 day and 999 days after the first instance of Kidney transplant | | |
|  | **Group 2B noDialysis, Death, SGLT2i** | | | | |
|  | cannot have |  | procedure | UMLS:CPT:1012740 | Dialysis Services and Procedures |
|  |  | or | procedure | UMLS:CPT:90945 | Dialysis procedure other than hemodialysis (eg, peritoneal dialysis, hemofiltration, or other continuous renal replacement therapies), with single evaluation by a physician or other qualified health care professional |
|  |  | or | procedure | UMLS:CPT:90947 | Dialysis procedure other than hemodialysis (eg, peritoneal dialysis, hemofiltration, or other continuous renal replacement therapies) requiring repeated evaluations by a physician or other qualified health care professional, with or without substantial revision of dialysis prescription |
|  |  | or | procedure | UMLS:CPT:1012752 | Hemodialysis Procedures |
|  |  | or | procedure | UMLS:SNOMED:302497006 | Hemodialysis |
|  |  | or | procedure | UMLS:CPT:90937 | Hemodialysis procedure requiring repeated evaluation(s) with or without substantial revision of dialysis prescription |
|  |  | or | procedure | UMLS:CPT:90935 | Hemodialysis procedure with single evaluation by a physician or other qualified health care professional |
|  |  | or | demographics | Deceased | Deceased |
|  |  | or | diagnosis | UMLS:ICD10CM:R99 | Ill-defined and unknown cause of mortality |
|  |  | or | medication | NLM:ATC:A10BK | Sodium-glucose co-transporter 2 (SGLT2) inhibitors |
| Group 3 | | | | | |
|  | **Group 3A Kidney transplant** | | | | |
|  | must have | any of | diagnosis | UMLS:ICD10CM:Z94.0 | Kidney transplant status |
|  |  |  | procedure | UMLS:ICD10PCS:0TY00Z0 | Transplantation of Right Kidney, Allogeneic, Open Approach |
|  |  |  | procedure | UMLS:ICD10PCS:0TY10Z0 | Transplantation of Left Kidney, Allogeneic, Open Approach |
|  |  |  | procedure | UMLS:ICD10PCS:0TY10Z1 | Transplantation of Left Kidney, Syngeneic, Open Approach |
|  |  |  | procedure | UMLS:ICD10PCS:0TY10Z2 | Transplantation of Left Kidney, Zooplastic, Open Approach |
|  |  |  | procedure | UMLS:ICD10PCS:0TY00Z1 | Transplantation of Right Kidney, Syngeneic, Open Approach |
|  |  |  | procedure | UMLS:ICD10PCS:0TY00Z2 | Transplantation of Right Kidney, Zooplastic, Open Approach |
|  |  |  | procedure | UMLS:ICD10PCS:0TY0 | Urinary System / Transplantation / Kidney, Right |
|  |  |  | procedure | UMLS:ICD10PCS:0TY1 | Urinary System / Transplantation / Kidney, Left |
|  | date constraint | | The terms in this group occurred at any time | | |
|  | event relationship | | Any instance of noSGLT2i occurred within 1000 days and 3006 days after the first instance of Kidney transplant | | |
|  | **Group 3B noSGLT2i** | | | | |
|  | cannot have |  | medication | NLM:ATC:A10BK | Sodium-glucose co-transporter 2 (SGLT2) inhibitors |
| Group 4 | | | | | |
|  | **Group 4A Kidney transplant** | | | | |
|  | must have | any of | diagnosis | UMLS:ICD10CM:Z94.0 | Kidney transplant status |
|  |  |  | procedure | UMLS:ICD10PCS:0TY00Z0 | Transplantation of Right Kidney, Allogeneic, Open Approach |
|  |  |  | procedure | UMLS:ICD10PCS:0TY10Z0 | Transplantation of Left Kidney, Allogeneic, Open Approach |
|  |  |  | procedure | UMLS:ICD10PCS:0TY10Z1 | Transplantation of Left Kidney, Syngeneic, Open Approach |
|  |  |  | procedure | UMLS:ICD10PCS:0TY10Z2 | Transplantation of Left Kidney, Zooplastic, Open Approach |
|  |  |  | procedure | UMLS:ICD10PCS:0TY00Z1 | Transplantation of Right Kidney, Syngeneic, Open Approach |
|  |  |  | procedure | UMLS:ICD10PCS:0TY00Z2 | Transplantation of Right Kidney, Zooplastic, Open Approach |
|  |  |  | procedure | UMLS:ICD10PCS:0TY0 | Urinary System / Transplantation / Kidney, Right |
|  |  |  | procedure | UMLS:ICD10PCS:0TY1 | Urinary System / Transplantation / Kidney, Left |
|  | date constraint | | The terms in this group occurred at any time | | |
|  | event relationship | | Any instance of noSGLT2i occurred at least 1 day before the first instance of Kidney transplant | | |
|  | **Group 4B noSGLT2i** | | | | |
|  | cannot have |  | medication | NLM:ATC:A10BK | Sodium-glucose co-transporter 2 (SGLT2) inhibitors |
| Group 5 | | | | | |
|  | **Group 5A Kidney transplant** | | | | |
|  | must have | any of | diagnosis | UMLS:ICD10CM:Z94.0 | Kidney transplant status |
|  |  |  | procedure | UMLS:ICD10PCS:0TY00Z0 | Transplantation of Right Kidney, Allogeneic, Open Approach |
|  |  |  | procedure | UMLS:ICD10PCS:0TY10Z0 | Transplantation of Left Kidney, Allogeneic, Open Approach |
|  |  |  | procedure | UMLS:ICD10PCS:0TY10Z1 | Transplantation of Left Kidney, Syngeneic, Open Approach |
|  |  |  | procedure | UMLS:ICD10PCS:0TY10Z2 | Transplantation of Left Kidney, Zooplastic, Open Approach |
|  |  |  | procedure | UMLS:ICD10PCS:0TY00Z1 | Transplantation of Right Kidney, Syngeneic, Open Approach |
|  |  |  | procedure | UMLS:ICD10PCS:0TY00Z2 | Transplantation of Right Kidney, Zooplastic, Open Approach |
|  |  |  | procedure | UMLS:ICD10PCS:0TY0 | Urinary System / Transplantation / Kidney, Right |
|  |  |  | procedure | UMLS:ICD10PCS:0TY1 | Urinary System / Transplantation / Kidney, Left |
|  | date constraint | | The terms in this group occurred at any time | | |
|  | event relationship | | Any instance of Visit occurred at least 1181 days after the first instance of Kidney transplant | | |
|  | **Group 5B Visit** | | | | |
|  | must have |  | visit | TNX:Visit | Visit |
| Group 6 | | | | | |
|  | **Group 6A Kidney transplant** | | | | |
|  | must have | any of | diagnosis | UMLS:ICD10CM:Z94.0 | Kidney transplant status |
|  |  |  | procedure | UMLS:ICD10PCS:0TY00Z0 | Transplantation of Right Kidney, Allogeneic, Open Approach |
|  |  |  | procedure | UMLS:ICD10PCS:0TY10Z0 | Transplantation of Left Kidney, Allogeneic, Open Approach |
|  |  |  | procedure | UMLS:ICD10PCS:0TY10Z1 | Transplantation of Left Kidney, Syngeneic, Open Approach |
|  |  |  | procedure | UMLS:ICD10PCS:0TY10Z2 | Transplantation of Left Kidney, Zooplastic, Open Approach |
|  |  |  | procedure | UMLS:ICD10PCS:0TY00Z1 | Transplantation of Right Kidney, Syngeneic, Open Approach |
|  |  |  | procedure | UMLS:ICD10PCS:0TY00Z2 | Transplantation of Right Kidney, Zooplastic, Open Approach |
|  |  |  | procedure | UMLS:ICD10PCS:0TY0 | Urinary System / Transplantation / Kidney, Right |
|  |  |  | procedure | UMLS:ICD10PCS:0TY1 | Urinary System / Transplantation / Kidney, Left |
|  | date constraint | | The terms in this group occurred at any time | | |
|  | event relationship | | Any instance of >29 BMI occurred within 1 day and 999 days after the first instance of Kidney transplant | | |
|  | **Group 6B >29 BMI** | | | | |
|  | must have |  | laboratory | TNX:9083 | BMI (at least 30.00 kg/m2) |

##### **Table S49. Code definitions and selection criteria for SGLT2i users – <15 eGFR.**

| Ungrouped terms | | | | | |
| --- | --- | --- | --- | --- | --- |
|  | must have |  | demographics | Age | Age (at least 18 years (most recent occurrence)) |
|  | cannot have |  | diagnosis | UMLS:ICD10CM:E08-E13 | Diabetes mellitus |
| Group 1 | | | | | |
|  | **Kidney transplant** | | | | |
|  | must have | any of | diagnosis | UMLS:ICD10CM:Z94.0 | Kidney transplant status |
|  |  |  | procedure | UMLS:ICD10PCS:0TY00Z0 | Transplantation of Right Kidney, Allogeneic, Open Approach |
|  |  |  | procedure | UMLS:ICD10PCS:0TY10Z0 | Transplantation of Left Kidney, Allogeneic, Open Approach |
|  |  |  | procedure | UMLS:ICD10PCS:0TY10Z1 | Transplantation of Left Kidney, Syngeneic, Open Approach |
|  |  |  | procedure | UMLS:ICD10PCS:0TY10Z2 | Transplantation of Left Kidney, Zooplastic, Open Approach |
|  |  |  | procedure | UMLS:ICD10PCS:0TY00Z1 | Transplantation of Right Kidney, Syngeneic, Open Approach |
|  |  |  | procedure | UMLS:ICD10PCS:0TY00Z2 | Transplantation of Right Kidney, Zooplastic, Open Approach |
|  |  |  | procedure | UMLS:ICD10PCS:0TY0 | Urinary System / Transplantation / Kidney, Right |
|  |  |  | procedure | UMLS:ICD10PCS:0TY1 | Urinary System / Transplantation / Kidney, Left |
|  | date constraint | | The terms in this group occurred between Jan 1, 2015 and Jan 1, 2022 | | |
| Group 2 | | | | | |
|  | **Group 2A Kidney transplant** | | | | |
|  | must have | any of | diagnosis | UMLS:ICD10CM:Z94.0 | Kidney transplant status |
|  |  |  | procedure | UMLS:ICD10PCS:0TY00Z0 | Transplantation of Right Kidney, Allogeneic, Open Approach |
|  |  |  | procedure | UMLS:ICD10PCS:0TY10Z0 | Transplantation of Left Kidney, Allogeneic, Open Approach |
|  |  |  | procedure | UMLS:ICD10PCS:0TY10Z1 | Transplantation of Left Kidney, Syngeneic, Open Approach |
|  |  |  | procedure | UMLS:ICD10PCS:0TY10Z2 | Transplantation of Left Kidney, Zooplastic, Open Approach |
|  |  |  | procedure | UMLS:ICD10PCS:0TY00Z1 | Transplantation of Right Kidney, Syngeneic, Open Approach |
|  |  |  | procedure | UMLS:ICD10PCS:0TY00Z2 | Transplantation of Right Kidney, Zooplastic, Open Approach |
|  |  |  | procedure | UMLS:ICD10PCS:0TY0 | Urinary System / Transplantation / Kidney, Right |
|  |  |  | procedure | UMLS:ICD10PCS:0TY1 | Urinary System / Transplantation / Kidney, Left |
|  | date constraint | | The terms in this group occurred at any time | | |
|  | event relationship | | Any instance of noDialysis, Death, SGLT2i occurred within 1 day and 999 days after the first instance of Kidney transplant | | |
|  | **Group 2B noDialysis, Death, SGLT2i** | | | | |
|  | cannot have |  | procedure | UMLS:CPT:1012740 | Dialysis Services and Procedures |
|  |  | or | procedure | UMLS:CPT:90945 | Dialysis procedure other than hemodialysis (eg, peritoneal dialysis, hemofiltration, or other continuous renal replacement therapies), with single evaluation by a physician or other qualified health care professional |
|  |  | or | procedure | UMLS:CPT:90947 | Dialysis procedure other than hemodialysis (eg, peritoneal dialysis, hemofiltration, or other continuous renal replacement therapies) requiring repeated evaluations by a physician or other qualified health care professional, with or without substantial revision of dialysis prescription |
|  |  | or | procedure | UMLS:CPT:1012752 | Hemodialysis Procedures |
|  |  | or | procedure | UMLS:SNOMED:302497006 | Hemodialysis |
|  |  | or | procedure | UMLS:CPT:90937 | Hemodialysis procedure requiring repeated evaluation(s) with or without substantial revision of dialysis prescription |
|  |  | or | procedure | UMLS:CPT:90935 | Hemodialysis procedure with single evaluation by a physician or other qualified health care professional |
|  |  | or | demographics | Deceased | Deceased |
|  |  | or | diagnosis | UMLS:ICD10CM:R99 | Ill-defined and unknown cause of mortality |
|  |  | or | medication | NLM:ATC:A10BK | Sodium-glucose co-transporter 2 (SGLT2) inhibitors |
| Group 3 | | | | | |
|  | **Group 3A Kidney transplant** | | | | |
|  | must have | any of | diagnosis | UMLS:ICD10CM:Z94.0 | Kidney transplant status |
|  |  |  | procedure | UMLS:ICD10PCS:0TY00Z0 | Transplantation of Right Kidney, Allogeneic, Open Approach |
|  |  |  | procedure | UMLS:ICD10PCS:0TY10Z0 | Transplantation of Left Kidney, Allogeneic, Open Approach |
|  |  |  | procedure | UMLS:ICD10PCS:0TY10Z1 | Transplantation of Left Kidney, Syngeneic, Open Approach |
|  |  |  | procedure | UMLS:ICD10PCS:0TY10Z2 | Transplantation of Left Kidney, Zooplastic, Open Approach |
|  |  |  | procedure | UMLS:ICD10PCS:0TY00Z1 | Transplantation of Right Kidney, Syngeneic, Open Approach |
|  |  |  | procedure | UMLS:ICD10PCS:0TY00Z2 | Transplantation of Right Kidney, Zooplastic, Open Approach |
|  |  |  | procedure | UMLS:ICD10PCS:0TY0 | Urinary System / Transplantation / Kidney, Right |
|  |  |  | procedure | UMLS:ICD10PCS:0TY1 | Urinary System / Transplantation / Kidney, Left |
|  | date constraint | | The terms in this group occurred at any time | | |
|  | event relationship | | Any instance of SGLT2i occurred at least 1000 days after the first instance of Kidney transplant | | |
|  | **Group 3B SGLT2i** | | | | |
|  | must have |  | medication | NLM:ATC:A10BK | Sodium-glucose co-transporter 2 (SGLT2) inhibitors |
| Group 4 | | | | | |
|  | **Group 4A Kidney transplant** | | | | |
|  | must have | any of | diagnosis | UMLS:ICD10CM:Z94.0 | Kidney transplant status |
|  |  |  | procedure | UMLS:ICD10PCS:0TY00Z0 | Transplantation of Right Kidney, Allogeneic, Open Approach |
|  |  |  | procedure | UMLS:ICD10PCS:0TY10Z0 | Transplantation of Left Kidney, Allogeneic, Open Approach |
|  |  |  | procedure | UMLS:ICD10PCS:0TY10Z1 | Transplantation of Left Kidney, Syngeneic, Open Approach |
|  |  |  | procedure | UMLS:ICD10PCS:0TY10Z2 | Transplantation of Left Kidney, Zooplastic, Open Approach |
|  |  |  | procedure | UMLS:ICD10PCS:0TY00Z1 | Transplantation of Right Kidney, Syngeneic, Open Approach |
|  |  |  | procedure | UMLS:ICD10PCS:0TY00Z2 | Transplantation of Right Kidney, Zooplastic, Open Approach |
|  |  |  | procedure | UMLS:ICD10PCS:0TY0 | Urinary System / Transplantation / Kidney, Right |
|  |  |  | procedure | UMLS:ICD10PCS:0TY1 | Urinary System / Transplantation / Kidney, Left |
|  | date constraint | | The terms in this group occurred at any time | | |
|  | event relationship | | Any instance of noSGLT2i occurred at least 1 day before the first instance of Kidney transplant | | |
|  | **Group 4B noSGLT2i** | | | | |
|  | cannot have |  | medication | NLM:ATC:A10BK | Sodium-glucose co-transporter 2 (SGLT2) inhibitors |
| Group 5 | | | | | |
|  | **Group 5A Kidney transplant** | | | | |
|  | must have | any of | diagnosis | UMLS:ICD10CM:Z94.0 | Kidney transplant status |
|  |  |  | procedure | UMLS:ICD10PCS:0TY00Z0 | Transplantation of Right Kidney, Allogeneic, Open Approach |
|  |  |  | procedure | UMLS:ICD10PCS:0TY10Z0 | Transplantation of Left Kidney, Allogeneic, Open Approach |
|  |  |  | procedure | UMLS:ICD10PCS:0TY10Z1 | Transplantation of Left Kidney, Syngeneic, Open Approach |
|  |  |  | procedure | UMLS:ICD10PCS:0TY10Z2 | Transplantation of Left Kidney, Zooplastic, Open Approach |
|  |  |  | procedure | UMLS:ICD10PCS:0TY00Z1 | Transplantation of Right Kidney, Syngeneic, Open Approach |
|  |  |  | procedure | UMLS:ICD10PCS:0TY00Z2 | Transplantation of Right Kidney, Zooplastic, Open Approach |
|  |  |  | procedure | UMLS:ICD10PCS:0TY0 | Urinary System / Transplantation / Kidney, Right |
|  |  |  | procedure | UMLS:ICD10PCS:0TY1 | Urinary System / Transplantation / Kidney, Left |
|  | date constraint | | The terms in this group occurred at any time | | |
|  | event relationship | | Any instance of Visit occurred at least 1181 days after the first instance of Kidney transplant | | |
|  | **Group 5B Visit** | | | | |
|  | must have |  | visit | TNX:Visit | Visit |
| Group 6 | | | | | |
|  | **Group 6A Kidney transplant** | | | | |
|  | must have | any of | diagnosis | UMLS:ICD10CM:Z94.0 | Kidney transplant status |
|  |  |  | procedure | UMLS:ICD10PCS:0TY00Z0 | Transplantation of Right Kidney, Allogeneic, Open Approach |
|  |  |  | procedure | UMLS:ICD10PCS:0TY10Z0 | Transplantation of Left Kidney, Allogeneic, Open Approach |
|  |  |  | procedure | UMLS:ICD10PCS:0TY10Z1 | Transplantation of Left Kidney, Syngeneic, Open Approach |
|  |  |  | procedure | UMLS:ICD10PCS:0TY10Z2 | Transplantation of Left Kidney, Zooplastic, Open Approach |
|  |  |  | procedure | UMLS:ICD10PCS:0TY00Z1 | Transplantation of Right Kidney, Syngeneic, Open Approach |
|  |  |  | procedure | UMLS:ICD10PCS:0TY00Z2 | Transplantation of Right Kidney, Zooplastic, Open Approach |
|  |  |  | procedure | UMLS:ICD10PCS:0TY0 | Urinary System / Transplantation / Kidney, Right |
|  |  |  | procedure | UMLS:ICD10PCS:0TY1 | Urinary System / Transplantation / Kidney, Left |
|  | date constraint | | The terms in this group occurred at any time | | |
|  | event relationship | | Any instance of <15 eGFR occurred within 1 day and 999 days after the first instance of Kidney transplant | | |
|  | **Group 6B <15 eGFR** | | | | |
|  | must have |  | laboratory | TNX:8001 | Glomerular filtration rate/1.73 sq M.predicted [Volume Rate/Area] in Serum, Plasma or Blood by Creatinine-based formula (MDRD) (between 0.00 and 15.00 mL/min/{1.73_m2}) |

##### **Table S50. Code definitions and selection criteria for SGLT2i non-users – <15 eGFR.**

| Ungrouped terms | | | | | |
| --- | --- | --- | --- | --- | --- |
|  | must have |  | demographics | Age | Age (at least 18 years (most recent occurrence)) |
|  | cannot have |  | diagnosis | UMLS:ICD10CM:E08-E13 | Diabetes mellitus |
| Group 1 | | | | | |
|  | **Kidney transplant** | | | | |
|  | must have | any of | diagnosis | UMLS:ICD10CM:Z94.0 | Kidney transplant status |
|  |  |  | procedure | UMLS:ICD10PCS:0TY00Z0 | Transplantation of Right Kidney, Allogeneic, Open Approach |
|  |  |  | procedure | UMLS:ICD10PCS:0TY10Z0 | Transplantation of Left Kidney, Allogeneic, Open Approach |
|  |  |  | procedure | UMLS:ICD10PCS:0TY10Z1 | Transplantation of Left Kidney, Syngeneic, Open Approach |
|  |  |  | procedure | UMLS:ICD10PCS:0TY10Z2 | Transplantation of Left Kidney, Zooplastic, Open Approach |
|  |  |  | procedure | UMLS:ICD10PCS:0TY00Z1 | Transplantation of Right Kidney, Syngeneic, Open Approach |
|  |  |  | procedure | UMLS:ICD10PCS:0TY00Z2 | Transplantation of Right Kidney, Zooplastic, Open Approach |
|  |  |  | procedure | UMLS:ICD10PCS:0TY0 | Urinary System / Transplantation / Kidney, Right |
|  |  |  | procedure | UMLS:ICD10PCS:0TY1 | Urinary System / Transplantation / Kidney, Left |
|  | date constraint | | The terms in this group occurred between Jan 1, 2015 and Jan 1, 2022 | | |
| Group 2 | | | | | |
|  | **Group 2A Kidney transplant** | | | | |
|  | must have | any of | diagnosis | UMLS:ICD10CM:Z94.0 | Kidney transplant status |
|  |  |  | procedure | UMLS:ICD10PCS:0TY00Z0 | Transplantation of Right Kidney, Allogeneic, Open Approach |
|  |  |  | procedure | UMLS:ICD10PCS:0TY10Z0 | Transplantation of Left Kidney, Allogeneic, Open Approach |
|  |  |  | procedure | UMLS:ICD10PCS:0TY10Z1 | Transplantation of Left Kidney, Syngeneic, Open Approach |
|  |  |  | procedure | UMLS:ICD10PCS:0TY10Z2 | Transplantation of Left Kidney, Zooplastic, Open Approach |
|  |  |  | procedure | UMLS:ICD10PCS:0TY00Z1 | Transplantation of Right Kidney, Syngeneic, Open Approach |
|  |  |  | procedure | UMLS:ICD10PCS:0TY00Z2 | Transplantation of Right Kidney, Zooplastic, Open Approach |
|  |  |  | procedure | UMLS:ICD10PCS:0TY0 | Urinary System / Transplantation / Kidney, Right |
|  |  |  | procedure | UMLS:ICD10PCS:0TY1 | Urinary System / Transplantation / Kidney, Left |
|  | date constraint | | The terms in this group occurred at any time | | |
|  | event relationship | | Any instance of noDialysis, Death, SGLT2i occurred within 1 day and 999 days after the first instance of Kidney transplant | | |
|  | **Group 2B noDialysis, Death, SGLT2i** | | | | |
|  | cannot have |  | procedure | UMLS:CPT:1012740 | Dialysis Services and Procedures |
|  |  | or | procedure | UMLS:CPT:90945 | Dialysis procedure other than hemodialysis (eg, peritoneal dialysis, hemofiltration, or other continuous renal replacement therapies), with single evaluation by a physician or other qualified health care professional |
|  |  | or | procedure | UMLS:CPT:90947 | Dialysis procedure other than hemodialysis (eg, peritoneal dialysis, hemofiltration, or other continuous renal replacement therapies) requiring repeated evaluations by a physician or other qualified health care professional, with or without substantial revision of dialysis prescription |
|  |  | or | procedure | UMLS:CPT:1012752 | Hemodialysis Procedures |
|  |  | or | procedure | UMLS:SNOMED:302497006 | Hemodialysis |
|  |  | or | procedure | UMLS:CPT:90937 | Hemodialysis procedure requiring repeated evaluation(s) with or without substantial revision of dialysis prescription |
|  |  | or | procedure | UMLS:CPT:90935 | Hemodialysis procedure with single evaluation by a physician or other qualified health care professional |
|  |  | or | demographics | Deceased | Deceased |
|  |  | or | diagnosis | UMLS:ICD10CM:R99 | Ill-defined and unknown cause of mortality |
|  |  | or | medication | NLM:ATC:A10BK | Sodium-glucose co-transporter 2 (SGLT2) inhibitors |
| Group 3 | | | | | |
|  | **Group 3A Kidney transplant** | | | | |
|  | must have | any of | diagnosis | UMLS:ICD10CM:Z94.0 | Kidney transplant status |
|  |  |  | procedure | UMLS:ICD10PCS:0TY00Z0 | Transplantation of Right Kidney, Allogeneic, Open Approach |
|  |  |  | procedure | UMLS:ICD10PCS:0TY10Z0 | Transplantation of Left Kidney, Allogeneic, Open Approach |
|  |  |  | procedure | UMLS:ICD10PCS:0TY10Z1 | Transplantation of Left Kidney, Syngeneic, Open Approach |
|  |  |  | procedure | UMLS:ICD10PCS:0TY10Z2 | Transplantation of Left Kidney, Zooplastic, Open Approach |
|  |  |  | procedure | UMLS:ICD10PCS:0TY00Z1 | Transplantation of Right Kidney, Syngeneic, Open Approach |
|  |  |  | procedure | UMLS:ICD10PCS:0TY00Z2 | Transplantation of Right Kidney, Zooplastic, Open Approach |
|  |  |  | procedure | UMLS:ICD10PCS:0TY0 | Urinary System / Transplantation / Kidney, Right |
|  |  |  | procedure | UMLS:ICD10PCS:0TY1 | Urinary System / Transplantation / Kidney, Left |
|  | date constraint | | The terms in this group occurred at any time | | |
|  | event relationship | | Any instance of SGLT2i occurred within 1000 days and 3006 days after the first instance of Kidney transplant | | |
|  | **Group 3B SGLT2i** | | | | |
|  | cannot have |  | medication | NLM:ATC:A10BK | Sodium-glucose co-transporter 2 (SGLT2) inhibitors |
| Group 4 | | | | | |
|  | **Group 4A Kidney transplant** | | | | |
|  | must have | any of | diagnosis | UMLS:ICD10CM:Z94.0 | Kidney transplant status |
|  |  |  | procedure | UMLS:ICD10PCS:0TY00Z0 | Transplantation of Right Kidney, Allogeneic, Open Approach |
|  |  |  | procedure | UMLS:ICD10PCS:0TY10Z0 | Transplantation of Left Kidney, Allogeneic, Open Approach |
|  |  |  | procedure | UMLS:ICD10PCS:0TY10Z1 | Transplantation of Left Kidney, Syngeneic, Open Approach |
|  |  |  | procedure | UMLS:ICD10PCS:0TY10Z2 | Transplantation of Left Kidney, Zooplastic, Open Approach |
|  |  |  | procedure | UMLS:ICD10PCS:0TY00Z1 | Transplantation of Right Kidney, Syngeneic, Open Approach |
|  |  |  | procedure | UMLS:ICD10PCS:0TY00Z2 | Transplantation of Right Kidney, Zooplastic, Open Approach |
|  |  |  | procedure | UMLS:ICD10PCS:0TY0 | Urinary System / Transplantation / Kidney, Right |
|  |  |  | procedure | UMLS:ICD10PCS:0TY1 | Urinary System / Transplantation / Kidney, Left |
|  | date constraint | | The terms in this group occurred at any time | | |
|  | event relationship | | Any instance of noSGLT2i occurred at least 1 day before the first instance of Kidney transplant | | |
|  | **Group 4B noSGLT2i** | | | | |
|  | cannot have |  | medication | NLM:ATC:A10BK | Sodium-glucose co-transporter 2 (SGLT2) inhibitors |
| Group 5 | | | | | |
|  | **Group 5A Kidney transplant** | | | | |
|  | must have | any of | diagnosis | UMLS:ICD10CM:Z94.0 | Kidney transplant status |
|  |  |  | procedure | UMLS:ICD10PCS:0TY00Z0 | Transplantation of Right Kidney, Allogeneic, Open Approach |
|  |  |  | procedure | UMLS:ICD10PCS:0TY10Z0 | Transplantation of Left Kidney, Allogeneic, Open Approach |
|  |  |  | procedure | UMLS:ICD10PCS:0TY10Z1 | Transplantation of Left Kidney, Syngeneic, Open Approach |
|  |  |  | procedure | UMLS:ICD10PCS:0TY10Z2 | Transplantation of Left Kidney, Zooplastic, Open Approach |
|  |  |  | procedure | UMLS:ICD10PCS:0TY00Z1 | Transplantation of Right Kidney, Syngeneic, Open Approach |
|  |  |  | procedure | UMLS:ICD10PCS:0TY00Z2 | Transplantation of Right Kidney, Zooplastic, Open Approach |
|  |  |  | procedure | UMLS:ICD10PCS:0TY0 | Urinary System / Transplantation / Kidney, Right |
|  |  |  | procedure | UMLS:ICD10PCS:0TY1 | Urinary System / Transplantation / Kidney, Left |
|  | date constraint | | The terms in this group occurred at any time | | |
|  | event relationship | | Any instance of Visit occurred at least 1181 days after the first instance of Kidney transplant | | |
|  | **Group 5B Visit** | | | | |
|  | must have |  | visit | TNX:Visit | Visit |
| Group 6 | | | | | |
|  | **Group 6A Kidney transplant** | | | | |
|  | must have | any of | diagnosis | UMLS:ICD10CM:Z94.0 | Kidney transplant status |
|  |  |  | procedure | UMLS:ICD10PCS:0TY00Z0 | Transplantation of Right Kidney, Allogeneic, Open Approach |
|  |  |  | procedure | UMLS:ICD10PCS:0TY10Z0 | Transplantation of Left Kidney, Allogeneic, Open Approach |
|  |  |  | procedure | UMLS:ICD10PCS:0TY10Z1 | Transplantation of Left Kidney, Syngeneic, Open Approach |
|  |  |  | procedure | UMLS:ICD10PCS:0TY10Z2 | Transplantation of Left Kidney, Zooplastic, Open Approach |
|  |  |  | procedure | UMLS:ICD10PCS:0TY00Z1 | Transplantation of Right Kidney, Syngeneic, Open Approach |
|  |  |  | procedure | UMLS:ICD10PCS:0TY00Z2 | Transplantation of Right Kidney, Zooplastic, Open Approach |
|  |  |  | procedure | UMLS:ICD10PCS:0TY0 | Urinary System / Transplantation / Kidney, Right |
|  |  |  | procedure | UMLS:ICD10PCS:0TY1 | Urinary System / Transplantation / Kidney, Left |
|  | date constraint | | The terms in this group occurred at any time | | |
|  | event relationship | | Any instance of <15 eGFR occurred within 1 day and 999 days after the first instance of Kidney transplant | | |
|  | **Group 6B <15 eGFR** | | | | |
|  | must have |  | laboratory | TNX:8001 | Glomerular filtration rate/1.73 sq M.predicted [Volume Rate/Area] in Serum, Plasma or Blood by Creatinine-based formula (MDRD) (between 0.00 and 15.00 mL/min/{1.73_m2}) |

##### **Table S51. Code definitions and selection criteria for SGLT2i users – 15-29 eGFR.**

| Ungrouped terms | | | | | |
| --- | --- | --- | --- | --- | --- |
|  | must have |  | demographics | Age | Age (at least 18 years (most recent occurrence)) |
|  | cannot have |  | diagnosis | UMLS:ICD10CM:E08-E13 | Diabetes mellitus |
| Group 1 | | | | | |
|  | **Kidney transplant** | | | | |
|  | must have | any of | diagnosis | UMLS:ICD10CM:Z94.0 | Kidney transplant status |
|  |  |  | procedure | UMLS:ICD10PCS:0TY00Z0 | Transplantation of Right Kidney, Allogeneic, Open Approach |
|  |  |  | procedure | UMLS:ICD10PCS:0TY10Z0 | Transplantation of Left Kidney, Allogeneic, Open Approach |
|  |  |  | procedure | UMLS:ICD10PCS:0TY10Z1 | Transplantation of Left Kidney, Syngeneic, Open Approach |
|  |  |  | procedure | UMLS:ICD10PCS:0TY10Z2 | Transplantation of Left Kidney, Zooplastic, Open Approach |
|  |  |  | procedure | UMLS:ICD10PCS:0TY00Z1 | Transplantation of Right Kidney, Syngeneic, Open Approach |
|  |  |  | procedure | UMLS:ICD10PCS:0TY00Z2 | Transplantation of Right Kidney, Zooplastic, Open Approach |
|  |  |  | procedure | UMLS:ICD10PCS:0TY0 | Urinary System / Transplantation / Kidney, Right |
|  |  |  | procedure | UMLS:ICD10PCS:0TY1 | Urinary System / Transplantation / Kidney, Left |
|  | date constraint | | The terms in this group occurred between Jan 1, 2015 and Jan 1, 2022 | | |
| Group 2 | | | | | |
|  | **Group 2A Kidney transplant** | | | | |
|  | must have | any of | diagnosis | UMLS:ICD10CM:Z94.0 | Kidney transplant status |
|  |  |  | procedure | UMLS:ICD10PCS:0TY00Z0 | Transplantation of Right Kidney, Allogeneic, Open Approach |
|  |  |  | procedure | UMLS:ICD10PCS:0TY10Z0 | Transplantation of Left Kidney, Allogeneic, Open Approach |
|  |  |  | procedure | UMLS:ICD10PCS:0TY10Z1 | Transplantation of Left Kidney, Syngeneic, Open Approach |
|  |  |  | procedure | UMLS:ICD10PCS:0TY10Z2 | Transplantation of Left Kidney, Zooplastic, Open Approach |
|  |  |  | procedure | UMLS:ICD10PCS:0TY00Z1 | Transplantation of Right Kidney, Syngeneic, Open Approach |
|  |  |  | procedure | UMLS:ICD10PCS:0TY00Z2 | Transplantation of Right Kidney, Zooplastic, Open Approach |
|  |  |  | procedure | UMLS:ICD10PCS:0TY0 | Urinary System / Transplantation / Kidney, Right |
|  |  |  | procedure | UMLS:ICD10PCS:0TY1 | Urinary System / Transplantation / Kidney, Left |
|  | date constraint | | The terms in this group occurred at any time | | |
|  | event relationship | | Any instance of noDialysis, Death, SGLT2i occurred within 1 day and 999 days after the first instance of Kidney transplant | | |
|  | **Group 2B noDialysis, Death, SGLT2i** | | | | |
|  | cannot have |  | procedure | UMLS:CPT:1012740 | Dialysis Services and Procedures |
|  |  | or | procedure | UMLS:CPT:90945 | Dialysis procedure other than hemodialysis (eg, peritoneal dialysis, hemofiltration, or other continuous renal replacement therapies), with single evaluation by a physician or other qualified health care professional |
|  |  | or | procedure | UMLS:CPT:90947 | Dialysis procedure other than hemodialysis (eg, peritoneal dialysis, hemofiltration, or other continuous renal replacement therapies) requiring repeated evaluations by a physician or other qualified health care professional, with or without substantial revision of dialysis prescription |
|  |  | or | procedure | UMLS:CPT:1012752 | Hemodialysis Procedures |
|  |  | or | procedure | UMLS:SNOMED:302497006 | Hemodialysis |
|  |  | or | procedure | UMLS:CPT:90937 | Hemodialysis procedure requiring repeated evaluation(s) with or without substantial revision of dialysis prescription |
|  |  | or | procedure | UMLS:CPT:90935 | Hemodialysis procedure with single evaluation by a physician or other qualified health care professional |
|  |  | or | demographics | Deceased | Deceased |
|  |  | or | diagnosis | UMLS:ICD10CM:R99 | Ill-defined and unknown cause of mortality |
|  |  | or | medication | NLM:ATC:A10BK | Sodium-glucose co-transporter 2 (SGLT2) inhibitors |
| Group 3 | | | | | |
|  | **Group 3A Kidney transplant** | | | | |
|  | must have | any of | diagnosis | UMLS:ICD10CM:Z94.0 | Kidney transplant status |
|  |  |  | procedure | UMLS:ICD10PCS:0TY00Z0 | Transplantation of Right Kidney, Allogeneic, Open Approach |
|  |  |  | procedure | UMLS:ICD10PCS:0TY10Z0 | Transplantation of Left Kidney, Allogeneic, Open Approach |
|  |  |  | procedure | UMLS:ICD10PCS:0TY10Z1 | Transplantation of Left Kidney, Syngeneic, Open Approach |
|  |  |  | procedure | UMLS:ICD10PCS:0TY10Z2 | Transplantation of Left Kidney, Zooplastic, Open Approach |
|  |  |  | procedure | UMLS:ICD10PCS:0TY00Z1 | Transplantation of Right Kidney, Syngeneic, Open Approach |
|  |  |  | procedure | UMLS:ICD10PCS:0TY00Z2 | Transplantation of Right Kidney, Zooplastic, Open Approach |
|  |  |  | procedure | UMLS:ICD10PCS:0TY0 | Urinary System / Transplantation / Kidney, Right |
|  |  |  | procedure | UMLS:ICD10PCS:0TY1 | Urinary System / Transplantation / Kidney, Left |
|  | date constraint | | The terms in this group occurred at any time | | |
|  | event relationship | | Any instance of SGLT2i occurred at least 1000 days after the first instance of Kidney transplant | | |
|  | **Group 3B SGLT2i** | | | | |
|  | must have |  | medication | NLM:ATC:A10BK | Sodium-glucose co-transporter 2 (SGLT2) inhibitors |
| Group 4 | | | | | |
|  | **Group 4A Kidney transplant** | | | | |
|  | must have | any of | diagnosis | UMLS:ICD10CM:Z94.0 | Kidney transplant status |
|  |  |  | procedure | UMLS:ICD10PCS:0TY00Z0 | Transplantation of Right Kidney, Allogeneic, Open Approach |
|  |  |  | procedure | UMLS:ICD10PCS:0TY10Z0 | Transplantation of Left Kidney, Allogeneic, Open Approach |
|  |  |  | procedure | UMLS:ICD10PCS:0TY10Z1 | Transplantation of Left Kidney, Syngeneic, Open Approach |
|  |  |  | procedure | UMLS:ICD10PCS:0TY10Z2 | Transplantation of Left Kidney, Zooplastic, Open Approach |
|  |  |  | procedure | UMLS:ICD10PCS:0TY00Z1 | Transplantation of Right Kidney, Syngeneic, Open Approach |
|  |  |  | procedure | UMLS:ICD10PCS:0TY00Z2 | Transplantation of Right Kidney, Zooplastic, Open Approach |
|  |  |  | procedure | UMLS:ICD10PCS:0TY0 | Urinary System / Transplantation / Kidney, Right |
|  |  |  | procedure | UMLS:ICD10PCS:0TY1 | Urinary System / Transplantation / Kidney, Left |
|  | date constraint | | The terms in this group occurred at any time | | |
|  | event relationship | | Any instance of noSGLT2i occurred at least 1 day before the first instance of Kidney transplant | | |
|  | **Group 4B noSGLT2i** | | | | |
|  | cannot have |  | medication | NLM:ATC:A10BK | Sodium-glucose co-transporter 2 (SGLT2) inhibitors |
| Group 5 | | | | | |
|  | **Group 5A Kidney transplant** | | | | |
|  | must have | any of | diagnosis | UMLS:ICD10CM:Z94.0 | Kidney transplant status |
|  |  |  | procedure | UMLS:ICD10PCS:0TY00Z0 | Transplantation of Right Kidney, Allogeneic, Open Approach |
|  |  |  | procedure | UMLS:ICD10PCS:0TY10Z0 | Transplantation of Left Kidney, Allogeneic, Open Approach |
|  |  |  | procedure | UMLS:ICD10PCS:0TY10Z1 | Transplantation of Left Kidney, Syngeneic, Open Approach |
|  |  |  | procedure | UMLS:ICD10PCS:0TY10Z2 | Transplantation of Left Kidney, Zooplastic, Open Approach |
|  |  |  | procedure | UMLS:ICD10PCS:0TY00Z1 | Transplantation of Right Kidney, Syngeneic, Open Approach |
|  |  |  | procedure | UMLS:ICD10PCS:0TY00Z2 | Transplantation of Right Kidney, Zooplastic, Open Approach |
|  |  |  | procedure | UMLS:ICD10PCS:0TY0 | Urinary System / Transplantation / Kidney, Right |
|  |  |  | procedure | UMLS:ICD10PCS:0TY1 | Urinary System / Transplantation / Kidney, Left |
|  | date constraint | | The terms in this group occurred at any time | | |
|  | event relationship | | Any instance of Visit occurred at least 1181 days after the first instance of Kidney transplant | | |
|  | **Group 5B Visit** | | | | |
|  | must have |  | visit | TNX:Visit | Visit |
| Group 6 | | | | | |
|  | **Group 6A Kidney transplant** | | | | |
|  | must have | any of | diagnosis | UMLS:ICD10CM:Z94.0 | Kidney transplant status |
|  |  |  | procedure | UMLS:ICD10PCS:0TY00Z0 | Transplantation of Right Kidney, Allogeneic, Open Approach |
|  |  |  | procedure | UMLS:ICD10PCS:0TY10Z0 | Transplantation of Left Kidney, Allogeneic, Open Approach |
|  |  |  | procedure | UMLS:ICD10PCS:0TY10Z1 | Transplantation of Left Kidney, Syngeneic, Open Approach |
|  |  |  | procedure | UMLS:ICD10PCS:0TY10Z2 | Transplantation of Left Kidney, Zooplastic, Open Approach |
|  |  |  | procedure | UMLS:ICD10PCS:0TY00Z1 | Transplantation of Right Kidney, Syngeneic, Open Approach |
|  |  |  | procedure | UMLS:ICD10PCS:0TY00Z2 | Transplantation of Right Kidney, Zooplastic, Open Approach |
|  |  |  | procedure | UMLS:ICD10PCS:0TY0 | Urinary System / Transplantation / Kidney, Right |
|  |  |  | procedure | UMLS:ICD10PCS:0TY1 | Urinary System / Transplantation / Kidney, Left |
|  | date constraint | | The terms in this group occurred at any time | | |
|  | event relationship | | Any instance of 15-29 eGFR occurred within 1 day and 999 days after the first instance of Kidney transplant | | |
|  | **Group 6B 15-29 eGFR** | | | | |
|  | must have |  | laboratory | TNX:8001 | Glomerular filtration rate/1.73 sq M.predicted [Volume Rate/Area] in Serum, Plasma or Blood by Creatinine-based formula (MDRD) (between 15.00 and 30.00 mL/min/{1.73_m2}) |

##### **Table S52. Code definitions and selection criteria for SGLT2i non-users – 15-29 eGFR.**

| Ungrouped terms | | | | | |
| --- | --- | --- | --- | --- | --- |
|  | must have |  | demographics | Age | Age (at least 18 years (most recent occurrence)) |
|  | cannot have |  | diagnosis | UMLS:ICD10CM:E08-E13 | Diabetes mellitus |
| Group 1 | | | | | |
|  | **Kidney transplant** | | | | |
|  | must have | any of | diagnosis | UMLS:ICD10CM:Z94.0 | Kidney transplant status |
|  |  |  | procedure | UMLS:ICD10PCS:0TY00Z0 | Transplantation of Right Kidney, Allogeneic, Open Approach |
|  |  |  | procedure | UMLS:ICD10PCS:0TY10Z0 | Transplantation of Left Kidney, Allogeneic, Open Approach |
|  |  |  | procedure | UMLS:ICD10PCS:0TY10Z1 | Transplantation of Left Kidney, Syngeneic, Open Approach |
|  |  |  | procedure | UMLS:ICD10PCS:0TY10Z2 | Transplantation of Left Kidney, Zooplastic, Open Approach |
|  |  |  | procedure | UMLS:ICD10PCS:0TY00Z1 | Transplantation of Right Kidney, Syngeneic, Open Approach |
|  |  |  | procedure | UMLS:ICD10PCS:0TY00Z2 | Transplantation of Right Kidney, Zooplastic, Open Approach |
|  |  |  | procedure | UMLS:ICD10PCS:0TY0 | Urinary System / Transplantation / Kidney, Right |
|  |  |  | procedure | UMLS:ICD10PCS:0TY1 | Urinary System / Transplantation / Kidney, Left |
|  | date constraint | | The terms in this group occurred between Jan 1, 2015 and Jan 1, 2022 | | |
| Group 2 | | | | | |
|  | **Group 2A Kidney transplant** | | | | |
|  | must have | any of | diagnosis | UMLS:ICD10CM:Z94.0 | Kidney transplant status |
|  |  |  | procedure | UMLS:ICD10PCS:0TY00Z0 | Transplantation of Right Kidney, Allogeneic, Open Approach |
|  |  |  | procedure | UMLS:ICD10PCS:0TY10Z0 | Transplantation of Left Kidney, Allogeneic, Open Approach |
|  |  |  | procedure | UMLS:ICD10PCS:0TY10Z1 | Transplantation of Left Kidney, Syngeneic, Open Approach |
|  |  |  | procedure | UMLS:ICD10PCS:0TY10Z2 | Transplantation of Left Kidney, Zooplastic, Open Approach |
|  |  |  | procedure | UMLS:ICD10PCS:0TY00Z1 | Transplantation of Right Kidney, Syngeneic, Open Approach |
|  |  |  | procedure | UMLS:ICD10PCS:0TY00Z2 | Transplantation of Right Kidney, Zooplastic, Open Approach |
|  |  |  | procedure | UMLS:ICD10PCS:0TY0 | Urinary System / Transplantation / Kidney, Right |
|  |  |  | procedure | UMLS:ICD10PCS:0TY1 | Urinary System / Transplantation / Kidney, Left |
|  | date constraint | | The terms in this group occurred at any time | | |
|  | event relationship | | Any instance of noDialysis, Death, SGLT2i occurred within 1 day and 999 days after the first instance of Kidney transplant | | |
|  | **Group 2B noDialysis, Death, SGLT2i** | | | | |
|  | cannot have |  | procedure | UMLS:CPT:1012740 | Dialysis Services and Procedures |
|  |  | or | procedure | UMLS:CPT:90945 | Dialysis procedure other than hemodialysis (eg, peritoneal dialysis, hemofiltration, or other continuous renal replacement therapies), with single evaluation by a physician or other qualified health care professional |
|  |  | or | procedure | UMLS:CPT:90947 | Dialysis procedure other than hemodialysis (eg, peritoneal dialysis, hemofiltration, or other continuous renal replacement therapies) requiring repeated evaluations by a physician or other qualified health care professional, with or without substantial revision of dialysis prescription |
|  |  | or | procedure | UMLS:CPT:1012752 | Hemodialysis Procedures |
|  |  | or | procedure | UMLS:SNOMED:302497006 | Hemodialysis |
|  |  | or | procedure | UMLS:CPT:90937 | Hemodialysis procedure requiring repeated evaluation(s) with or without substantial revision of dialysis prescription |
|  |  | or | procedure | UMLS:CPT:90935 | Hemodialysis procedure with single evaluation by a physician or other qualified health care professional |
|  |  | or | demographics | Deceased | Deceased |
|  |  | or | diagnosis | UMLS:ICD10CM:R99 | Ill-defined and unknown cause of mortality |
|  |  | or | medication | NLM:ATC:A10BK | Sodium-glucose co-transporter 2 (SGLT2) inhibitors |
| Group 3 | | | | | |
|  | **Group 3A Kidney transplant** | | | | |
|  | must have | any of | diagnosis | UMLS:ICD10CM:Z94.0 | Kidney transplant status |
|  |  |  | procedure | UMLS:ICD10PCS:0TY00Z0 | Transplantation of Right Kidney, Allogeneic, Open Approach |
|  |  |  | procedure | UMLS:ICD10PCS:0TY10Z0 | Transplantation of Left Kidney, Allogeneic, Open Approach |
|  |  |  | procedure | UMLS:ICD10PCS:0TY10Z1 | Transplantation of Left Kidney, Syngeneic, Open Approach |
|  |  |  | procedure | UMLS:ICD10PCS:0TY10Z2 | Transplantation of Left Kidney, Zooplastic, Open Approach |
|  |  |  | procedure | UMLS:ICD10PCS:0TY00Z1 | Transplantation of Right Kidney, Syngeneic, Open Approach |
|  |  |  | procedure | UMLS:ICD10PCS:0TY00Z2 | Transplantation of Right Kidney, Zooplastic, Open Approach |
|  |  |  | procedure | UMLS:ICD10PCS:0TY0 | Urinary System / Transplantation / Kidney, Right |
|  |  |  | procedure | UMLS:ICD10PCS:0TY1 | Urinary System / Transplantation / Kidney, Left |
|  | date constraint | | The terms in this group occurred at any time | | |
|  | event relationship | | Any instance of noSGLT2i occurred within 1000 days and 3006 days after the first instance of Kidney transplant | | |
|  | **Group 3B noSGLT2i** | | | | |
|  | cannot have |  | medication | NLM:ATC:A10BK | Sodium-glucose co-transporter 2 (SGLT2) inhibitors |
| Group 4 | | | | | |
|  | **Group 4A Kidney transplant** | | | | |
|  | must have | any of | diagnosis | UMLS:ICD10CM:Z94.0 | Kidney transplant status |
|  |  |  | procedure | UMLS:ICD10PCS:0TY00Z0 | Transplantation of Right Kidney, Allogeneic, Open Approach |
|  |  |  | procedure | UMLS:ICD10PCS:0TY10Z0 | Transplantation of Left Kidney, Allogeneic, Open Approach |
|  |  |  | procedure | UMLS:ICD10PCS:0TY10Z1 | Transplantation of Left Kidney, Syngeneic, Open Approach |
|  |  |  | procedure | UMLS:ICD10PCS:0TY10Z2 | Transplantation of Left Kidney, Zooplastic, Open Approach |
|  |  |  | procedure | UMLS:ICD10PCS:0TY00Z1 | Transplantation of Right Kidney, Syngeneic, Open Approach |
|  |  |  | procedure | UMLS:ICD10PCS:0TY00Z2 | Transplantation of Right Kidney, Zooplastic, Open Approach |
|  |  |  | procedure | UMLS:ICD10PCS:0TY0 | Urinary System / Transplantation / Kidney, Right |
|  |  |  | procedure | UMLS:ICD10PCS:0TY1 | Urinary System / Transplantation / Kidney, Left |
|  | date constraint | | The terms in this group occurred at any time | | |
|  | event relationship | | Any instance of noSGLT2i occurred at least 1 day before the first instance of Kidney transplant | | |
|  | **Group 4B noSGLT2i** | | | | |
|  | cannot have |  | medication | NLM:ATC:A10BK | Sodium-glucose co-transporter 2 (SGLT2) inhibitors |
| Group 5 | | | | | |
|  | **Group 5A Kidney transplant** | | | | |
|  | must have | any of | diagnosis | UMLS:ICD10CM:Z94.0 | Kidney transplant status |
|  |  |  | procedure | UMLS:ICD10PCS:0TY00Z0 | Transplantation of Right Kidney, Allogeneic, Open Approach |
|  |  |  | procedure | UMLS:ICD10PCS:0TY10Z0 | Transplantation of Left Kidney, Allogeneic, Open Approach |
|  |  |  | procedure | UMLS:ICD10PCS:0TY10Z1 | Transplantation of Left Kidney, Syngeneic, Open Approach |
|  |  |  | procedure | UMLS:ICD10PCS:0TY10Z2 | Transplantation of Left Kidney, Zooplastic, Open Approach |
|  |  |  | procedure | UMLS:ICD10PCS:0TY00Z1 | Transplantation of Right Kidney, Syngeneic, Open Approach |
|  |  |  | procedure | UMLS:ICD10PCS:0TY00Z2 | Transplantation of Right Kidney, Zooplastic, Open Approach |
|  |  |  | procedure | UMLS:ICD10PCS:0TY0 | Urinary System / Transplantation / Kidney, Right |
|  |  |  | procedure | UMLS:ICD10PCS:0TY1 | Urinary System / Transplantation / Kidney, Left |
|  | date constraint | | The terms in this group occurred at any time | | |
|  | event relationship | | Any instance of Visit occurred at least 1181 days after the first instance of Kidney transplant | | |
|  | **Group 5B Visit** | | | | |
|  | must have |  | visit | TNX:Visit | Visit |
| Group 6 | | | | | |
|  | **Group 6A Kidney transplant** | | | | |
|  | must have | any of | diagnosis | UMLS:ICD10CM:Z94.0 | Kidney transplant status |
|  |  |  | procedure | UMLS:ICD10PCS:0TY00Z0 | Transplantation of Right Kidney, Allogeneic, Open Approach |
|  |  |  | procedure | UMLS:ICD10PCS:0TY10Z0 | Transplantation of Left Kidney, Allogeneic, Open Approach |
|  |  |  | procedure | UMLS:ICD10PCS:0TY10Z1 | Transplantation of Left Kidney, Syngeneic, Open Approach |
|  |  |  | procedure | UMLS:ICD10PCS:0TY10Z2 | Transplantation of Left Kidney, Zooplastic, Open Approach |
|  |  |  | procedure | UMLS:ICD10PCS:0TY00Z1 | Transplantation of Right Kidney, Syngeneic, Open Approach |
|  |  |  | procedure | UMLS:ICD10PCS:0TY00Z2 | Transplantation of Right Kidney, Zooplastic, Open Approach |
|  |  |  | procedure | UMLS:ICD10PCS:0TY0 | Urinary System / Transplantation / Kidney, Right |
|  |  |  | procedure | UMLS:ICD10PCS:0TY1 | Urinary System / Transplantation / Kidney, Left |
|  | date constraint | | The terms in this group occurred at any time | | |
|  | event relationship | | Any instance of 15-29 eGFR occurred within 1 day and 999 days after the first instance of Kidney transplant | | |
|  | **Group 6B 15-29 eGFR** | | | | |
|  | must have |  | laboratory | TNX:8001 | Glomerular filtration rate/1.73 sq M.predicted [Volume Rate/Area] in Serum, Plasma or Blood by Creatinine-based formula (MDRD) (between 15.00 and 30.00 mL/min/{1.73_m2}) |

##### **Table S53. Code definitions and selection criteria for SGLT2i users – 30-59 eGFR.**

| Ungrouped terms | | | | | |
| --- | --- | --- | --- | --- | --- |
|  | must have |  | demographics | Age | Age (at least 18 years (most recent occurrence)) |
|  | cannot have |  | diagnosis | UMLS:ICD10CM:E08-E13 | Diabetes mellitus |
| Group 1 | | | | | |
|  | **Kidney transplant** | | | | |
|  | must have | any of | diagnosis | UMLS:ICD10CM:Z94.0 | Kidney transplant status |
|  |  |  | procedure | UMLS:ICD10PCS:0TY00Z0 | Transplantation of Right Kidney, Allogeneic, Open Approach |
|  |  |  | procedure | UMLS:ICD10PCS:0TY10Z0 | Transplantation of Left Kidney, Allogeneic, Open Approach |
|  |  |  | procedure | UMLS:ICD10PCS:0TY10Z1 | Transplantation of Left Kidney, Syngeneic, Open Approach |
|  |  |  | procedure | UMLS:ICD10PCS:0TY10Z2 | Transplantation of Left Kidney, Zooplastic, Open Approach |
|  |  |  | procedure | UMLS:ICD10PCS:0TY00Z1 | Transplantation of Right Kidney, Syngeneic, Open Approach |
|  |  |  | procedure | UMLS:ICD10PCS:0TY00Z2 | Transplantation of Right Kidney, Zooplastic, Open Approach |
|  |  |  | procedure | UMLS:ICD10PCS:0TY0 | Urinary System / Transplantation / Kidney, Right |
|  |  |  | procedure | UMLS:ICD10PCS:0TY1 | Urinary System / Transplantation / Kidney, Left |
|  | date constraint | | The terms in this group occurred between Jan 1, 2015 and Jan 1, 2022 | | |
| Group 2 | | | | | |
|  | **Group 2A Kidney transplant** | | | | |
|  | must have | any of | diagnosis | UMLS:ICD10CM:Z94.0 | Kidney transplant status |
|  |  |  | procedure | UMLS:ICD10PCS:0TY00Z0 | Transplantation of Right Kidney, Allogeneic, Open Approach |
|  |  |  | procedure | UMLS:ICD10PCS:0TY10Z0 | Transplantation of Left Kidney, Allogeneic, Open Approach |
|  |  |  | procedure | UMLS:ICD10PCS:0TY10Z1 | Transplantation of Left Kidney, Syngeneic, Open Approach |
|  |  |  | procedure | UMLS:ICD10PCS:0TY10Z2 | Transplantation of Left Kidney, Zooplastic, Open Approach |
|  |  |  | procedure | UMLS:ICD10PCS:0TY00Z1 | Transplantation of Right Kidney, Syngeneic, Open Approach |
|  |  |  | procedure | UMLS:ICD10PCS:0TY00Z2 | Transplantation of Right Kidney, Zooplastic, Open Approach |
|  |  |  | procedure | UMLS:ICD10PCS:0TY0 | Urinary System / Transplantation / Kidney, Right |
|  |  |  | procedure | UMLS:ICD10PCS:0TY1 | Urinary System / Transplantation / Kidney, Left |
|  | date constraint | | The terms in this group occurred at any time | | |
|  | event relationship | | Any instance of noDialysis, Death, SGLT2i occurred within 1 day and 999 days after the first instance of Kidney transplant | | |
|  | **Group 2B noDialysis, Death, SGLT2i** | | | | |
|  | cannot have |  | procedure | UMLS:CPT:1012740 | Dialysis Services and Procedures |
|  |  | or | procedure | UMLS:CPT:90945 | Dialysis procedure other than hemodialysis (eg, peritoneal dialysis, hemofiltration, or other continuous renal replacement therapies), with single evaluation by a physician or other qualified health care professional |
|  |  | or | procedure | UMLS:CPT:90947 | Dialysis procedure other than hemodialysis (eg, peritoneal dialysis, hemofiltration, or other continuous renal replacement therapies) requiring repeated evaluations by a physician or other qualified health care professional, with or without substantial revision of dialysis prescription |
|  |  | or | procedure | UMLS:CPT:1012752 | Hemodialysis Procedures |
|  |  | or | procedure | UMLS:SNOMED:302497006 | Hemodialysis |
|  |  | or | procedure | UMLS:CPT:90937 | Hemodialysis procedure requiring repeated evaluation(s) with or without substantial revision of dialysis prescription |
|  |  | or | procedure | UMLS:CPT:90935 | Hemodialysis procedure with single evaluation by a physician or other qualified health care professional |
|  |  | or | demographics | Deceased | Deceased |
|  |  | or | diagnosis | UMLS:ICD10CM:R99 | Ill-defined and unknown cause of mortality |
|  |  | or | medication | NLM:ATC:A10BK | Sodium-glucose co-transporter 2 (SGLT2) inhibitors |
| Group 3 | | | | | |
|  | **Group 3A Kidney transplant** | | | | |
|  | must have | any of | diagnosis | UMLS:ICD10CM:Z94.0 | Kidney transplant status |
|  |  |  | procedure | UMLS:ICD10PCS:0TY00Z0 | Transplantation of Right Kidney, Allogeneic, Open Approach |
|  |  |  | procedure | UMLS:ICD10PCS:0TY10Z0 | Transplantation of Left Kidney, Allogeneic, Open Approach |
|  |  |  | procedure | UMLS:ICD10PCS:0TY10Z1 | Transplantation of Left Kidney, Syngeneic, Open Approach |
|  |  |  | procedure | UMLS:ICD10PCS:0TY10Z2 | Transplantation of Left Kidney, Zooplastic, Open Approach |
|  |  |  | procedure | UMLS:ICD10PCS:0TY00Z1 | Transplantation of Right Kidney, Syngeneic, Open Approach |
|  |  |  | procedure | UMLS:ICD10PCS:0TY00Z2 | Transplantation of Right Kidney, Zooplastic, Open Approach |
|  |  |  | procedure | UMLS:ICD10PCS:0TY0 | Urinary System / Transplantation / Kidney, Right |
|  |  |  | procedure | UMLS:ICD10PCS:0TY1 | Urinary System / Transplantation / Kidney, Left |
|  | date constraint | | The terms in this group occurred at any time | | |
|  | event relationship | | Any instance of SGLT2i occurred at least 1000 days after the first instance of Kidney transplant | | |
|  | **Group 3B SGLT2i** | | | | |
|  | must have |  | medication | NLM:ATC:A10BK | Sodium-glucose co-transporter 2 (SGLT2) inhibitors |
| Group 4 | | | | | |
|  | **Group 4A Kidney transplant** | | | | |
|  | must have | any of | diagnosis | UMLS:ICD10CM:Z94.0 | Kidney transplant status |
|  |  |  | procedure | UMLS:ICD10PCS:0TY00Z0 | Transplantation of Right Kidney, Allogeneic, Open Approach |
|  |  |  | procedure | UMLS:ICD10PCS:0TY10Z0 | Transplantation of Left Kidney, Allogeneic, Open Approach |
|  |  |  | procedure | UMLS:ICD10PCS:0TY10Z1 | Transplantation of Left Kidney, Syngeneic, Open Approach |
|  |  |  | procedure | UMLS:ICD10PCS:0TY10Z2 | Transplantation of Left Kidney, Zooplastic, Open Approach |
|  |  |  | procedure | UMLS:ICD10PCS:0TY00Z1 | Transplantation of Right Kidney, Syngeneic, Open Approach |
|  |  |  | procedure | UMLS:ICD10PCS:0TY00Z2 | Transplantation of Right Kidney, Zooplastic, Open Approach |
|  |  |  | procedure | UMLS:ICD10PCS:0TY0 | Urinary System / Transplantation / Kidney, Right |
|  |  |  | procedure | UMLS:ICD10PCS:0TY1 | Urinary System / Transplantation / Kidney, Left |
|  | date constraint | | The terms in this group occurred at any time | | |
|  | event relationship | | Any instance of noSGLT2i occurred at least 1 day before the first instance of Kidney transplant | | |
|  | **Group 4B noSGLT2i** | | | | |
|  | cannot have |  | medication | NLM:ATC:A10BK | Sodium-glucose co-transporter 2 (SGLT2) inhibitors |
| Group 5 | | | | | |
|  | **Group 5A Kidney transplant** | | | | |
|  | must have | any of | diagnosis | UMLS:ICD10CM:Z94.0 | Kidney transplant status |
|  |  |  | procedure | UMLS:ICD10PCS:0TY00Z0 | Transplantation of Right Kidney, Allogeneic, Open Approach |
|  |  |  | procedure | UMLS:ICD10PCS:0TY10Z0 | Transplantation of Left Kidney, Allogeneic, Open Approach |
|  |  |  | procedure | UMLS:ICD10PCS:0TY10Z1 | Transplantation of Left Kidney, Syngeneic, Open Approach |
|  |  |  | procedure | UMLS:ICD10PCS:0TY10Z2 | Transplantation of Left Kidney, Zooplastic, Open Approach |
|  |  |  | procedure | UMLS:ICD10PCS:0TY00Z1 | Transplantation of Right Kidney, Syngeneic, Open Approach |
|  |  |  | procedure | UMLS:ICD10PCS:0TY00Z2 | Transplantation of Right Kidney, Zooplastic, Open Approach |
|  |  |  | procedure | UMLS:ICD10PCS:0TY0 | Urinary System / Transplantation / Kidney, Right |
|  |  |  | procedure | UMLS:ICD10PCS:0TY1 | Urinary System / Transplantation / Kidney, Left |
|  | date constraint | | The terms in this group occurred at any time | | |
|  | event relationship | | Any instance of Visit occurred at least 1181 days after the first instance of Kidney transplant | | |
|  | **Group 5B Visit** | | | | |
|  | must have |  | visit | TNX:Visit | Visit |
| Group 6 | | | | | |
|  | **Group 6A Kidney transplant** | | | | |
|  | must have | any of | diagnosis | UMLS:ICD10CM:Z94.0 | Kidney transplant status |
|  |  |  | procedure | UMLS:ICD10PCS:0TY00Z0 | Transplantation of Right Kidney, Allogeneic, Open Approach |
|  |  |  | procedure | UMLS:ICD10PCS:0TY10Z0 | Transplantation of Left Kidney, Allogeneic, Open Approach |
|  |  |  | procedure | UMLS:ICD10PCS:0TY10Z1 | Transplantation of Left Kidney, Syngeneic, Open Approach |
|  |  |  | procedure | UMLS:ICD10PCS:0TY10Z2 | Transplantation of Left Kidney, Zooplastic, Open Approach |
|  |  |  | procedure | UMLS:ICD10PCS:0TY00Z1 | Transplantation of Right Kidney, Syngeneic, Open Approach |
|  |  |  | procedure | UMLS:ICD10PCS:0TY00Z2 | Transplantation of Right Kidney, Zooplastic, Open Approach |
|  |  |  | procedure | UMLS:ICD10PCS:0TY0 | Urinary System / Transplantation / Kidney, Right |
|  |  |  | procedure | UMLS:ICD10PCS:0TY1 | Urinary System / Transplantation / Kidney, Left |
|  | date constraint | | The terms in this group occurred at any time | | |
|  | event relationship | | Any instance of 30-59 eGFR occurred within 1 day and 999 days after the first instance of Kidney transplant | | |
|  | **Group 6B 30-59 eGFR** | | | | |
|  | must have |  | laboratory | TNX:8001 | Glomerular filtration rate/1.73 sq M.predicted [Volume Rate/Area] in Serum, Plasma or Blood by Creatinine-based formula (MDRD) (between 30.00 and 60.00 mL/min/{1.73_m2}) |

##### **Table S54. Code definitions and selection criteria for SGLT2i non-users – 30-59 eGFR.**

| Ungrouped terms | | | | | |
| --- | --- | --- | --- | --- | --- |
|  | must have |  | demographics | Age | Age (at least 18 years (most recent occurrence)) |
|  | cannot have |  | diagnosis | UMLS:ICD10CM:E08-E13 | Diabetes mellitus |
| Group 1 | | | | | |
|  | **Kidney transplant** | | | | |
|  | must have | any of | diagnosis | UMLS:ICD10CM:Z94.0 | Kidney transplant status |
|  |  |  | procedure | UMLS:ICD10PCS:0TY00Z0 | Transplantation of Right Kidney, Allogeneic, Open Approach |
|  |  |  | procedure | UMLS:ICD10PCS:0TY10Z0 | Transplantation of Left Kidney, Allogeneic, Open Approach |
|  |  |  | procedure | UMLS:ICD10PCS:0TY10Z1 | Transplantation of Left Kidney, Syngeneic, Open Approach |
|  |  |  | procedure | UMLS:ICD10PCS:0TY10Z2 | Transplantation of Left Kidney, Zooplastic, Open Approach |
|  |  |  | procedure | UMLS:ICD10PCS:0TY00Z1 | Transplantation of Right Kidney, Syngeneic, Open Approach |
|  |  |  | procedure | UMLS:ICD10PCS:0TY00Z2 | Transplantation of Right Kidney, Zooplastic, Open Approach |
|  |  |  | procedure | UMLS:ICD10PCS:0TY0 | Urinary System / Transplantation / Kidney, Right |
|  |  |  | procedure | UMLS:ICD10PCS:0TY1 | Urinary System / Transplantation / Kidney, Left |
|  | date constraint | | The terms in this group occurred between Jan 1, 2015 and Jan 1, 2022 | | |
| Group 2 | | | | | |
|  | **Group 2A Kidney transplant** | | | | |
|  | must have | any of | diagnosis | UMLS:ICD10CM:Z94.0 | Kidney transplant status |
|  |  |  | procedure | UMLS:ICD10PCS:0TY00Z0 | Transplantation of Right Kidney, Allogeneic, Open Approach |
|  |  |  | procedure | UMLS:ICD10PCS:0TY10Z0 | Transplantation of Left Kidney, Allogeneic, Open Approach |
|  |  |  | procedure | UMLS:ICD10PCS:0TY10Z1 | Transplantation of Left Kidney, Syngeneic, Open Approach |
|  |  |  | procedure | UMLS:ICD10PCS:0TY10Z2 | Transplantation of Left Kidney, Zooplastic, Open Approach |
|  |  |  | procedure | UMLS:ICD10PCS:0TY00Z1 | Transplantation of Right Kidney, Syngeneic, Open Approach |
|  |  |  | procedure | UMLS:ICD10PCS:0TY00Z2 | Transplantation of Right Kidney, Zooplastic, Open Approach |
|  |  |  | procedure | UMLS:ICD10PCS:0TY0 | Urinary System / Transplantation / Kidney, Right |
|  |  |  | procedure | UMLS:ICD10PCS:0TY1 | Urinary System / Transplantation / Kidney, Left |
|  | date constraint | | The terms in this group occurred at any time | | |
|  | event relationship | | Any instance of noDialysis, Death, SGLT2i occurred within 1 day and 999 days after the first instance of Kidney transplant | | |
|  | **Group 2B noDialysis, Death, SGLT2i** | | | | |
|  | cannot have |  | procedure | UMLS:CPT:1012740 | Dialysis Services and Procedures |
|  |  | or | procedure | UMLS:CPT:90945 | Dialysis procedure other than hemodialysis (eg, peritoneal dialysis, hemofiltration, or other continuous renal replacement therapies), with single evaluation by a physician or other qualified health care professional |
|  |  | or | procedure | UMLS:CPT:90947 | Dialysis procedure other than hemodialysis (eg, peritoneal dialysis, hemofiltration, or other continuous renal replacement therapies) requiring repeated evaluations by a physician or other qualified health care professional, with or without substantial revision of dialysis prescription |
|  |  | or | procedure | UMLS:CPT:1012752 | Hemodialysis Procedures |
|  |  | or | procedure | UMLS:SNOMED:302497006 | Hemodialysis |
|  |  | or | procedure | UMLS:CPT:90937 | Hemodialysis procedure requiring repeated evaluation(s) with or without substantial revision of dialysis prescription |
|  |  | or | procedure | UMLS:CPT:90935 | Hemodialysis procedure with single evaluation by a physician or other qualified health care professional |
|  |  | or | demographics | Deceased | Deceased |
|  |  | or | diagnosis | UMLS:ICD10CM:R99 | Ill-defined and unknown cause of mortality |
|  |  | or | medication | NLM:ATC:A10BK | Sodium-glucose co-transporter 2 (SGLT2) inhibitors |
| Group 3 | | | | | |
|  | **Group 3A Kidney transplant** | | | | |
|  | must have | any of | diagnosis | UMLS:ICD10CM:Z94.0 | Kidney transplant status |
|  |  |  | procedure | UMLS:ICD10PCS:0TY00Z0 | Transplantation of Right Kidney, Allogeneic, Open Approach |
|  |  |  | procedure | UMLS:ICD10PCS:0TY10Z0 | Transplantation of Left Kidney, Allogeneic, Open Approach |
|  |  |  | procedure | UMLS:ICD10PCS:0TY10Z1 | Transplantation of Left Kidney, Syngeneic, Open Approach |
|  |  |  | procedure | UMLS:ICD10PCS:0TY10Z2 | Transplantation of Left Kidney, Zooplastic, Open Approach |
|  |  |  | procedure | UMLS:ICD10PCS:0TY00Z1 | Transplantation of Right Kidney, Syngeneic, Open Approach |
|  |  |  | procedure | UMLS:ICD10PCS:0TY00Z2 | Transplantation of Right Kidney, Zooplastic, Open Approach |
|  |  |  | procedure | UMLS:ICD10PCS:0TY0 | Urinary System / Transplantation / Kidney, Right |
|  |  |  | procedure | UMLS:ICD10PCS:0TY1 | Urinary System / Transplantation / Kidney, Left |
|  | date constraint | | The terms in this group occurred at any time | | |
|  | event relationship | | Any instance of noSGLT2i occurred within 1000 days and 3006 days after the first instance of Kidney transplant | | |
|  | **Group 3B noSGLT2i** | | | | |
|  | cannot have |  | medication | NLM:ATC:A10BK | Sodium-glucose co-transporter 2 (SGLT2) inhibitors |
| Group 4 | | | | | |
|  | **Group 4A Kidney transplant** | | | | |
|  | must have | any of | diagnosis | UMLS:ICD10CM:Z94.0 | Kidney transplant status |
|  |  |  | procedure | UMLS:ICD10PCS:0TY00Z0 | Transplantation of Right Kidney, Allogeneic, Open Approach |
|  |  |  | procedure | UMLS:ICD10PCS:0TY10Z0 | Transplantation of Left Kidney, Allogeneic, Open Approach |
|  |  |  | procedure | UMLS:ICD10PCS:0TY10Z1 | Transplantation of Left Kidney, Syngeneic, Open Approach |
|  |  |  | procedure | UMLS:ICD10PCS:0TY10Z2 | Transplantation of Left Kidney, Zooplastic, Open Approach |
|  |  |  | procedure | UMLS:ICD10PCS:0TY00Z1 | Transplantation of Right Kidney, Syngeneic, Open Approach |
|  |  |  | procedure | UMLS:ICD10PCS:0TY00Z2 | Transplantation of Right Kidney, Zooplastic, Open Approach |
|  |  |  | procedure | UMLS:ICD10PCS:0TY0 | Urinary System / Transplantation / Kidney, Right |
|  |  |  | procedure | UMLS:ICD10PCS:0TY1 | Urinary System / Transplantation / Kidney, Left |
|  | date constraint | | The terms in this group occurred at any time | | |
|  | event relationship | | Any instance of noSGLT2i occurred at least 1 day before the first instance of Kidney transplant | | |
|  | **Group 4B noSGLT2i** | | | | |
|  | cannot have |  | medication | NLM:ATC:A10BK | Sodium-glucose co-transporter 2 (SGLT2) inhibitors |
| Group 5 | | | | | |
|  | **Group 5A Kidney transplant** | | | | |
|  | must have | any of | diagnosis | UMLS:ICD10CM:Z94.0 | Kidney transplant status |
|  |  |  | procedure | UMLS:ICD10PCS:0TY00Z0 | Transplantation of Right Kidney, Allogeneic, Open Approach |
|  |  |  | procedure | UMLS:ICD10PCS:0TY10Z0 | Transplantation of Left Kidney, Allogeneic, Open Approach |
|  |  |  | procedure | UMLS:ICD10PCS:0TY10Z1 | Transplantation of Left Kidney, Syngeneic, Open Approach |
|  |  |  | procedure | UMLS:ICD10PCS:0TY10Z2 | Transplantation of Left Kidney, Zooplastic, Open Approach |
|  |  |  | procedure | UMLS:ICD10PCS:0TY00Z1 | Transplantation of Right Kidney, Syngeneic, Open Approach |
|  |  |  | procedure | UMLS:ICD10PCS:0TY00Z2 | Transplantation of Right Kidney, Zooplastic, Open Approach |
|  |  |  | procedure | UMLS:ICD10PCS:0TY0 | Urinary System / Transplantation / Kidney, Right |
|  |  |  | procedure | UMLS:ICD10PCS:0TY1 | Urinary System / Transplantation / Kidney, Left |
|  | date constraint | | The terms in this group occurred at any time | | |
|  | event relationship | | Any instance of Visit occurred at least 1181 days after the first instance of Kidney transplant | | |
|  | **Group 5B Visit** | | | | |
|  | must have |  | visit | TNX:Visit | Visit |
| Group 6 | | | | | |
|  | **Group 6A Kidney transplant** | | | | |
|  | must have | any of | diagnosis | UMLS:ICD10CM:Z94.0 | Kidney transplant status |
|  |  |  | procedure | UMLS:ICD10PCS:0TY00Z0 | Transplantation of Right Kidney, Allogeneic, Open Approach |
|  |  |  | procedure | UMLS:ICD10PCS:0TY10Z0 | Transplantation of Left Kidney, Allogeneic, Open Approach |
|  |  |  | procedure | UMLS:ICD10PCS:0TY10Z1 | Transplantation of Left Kidney, Syngeneic, Open Approach |
|  |  |  | procedure | UMLS:ICD10PCS:0TY10Z2 | Transplantation of Left Kidney, Zooplastic, Open Approach |
|  |  |  | procedure | UMLS:ICD10PCS:0TY00Z1 | Transplantation of Right Kidney, Syngeneic, Open Approach |
|  |  |  | procedure | UMLS:ICD10PCS:0TY00Z2 | Transplantation of Right Kidney, Zooplastic, Open Approach |
|  |  |  | procedure | UMLS:ICD10PCS:0TY0 | Urinary System / Transplantation / Kidney, Right |
|  |  |  | procedure | UMLS:ICD10PCS:0TY1 | Urinary System / Transplantation / Kidney, Left |
|  | date constraint | | The terms in this group occurred at any time | | |
|  | event relationship | | Any instance of 30-59 eGFR occurred within 1 day and 999 days after the first instance of Kidney transplant | | |
|  | **Group 6B 30-59 eGFR** | | | | |
|  | must have |  | laboratory | TNX:8001 | Glomerular filtration rate/1.73 sq M.predicted [Volume Rate/Area] in Serum, Plasma or Blood by Creatinine-based formula (MDRD) (between 30.00 and 60.00 mL/min/{1.73_m2}) |

##### **Table S55. Code definitions and selection criteria for SGLT2i users – ≥60 eGFR.**

| Ungrouped terms | | | | | |
| --- | --- | --- | --- | --- | --- |
|  | must have |  | demographics | Age | Age (at least 18 years (most recent occurrence)) |
|  | cannot have |  | diagnosis | UMLS:ICD10CM:E08-E13 | Diabetes mellitus |
| Group 1 | | | | | |
|  | **Kidney transplant** | | | | |
|  | must have | any of | diagnosis | UMLS:ICD10CM:Z94.0 | Kidney transplant status |
|  |  |  | procedure | UMLS:ICD10PCS:0TY00Z0 | Transplantation of Right Kidney, Allogeneic, Open Approach |
|  |  |  | procedure | UMLS:ICD10PCS:0TY10Z0 | Transplantation of Left Kidney, Allogeneic, Open Approach |
|  |  |  | procedure | UMLS:ICD10PCS:0TY10Z1 | Transplantation of Left Kidney, Syngeneic, Open Approach |
|  |  |  | procedure | UMLS:ICD10PCS:0TY10Z2 | Transplantation of Left Kidney, Zooplastic, Open Approach |
|  |  |  | procedure | UMLS:ICD10PCS:0TY00Z1 | Transplantation of Right Kidney, Syngeneic, Open Approach |
|  |  |  | procedure | UMLS:ICD10PCS:0TY00Z2 | Transplantation of Right Kidney, Zooplastic, Open Approach |
|  |  |  | procedure | UMLS:ICD10PCS:0TY0 | Urinary System / Transplantation / Kidney, Right |
|  |  |  | procedure | UMLS:ICD10PCS:0TY1 | Urinary System / Transplantation / Kidney, Left |
|  | date constraint | | The terms in this group occurred between Jan 1, 2015 and Jan 1, 2022 | | |
| Group 2 | | | | | |
|  | **Group 2A Kidney transplant** | | | | |
|  | must have | any of | diagnosis | UMLS:ICD10CM:Z94.0 | Kidney transplant status |
|  |  |  | procedure | UMLS:ICD10PCS:0TY00Z0 | Transplantation of Right Kidney, Allogeneic, Open Approach |
|  |  |  | procedure | UMLS:ICD10PCS:0TY10Z0 | Transplantation of Left Kidney, Allogeneic, Open Approach |
|  |  |  | procedure | UMLS:ICD10PCS:0TY10Z1 | Transplantation of Left Kidney, Syngeneic, Open Approach |
|  |  |  | procedure | UMLS:ICD10PCS:0TY10Z2 | Transplantation of Left Kidney, Zooplastic, Open Approach |
|  |  |  | procedure | UMLS:ICD10PCS:0TY00Z1 | Transplantation of Right Kidney, Syngeneic, Open Approach |
|  |  |  | procedure | UMLS:ICD10PCS:0TY00Z2 | Transplantation of Right Kidney, Zooplastic, Open Approach |
|  |  |  | procedure | UMLS:ICD10PCS:0TY0 | Urinary System / Transplantation / Kidney, Right |
|  |  |  | procedure | UMLS:ICD10PCS:0TY1 | Urinary System / Transplantation / Kidney, Left |
|  | date constraint | | The terms in this group occurred at any time | | |
|  | event relationship | | Any instance of noDialysis, Death, SGLT2i occurred within 1 day and 999 days after the first instance of Kidney transplant | | |
|  | **Group 2B noDialysis, Death, SGLT2i** | | | | |
|  | cannot have |  | procedure | UMLS:CPT:1012740 | Dialysis Services and Procedures |
|  |  | or | procedure | UMLS:CPT:90945 | Dialysis procedure other than hemodialysis (eg, peritoneal dialysis, hemofiltration, or other continuous renal replacement therapies), with single evaluation by a physician or other qualified health care professional |
|  |  | or | procedure | UMLS:CPT:90947 | Dialysis procedure other than hemodialysis (eg, peritoneal dialysis, hemofiltration, or other continuous renal replacement therapies) requiring repeated evaluations by a physician or other qualified health care professional, with or without substantial revision of dialysis prescription |
|  |  | or | procedure | UMLS:CPT:1012752 | Hemodialysis Procedures |
|  |  | or | procedure | UMLS:SNOMED:302497006 | Hemodialysis |
|  |  | or | procedure | UMLS:CPT:90937 | Hemodialysis procedure requiring repeated evaluation(s) with or without substantial revision of dialysis prescription |
|  |  | or | procedure | UMLS:CPT:90935 | Hemodialysis procedure with single evaluation by a physician or other qualified health care professional |
|  |  | or | demographics | Deceased | Deceased |
|  |  | or | diagnosis | UMLS:ICD10CM:R99 | Ill-defined and unknown cause of mortality |
|  |  | or | medication | NLM:ATC:A10BK | Sodium-glucose co-transporter 2 (SGLT2) inhibitors |
| Group 3 | | | | | |
|  | **Group 3A Kidney transplant** | | | | |
|  | must have | any of | diagnosis | UMLS:ICD10CM:Z94.0 | Kidney transplant status |
|  |  |  | procedure | UMLS:ICD10PCS:0TY00Z0 | Transplantation of Right Kidney, Allogeneic, Open Approach |
|  |  |  | procedure | UMLS:ICD10PCS:0TY10Z0 | Transplantation of Left Kidney, Allogeneic, Open Approach |
|  |  |  | procedure | UMLS:ICD10PCS:0TY10Z1 | Transplantation of Left Kidney, Syngeneic, Open Approach |
|  |  |  | procedure | UMLS:ICD10PCS:0TY10Z2 | Transplantation of Left Kidney, Zooplastic, Open Approach |
|  |  |  | procedure | UMLS:ICD10PCS:0TY00Z1 | Transplantation of Right Kidney, Syngeneic, Open Approach |
|  |  |  | procedure | UMLS:ICD10PCS:0TY00Z2 | Transplantation of Right Kidney, Zooplastic, Open Approach |
|  |  |  | procedure | UMLS:ICD10PCS:0TY0 | Urinary System / Transplantation / Kidney, Right |
|  |  |  | procedure | UMLS:ICD10PCS:0TY1 | Urinary System / Transplantation / Kidney, Left |
|  | date constraint | | The terms in this group occurred at any time | | |
|  | event relationship | | Any instance of SGLT2i occurred at least 1000 days after the first instance of Kidney transplant | | |
|  | **Group 3B SGLT2i** | | | | |
|  | must have |  | medication | NLM:ATC:A10BK | Sodium-glucose co-transporter 2 (SGLT2) inhibitors |
| Group 4 | | | | | |
|  | **Group 4A Kidney transplant** | | | | |
|  | must have | any of | diagnosis | UMLS:ICD10CM:Z94.0 | Kidney transplant status |
|  |  |  | procedure | UMLS:ICD10PCS:0TY00Z0 | Transplantation of Right Kidney, Allogeneic, Open Approach |
|  |  |  | procedure | UMLS:ICD10PCS:0TY10Z0 | Transplantation of Left Kidney, Allogeneic, Open Approach |
|  |  |  | procedure | UMLS:ICD10PCS:0TY10Z1 | Transplantation of Left Kidney, Syngeneic, Open Approach |
|  |  |  | procedure | UMLS:ICD10PCS:0TY10Z2 | Transplantation of Left Kidney, Zooplastic, Open Approach |
|  |  |  | procedure | UMLS:ICD10PCS:0TY00Z1 | Transplantation of Right Kidney, Syngeneic, Open Approach |
|  |  |  | procedure | UMLS:ICD10PCS:0TY00Z2 | Transplantation of Right Kidney, Zooplastic, Open Approach |
|  |  |  | procedure | UMLS:ICD10PCS:0TY0 | Urinary System / Transplantation / Kidney, Right |
|  |  |  | procedure | UMLS:ICD10PCS:0TY1 | Urinary System / Transplantation / Kidney, Left |
|  | date constraint | | The terms in this group occurred at any time | | |
|  | event relationship | | Any instance of noSGLT2i occurred at least 1 day before the first instance of Kidney transplant | | |
|  | **Group 4B noSGLT2i** | | | | |
|  | cannot have |  | medication | NLM:ATC:A10BK | Sodium-glucose co-transporter 2 (SGLT2) inhibitors |
| Group 5 | | | | | |
|  | **Group 5A Kidney transplant** | | | | |
|  | must have | any of | diagnosis | UMLS:ICD10CM:Z94.0 | Kidney transplant status |
|  |  |  | procedure | UMLS:ICD10PCS:0TY00Z0 | Transplantation of Right Kidney, Allogeneic, Open Approach |
|  |  |  | procedure | UMLS:ICD10PCS:0TY10Z0 | Transplantation of Left Kidney, Allogeneic, Open Approach |
|  |  |  | procedure | UMLS:ICD10PCS:0TY10Z1 | Transplantation of Left Kidney, Syngeneic, Open Approach |
|  |  |  | procedure | UMLS:ICD10PCS:0TY10Z2 | Transplantation of Left Kidney, Zooplastic, Open Approach |
|  |  |  | procedure | UMLS:ICD10PCS:0TY00Z1 | Transplantation of Right Kidney, Syngeneic, Open Approach |
|  |  |  | procedure | UMLS:ICD10PCS:0TY00Z2 | Transplantation of Right Kidney, Zooplastic, Open Approach |
|  |  |  | procedure | UMLS:ICD10PCS:0TY0 | Urinary System / Transplantation / Kidney, Right |
|  |  |  | procedure | UMLS:ICD10PCS:0TY1 | Urinary System / Transplantation / Kidney, Left |
|  | date constraint | | The terms in this group occurred at any time | | |
|  | event relationship | | Any instance of Visit occurred at least 1181 days after the first instance of Kidney transplant | | |
|  | **Group 5B Visit** | | | | |
|  | must have |  | visit | TNX:Visit | Visit |
| Group 6 | | | | | |
|  | **Group 6A Kidney transplant** | | | | |
|  | must have | any of | diagnosis | UMLS:ICD10CM:Z94.0 | Kidney transplant status |
|  |  |  | procedure | UMLS:ICD10PCS:0TY00Z0 | Transplantation of Right Kidney, Allogeneic, Open Approach |
|  |  |  | procedure | UMLS:ICD10PCS:0TY10Z0 | Transplantation of Left Kidney, Allogeneic, Open Approach |
|  |  |  | procedure | UMLS:ICD10PCS:0TY10Z1 | Transplantation of Left Kidney, Syngeneic, Open Approach |
|  |  |  | procedure | UMLS:ICD10PCS:0TY10Z2 | Transplantation of Left Kidney, Zooplastic, Open Approach |
|  |  |  | procedure | UMLS:ICD10PCS:0TY00Z1 | Transplantation of Right Kidney, Syngeneic, Open Approach |
|  |  |  | procedure | UMLS:ICD10PCS:0TY00Z2 | Transplantation of Right Kidney, Zooplastic, Open Approach |
|  |  |  | procedure | UMLS:ICD10PCS:0TY0 | Urinary System / Transplantation / Kidney, Right |
|  |  |  | procedure | UMLS:ICD10PCS:0TY1 | Urinary System / Transplantation / Kidney, Left |
|  | date constraint | | The terms in this group occurred at any time | | |
|  | event relationship | | Any instance of >59 eGFR occurred within 1 day and 999 days after the first instance of Kidney transplant | | |
|  | **Group 6B >59 eGFR** | | | | |
|  | must have |  | laboratory | TNX:8001 | Glomerular filtration rate/1.73 sq M.predicted [Volume Rate/Area] in Serum, Plasma or Blood by Creatinine-based formula (MDRD) (at least 60.00 mL/min/{1.73_m2}) |

##### **Table S56. Code definitions and selection criteria for SGLT2i non-users – ≥60 eGFR.**

| Ungrouped terms | | | | | |
| --- | --- | --- | --- | --- | --- |
|  | must have |  | demographics | Age | Age (at least 18 years (most recent occurrence)) |
|  | cannot have |  | diagnosis | UMLS:ICD10CM:E08-E13 | Diabetes mellitus |
| Group 1 | | | | | |
|  | **Kidney transplant** | | | | |
|  | must have | any of | diagnosis | UMLS:ICD10CM:Z94.0 | Kidney transplant status |
|  |  |  | procedure | UMLS:ICD10PCS:0TY00Z0 | Transplantation of Right Kidney, Allogeneic, Open Approach |
|  |  |  | procedure | UMLS:ICD10PCS:0TY10Z0 | Transplantation of Left Kidney, Allogeneic, Open Approach |
|  |  |  | procedure | UMLS:ICD10PCS:0TY10Z1 | Transplantation of Left Kidney, Syngeneic, Open Approach |
|  |  |  | procedure | UMLS:ICD10PCS:0TY10Z2 | Transplantation of Left Kidney, Zooplastic, Open Approach |
|  |  |  | procedure | UMLS:ICD10PCS:0TY00Z1 | Transplantation of Right Kidney, Syngeneic, Open Approach |
|  |  |  | procedure | UMLS:ICD10PCS:0TY00Z2 | Transplantation of Right Kidney, Zooplastic, Open Approach |
|  |  |  | procedure | UMLS:ICD10PCS:0TY0 | Urinary System / Transplantation / Kidney, Right |
|  |  |  | procedure | UMLS:ICD10PCS:0TY1 | Urinary System / Transplantation / Kidney, Left |
|  | date constraint | | The terms in this group occurred between Jan 1, 2015 and Jan 1, 2022 | | |
| Group 2 | | | | | |
|  | **Group 2A Kidney transplant** | | | | |
|  | must have | any of | diagnosis | UMLS:ICD10CM:Z94.0 | Kidney transplant status |
|  |  |  | procedure | UMLS:ICD10PCS:0TY00Z0 | Transplantation of Right Kidney, Allogeneic, Open Approach |
|  |  |  | procedure | UMLS:ICD10PCS:0TY10Z0 | Transplantation of Left Kidney, Allogeneic, Open Approach |
|  |  |  | procedure | UMLS:ICD10PCS:0TY10Z1 | Transplantation of Left Kidney, Syngeneic, Open Approach |
|  |  |  | procedure | UMLS:ICD10PCS:0TY10Z2 | Transplantation of Left Kidney, Zooplastic, Open Approach |
|  |  |  | procedure | UMLS:ICD10PCS:0TY00Z1 | Transplantation of Right Kidney, Syngeneic, Open Approach |
|  |  |  | procedure | UMLS:ICD10PCS:0TY00Z2 | Transplantation of Right Kidney, Zooplastic, Open Approach |
|  |  |  | procedure | UMLS:ICD10PCS:0TY0 | Urinary System / Transplantation / Kidney, Right |
|  |  |  | procedure | UMLS:ICD10PCS:0TY1 | Urinary System / Transplantation / Kidney, Left |
|  | date constraint | | The terms in this group occurred at any time | | |
|  | event relationship | | Any instance of noDialysis, Death, SGLT2i occurred within 1 day and 999 days after the first instance of Kidney transplant | | |
|  | **Group 2B noDialysis, Death, SGLT2i** | | | | |
|  | cannot have |  | procedure | UMLS:CPT:1012740 | Dialysis Services and Procedures |
|  |  | or | procedure | UMLS:CPT:90945 | Dialysis procedure other than hemodialysis (eg, peritoneal dialysis, hemofiltration, or other continuous renal replacement therapies), with single evaluation by a physician or other qualified health care professional |
|  |  | or | procedure | UMLS:CPT:90947 | Dialysis procedure other than hemodialysis (eg, peritoneal dialysis, hemofiltration, or other continuous renal replacement therapies) requiring repeated evaluations by a physician or other qualified health care professional, with or without substantial revision of dialysis prescription |
|  |  | or | procedure | UMLS:CPT:1012752 | Hemodialysis Procedures |
|  |  | or | procedure | UMLS:SNOMED:302497006 | Hemodialysis |
|  |  | or | procedure | UMLS:CPT:90937 | Hemodialysis procedure requiring repeated evaluation(s) with or without substantial revision of dialysis prescription |
|  |  | or | procedure | UMLS:CPT:90935 | Hemodialysis procedure with single evaluation by a physician or other qualified health care professional |
|  |  | or | demographics | Deceased | Deceased |
|  |  | or | diagnosis | UMLS:ICD10CM:R99 | Ill-defined and unknown cause of mortality |
|  |  | or | medication | NLM:ATC:A10BK | Sodium-glucose co-transporter 2 (SGLT2) inhibitors |
| Group 3 | | | | | |
|  | **Group 3A Kidney transplant** | | | | |
|  | must have | any of | diagnosis | UMLS:ICD10CM:Z94.0 | Kidney transplant status |
|  |  |  | procedure | UMLS:ICD10PCS:0TY00Z0 | Transplantation of Right Kidney, Allogeneic, Open Approach |
|  |  |  | procedure | UMLS:ICD10PCS:0TY10Z0 | Transplantation of Left Kidney, Allogeneic, Open Approach |
|  |  |  | procedure | UMLS:ICD10PCS:0TY10Z1 | Transplantation of Left Kidney, Syngeneic, Open Approach |
|  |  |  | procedure | UMLS:ICD10PCS:0TY10Z2 | Transplantation of Left Kidney, Zooplastic, Open Approach |
|  |  |  | procedure | UMLS:ICD10PCS:0TY00Z1 | Transplantation of Right Kidney, Syngeneic, Open Approach |
|  |  |  | procedure | UMLS:ICD10PCS:0TY00Z2 | Transplantation of Right Kidney, Zooplastic, Open Approach |
|  |  |  | procedure | UMLS:ICD10PCS:0TY0 | Urinary System / Transplantation / Kidney, Right |
|  |  |  | procedure | UMLS:ICD10PCS:0TY1 | Urinary System / Transplantation / Kidney, Left |
|  | date constraint | | The terms in this group occurred at any time | | |
|  | event relationship | | Any instance of noSGLT2i occurred within 1000 days and 3006 days after the first instance of Kidney transplant | | |
|  | **Group 3B noSGLT2i** | | | | |
|  | cannot have |  | medication | NLM:ATC:A10BK | Sodium-glucose co-transporter 2 (SGLT2) inhibitors |
| Group 4 | | | | | |
|  | **Group 4A Kidney transplant** | | | | |
|  | must have | any of | diagnosis | UMLS:ICD10CM:Z94.0 | Kidney transplant status |
|  |  |  | procedure | UMLS:ICD10PCS:0TY00Z0 | Transplantation of Right Kidney, Allogeneic, Open Approach |
|  |  |  | procedure | UMLS:ICD10PCS:0TY10Z0 | Transplantation of Left Kidney, Allogeneic, Open Approach |
|  |  |  | procedure | UMLS:ICD10PCS:0TY10Z1 | Transplantation of Left Kidney, Syngeneic, Open Approach |
|  |  |  | procedure | UMLS:ICD10PCS:0TY10Z2 | Transplantation of Left Kidney, Zooplastic, Open Approach |
|  |  |  | procedure | UMLS:ICD10PCS:0TY00Z1 | Transplantation of Right Kidney, Syngeneic, Open Approach |
|  |  |  | procedure | UMLS:ICD10PCS:0TY00Z2 | Transplantation of Right Kidney, Zooplastic, Open Approach |
|  |  |  | procedure | UMLS:ICD10PCS:0TY0 | Urinary System / Transplantation / Kidney, Right |
|  |  |  | procedure | UMLS:ICD10PCS:0TY1 | Urinary System / Transplantation / Kidney, Left |
|  | date constraint | | The terms in this group occurred at any time | | |
|  | event relationship | | Any instance of noSGLT2i occurred at least 1 day before the first instance of Kidney transplant | | |
|  | **Group 4B noSGLT2i** | | | | |
|  | cannot have |  | medication | NLM:ATC:A10BK | Sodium-glucose co-transporter 2 (SGLT2) inhibitors |
| Group 5 | | | | | |
|  | **Group 5A Kidney transplant** | | | | |
|  | must have | any of | diagnosis | UMLS:ICD10CM:Z94.0 | Kidney transplant status |
|  |  |  | procedure | UMLS:ICD10PCS:0TY00Z0 | Transplantation of Right Kidney, Allogeneic, Open Approach |
|  |  |  | procedure | UMLS:ICD10PCS:0TY10Z0 | Transplantation of Left Kidney, Allogeneic, Open Approach |
|  |  |  | procedure | UMLS:ICD10PCS:0TY10Z1 | Transplantation of Left Kidney, Syngeneic, Open Approach |
|  |  |  | procedure | UMLS:ICD10PCS:0TY10Z2 | Transplantation of Left Kidney, Zooplastic, Open Approach |
|  |  |  | procedure | UMLS:ICD10PCS:0TY00Z1 | Transplantation of Right Kidney, Syngeneic, Open Approach |
|  |  |  | procedure | UMLS:ICD10PCS:0TY00Z2 | Transplantation of Right Kidney, Zooplastic, Open Approach |
|  |  |  | procedure | UMLS:ICD10PCS:0TY0 | Urinary System / Transplantation / Kidney, Right |
|  |  |  | procedure | UMLS:ICD10PCS:0TY1 | Urinary System / Transplantation / Kidney, Left |
|  | date constraint | | The terms in this group occurred at any time | | |
|  | event relationship | | Any instance of Visit occurred at least 1181 days after the first instance of Kidney transplant | | |
|  | **Group 5B Visit** | | | | |
|  | must have |  | visit | TNX:Visit | Visit |
| Group 6 | | | | | |
|  | **Group 6A Kidney transplant** | | | | |
|  | must have | any of | diagnosis | UMLS:ICD10CM:Z94.0 | Kidney transplant status |
|  |  |  | procedure | UMLS:ICD10PCS:0TY00Z0 | Transplantation of Right Kidney, Allogeneic, Open Approach |
|  |  |  | procedure | UMLS:ICD10PCS:0TY10Z0 | Transplantation of Left Kidney, Allogeneic, Open Approach |
|  |  |  | procedure | UMLS:ICD10PCS:0TY10Z1 | Transplantation of Left Kidney, Syngeneic, Open Approach |
|  |  |  | procedure | UMLS:ICD10PCS:0TY10Z2 | Transplantation of Left Kidney, Zooplastic, Open Approach |
|  |  |  | procedure | UMLS:ICD10PCS:0TY00Z1 | Transplantation of Right Kidney, Syngeneic, Open Approach |
|  |  |  | procedure | UMLS:ICD10PCS:0TY00Z2 | Transplantation of Right Kidney, Zooplastic, Open Approach |
|  |  |  | procedure | UMLS:ICD10PCS:0TY0 | Urinary System / Transplantation / Kidney, Right |
|  |  |  | procedure | UMLS:ICD10PCS:0TY1 | Urinary System / Transplantation / Kidney, Left |
|  | date constraint | | The terms in this group occurred at any time | | |
|  | event relationship | | Any instance of >59 eGFR occurred within 1 day and 999 days after the first instance of Kidney transplant | | |
|  | **Group 6B >59 eGFR** | | | | |
|  | must have |  | laboratory | TNX:8001 | Glomerular filtration rate/1.73 sq M.predicted [Volume Rate/Area] in Serum, Plasma or Blood by Creatinine-based formula (MDRD) (at least 60.00 mL/min/{1.73_m2}) |

##### **Table S57. Covariate definitions used in the Cox regression analysis.**

| **Cox Model Covariates** | | | |
| --- | --- | --- | --- |
|  |  | Covariate | Type |
|  |  | Cohort 1 or Cohort 2 Membership |  |
|  | UMLS:HL7V3.0:Gender:M | Male | Present/Absent |
|  | AgeAtIndex | Age at Index | Continuous |
|  | UMLS:HL7V3.0:Ethnicity:2186-5 | Not Hispanic or Latino | Present/Absent |
|  | UMLS:HL7V3.0:Race:2106-3 | White | Present/Absent |
|  | UMLS:ICD10CM:I10-I1A | Hypertensive diseases | Present/Absent |
|  | UMLS:ICD10CM:I70-I79 | Diseases of arteries, arterioles and capillaries | Present/Absent |
|  | UMLS:ICD10CM:I60-I69 | Cerebrovascular diseases | Present/Absent |
|  | UMLS:ICD10CM:I26-I28 | Pulmonary heart disease and diseases of pulmonary circulation | Present/Absent |
|  | UMLS:ICD10CM:E66 | Overweight and obesity | Present/Absent |
|  | UMLS:ICD10CM:C00-D49 | Neoplasms | Present/Absent |
|  | UMLS:ICD10CM:K70-K77 | Diseases of liver | Present/Absent |
|  | UMLS:ICD10CM:R80 | Proteinuria | Present/Absent |
|  | UMLS:ICD10CM:M30-M36 | Systemic connective tissue disorders | Present/Absent |
|  | UMLS:ICD10CM:I10 | Essential (primary) hypertension | Present/Absent |
|  | UMLS:ICD10CM:I50.2 | Systolic (congestive) heart failure | Present/Absent |
|  | UMLS:ICD10CM:I50.3 | Diastolic (congestive) heart failure | Present/Absent |
|  | UMLS:ICD10CM:N39.0 | Urinary tract infection, site not specified | Present/Absent |
|  | UMLS:ICD10CM:I20-I25 | Ischemic heart diseases | Present/Absent |
|  | UMLS:ICD10CM:J40-J4A | Chronic lower respiratory diseases | Present/Absent |
|  | UMLS:ICD10CM:E78 | Disorders of lipoprotein metabolism and other lipidemias | Present/Absent |
|  | UMLS:ICD10CM:I21 | Acute myocardial infarction | Present/Absent |
|  | UMLS:ICD10CM:I50.9 | Heart failure, unspecified | Present/Absent |
|  | UMLS:ICD10CM:N17 | Acute kidney failure | Present/Absent |
|  | UMLS:ICD10CM:N18 | Chronic kidney disease (CKD) | Present/Absent |
|  | UMLS:ICD10CM:N19 | Unspecified kidney failure | Present/Absent |
|  | UMLS:ICD10CM:Q61 | Cystic kidney disease | Present/Absent |
|  | NLM:VA:CV100 | BETA BLOCKERS/RELATED | Present/Absent |
|  | NLM:VA:CV200 | CALCIUM CHANNEL BLOCKERS | Present/Absent |
|  | NLM:VA:CV300 | ANTIARRHYTHMICS | Present/Absent |
|  | NLM:VA:CV700 | DIURETICS | Present/Absent |
|  | NLM:VA:CV350 | ANTILIPEMIC AGENTS | Present/Absent |
|  | NLM:VA:CV800 | ACE INHIBITORS | Present/Absent |
|  | NLM:VA:CV805 | ANGIOTENSIN II INHIBITOR | Present/Absent |
|  | NLM:VA:CV250 | ANTIANGINALS | Present/Absent |
|  | NLM:ATC:C10AA | HMG CoA reductase inhibitors | Present/Absent |
|  | NLM:VA:HS502 | ORAL HYPOGLYCEMIC AGENTS,ORAL | Present/Absent |
|  | TNX:9002 | Cholesterol in LDL [Mass/volume] in Serum or Plasma | Categorical |
|  |  |  | 0 - 55 mg/dL |
|  |  |  | 55 - 100 mg/dL |
|  |  |  | 100 - 130 mg/dL |
|  |  |  | 130 - 160 mg/dL |
|  |  |  | 160 - 190 mg/dL |
|  |  |  | 190 - 0 mg/dL |
|  | TNX:9001 | Cholesterol in HDL [Mass/volume] in Serum or Plasma | Categorical |
|  |  |  | 0 - 45 mg/dL |
|  |  |  | 45 - 60 mg/dL |
|  |  |  | 60 - 0 mg/dL |
|  | TNX:9083 | BMI | Categorical |
|  |  |  | 0 - 21 kg/m2 |
|  |  |  | 21 - 26 kg/m2 |
|  |  |  | 26 - 31 kg/m2 |
|  |  |  | 31 - 36 kg/m2 |
|  |  |  | 36 - 0 kg/m2 |
|  | TNX:9085 | Blood Pressure, Systolic | Categorical |
|  |  |  | 0 - 110 mm[Hg] |
|  |  |  | 110 - 130 mm[Hg] |
|  |  |  | 130 - 140 mm[Hg] |
|  |  |  | 140 - 160 mm[Hg] |
|  |  |  | 160 - 0 mm[Hg] |
|  | TNX:9086 | Blood Pressure, Diastolic | Categorical |
|  |  |  | 0 - 70 mm[Hg] |
|  |  |  | 70 - 80 mm[Hg] |
|  |  |  | 80 - 90 mm[Hg] |
|  |  |  | 90 - 0 mm[Hg] |
|  | TNX:9004 | Triglyceride [Mass/volume] in Serum, Plasma or Blood | Categorical |
|  |  |  | 0 - 150 mg/dL |
|  |  |  | 150 - 200 mg/dL |
|  |  |  | 200 - 500 mg/dL |
|  |  |  | 500 - 0 mg/dL |
|  | TNX:9014 | Hemoglobin [Mass/volume] in Blood | Categorical |
|  |  |  | 0 - 10 g/dL |
|  |  |  | 10 - 12 g/dL |
|  |  |  | 12 - 14 g/dL |
|  |  |  | 14 - 16 g/dL |
|  |  |  | 16 - 0 g/dL |
|  | TNX:8001 | Glomerular filtration rate/1.73 sq M.predicted [Volume Rate/Area] in Serum, Plasma or Blood by Creatinine-based formula (MDRD) | Categorical |
|  |  |  | 0 - 15 mL/min/{1.73_m2} |
|  |  |  | 15 - 30 mL/min/{1.73_m2} |
|  |  |  | 30 - 45 mL/min/{1.73_m2} |
|  |  |  | 45 - 60 mL/min/{1.73_m2} |
|  |  |  | 60 - 0 mL/min/{1.73_m2} |
|  | TNX:9029 | Sodium [Moles/volume] in Serum, Plasma or Blood | Categorical |
|  |  |  | 0 - 135 mmol/L |
|  |  |  | 135 - 145 mmol/L |
|  |  |  | 145 - 200 mmol/L |
|  |  |  | 200 - 0 mmol/L |
|  | TNX:9028 | Potassium [Moles/volume] in Serum, Plasma or Blood | Categorical |
|  |  |  | 0 - 3.50 mmol/L |
|  |  |  | 3.60 - 5 mmol/L |
|  |  |  | 5.10 - 5.50 mmol/L |
|  |  |  | 5.50 - 0 mmol/L |

### **10. Multivariable Cox regression analyses for clinical outcomes**

##### **Table S58. MAKE outcomes after multivariable Cox regression analysis.**

| **Cohort Information** | | |
| --- | --- | --- |
|  | **Cohort Name** | **Index Count** |
|  | SGLT-2i-User | 792 |
|  | noSGLT-2i-User | 45292 |

| **Cox Model Results** | | | | | | | |
| --- | --- | --- | --- | --- | --- | --- | --- |
|  | **Covariate** | **Hazard Ratio** | **Coefficient** | **Standard Error** | **z** | **P > \|z\|** | **95% Confidence Interval** |
|  | Cohort 1 or Cohort 2 Membership | 0.492 | -0.709 | 0.125 | -5.649 | 0.000 | (0.385, 0.630) |
| M | Male | 1.202 | 0.184 | 0.030 | 6.157 | 0.000 | (1.134, 1.275) |
| AI | Age at Index | 1.027 | 0.027 | 0.001 | 25.747 | 0 | (1.025, 1.029) |
| 2186-5 | Not Hispanic or Latino | 1.118 | 0.112 | 0.032 | 3.475 | 0.001 | (1.050, 1.191) |
| 2106-3 | White | 0.946 | -0.056 | 0.030 | -1.841 | 0.066 | (0.892, 1.004) |
| I10-I1A | Hypertensive diseases | 1.120 | 0.113 | 0.059 | 1.918 | 0.055 | (0.998, 1.258) |
| I70-I79 | Diseases of arteries, arterioles and capillaries | 1.094 | 0.090 | 0.061 | 1.464 | 0.143 | (0.970, 1.234) |
| I60-I69 | Cerebrovascular diseases | 1.356 | 0.305 | 0.086 | 3.535 | 0.000 | (1.145, 1.605) |
| I26-I28 | Pulmonary heart disease and diseases of pulmonary circulation | 0.987 | -0.013 | 0.098 | -0.129 | 0.897 | (0.815, 1.197) |
| E66 | Overweight and obesity | 1.039 | 0.039 | 0.083 | 0.466 | 0.641 | (0.884, 1.223) |
| C00-D49 | Neoplasms | 1.151 | 0.141 | 0.046 | 3.091 | 0.002 | (1.053, 1.258) |
| K70-K77 | Diseases of liver | 0.809 | -0.212 | 0.078 | -2.732 | 0.006 | (0.695, 0.942) |
| R80 | Proteinuria | 1.112 | 0.106 | 0.067 | 1.583 | 0.114 | (0.975, 1.267) |
| M30-M36 | Systemic connective tissue disorders | 1.073 | 0.071 | 0.080 | 0.882 | 0.378 | (0.917, 1.256) |
| I10 | Essential (primary) hypertension | 0.853 | -0.159 | 0.056 | -2.824 | 0.005 | (0.764, 0.953) |
| I50.2 | Systolic (congestive) heart failure | 1.358 | 0.306 | 0.146 | 2.096 | 0.036 | (1.020, 1.809) |
| I50.3 | Diastolic (congestive) heart failure | 1.400 | 0.336 | 0.143 | 2.347 | 0.019 | (1.057, 1.854) |
| N39.0 | Urinary tract infection, site not specified | 1.121 | 0.114 | 0.061 | 1.870 | 0.061 | (0.995, 1.263) |
| I20-I25 | Ischemic heart diseases | 1.320 | 0.278 | 0.064 | 4.349 | 0.000 | (1.165, 1.496) |
| J40-J4A | Chronic lower respiratory diseases | 1.136 | 0.128 | 0.070 | 1.818 | 0.069 | (0.990, 1.303) |
| E78 | Disorders of lipoprotein metabolism and other lipidemias | 0.911 | -0.093 | 0.047 | -1.982 | 0.048 | (0.831, 0.999) |
| I21 | Acute myocardial infarction | 0.616 | -0.485 | 0.199 | -2.441 | 0.015 | (0.417, 0.909) |
| I50.9 | Heart failure, unspecified | 1.238 | 0.213 | 0.100 | 2.132 | 0.033 | (1.017, 1.506) |
| N17 | Acute kidney failure | 1.184 | 0.169 | 0.064 | 2.642 | 0.008 | (1.045, 1.343) |
| N18 | Chronic kidney disease (CKD) | 1.138 | 0.129 | 0.039 | 3.348 | 0.001 | (1.055, 1.227) |
| N19 | Unspecified kidney failure | 1.115 | 0.109 | 0.089 | 1.229 | 0.219 | (0.937, 1.327) |
| Q61 | Cystic kidney disease | 0.782 | -0.246 | 0.069 | -3.587 | 0.000 | (0.684, 0.895) |
| CV100 | BETA BLOCKERS/RELATED | 1.052 | 0.051 | 0.042 | 1.196 | 0.232 | (0.968, 1.143) |
| CV200 | CALCIUM CHANNEL BLOCKERS | 0.998 | -0.002 | 0.043 | -0.043 | 0.966 | (0.917, 1.087) |
| CV300 | ANTIARRHYTHMICS | 0.998 | -0.002 | 0.050 | -0.049 | 0.961 | (0.905, 1.099) |
| CV700 | DIURETICS | 0.945 | -0.057 | 0.048 | -1.190 | 0.234 | (0.861, 1.037) |
| CV350 | ANTILIPEMIC AGENTS | 1.060 | 0.059 | 0.127 | 0.460 | 0.646 | (0.826, 1.361) |
| CV800 | ACE INHIBITORS | 1.085 | 0.082 | 0.053 | 1.540 | 0.124 | (0.978, 1.204) |
| CV805 | ANGIOTENSIN II INHIBITOR | 0.925 | -0.078 | 0.059 | -1.327 | 0.185 | (0.824, 1.038) |
| CV250 | ANTIANGINALS | 1.023 | 0.022 | 0.086 | 0.259 | 0.796 | (0.863, 1.211) |
| C10AA | HMG CoA reductase inhibitors | 0.926 | -0.077 | 0.130 | -0.592 | 0.554 | (0.717, 1.195) |
| HS502 | ORAL HYPOGLYCEMIC AGENTS,ORAL | 1.043 | 0.043 | 0.240 | 0.177 | 0.859 | (0.652, 1.669) |
| 9002 | Cholesterol in LDL [Mass/volume] in Serum or Plasma |  |  |  |  |  |  |
|  | 0 - 55 mg/dL | 1.178 | 0.164 | 0.099 | 1.663 | 0.096 | (0.971, 1.429) |
|  | 55 - 100 mg/dL | 1.370 | 0.315 | 0.067 | 4.700 | 0.000 | (1.201, 1.562) |
|  | 100 - 130 mg/dL | 1.213 | 0.193 | 0.070 | 2.781 | 0.005 | (1.059, 1.390) |
|  | 130 - 160 mg/dL | 1.286 | 0.252 | 0.091 | 2.774 | 0.006 | (1.077, 1.537) |
|  | 160 - 190 mg/dL | 1.221 | 0.200 | 0.140 | 1.426 | 0.154 | (0.928, 1.608) |
|  | 190 - 0 mg/dL | 0.920 | -0.084 | 0.226 | -0.372 | 0.710 | (0.591, 1.431) |
| 9001 | Cholesterol in HDL [Mass/volume] in Serum or Plasma |  |  |  |  |  |  |
|  | 0 - 45 mg/dL | 0.983 | -0.017 | 0.079 | -0.221 | 0.825 | (0.842, 1.147) |
|  | 45 - 60 mg/dL | 1.031 | 0.030 | 0.074 | 0.406 | 0.685 | (0.891, 1.192) |
|  | 60 - 0 mg/dL | 1.046 | 0.045 | 0.084 | 0.533 | 0.594 | (0.888, 1.232) |
| 9083 | BMI |  |  |  |  |  |  |
|  | 0 - 21 kg/m2 | 1.069 | 0.066 | 0.062 | 1.076 | 0.282 | (0.947, 1.206) |
|  | 21 - 26 kg/m2 | 0.966 | -0.034 | 0.045 | -0.770 | 0.441 | (0.885, 1.055) |
|  | 26 - 31 kg/m2 | 0.933 | -0.069 | 0.045 | -1.528 | 0.126 | (0.854, 1.020) |
|  | 31 - 36 kg/m2 | 1.013 | 0.013 | 0.057 | 0.223 | 0.824 | (0.906, 1.132) |
|  | 36 - 0 kg/m2 | 1.061 | 0.060 | 0.078 | 0.764 | 0.445 | (0.911, 1.237) |
| 9085 | Blood Pressure, Systolic |  |  |  |  |  |  |
|  | 0 - 110 mm[Hg] | 0.975 | -0.025 | 0.057 | -0.440 | 0.660 | (0.873, 1.090) |
|  | 110 - 130 mm[Hg] | 0.992 | -0.008 | 0.055 | -0.152 | 0.879 | (0.891, 1.104) |
|  | 130 - 140 mm[Hg] | 0.900 | -0.105 | 0.051 | -2.045 | 0.041 | (0.814, 0.996) |
|  | 140 - 160 mm[Hg] | 1.055 | 0.054 | 0.052 | 1.028 | 0.304 | (0.953, 1.169) |
|  | 160 - 0 mm[Hg] | 1.139 | 0.130 | 0.054 | 2.416 | 0.016 | (1.025, 1.266) |
| 9086 | Blood Pressure, Diastolic |  |  |  |  |  |  |
|  | 0 - 70 mm[Hg] | 1.149 | 0.139 | 0.056 | 2.483 | 0.013 | (1.030, 1.282) |
|  | 70 - 80 mm[Hg] | 1.045 | 0.044 | 0.053 | 0.829 | 0.407 | (0.941, 1.160) |
|  | 80 - 90 mm[Hg] | 0.894 | -0.112 | 0.052 | -2.172 | 0.030 | (0.807, 0.989) |
|  | 90 - 0 mm[Hg] | 1.090 | 0.086 | 0.054 | 1.604 | 0.109 | (0.981, 1.212) |
| 9004 | Triglyceride [Mass/volume] in Serum, Plasma or Blood |  |  |  |  |  |  |
|  | 0 - 150 mg/dL | 0.721 | -0.327 | 0.065 | -5.060 | 0.000 | (0.635, 0.818) |
|  | 150 - 200 mg/dL | 0.821 | -0.198 | 0.074 | -2.688 | 0.007 | (0.710, 0.948) |
|  | 200 - 500 mg/dL | 0.812 | -0.208 | 0.080 | -2.600 | 0.009 | (0.695, 0.950) |
|  | 500 - 0 mg/dL | 0.994 | -0.006 | 0.255 | -0.025 | 0.980 | (0.603, 1.637) |
| 9014 | Hemoglobin [Mass/volume] in Blood |  |  |  |  |  |  |
|  | 0 - 10 g/dL | 1.184 | 0.169 | 0.054 | 3.149 | 0.002 | (1.066, 1.315) |
|  | 10 - 12 g/dL | 1.120 | 0.113 | 0.047 | 2.422 | 0.015 | (1.022, 1.227) |
|  | 12 - 14 g/dL | 0.911 | -0.093 | 0.042 | -2.242 | 0.025 | (0.840, 0.988) |
|  | 14 - 16 g/dL | 0.855 | -0.157 | 0.053 | -2.947 | 0.003 | (0.770, 0.949) |
|  | 16 - 0 g/dL | 0.878 | -0.131 | 0.099 | -1.324 | 0.185 | (0.723, 1.065) |
| 8001 | Glomerular filtration rate/1.73 sq M.predicted [Volume Rate/Area] in Serum, Plasma or Blood by Creatinine-based formula (MDRD) |  |  |  |  |  |  |
|  | 0 - 15 mL/min/{1.73_m2} | 0.753 | -0.284 | 0.051 | -5.611 | 0.000 | (0.682, 0.831) |
|  | 15 - 30 mL/min/{1.73_m2} | 1.219 | 0.198 | 0.051 | 3.892 | 0.000 | (1.103, 1.347) |
|  | 30 - 45 mL/min/{1.73_m2} | 1.161 | 0.149 | 0.052 | 2.869 | 0.004 | (1.048, 1.285) |
|  | 45 - 60 mL/min/{1.73_m2} | 0.957 | -0.044 | 0.050 | -0.879 | 0.379 | (0.867, 1.056) |
|  | 60 - 0 mL/min/{1.73_m2} | 0.785 | -0.242 | 0.051 | -4.764 | 0.000 | (0.711, 0.867) |
| 9029 | Sodium [Moles/volume] in Serum, Plasma or Blood |  |  |  |  |  |  |
|  | 0 - 135 mmol/L | 0.953 | -0.048 | 0.052 | -0.923 | 0.356 | (0.861, 1.055) |
|  | 135 - 145 mmol/L | 1.044 | 0.043 | 0.063 | 0.687 | 0.492 | (0.923, 1.180) |
|  | 145 - 200 mmol/L | 1.143 | 0.133 | 0.077 | 1.724 | 0.085 | (0.982, 1.330) |
|  | 200 - 0 mmol/L | 0.001 | -7.274 | 203.068 | -0.036 | 0.971 | (0, Infinity) |
| 9028 | Potassium [Moles/volume] in Serum, Plasma or Blood |  |  |  |  |  |  |
|  | 0 - 3.50 mmol/L | 1.028 | 0.028 | 0.055 | 0.511 | 0.610 | (0.923, 1.145) |
|  | 3.60 - 5 mmol/L | 0.883 | -0.124 | 0.060 | -2.079 | 0.038 | (0.785, 0.993) |
|  | 5.10 - 5.50 mmol/L | 0.891 | -0.116 | 0.055 | -2.086 | 0.037 | (0.799, 0.993) |
|  | 5.50 - 0 mmol/L | 1.063 | 0.061 | 0.064 | 0.958 | 0.338 | (0.938, 1.204) |

##### **Table S59. All-cause mortality outcomes after multivariable Cox regression analysis.**

| **Cohort Information** | | |
| --- | --- | --- |
|  | **Cohort Name** | **Index Count** |
|  | SGLT-2i-User | 792 |
|  | noSGLT-2i-User | 45292 |

| **Cox Model Results** | | | | | | | |
| --- | --- | --- | --- | --- | --- | --- | --- |
|  | **Covariate** | **Hazard Ratio** | **Coefficient** | **Standard Error** | **z** | **P > \|z\|** | **95% Confidence Interval** |
|  | Cohort 1 or Cohort 2 Membership | 0.324 | -1.126 | 0.173 | -6.509 | 0.000 | (0.231, 0.455) |
| M | Male | 1.238 | 0.214 | 0.035 | 6.048 | 0.000 | (1.155, 1.327) |
| AI | Age at Index | 1.055 | 0.054 | 0.001 | 39.083 | 0 | (1.052, 1.058) |
| 2186-5 | Not Hispanic or Latino | 1.074 | 0.071 | 0.038 | 1.857 | 0.063 | (0.996, 1.158) |
| 2106-3 | White | 1.072 | 0.070 | 0.036 | 1.923 | 0.054 | (0.999, 1.152) |
| I10-I1A | Hypertensive diseases | 1.006 | 0.006 | 0.073 | 0.078 | 0.938 | (0.872, 1.160) |
| I70-I79 | Diseases of arteries, arterioles and capillaries | 1.019 | 0.019 | 0.072 | 0.265 | 0.791 | (0.885, 1.174) |
| I60-I69 | Cerebrovascular diseases | 1.403 | 0.339 | 0.096 | 3.518 | 0.000 | (1.162, 1.695) |
| I26-I28 | Pulmonary heart disease and diseases of pulmonary circulation | 1.124 | 0.117 | 0.109 | 1.076 | 0.282 | (0.908, 1.392) |
| E66 | Overweight and obesity | 1.132 | 0.124 | 0.096 | 1.289 | 0.197 | (0.937, 1.368) |
| C00-D49 | Neoplasms | 1.150 | 0.140 | 0.051 | 2.733 | 0.006 | (1.040, 1.272) |
| K70-K77 | Diseases of liver | 0.809 | -0.212 | 0.090 | -2.366 | 0.018 | (0.679, 0.964) |
| R80 | Proteinuria | 1.115 | 0.109 | 0.081 | 1.344 | 0.179 | (0.951, 1.308) |
| M30-M36 | Systemic connective tissue disorders | 1.087 | 0.083 | 0.102 | 0.817 | 0.414 | (0.890, 1.327) |
| I10 | Essential (primary) hypertension | 0.865 | -0.145 | 0.070 | -2.074 | 0.038 | (0.755, 0.992) |
| I50.2 | Systolic (congestive) heart failure | 1.521 | 0.419 | 0.158 | 2.657 | 0.008 | (1.116, 2.072) |
| I50.3 | Diastolic (congestive) heart failure | 1.232 | 0.209 | 0.161 | 1.296 | 0.195 | (0.899, 1.689) |
| N39.0 | Urinary tract infection, site not specified | 1.017 | 0.017 | 0.074 | 0.225 | 0.822 | (0.879, 1.176) |
| I20-I25 | Ischemic heart diseases | 1.323 | 0.280 | 0.071 | 3.927 | 0.000 | (1.150, 1.521) |
| J40-J4A | Chronic lower respiratory diseases | 1.096 | 0.092 | 0.083 | 1.113 | 0.266 | (0.932, 1.289) |
| E78 | Disorders of lipoprotein metabolism and other lipidemias | 0.870 | -0.139 | 0.055 | -2.519 | 0.012 | (0.781, 0.970) |
| I21 | Acute myocardial infarction | 0.542 | -0.612 | 0.224 | -2.725 | 0.006 | (0.349, 0.842) |
| I50.9 | Heart failure, unspecified | 1.407 | 0.341 | 0.112 | 3.047 | 0.002 | (1.130, 1.752) |
| N17 | Acute kidney failure | 1.052 | 0.051 | 0.080 | 0.635 | 0.526 | (0.900, 1.230) |
| N18 | Chronic kidney disease (CKD) | 1.088 | 0.085 | 0.046 | 1.845 | 0.065 | (0.995, 1.190) |
| N19 | Unspecified kidney failure | 1.164 | 0.152 | 0.110 | 1.382 | 0.167 | (0.938, 1.445) |
| Q61 | Cystic kidney disease | 0.811 | -0.210 | 0.080 | -2.615 | 0.009 | (0.693, 0.949) |
| CV100 | BETA BLOCKERS/RELATED | 1.098 | 0.094 | 0.050 | 1.873 | 0.061 | (0.996, 1.212) |
| CV200 | CALCIUM CHANNEL BLOCKERS | 0.925 | -0.078 | 0.052 | -1.509 | 0.131 | (0.835, 1.024) |
| CV300 | ANTIARRHYTHMICS | 1.056 | 0.055 | 0.059 | 0.934 | 0.350 | (0.942, 1.185) |
| CV700 | DIURETICS | 1.039 | 0.038 | 0.055 | 0.688 | 0.491 | (0.932, 1.158) |
| CV350 | ANTILIPEMIC AGENTS | 1.127 | 0.120 | 0.145 | 0.826 | 0.409 | (0.848, 1.497) |
| CV800 | ACE INHIBITORS | 0.986 | -0.014 | 0.065 | -0.214 | 0.831 | (0.868, 1.120) |
| CV805 | ANGIOTENSIN II INHIBITOR | 0.924 | -0.079 | 0.070 | -1.124 | 0.261 | (0.805, 1.061) |
| CV250 | ANTIANGINALS | 0.970 | -0.030 | 0.101 | -0.300 | 0.764 | (0.796, 1.183) |
| C10AA | HMG CoA reductase inhibitors | 0.908 | -0.096 | 0.148 | -0.650 | 0.516 | (0.680, 1.214) |
| HS502 | ORAL HYPOGLYCEMIC AGENTS,ORAL | 1.241 | 0.216 | 0.250 | 0.862 | 0.388 | (0.760, 2.024) |
| 9002 | Cholesterol in LDL [Mass/volume] in Serum or Plasma |  |  |  |  |  |  |
|  | 0 - 55 mg/dL | 1.132 | 0.124 | 0.113 | 1.092 | 0.275 | (0.906, 1.414) |
|  | 55 - 100 mg/dL | 1.164 | 0.152 | 0.079 | 1.916 | 0.055 | (0.997, 1.360) |
|  | 100 - 130 mg/dL | 1.045 | 0.044 | 0.083 | 0.534 | 0.593 | (0.889, 1.230) |
|  | 130 - 160 mg/dL | 1.052 | 0.051 | 0.110 | 0.459 | 0.646 | (0.847, 1.306) |
|  | 160 - 190 mg/dL | 1.155 | 0.144 | 0.165 | 0.873 | 0.383 | (0.836, 1.594) |
|  | 190 - 0 mg/dL | 0.834 | -0.181 | 0.276 | -0.655 | 0.513 | (0.485, 1.434) |
| 9001 | Cholesterol in HDL [Mass/volume] in Serum or Plasma |  |  |  |  |  |  |
|  | 0 - 45 mg/dL | 1.030 | 0.030 | 0.092 | 0.327 | 0.744 | (0.861, 1.233) |
|  | 45 - 60 mg/dL | 1.110 | 0.104 | 0.086 | 1.217 | 0.224 | (0.938, 1.314) |
|  | 60 - 0 mg/dL | 1.121 | 0.114 | 0.096 | 1.195 | 0.232 | (0.929, 1.352) |
| 9083 | BMI |  |  |  |  |  |  |
|  | 0 - 21 kg/m2 | 1.020 | 0.020 | 0.078 | 0.258 | 0.797 | (0.876, 1.188) |
|  | 21 - 26 kg/m2 | 0.973 | -0.028 | 0.053 | -0.520 | 0.603 | (0.876, 1.080) |
|  | 26 - 31 kg/m2 | 0.993 | -0.007 | 0.052 | -0.137 | 0.891 | (0.896, 1.100) |
|  | 31 - 36 kg/m2 | 1.059 | 0.057 | 0.066 | 0.874 | 0.382 | (0.931, 1.205) |
|  | 36 - 0 kg/m2 | 1.088 | 0.084 | 0.092 | 0.913 | 0.361 | (0.908, 1.303) |
| 9085 | Blood Pressure, Systolic |  |  |  |  |  |  |
|  | 0 - 110 mm[Hg] | 1.060 | 0.058 | 0.067 | 0.878 | 0.380 | (0.930, 1.208) |
|  | 110 - 130 mm[Hg] | 0.968 | -0.032 | 0.064 | -0.503 | 0.615 | (0.854, 1.098) |
|  | 130 - 140 mm[Hg] | 0.817 | -0.203 | 0.060 | -3.371 | 0.001 | (0.726, 0.919) |
|  | 140 - 160 mm[Hg] | 1.086 | 0.083 | 0.060 | 1.371 | 0.171 | (0.965, 1.223) |
|  | 160 - 0 mm[Hg] | 1.126 | 0.119 | 0.063 | 1.880 | 0.060 | (0.995, 1.275) |
| 9086 | Blood Pressure, Diastolic |  |  |  |  |  |  |
|  | 0 - 70 mm[Hg] | 1.120 | 0.113 | 0.066 | 1.710 | 0.087 | (0.984, 1.275) |
|  | 70 - 80 mm[Hg] | 1.133 | 0.124 | 0.063 | 1.988 | 0.047 | (1.002, 1.280) |
|  | 80 - 90 mm[Hg] | 0.920 | -0.083 | 0.060 | -1.389 | 0.165 | (0.818, 1.035) |
|  | 90 - 0 mm[Hg] | 1.006 | 0.006 | 0.064 | 0.094 | 0.925 | (0.888, 1.140) |
| 9004 | Triglyceride [Mass/volume] in Serum, Plasma or Blood |  |  |  |  |  |  |
|  | 0 - 150 mg/dL | 0.834 | -0.182 | 0.075 | -2.418 | 0.016 | (0.719, 0.966) |
|  | 150 - 200 mg/dL | 0.951 | -0.050 | 0.084 | -0.598 | 0.550 | (0.806, 1.122) |
|  | 200 - 500 mg/dL | 0.903 | -0.102 | 0.093 | -1.093 | 0.274 | (0.752, 1.084) |
|  | 500 - 0 mg/dL | 0.988 | -0.012 | 0.321 | -0.039 | 0.969 | (0.526, 1.853) |
| 9014 | Hemoglobin [Mass/volume] in Blood |  |  |  |  |  |  |
|  | 0 - 10 g/dL | 1.104 | 0.099 | 0.066 | 1.493 | 0.135 | (0.970, 1.256) |
|  | 10 - 12 g/dL | 1.117 | 0.110 | 0.056 | 1.965 | 0.049 | (1.000, 1.246) |
|  | 12 - 14 g/dL | 0.923 | -0.080 | 0.049 | -1.624 | 0.104 | (0.837, 1.017) |
|  | 14 - 16 g/dL | 0.906 | -0.099 | 0.061 | -1.615 | 0.106 | (0.803, 1.021) |
|  | 16 - 0 g/dL | 0.958 | -0.043 | 0.111 | -0.386 | 0.699 | (0.771, 1.191) |
| 8001 | Glomerular filtration rate/1.73 sq M.predicted [Volume Rate/Area] in Serum, Plasma or Blood by Creatinine-based formula (MDRD) |  |  |  |  |  |  |
|  | 0 - 15 mL/min/{1.73_m2} | 0.842 | -0.172 | 0.061 | -2.800 | 0.005 | (0.747, 0.950) |
|  | 15 - 30 mL/min/{1.73_m2} | 1.280 | 0.247 | 0.061 | 4.055 | 0.000 | (1.136, 1.442) |
|  | 30 - 45 mL/min/{1.73_m2} | 1.150 | 0.140 | 0.060 | 2.326 | 0.020 | (1.022, 1.295) |
|  | 45 - 60 mL/min/{1.73_m2} | 1.047 | 0.046 | 0.057 | 0.803 | 0.422 | (0.936, 1.172) |
|  | 60 - 0 mL/min/{1.73_m2} | 0.891 | -0.115 | 0.058 | -1.984 | 0.047 | (0.796, 0.999) |
| 9029 | Sodium [Moles/volume] in Serum, Plasma or Blood |  |  |  |  |  |  |
|  | 0 - 135 mmol/L | 0.908 | -0.097 | 0.063 | -1.540 | 0.124 | (0.803, 1.027) |
|  | 135 - 145 mmol/L | 1.026 | 0.025 | 0.073 | 0.346 | 0.730 | (0.888, 1.184) |
|  | 145 - 200 mmol/L | 1.106 | 0.101 | 0.091 | 1.110 | 0.267 | (0.926, 1.321) |
|  | 200 - 0 mmol/L | 0.003 | -5.967 | 282.501 | -0.021 | 0.983 | (0, Infinity) |
| 9028 | Potassium [Moles/volume] in Serum, Plasma or Blood |  |  |  |  |  |  |
|  | 0 - 3.50 mmol/L | 1.036 | 0.035 | 0.065 | 0.537 | 0.591 | (0.912, 1.176) |
|  | 3.60 - 5 mmol/L | 0.803 | -0.220 | 0.070 | -3.146 | 0.002 | (0.700, 0.920) |
|  | 5.10 - 5.50 mmol/L | 0.856 | -0.155 | 0.067 | -2.316 | 0.021 | (0.751, 0.976) |
|  | 5.50 - 0 mmol/L | 1.033 | 0.033 | 0.078 | 0.417 | 0.677 | (0.886, 1.204) |

##### **Table S60. MACE outcomes after multivariable Cox regression analysis.**

| **Cohort Information** | | |
| --- | --- | --- |
|  | **Cohort Name** | **Index Count** |
|  | SGLT-2i-User | 732 |
|  | noSGLT-2i-User | 42095 |

| **Cox Model Results** | | | | | | | |
| --- | --- | --- | --- | --- | --- | --- | --- |
|  | **Covariate** | **Hazard Ratio** | **Coefficient** | **Standard Error** | **z** | **P > \|z\|** | **95% Confidence Interval** |
|  | Cohort 1 or Cohort 2 Membership | 0.763 | -0.271 | 0.105 | -2.570 | 0.010 | (0.620, 1.028) |
| M | Male | 1.177 | 0.163 | 0.031 | 5.208 | 0.000 | (1.107, 1.251) |
| AI | Age at Index | 1.046 | 0.045 | 0.001 | 38.035 | 0 | (1.043, 1.048) |
| 2186-5 | Not Hispanic or Latino | 1.113 | 0.107 | 0.034 | 3.112 | 0.002 | (1.040, 1.190) |
| 2106-3 | White | 1.028 | 0.028 | 0.032 | 0.880 | 0.379 | (0.966, 1.095) |
| I10-I1A | Hypertensive diseases | 0.995 | -0.005 | 0.064 | -0.072 | 0.943 | (0.877, 1.129) |
| I70-I79 | Diseases of arteries, arterioles and capillaries | 1.046 | 0.045 | 0.063 | 0.719 | 0.472 | (0.925, 1.184) |
| I60-I69 | Cerebrovascular diseases | 2.003 | 0.695 | 0.078 | 8.945 | 0.000 | (1.720, 2.333) |
| I26-I28 | Pulmonary heart disease and diseases of pulmonary circulation | 1.188 | 0.172 | 0.098 | 1.747 | 0.081 | (0.979, 1.440) |
| E66 | Overweight and obesity | 1.070 | 0.068 | 0.086 | 0.791 | 0.429 | (0.905, 1.265) |
| C00-D49 | Neoplasms | 1.169 | 0.156 | 0.046 | 3.353 | 0.001 | (1.067, 1.280) |
| K70-K77 | Diseases of liver | 0.859 | -0.152 | 0.076 | -1.982 | 0.047 | (0.740, 0.998) |
| R80 | Proteinuria | 1.142 | 0.133 | 0.071 | 1.877 | 0.060 | (0.994, 1.312) |
| M30-M36 | Systemic connective tissue disorders | 1.152 | 0.141 | 0.086 | 1.638 | 0.101 | (0.973, 1.364) |
| I10 | Essential (primary) hypertension | 0.899 | -0.106 | 0.062 | -1.730 | 0.084 | (0.797, 1.014) |
| I50.2 | Systolic (congestive) heart failure | 1.463 | 0.380 | 0.139 | 2.732 | 0.006 | (1.113, 1.922) |
| I50.3 | Diastolic (congestive) heart failure | 1.316 | 0.275 | 0.141 | 1.952 | 0.051 | (0.999, 1.735) |
| N39.0 | Urinary tract infection, site not specified | 1.077 | 0.074 | 0.065 | 1.131 | 0.258 | (0.947, 1.223) |
| I20-I25 | Ischemic heart diseases | 1.483 | 0.394 | 0.063 | 6.269 | 0.000 | (1.311, 1.678) |
| J40-J4A | Chronic lower respiratory diseases | 1.214 | 0.194 | 0.072 | 2.687 | 0.007 | (1.054, 1.398) |
| E78 | Disorders of lipoprotein metabolism and other lipidemias | 0.920 | -0.084 | 0.049 | -1.699 | 0.089 | (0.835, 1.013) |
| I21 | Acute myocardial infarction | 1.190 | 0.174 | 0.150 | 1.162 | 0.245 | (0.888, 1.595) |
| I50.9 | Heart failure, unspecified | 1.100 | 0.095 | 0.104 | 0.913 | 0.361 | (0.897, 1.350) |
| N17 | Acute kidney failure | 1.028 | 0.027 | 0.072 | 0.381 | 0.703 | (0.893, 1.183) |
| N18 | Chronic kidney disease (CKD) | 1.042 | 0.041 | 0.041 | 1.001 | 0.317 | (0.962, 1.129) |
| N19 | Unspecified kidney failure | 1.164 | 0.152 | 0.098 | 1.553 | 0.120 | (0.961, 1.409) |
| Q61 | Cystic kidney disease | 0.869 | -0.141 | 0.072 | -1.965 | 0.049 | (0.755, 1.000) |
| CV100 | BETA BLOCKERS/RELATED | 1.129 | 0.121 | 0.044 | 2.762 | 0.006 | (1.036, 1.231) |
| CV200 | CALCIUM CHANNEL BLOCKERS | 0.969 | -0.031 | 0.046 | -0.684 | 0.494 | (0.886, 1.060) |
| CV300 | ANTIARRHYTHMICS | 1.024 | 0.024 | 0.052 | 0.453 | 0.650 | (0.924, 1.135) |
| CV700 | DIURETICS | 1.004 | 0.004 | 0.049 | 0.082 | 0.935 | (0.912, 1.106) |
| CV350 | ANTILIPEMIC AGENTS | 1.053 | 0.052 | 0.135 | 0.384 | 0.701 | (0.808, 1.373) |
| CV800 | ACE INHIBITORS | 1.101 | 0.096 | 0.055 | 1.740 | 0.082 | (0.988, 1.228) |
| CV805 | ANGIOTENSIN II INHIBITOR | 0.939 | -0.063 | 0.061 | -1.041 | 0.298 | (0.833, 1.058) |
| CV250 | ANTIANGINALS | 1.076 | 0.073 | 0.083 | 0.882 | 0.378 | (0.914, 1.267) |
| C10AA | HMG CoA reductase inhibitors | 0.943 | -0.059 | 0.138 | -0.426 | 0.670 | (0.719, 1.236) |
| HS502 | ORAL HYPOGLYCEMIC AGENTS,ORAL | 0.928 | -0.075 | 0.228 | -0.329 | 0.742 | (0.593, 1.452) |
| 9002 | Cholesterol in LDL [Mass/volume] in Serum or Plasma |  |  |  |  |  |  |
|  | 0 - 55 mg/dL | 1.151 | 0.140 | 0.102 | 1.377 | 0.168 | (0.942, 1.405) |
|  | 55 - 100 mg/dL | 1.199 | 0.181 | 0.072 | 2.514 | 0.012 | (1.041, 1.380) |
|  | 100 - 130 mg/dL | 1.080 | 0.077 | 0.076 | 1.018 | 0.309 | (0.931, 1.254) |
|  | 130 - 160 mg/dL | 1.193 | 0.176 | 0.096 | 1.831 | 0.067 | (0.988, 1.441) |
|  | 160 - 190 mg/dL | 1.254 | 0.226 | 0.142 | 1.589 | 0.112 | (0.949, 1.658) |
|  | 190 - 0 mg/dL | 0.961 | -0.040 | 0.223 | -0.180 | 0.857 | (0.620, 1.488) |
| 9001 | Cholesterol in HDL [Mass/volume] in Serum or Plasma |  |  |  |  |  |  |
|  | 0 - 45 mg/dL | 0.958 | -0.043 | 0.084 | -0.510 | 0.610 | (0.814, 1.129) |
|  | 45 - 60 mg/dL | 0.985 | -0.016 | 0.079 | -0.196 | 0.844 | (0.843, 1.150) |
|  | 60 - 0 mg/dL | 0.936 | -0.067 | 0.089 | -0.745 | 0.457 | (0.785, 1.115) |
| 9083 | BMI |  |  |  |  |  |  |
|  | 0 - 21 kg/m2 | 1.049 | 0.048 | 0.069 | 0.695 | 0.487 | (0.917, 1.200) |
|  | 21 - 26 kg/m2 | 0.986 | -0.014 | 0.047 | -0.304 | 0.761 | (0.899, 1.081) |
|  | 26 - 31 kg/m2 | 1.032 | 0.031 | 0.046 | 0.676 | 0.499 | (0.942, 1.130) |
|  | 31 - 36 kg/m2 | 1.057 | 0.056 | 0.059 | 0.947 | 0.344 | (0.942, 1.186) |
|  | 36 - 0 kg/m2 | 1.045 | 0.044 | 0.082 | 0.533 | 0.594 | (0.889, 1.228) |
| 9085 | Blood Pressure, Systolic |  |  |  |  |  |  |
|  | 0 - 110 mm[Hg] | 1.070 | 0.068 | 0.061 | 1.120 | 0.263 | (0.950, 1.206) |
|  | 110 - 130 mm[Hg] | 1.059 | 0.057 | 0.057 | 1.004 | 0.315 | (0.947, 1.185) |
|  | 130 - 140 mm[Hg] | 0.856 | -0.156 | 0.054 | -2.869 | 0.004 | (0.769, 0.952) |
|  | 140 - 160 mm[Hg] | 1.064 | 0.062 | 0.055 | 1.129 | 0.259 | (0.956, 1.184) |
|  | 160 - 0 mm[Hg] | 1.149 | 0.139 | 0.057 | 2.445 | 0.014 | (1.028, 1.285) |
| 9086 | Blood Pressure, Diastolic |  |  |  |  |  |  |
|  | 0 - 70 mm[Hg] | 1.010 | 0.010 | 0.060 | 0.167 | 0.868 | (0.898, 1.136) |
|  | 70 - 80 mm[Hg] | 0.986 | -0.014 | 0.056 | -0.255 | 0.799 | (0.883, 1.101) |
|  | 80 - 90 mm[Hg] | 0.957 | -0.044 | 0.054 | -0.817 | 0.414 | (0.860, 1.064) |
|  | 90 - 0 mm[Hg] | 0.993 | -0.008 | 0.057 | -0.131 | 0.896 | (0.887, 1.111) |
| 9004 | Triglyceride [Mass/volume] in Serum, Plasma or Blood |  |  |  |  |  |  |
|  | 0 - 150 mg/dL | 0.866 | -0.144 | 0.068 | -2.131 | 0.033 | (0.758, 0.989) |
|  | 150 - 200 mg/dL | 1.044 | 0.043 | 0.074 | 0.584 | 0.559 | (0.903, 1.208) |
|  | 200 - 500 mg/dL | 1.014 | 0.014 | 0.081 | 0.175 | 0.861 | (0.866, 1.188) |
|  | 500 - 0 mg/dL | 1.078 | 0.075 | 0.263 | 0.285 | 0.776 | (0.644, 1.804) |
| 9014 | Hemoglobin [Mass/volume] in Blood |  |  |  |  |  |  |
|  | 0 - 10 g/dL | 1.086 | 0.083 | 0.058 | 1.421 | 0.155 | (0.969, 1.218) |
|  | 10 - 12 g/dL | 1.070 | 0.068 | 0.050 | 1.362 | 0.173 | (0.971, 1.180) |
|  | 12 - 14 g/dL | 0.899 | -0.106 | 0.044 | -2.421 | 0.015 | (0.825, 0.980) |
|  | 14 - 16 g/dL | 0.901 | -0.105 | 0.056 | -1.886 | 0.059 | (0.808, 1.004) |
|  | 16 - 0 g/dL | 0.909 | -0.096 | 0.103 | -0.927 | 0.354 | (0.742, 1.113) |
| 8001 | Glomerular filtration rate/1.73 sq M.predicted [Volume Rate/Area] in Serum, Plasma or Blood by Creatinine-based formula (MDRD) |  |  |  |  |  |  |
|  | 0 - 15 mL/min/{1.73_m2} | 0.829 | -0.188 | 0.054 | -3.493 | 0.000 | (0.746, 0.921) |
|  | 15 - 30 mL/min/{1.73_m2} | 1.226 | 0.203 | 0.054 | 3.754 | 0.000 | (1.102, 1.363) |
|  | 30 - 45 mL/min/{1.73_m2} | 1.253 | 0.226 | 0.054 | 4.180 | 0.000 | (1.127, 1.393) |
|  | 45 - 60 mL/min/{1.73_m2} | 0.980 | -0.020 | 0.052 | -0.378 | 0.705 | (0.885, 1.086) |
|  | 60 - 0 mL/min/{1.73_m2} | 0.888 | -0.118 | 0.052 | -2.268 | 0.023 | (0.802, 0.984) |
| 9029 | Sodium [Moles/volume] in Serum, Plasma or Blood |  |  |  |  |  |  |
|  | 0 - 135 mmol/L | 0.957 | -0.044 | 0.056 | -0.778 | 0.437 | (0.858, 1.068) |
|  | 135 - 145 mmol/L | 1.094 | 0.090 | 0.065 | 1.374 | 0.170 | (0.962, 1.243) |
|  | 145 - 200 mmol/L | 0.985 | -0.015 | 0.084 | -0.177 | 0.860 | (0.835, 1.162) |
|  | 200 - 0 mmol/L | 0.001 | -6.508 | 239.378 | -0.027 | 0.978 | (0, Infinity) |
| 9028 | Potassium [Moles/volume] in Serum, Plasma or Blood |  |  |  |  |  |  |
|  | 0 - 3.50 mmol/L | 1.046 | 0.045 | 0.059 | 0.761 | 0.447 | (0.932, 1.173) |
|  | 3.60 - 5 mmol/L | 0.812 | -0.208 | 0.062 | -3.348 | 0.001 | (0.719, 0.917) |
|  | 5.10 - 5.50 mmol/L | 0.851 | -0.162 | 0.059 | -2.722 | 0.006 | (0.757, 0.956) |
|  | 5.50 - 0 mmol/L | 1.064 | 0.062 | 0.068 | 0.917 | 0.359 | (0.931, 1.216) |
